# First-Time Prescribing of GLP-1 Receptor Agonists from 2018-2023: A Descriptive Analysis

**DOI:** 10.1101/2023.08.22.23294277

**Authors:** Patricia J Rodriguez, Samuel Gratzl, Brianna M Goodwin Cartwright, Rajdeep Brar, Ty J Gluckman, Nicholas L Stucky

**Affiliations:** Truveta, Inc, Bellevue, WA; Center for Cardiovascular Analytics, Research and Data Science (CARDS), Providence Heart Institute, Providence St Joseph Health, Portland, OR; Providence St Joseph Health, Portland, OR

**Author notes:** **Corresponding author:** Dr Nicholas Stucky, Truveta, Inc, 1745 114th Ave SE, Bellevue, WA 98004.

**Keywords:** glucagon-like peptide 1 receptor agonists, anti-obesity medication, ant-diabetic medication, real-world data

## Abstract

**Aims:** Limited recent data exist on prescribing patterns and patient characteristics for glucagon-like peptide 1 receptor agonists (GLP-1 RAs), an important drug class for patients with type 2 diabetes mellitus (T2D) and/or overweight or obesity. We describe trends in first-time prescribing of GLP-1 RAs.

**Materials and Methods:** Using aggregated US electronic health record data, we identified first-time prescriptions of GLP-1 RAs between January 2018 and June 2023 for adults receiving regular care. We describe prescribing volumes stratified by patient characteristics, specific drug, and FDA-labelled indication. The 5.5-year analysis period was divided into six-month periods.

**Results:** In total, 274,562 patients were newly prescribed a GLP-1 RA, with a significant increase over time (January to June, 2018: 9,642 versus 2023: 66,569; p < 0.001). Overall, 181,860 (66.2%) patients had T2D and 229,715 (83.7%) had obesity or overweight. The proportion with T2D decreased over time (January through June, 2018: 84.1% versus 2023: 49%; p < 0.001), while the proportion with overweight or obesity increased (January through June, 2018: 75.9% versus 2023: 90.7%; p <.001). Of prescriptions with a known FDA-labelled indication (74.2%), 87% were labelled for T2D and 13% were labelled for overweight or obesity. Patients first prescribed a GLP-1 RA labelled for T2D were 59.1% female with a mean (SD) age of 58.6 (13) years, while those prescribed a GLP-1 RA labelled for overweight or obesity were 82.9% female, with a mean (SD) age of 48.2 (12.2) years.

**Conclusions:** We observed an increase in first-time prescribing of GLP-1 RAs overall, and a shift away from a predominately T2D population.

## Introduction

Obesity and type 2 diabetes mellitus (T2D) are interconnected epidemics with a high prevalence as well as individual and combined negative impacts on health [1, 2]. Glucagon-like peptide-1 receptor agonists (GLP-1 RAs), have been shown to improve glycemic control and and cardiovascular outcomes in patients with T2D and as such represent a key medication class for this population [3–6]. Some GLP-1 RAs have additionally demonstrated significant weight loss in patients with and without T2D [7–11]. Interest in GLP-1 RAs for weight loss has recently accelerated [12, 13], and both increased demand and supply chain disruptions are thought to have contributed to widespread shortages of certain GLP-1 RAs over the last year [14, 15].

Limited clinical data exists on first-time prescribing volumes of GLP-1 RAs over time, segmented by patient characteristics, specific drug, and FDA-labelled use. Accordingly, we used near real-time, aggregated electronic health record (EHR) data from a group of US healthcare systems to describe first-time prescribing of GLP-1 RA medications from January 2018 to June 2023. Importantly, our analysis also includes the dual GLP-1 RA/gastric inhibitory polypeptide (GIP) receptor agonist, tirzepatide.

## Materials and Methods

### Data

We evaluated first-time prescribing of GLP-1 RAs between January 2018 and June 2023 using a subset of Truveta Data. Truveta provides access to continuously updated and linked EHR from a collective of US health systems to enable health services and comparative effectiveness research. Updated data are provided daily to Truveta. Data are normalized into a common data model through syntactic and semantic normalization. The data are then de-identified by expert determination under the HIPAA Privacy Rule. Once de-identified, the data are available for analysis in R or Python using Truveta Studio. Only data available for the full study period were included.

### Population

We sought to capture all first-time GLP-1 RA prescribing among patients with an encounter at a Truveta-constituent healthcare system during the study period, including off-label prescribing. Therefore, patients in our study were not required to have T2D, overweight, or obesity at baseline. Patients first prescribed a GLP-1 RA between January 2018 and June 2023 were identified using RxNorm codes (Supplement). For this analysis, GLP-1 RA prescribing did not require that a GLP-1 RA was filled. We restricted the cohort to adults (≥ 18 years) and excluded patients with recorded type I or gestational diabetes mellitus in the preceding two years. To reduce the risk of exposure and comorbidity misclassification, only patients who received regular care at a Truveta constituent healthcare system, defined as having at least one outpatient encounter in each of the 2 consecutive 6-month periods before GLP-1 RA initiation (e.g.,1–182 days before and 183-365 days before new GLP-1 RA prescribing) were included in the analysis (Figure 1).

**Figure 1:**
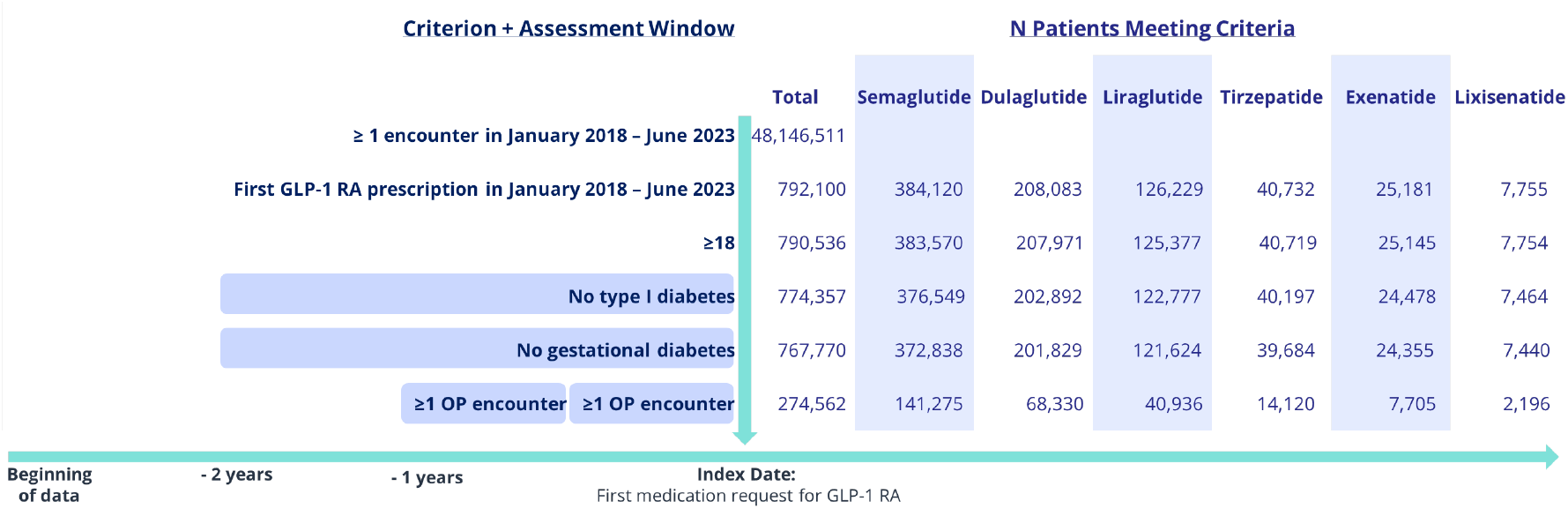
Diagram of inclusion and index criteria and patient counts

### Observation Window

We evaluated first-time GLP-1 RA prescribing, with a patient’s first GLP-1 RA medication request serving as their index date. A two-year look-back window was used to ascertain presence of comorbidities and previous medication use. For baseline quantitative measures of weight, body mass index (BMI), and hemoglobin A1c (HbA1c), we used the most recent value in the 12 months before to up to one-day after the index date. The one-day post-index buffer allowed us to more easily reconcile minor differences in timestamps. The 5.5-year analysis period was divided into 11 six-month periods (January-June and July-December of each year) to facilitate comparison with partial-year 2023.

### Patient Health History

Patients were classified as having T2D if they had a T2D diagnosis, used insulin, dipeptidyl peptidase 4 (DPP-4) inhibitor, or a sulfonylurea, or had an HbA1c level of ≥7.5% in the two years before their index date. Patients were classified as having overweight or obesity if they had a BMI ≥27 kg/m^2^ or a diagnosis of overweight (restricted to diagnoses associated with BMI ≥27 kg/m^2^) or obesity in the two years before their index date. A BMI range of ≥27 to <30 kg/m^2^, rather than the more common ≥25 to <30 kg/m^2^ [16], was used to define overweight to maintain consistency with the FDA label for GLP-1 RAs approved for overweight and obesity [17, 18]. Codes for all data definitions are provided in the Supplement.

Patient demographics, clinical comorbidities, use of other anti-diabetic medication (ADM) (e.g., metformin, sulfonylurea, sodium/glucose cotransporter-2 inhibitors [SGLT2i], DPP-4), use of other anti-obesity medication (AOM) (e.g.,orlistat, phentermine topiramate), and history of bariatric surgery in the 2 years before the index date were assessed. Codes for all comorbidities are provided in the Supplement.

For weight, BMI, and HBA1c, the most recent value within 12 months before to one day after the patient’s index date was considered the baseline value. Pre-processing steps applied to weight, BMI, and HbA1c values are described in the Supplement.

### Brand and Label Classification

Where possible, we attributed a specific brand name and FDA-labelled use (ADM versus AOM) to the GLP-1 RA prescription. An RxNorm code corresponding to a specific brand (e.g., RxNorm code 2619154: 0.25 MG, 0.5 MG Dose 3 ML semaglutide 0.68 MG/ML Pen Injector [Ozempic]) was present for most first-time prescriptions. When no brand-specific RxNorm code was present, the brand was inferred if only one option was available (e.g., all tirzepatide prescriptions were presumed to be brand name Mounjaro; all oral semaglutide prescriptions were presumed to be brand name Rybelsus). When the brand name was not documented and could not be inferred, we classified the brand as unknown. For example, RxNorm code 2619152 (0.25 MG, 0.5 MG Dose 3 ML semaglutide 0.68 MG/ML Pen Injector) was classified as unknown because it could correspond to either brand name Ozempic or Wegovy.

Using the brand name classifications above, prescriptions were categorized as having an FDA-labelled use for either T2D (ADM), overweight/obesity (AOM), or unknown based on approvals as of July 2023. Only prescriptions with a known FDA-labelled use were included in on-versus off-label analyses. The prescription was classified as on-label if the patient had a history of the labelled condition in the previous two years and otherwise it was classified as off-label.

### Analyses

We summarized total monthly prescriptions overall and stratified by specific medication, then applied seasonally-adjusted autoregressive (AR(1]) models to test for trends over time. We compared differences in proportions between January - June 2018 and January - June 2023 using two-sample tests of proportions, and differences in mean quantitative measures using t-tests.

All analyses were conducted in R version 4.1.3. [19, 20] utilizing the following packages: arrow, broom, dplyr, forcats, ggplot2, knitr, lubridate, tidyr, and xtable.[21–29]

## Results

### Population

Overall, 48,146,511 patients in the subset of Truveta Data used for this study had encounters between January 2018 and June 2023, including 40,001,234 adults, 4,363,619 adults with T2D and 13,408,738 adults with overweight or obesity (T2D and overweight or obesity are not mutually exclusive). Of these, 12,186,837 adults satisfied the additional study criteria (at least one outpatient visit in each consecutive six-month period preceding an encounter and no Type I or gestational diabetes mellitus), of which, 2,463,012 (20.2%) had T2D, and 6,669,448 (54.7%) had overweight or obesity (Supplement, section 2).

The final study cohort included 274,562 adults who were first prescribed a GLP-1 RA during the analysis period, of which 181,860 (66.2%) had T2D and 229,715 (83.7%) had obesity or overweight. The mean (SD) patient age at baseline was 56.5 (13.5) years, with 51% of patients between 45 and 64 years. Overall, 63.8% of patients were female and 71.1% were white.

### Changes over Time

First-time GLP-1 RA prescribing volumes increased over the study period (Figure 2), with 9,642 new prescriptions between January and June 2018 and 66,569 new prescriptions between January and June 2023 (p < 0.001) (Supplement). Increases in GLP-1 RA prescribing were driven by higher prescribing of semaglutide and tirzepatide.

**Figure 2:**
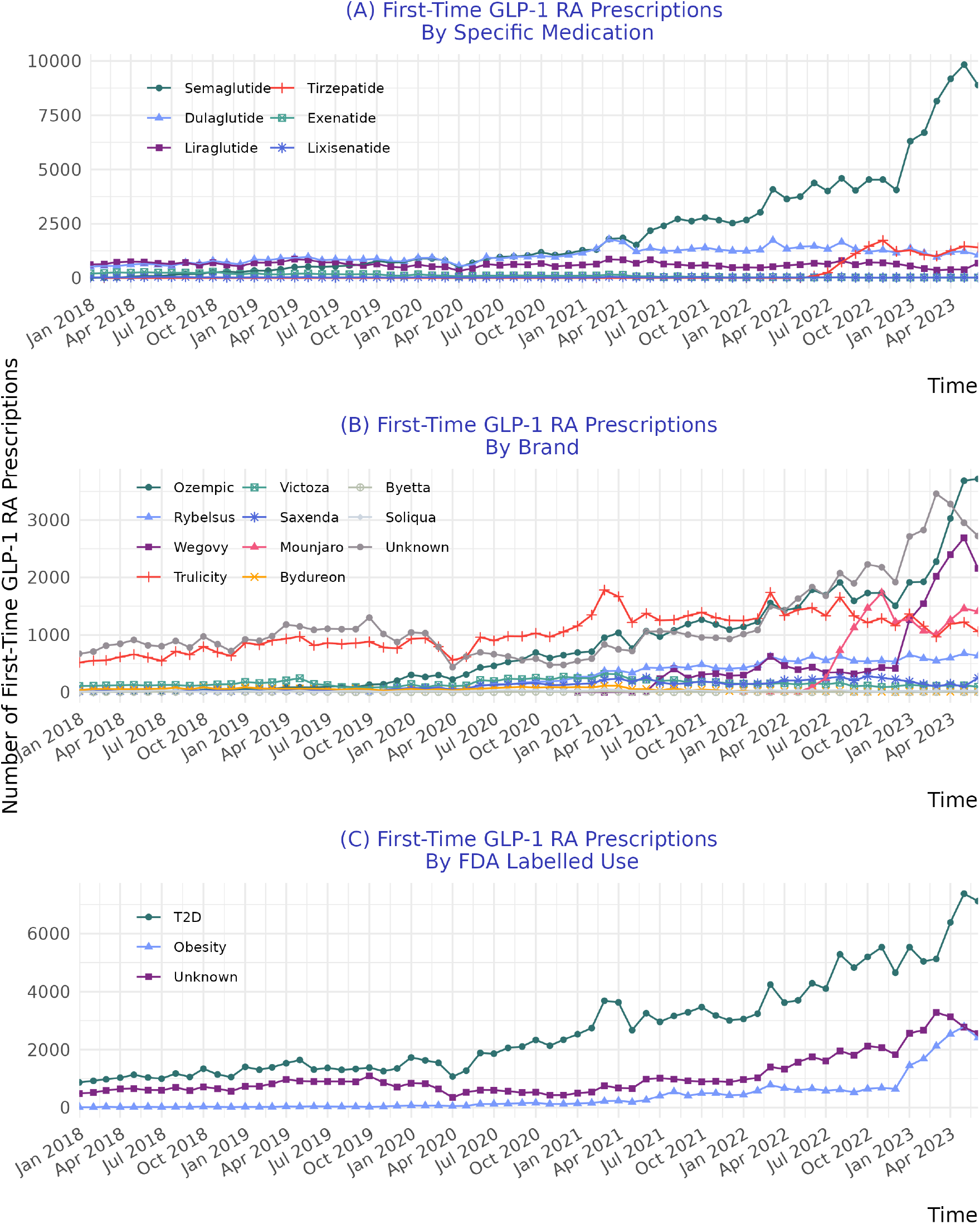
First-Time GLP-1 RA prescribing volume by (A) specific medication, (B) brand, and (C) FDA labelled use.

There were notable differences in characteristics of patients first prescribed a GLP-1 RA over time. Whereas the proportion of patients with T2D was higher during the early time period (84.1% [January to June 2018] versus 49% [January to June 2023]; p < 0.001), the proportion of patients with overweight or obesity was higher during the later time period (75.9% [January - June 2018] versus 90.7% [January - June 2023]; p <.001). During the later period, it was also noted that a smaller proportion of patients had an HbA1c measured prior to GLP-1 RA initiation (p <0.001), a smaller proportion of patients used other ADMs (p <0.001), and the mean HbA1c level was lower at baseline (p <0.001).

### Characteristics by Specific Medication

Semaglutide was the most commonly prescribed GLP-1 RA for all months between March 2021 and June 2023, representing 62.9% of new GLP-1 RA prescriptions during this period. Of the 141,275 first-time semaglutide prescriptions during the study period, a brand name was established for 89,417 (63.3%), of which 60.5% were for Ozempic, 18.5% were for Rybelsus, and 21% were for Wegovy. Patients prescribed Wegovy were more likely to be female (81.8%) and younger (mean age 48.5 years), compared to those prescribed Ozempic (62.9% female [p < 0.001]; mean age 57.5 years [p < 0.001]) or Rybelsus (56.2% female [p < 0.001]; mean age 60.5 years [p < 0.001]).

Tirzepatide, which was approved for treatment of T2D in May 2022, was prescribed to 14,120 patients. Of these, 47.3% had evidence of T2D, which was the lowest proportion amoung GLP-1 RAs (by medication type). Consistent with this, availability of a baseline HbA1c was lower for patients prescribed tirzepatide compared to those prescribed other GLP-1 RAs (62.6% versus 71.9%; p < 0.001), and among those with a baseline HbA1c available, the mean value was lower for those prescribed tirzepatide (mean [SD] 6.8 [1.6]) compared to other GLP-1 RAs (mean [sd] 7.8 [2]; p < 0.001). Finally, patients prescribed tirzepatide had a lower mean age (52.5 versus 56.7; p < 0.001) and were more likely to be female (71.6% versus 63.4%; p < 0.001).

When the analysis was restricted to the period after tirzepatide’s approval (June 2022 to June 2023), however, greater similarity was noted among those on other GLP-1 RAs, with younger mean ages, higher female proportion, and lesser evidence of T2D (Supplement).

**Table 1:**
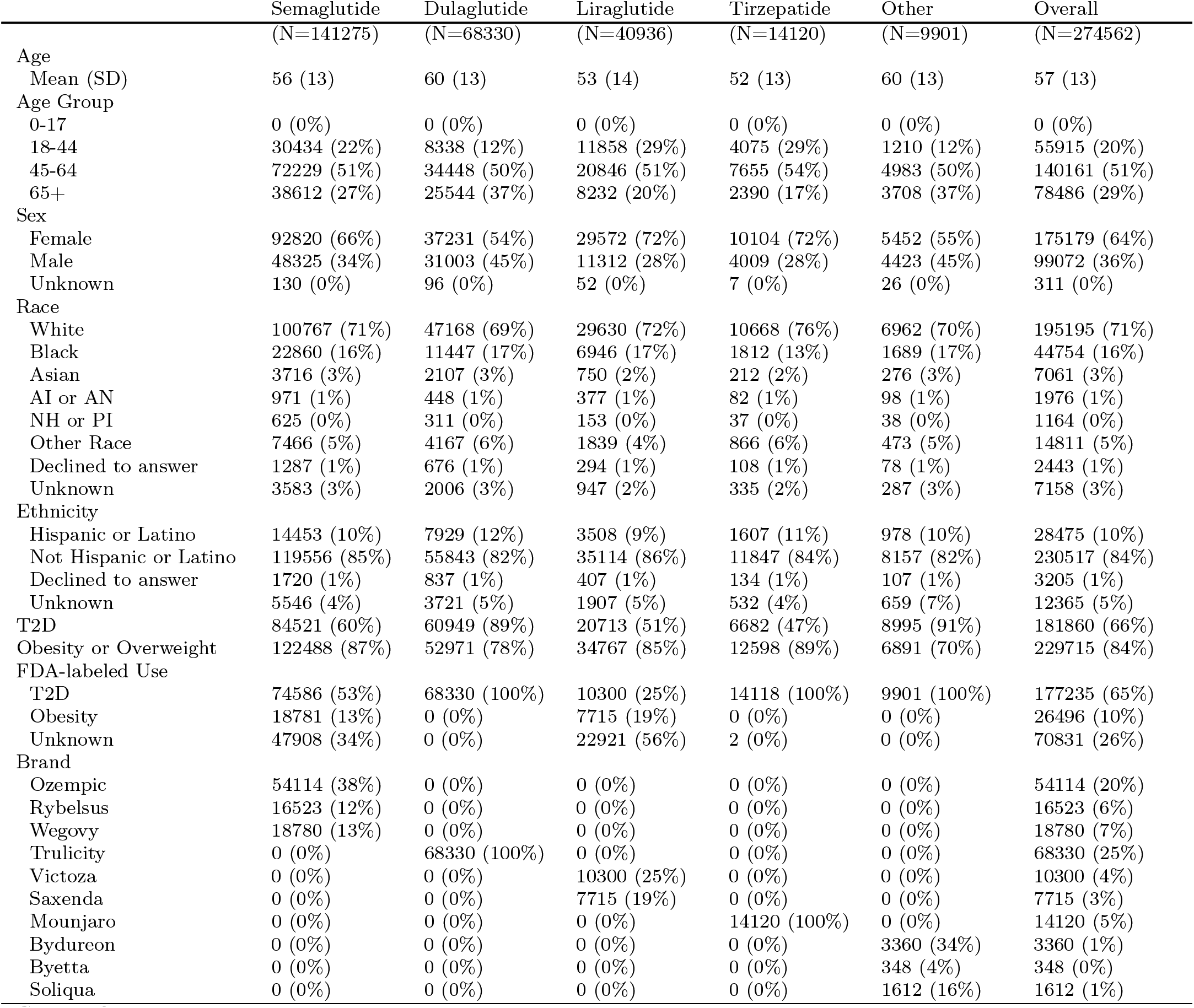
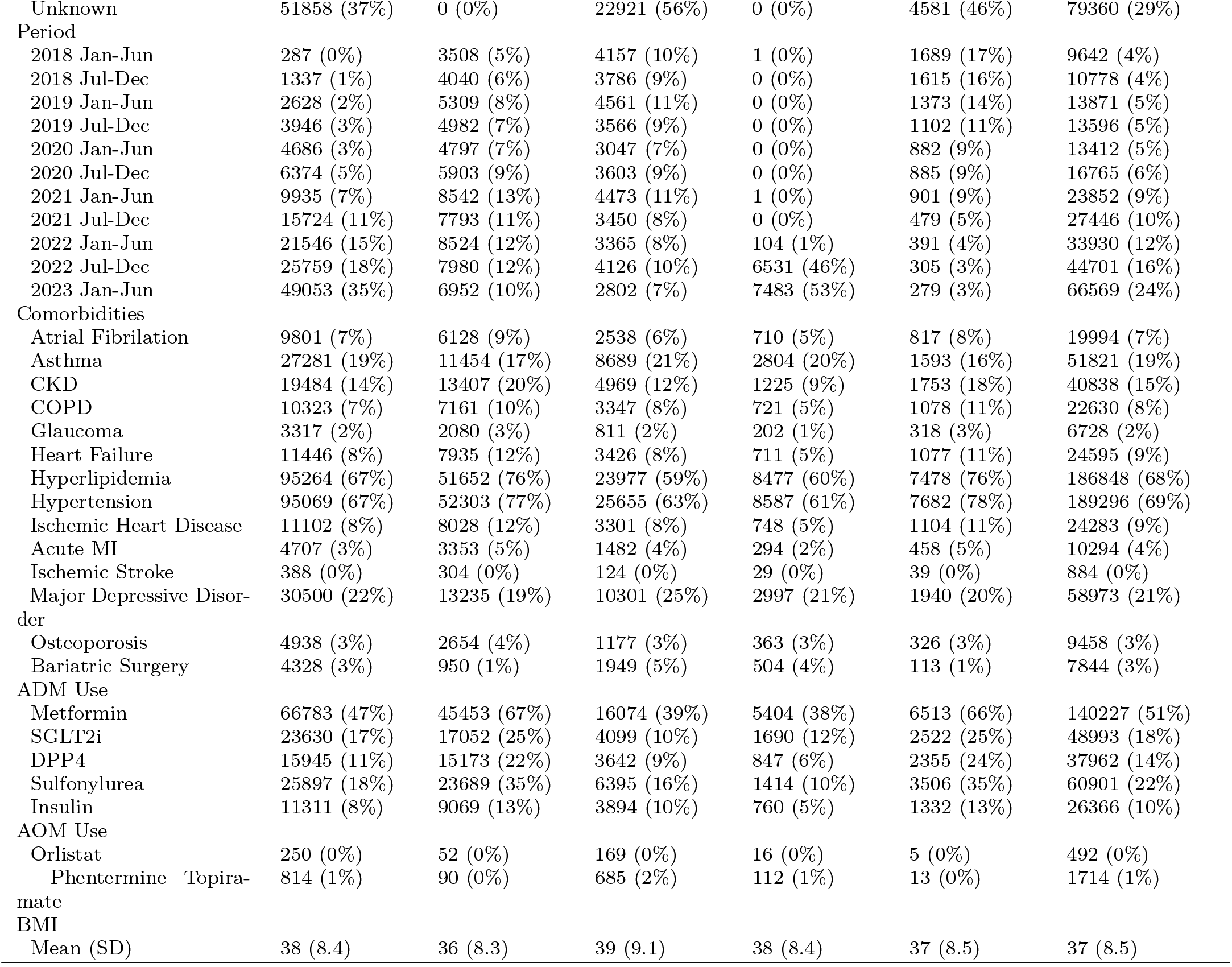
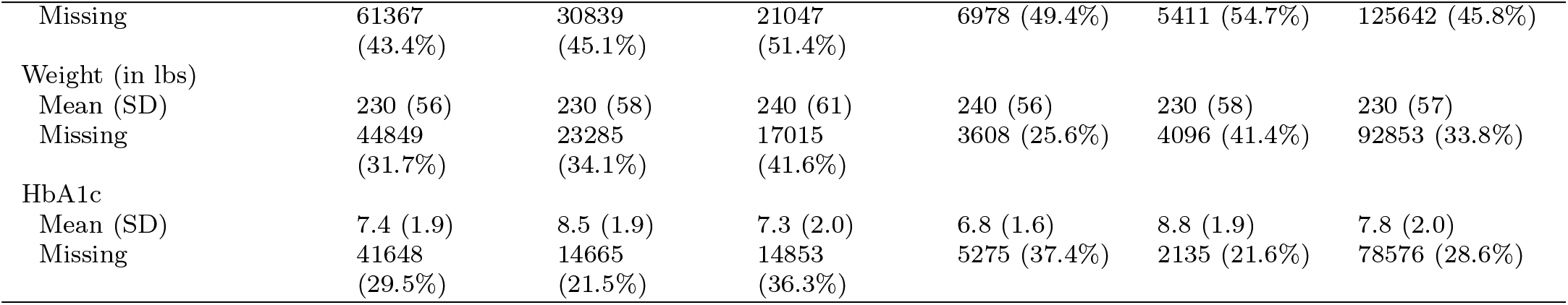
Characteristics of patients first prescribed a GLP-1 RA, by specific medication Other specific medication includes the two smallest groups: Exenatide and Lixisenatide.: ADM=anti-diabetic medication, AI = American Indian, AN = Alaska Native, AOM=anti-obesity medication, Black = Black or African American, BMI = body mass index, CKD=chronic kidney disease, COPD=chronic obstructive pulmonary disease, DPP4= dipeptidyl peptidase 4 inhibitor, FDA=Food and Drug Administration, HbA1c=hemoglobin A1C, MI=myocardial infarction, NH = Native Hawaiian, PI = Pacific Islander, SD=standard deviation, SGLT2i=sodium/glucose cotransporter-2 inhibitor, T2D=type 2 diabetes mellitus.

### On-label Use

FDA-labelled was established for 203,731 (74.2%) of first-time GLP-1 RA prescriptions. Of these, 177,235 were for medications labelled for T2D, with 137,762 (77.7%) prescribed on-label (to patients with T2D). On-label prescribing for this indication was lower between January and June 2023 (67.1% on-label), compared to January and June 2018 (91.2% on-label; p < 0.001) (Figure 3).

**Figure 3:**
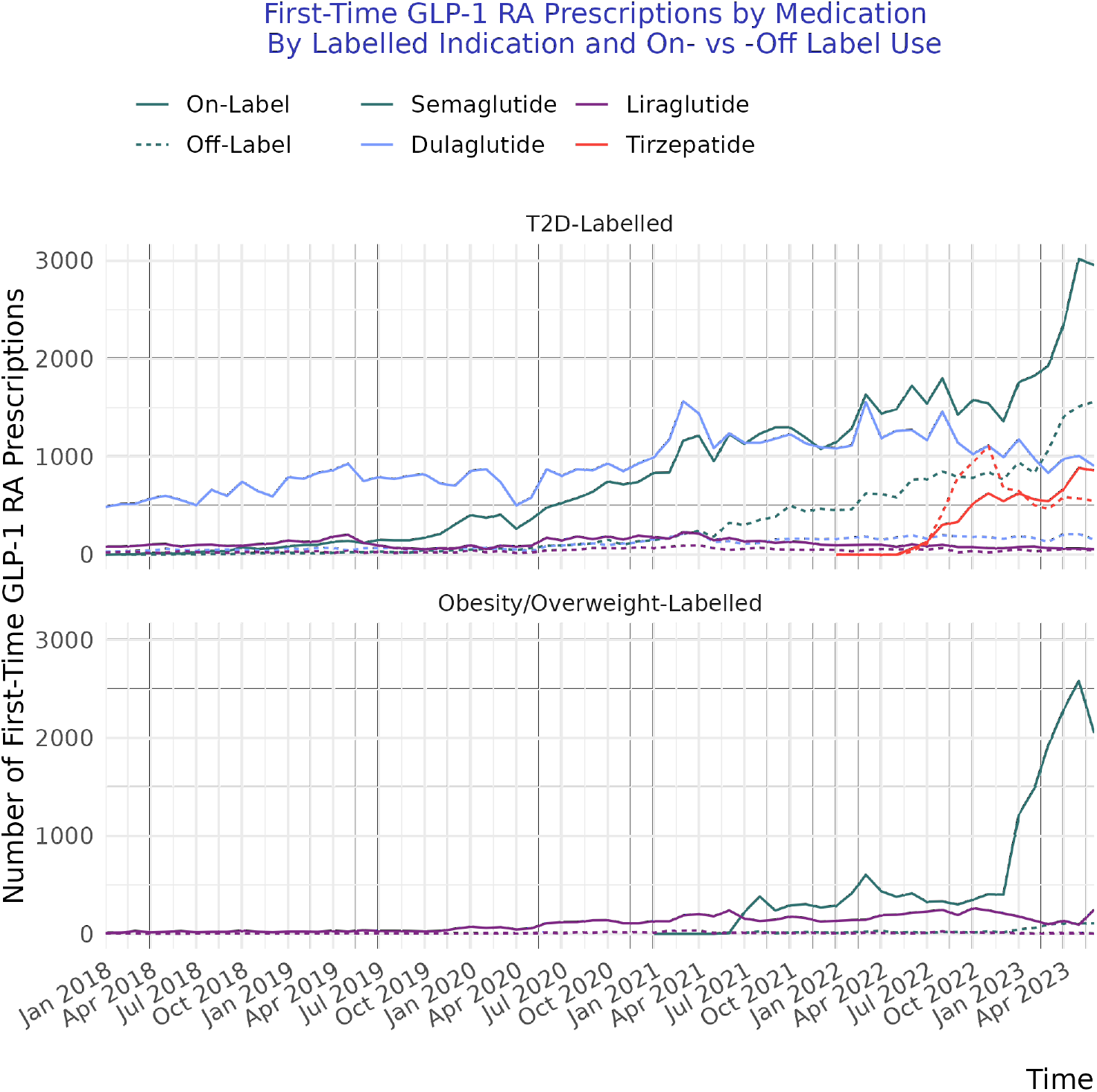
First-Time GLP-1 RA prescribing volume, stratified by labelled-use and on vs off-label for semaglutide, dulaglutide, liraglutide, and tirzepatide

The rate of on-label use was consistently higher for medications labelled for overweight and obesity. Of 26,496 first-time prescriptions labelled for obesity and overweight, 94% were prescribed on-label.

## Discussion

In this analysis, GLP-1 RA prescribing increased substantially over time, with 6.9-fold higher volume in the first 6 months of 2023 compared to the first 6 months of 2018. Over the same period, the population first-prescribed GLP-1 RAs shifted from one dominated by those with T2D (the majority of whom had comorbid overweight or obesity), to a mix of patients with and without T2D. Finally, GLP-1 RAs labelled for T2D were commonly used both on- and off-label, while GLP-1 RAs labelled for overweight or obesity were largely used on-label.

Consistent with age and sex characteristics from previous descriptive studies of GLP-1 RAs initiated for T2D [30, 31], patients first prescribed a GLP-1 RA labelled for T2D in our study were 59.1% female, with a mean age of 58.6 years. In contrast, those initiated on a GLP-1 RA labelled for overweight and obesity in our were predominately female (82.9%) with a mean age of 48.2 years. Previous studies on this topic have largely predated 2020 and have been limited to patients with T2D [30–32]. Such studies have also largely relied on pharmacy claims data, which may under-capture GLP-1 RAs prescribed for obesity due to limited insurance coverage for this population [33]. Given growing attention to GLP-1 RAs for weight loss, improved understanding of current prescribing trends in a real-world population is critically important.

Our study is subject to several limitations. First, while our study cohort from a group of US heathcare systems included 15 states with ≥ 1,000 patients, the prescribing patterns observed in our study may not be representative of the broader US. Second, we only captured care delivered at Truveta constituent heathcare systems; as such, any care delivered elsewhere would be missed. To reduce the risk of misclassification due to care received elsewhere, we restricted our analysis to patients receiving regular care at the heathcare system prior to their first GLP-1 RA prescription. It remains possible, however, that additional prescriptions were initiated elsewhere. Finally, we sought to describe first-time prescribing of GLP-1 RAs, which may differ from medication dispensing or medication use patterns. Patients prescribed a GLP-1 RA may not have filled or initiated the medication due to cost or other factors, or alternatively may have initiated a different GLP-1 RA medication than that initially prescribed because of stock-outs. GLP-1 RA dispensing patterns and barriers to access represent an important topic of future research given the high cost, limited insurance coverage of AOMs, and disproportionate prevalence of overweight and obesity by social drivers of health [34–36].

## Conclusions

Increased understanding of GLP-1 RA prescribing patterns is an important topic given the high prevalence and clinical impacts of T2D as well as overweight and obesity [35, 36]. Our findings document appreciable shifts in the clinical characteristics and demographics of those prescribed a GLP-1 RA in a large US population. Growth in GLP-1 RA prescribing was driven disproportionately by semaglutide and tirzepatide, with increased prescribing among younger, female patients for overweight or obesity.

## Data Availability

The data used in this study is available to all Truveta subscribers and may be accessed at studio.truveta.com.

