## Supplemental Material for "First-Time Prescribing of GLP-1 Receptor Agonists from 2018-2023: A Descriptive Analysis"

**4 Patient Characteristics by Brand** **11**
4.1 Patient Characteristics by Brand: Semaglutide and Liraglutide . . . 11
4.2 Patient Characteristics by Brand: Dulaglutide, Tirzepatide, and Other . . . 14

**5 Patient Characteristics by FDA-labelled Use** **17**

**6 Patient Characteristics by Specific Medication After May 2022** **19**

**7 Patient Characteristics for Ozempic, Wegovy, and Mounjaro After May 2022** **22**

**8 Data Definitions** **24**

### 1 Quantitative Measure Pre-processing

#### 1.1 Weight

Weight measures include both a numeric value and a unit of measure (ounce, pounds, kilogram, gram). When the unit of measure was missing but a numeric measure was present, we assumed weight values $> 1000$ were measured in ounces (the most common unit in Truveta Data), and weight values $< 125$ were measured in kilograms. Values between 125 and 1000 with a missing unit were treated as missing. We converted all weights to pounds and excluded and measures $<90$ or $>700$ pounds, as these were likely data entry errors.

#### 1.2 BMI

If multiple BMI values were present on the most recent measurement date in the 12 months before to one day after index, we used the mean of BMI values on that date as the baseline value, except when the standard deviation of BMI values on this date was $>1$. If the standard deviation was $>1$, we treated the baseline BMI value as missing. We also removed BMI values $<10$ or $> 100$, which were assumed to be entered in error.

#### 1.3 HbA1c

We removed HbA1c values of 0 or $>20$, which were assumed to be entered in error.

### 2 Volume of Patients with Encounters

#### 2.1 Patients with Encounters, by T2D and Overweight or Obesity

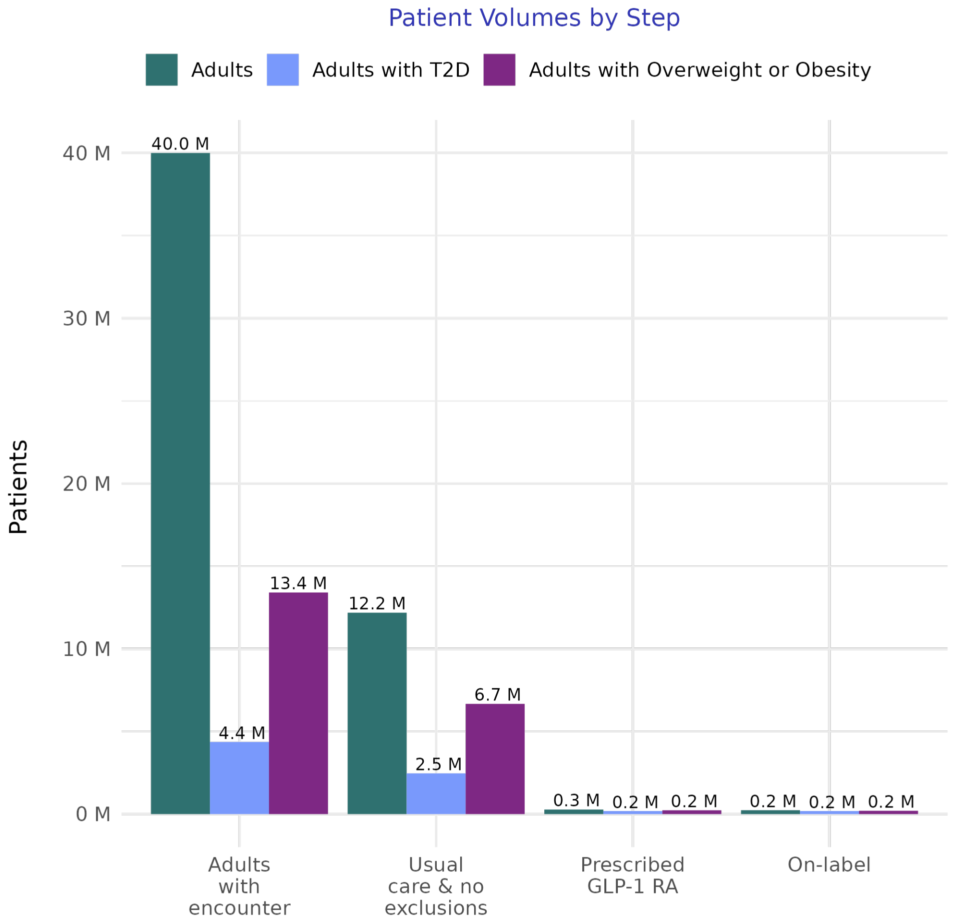

Figure 1: The number of patients in the subset of Truveta Data used for this study with an encounter, by presence of T2D and overweight or obesity. T2D and overweight or obesity are not mutually exclusive. Usual care indicates at least one outpatient encounter in each consecutive six-month period prior to the encounter. Exclusions refer to Type I and gestational diabetes mellitus. Abbreviations: T2D = Type II diabetes mellitus

#### 2.2 Proportion of Patients with Encounters by Mutually Exclusive Condition Group

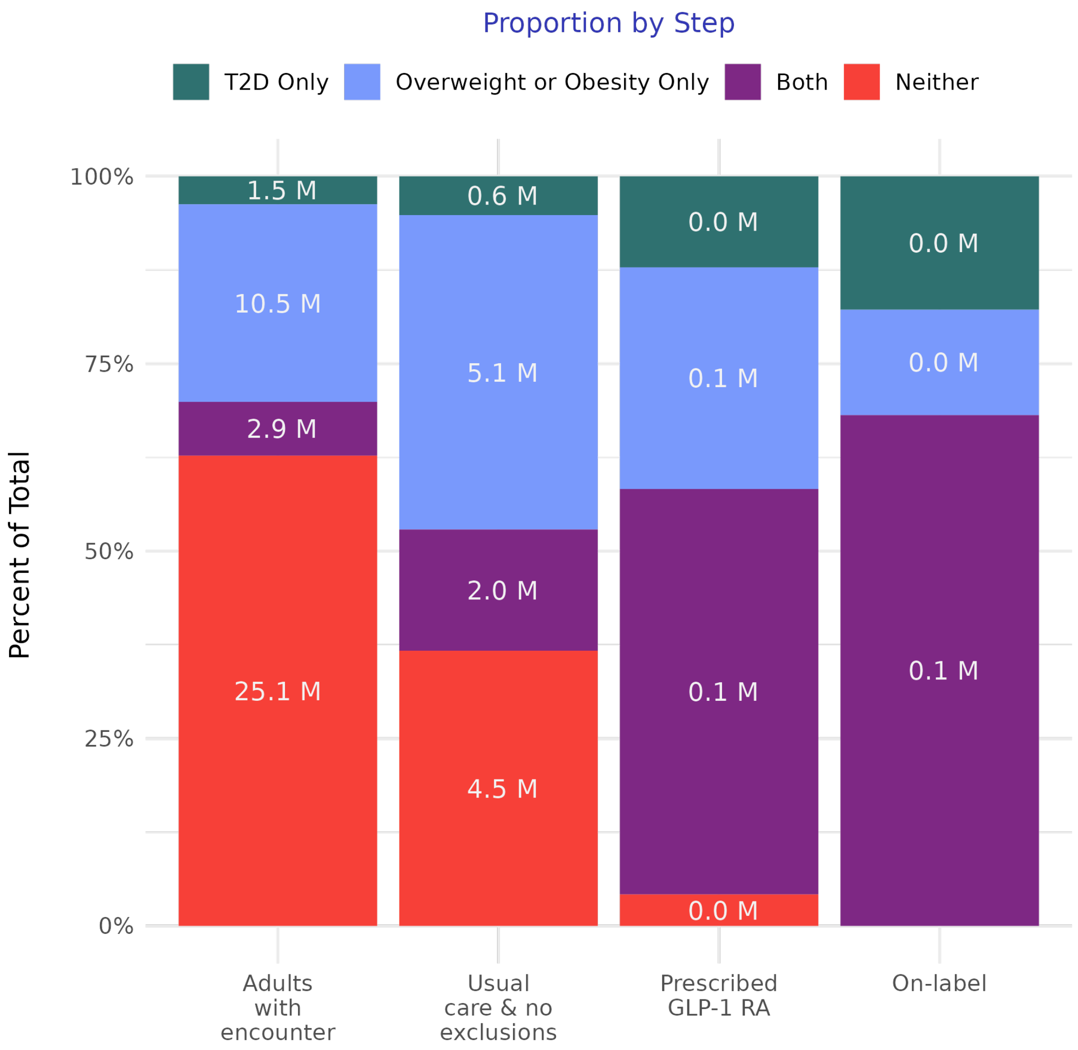

Figure 2: The number of patients in the subset of Truveta Data used for this study with an encounter, by week. Adult indicates patient was ≥18 at the time of the encounter. Presence of T2D and overweight/obesity are assessed in the 2 year window up to the encounter. Abbreviations: T2D = type II diabetes mellitus.

#### 2.3 Patients with Encounters over Time

The number of patients, by week, with an encounter in the subset of Truveta Data used for this study.

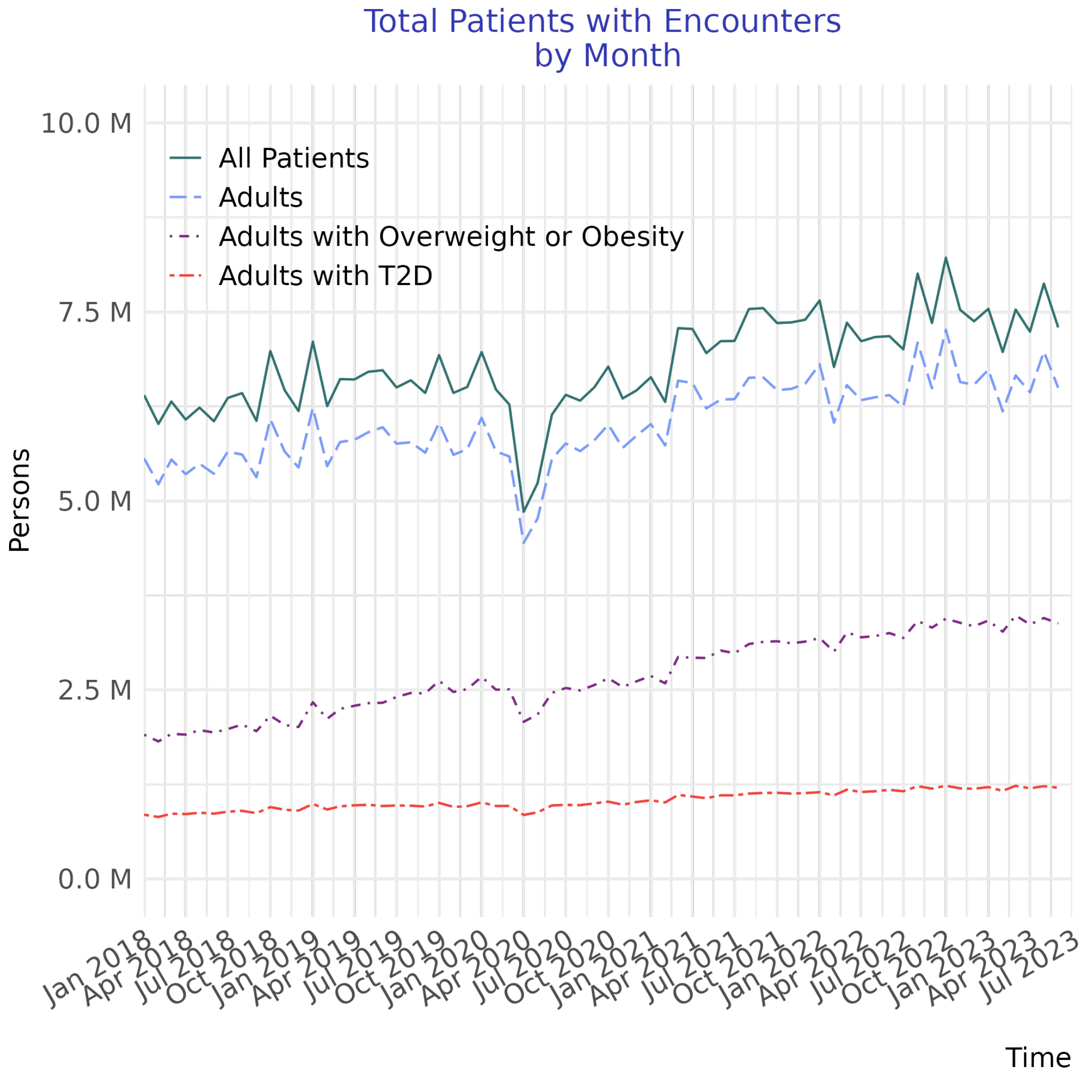

Figure 3: The number of patients in the subset of Truveta Data used for this study with an encounter, by month. Adult indicates patient was ≥18 at the time of the encounter. Presence of T2D and overweight/obesity are assessed in the 2 year window up to the encounter. T2D and overweight or obesity are not mutually exclusive. Abbreviations: T2D = type II diabetes mellitus.

### 3 Patient Characteristics by Period

We split each year into January-June and July-December to create 11 equal time intervals. The table is separated into 2018 - 2020 and 2021 -2023.

#### 3.1 Patient Characteristics by Period: 2018-2020

| | 2018 Jan-Jun (N=9642) | 2018 Jul-Dec (N=10778) | 2019 Jan-Jun (N=13871) | 2019 Jul-Dec (N=13596) | 2020 Jan-Jun (N=13412) | 2020 Jul-Dec (N=16765) |
|---|---|---|---|---|---|---|
| Age | | | | | | |
| Mean (SD) | 58 (12) | 58 (12) | 58 (13) | 58 (13) | 58 (13) | 58 (13) |
| Age Group | | | | | | |
| 0-17 | 0 (0%) | 0 (0%) | 0 (0%) | 0 (0%) | 0 (0%) | 0 (0%) |
| 18-44 | 1576 (16%) | 1616 (15%) | 2134 (15%) | 2163 (16%) | 2126 (16%) | 2625 (16%) |
| 45-64 | 5260 (55%) | 5886 (55%) | 7442 (54%) | 7180 (53%) | 7043 (53%) | 8682 (52%) |
| 65+ | 2806 (29%) | 3276 (30%) | 4295 (31%) | 4253 (31%) | 4243 (32%) | 5458 (33%) |
| Sex | | | | | | |
| Female | 5656 (59%) | 6313 (59%) | 7909 (57%) | 7838 (58%) | 7651 (57%) | 9763 (58%) |
| Male | 3959 (41%) | 4439 (41%) | 5934 (43%) | 5719 (42%) | 5737 (43%) | 6981 (42%) |
| Unknown | 27 (0%) | 26 (0%) | 28 (0%) | 39 (0%) | 24 (0%) | 21 (0%) |
| Race | | | | | | |
| White | 7142 (74%) | 7988 (74%) | 10041 (72%) | 9698 (71%) | 9500 (71%) | 11742 (70%) |
| Black | 1671 (17%) | 1699 (16%) | 2210 (16%) | 2341 (17%) | 2328 (17%) | 2736 (16%) |
| Asian | 135 (1%) | 233 (2%) | 364 (3%) | 316 (2%) | 374 (3%) | 461 (3%) |
| AI or AN | 63 (1%) | 73 (1%) | 101 (1%) | 84 (1%) | 87 (1%) | 130 (1%) |
| NH or PI | 30 (0%) | 34 (0%) | 50 (0%) | 49 (0%) | 53 (0%) | 70 (0%) |
| Other Race | 288 (3%) | 388 (4%) | 592 (4%) | 665 (5%) | 608 (5%) | 1028 (6%) |
| Declined to answer | 57 (1%) | 68 (1%) | 91 (1%) | 94 (1%) | 115 (1%) | 171 (1%) |
| Unknown | 256 (3%) | 295 (3%) | 422 (3%) | 349 (3%) | 347 (3%) | 427 (3%) |
| Ethnicity | | | | | | |
| Hispanic or Latino | 576 (6%) | 741 (7%) | 1070 (8%) | 1209 (9%) | 1200 (9%) | 1911 (11%) |
| Not Hispanic or Latino | 8327 (86%) | 9260 (86%) | 11770 (85%) | 11416 (84%) | 11294 (84%) | 13796 (82%) |
| Declined to answer | 84 (1%) | 105 (1%) | 161 (1%) | 159 (1%) | 152 (1%) | 203 (1%) |
| Unknown | 655 (7%) | 672 (6%) | 870 (6%) | 812 (6%) | 766 (6%) | 855 (5%) |
| T2D | 8106 (84%) | 9063 (84%) | 11605 (84%) | 11298 (83%) | 11074 (83%) | 13395 (80%) |
| Obesity or Overweight | 7318 (76%) | 8134 (75%) | 10424 (75%) | 10947 (81%) | 10558 (79%) | 12379 (74%) |
| FDA-labeled Use | | | | | | |
| T2D | 5977 (62%) | 6783 (63%) | 8589 (62%) | 8002 (59%) | 9142 (68%) | 12838 (77%) |
| Obesity | 145 (2%) | 165 (2%) | 202 (1%) | 233 (2%) | 467 (3%) | 839 (5%) |
| Unknown | 3520 (37%) | 3830 (36%) | 5080 (37%) | 5361 (39%) | 3803 (28%) | 3088 (18%) |
| Generic | | | | | | |
| Semaglutide | 287 (3%) | 1337 (12%) | 2628 (19%) | 3946 (29%) | 4686 (35%) | 6374 (38%) |
| Dulaglutide | 3508 (36%) | 4040 (37%) | 5309 (38%) | 4982 (37%) | 4797 (36%) | 5903 (35%) |
| Liraglutide | 4157 (43%) | 3786 (35%) | 4561 (33%) | 3566 (26%) | 3047 (23%) | 3603 (21%) |
| Tirzepatide | 1 (0%) | 0 (0%) | 0 (0%) | 0 (0%) | 0 (0%) | 0 (0%) |
| Exenatide | 1448 (15%) | 1382 (13%) | 1149 (8%) | 921 (7%) | 691 (5%) | 685 (4%) |
| Lixisenatide | 241 (2%) | 233 (2%) | 224 (2%) | 181 (1%) | 191 (1%) | 200 (1%) |
| Brand | | | | | | |
| Ozempic | 44 (0%) | 201 (2%) | 448 (3%) | 774 (6%) | 1837 (14%) | 3491 (21%) |

Continued on next page

**Table1 – continued from previous page**

| Feature | 2018 Jan-Jun | 2018 Jul-Dec | 2019 Jan-Jun | 2019 Jul-Dec | 2020 Jan-Jun | 2020 Jul-Dec |
|---|---|---|---|---|---|---|
| Rybelsus | 0 (0%) | 0 (0%) | 0 (0%) | 110 (1%) | 443 (3%) | 1019 (6%) |
| Wegovy | 0 (0%) | 0 (0%) | 0 (0%) | 0 (0%) | 0 (0%) | 0 (0%) |
| Trulicity | 3508 (36%) | 4040 (37%) | 5309 (38%) | 4982 (37%) | 4797 (36%) | 5903 (35%) |
| Victoza | 712 (7%) | 768 (7%) | 1103 (8%) | 568 (4%) | 790 (6%) | 1394 (8%) |
| Saxenda | 145 (2%) | 165 (2%) | 202 (1%) | 233 (2%) | 467 (3%) | 839 (5%) |
| Mounjaro | 1 (0%) | 0 (0%) | 0 (0%) | 0 (0%) | 0 (0%) | 0 (0%) |
| Bydureon | 326 (3%) | 406 (4%) | 419 (3%) | 295 (2%) | 312 (2%) | 512 (3%) |
| Byetta | 32 (0%) | 75 (1%) | 62 (0%) | 19 (0%) | 17 (0%) | 36 (0%) |
| Soliqua | 105 (1%) | 121 (1%) | 118 (1%) | 112 (1%) | 127 (1%) | 185 (1%) |
| Unknown | 4769 (49%) | 5002 (46%) | 6210 (45%) | 6503 (48%) | 4622 (34%) | 3386 (20%) |
| Comorbidities | | | | | | |
| Atrial Fibrilation | 736 (8%) | 853 (8%) | 1115 (8%) | 1144 (8%) | 1101 (8%) | 1386 (8%) |
| Asthma | 1605 (17%) | 1867 (17%) | 2383 (17%) | 2408 (18%) | 2319 (17%) | 2874 (17%) |
| CKD | 1434 (15%) | 1676 (16%) | 2182 (16%) | 2402 (18%) | 2443 (18%) | 2928 (17%) |
| COPD | 964 (10%) | 1068 (10%) | 1243 (9%) | 1352 (10%) | 1357 (10%) | 1543 (9%) |
| Glaucoma | 259 (3%) | 286 (3%) | 385 (3%) | 408 (3%) | 373 (3%) | 450 (3%) |
| Heart Failure | 944 (10%) | 1043 (10%) | 1349 (10%) | 1487 (11%) | 1502 (11%) | 1797 (11%) |
| Hyperlipidemia | 7310 (76%) | 8215 (76%) | 10470 (75%) | 10208 (75%) | 9913 (74%) | 11628 (69%) |
| Hypertension | 7499 (78%) | 8310 (77%) | 10627 (77%) | 10443 (77%) | 10045 (75%) | 11774 (70%) |
| Ischemic Heart Disease | 1084 (11%) | 1189 (11%) | 1528 (11%) | 1613 (12%) | 1559 (12%) | 1759 (10%) |
| Acute MI | 468 (5%) | 498 (5%) | 640 (5%) | 693 (5%) | 665 (5%) | 761 (5%) |
| Ischemic Stroke | 28 (0%) | 42 (0%) | 63 (0%) | 65 (0%) | 50 (0%) | 59 (0%) |
| Major Depressive Disorder | 1874 (19%) | 2177 (20%) | 2917 (21%) | 2846 (21%) | 2826 (21%) | 3211 (19%) |
| Osteoporosis | 301 (3%) | 347 (3%) | 471 (3%) | 470 (3%) | 429 (3%) | 582 (3%) |
| Bariatric Surgery | 180 (2%) | 219 (2%) | 286 (2%) | 274 (2%) | 298 (2%) | 336 (2%) |
| ADM Use | | | | | | |
| Metformin | 6134 (64%) | 6686 (62%) | 8655 (62%) | 8579 (63%) | 8428 (63%) | 9949 (59%) |
| SGLT2i | 2156 (22%) | 2294 (21%) | 2761 (20%) | 2917 (21%) | 2843 (21%) | 3382 (20%) |
| DPP4 | 2469 (26%) | 2521 (23%) | 3026 (22%) | 3041 (22%) | 2864 (21%) | 3079 (18%) |
| Sulfonylurea | 3268 (34%) | 3578 (33%) | 4387 (32%) | 4503 (33%) | 4352 (32%) | 4999 (30%) |
| Insulin | 1289 (13%) | 1320 (12%) | 1665 (12%) | 1627 (12%) | 1607 (12%) | 1938 (12%) |
| ADM Use | | | | | | |
| Orlistat | 12 (0%) | 12 (0%) | 19 (0%) | 20 (0%) | 23 (0%) | 24 (0%) |
| Phentermine Topiramate | 90 (1%) | 115 (1%) | 116 (1%) | 85 (1%) | 80 (1%) | 73 (0%) |
| BMI | | | | | | |
| Mean (SD) | 38 (8.2) | 37 (8.2) | 37 (8.4) | 37 (8.4) | 37 (8.4) | 37 (8.6) |
| Missing | 4812 (49.9%) | 5475 (50.8%) | 7540 (54.4%) | 6164 (45.3%) | 6154 (45.9%) | 8669 (51.7%) |
| Weight (in lbs) | | | | | | |
| Mean (SD) | 240 (57) | 230 (58) | 240 (58) | 230 (58) | 230 (59) | 240 (60) |
| Missing | 3270 (33.9%) | 3997 (37.1%) | 5781 (41.7%) | 4580 (33.7%) | 4943 (36.9%) | 7907 (47.2%) |
| HbA1c | | | | | | |
| Mean (SD) | 8.4 (1.9) | 8.4 (1.9) | 8.4 (1.9) | 8.4 (1.9) | 8.4 (2.0) | 8.3 (2.0) |
| Missing | 2232 (23.1%) | 2500 (23.2%) | 3057 (22.0%) | 3068 (22.6%) | 2974 (22.2%) | 3746 (22.3%) |

Table 1: Characteristics of patients initiating a GLP-1 RA, by period (part 1).: ADM=anti-diabetic medication, AI = American Indian, AN = Alaska Native, AOM=anti-obesity medication, Black = Black or African American, BMI = body mass index, CKD=chronic kidney disease, COPD=chronic obstructive pulmonary disease, DPP4= dipeptidyl peptidase 4 inhibitor, FDA=Food and Drug Administration, HbA1c=hemoglobin A1C, MI=myocardial infarction, NH = Native Hawaiian, PI = Pacific Islander, SD=standard deviation, SGLT2i=sodium/glucose cotransporter-2 inhibitor, T2D=type 2 diabetes mellitus.

#### 3.2 Patient Characteristics by Period: 2021-2023

| | 2021 Jan-Jun | 2021 Jul-Dec | 2022 Jan-Jun | 2022 Jul-Dec | 2023 Jan-Jun |
|---|---|---|---|---|---|
| | (N=23852) | (N=27446) | (N=33930) | (N=44701) | (N=66569) |
| Age | | | | | |
| Mean (SD) | 59 (13) | 57 (14) | 56 (14) | 55 (14) | 55 (14) |
| Age Group | | | | | |
| 0-17 | 0 (0%) | 0 (0%) | 0 (0%) | 0 (0%) | 0 (0%) |
| 18-44 | 3842 (16%) | 5333 (19%) | 7131 (21%) | 10763 (24%) | 16606 (25%) |
| 45-64 | 11807 (50%) | 13868 (51%) | 16928 (50%) | 22636 (51%) | 33429 (50%) |
| 65+ | 8203 (34%) | 8245 (30%) | 9871 (29%) | 11302 (25%) | 16534 (25%) |
| Sex | | | | | |
| Female | 13903 (58%) | 17366 (63%) | 21757 (64%) | 30638 (69%) | 46385 (70%) |
| Male | 9921 (42%) | 10046 (37%) | 12147 (36%) | 14033 (31%) | 20156 (30%) |
| Unknown | 28 (0%) | 34 (0%) | 26 (0%) | 30 (0%) | 28 (0%) |
| Race | | | | | |
| White | 16961 (71%) | 19049 (69%) | 23644 (70%) | 31997 (72%) | 47433 (71%) |
| Black | 3536 (15%) | 4614 (17%) | 5787 (17%) | 7390 (17%) | 10442 (16%) |
| Asian | 766 (3%) | 799 (3%) | 955 (3%) | 1051 (2%) | 1607 (2%) |
| AI or AN | 199 (1%) | 222 (1%) | 255 (1%) | 346 (1%) | 416 (1%) |
| NH or PI | 118 (0%) | 113 (0%) | 154 (0%) | 220 (0%) | 273 (0%) |
| Other Race | 1374 (6%) | 1712 (6%) | 1988 (6%) | 2343 (5%) | 3825 (6%) |
| Declined to answer | 351 (1%) | 236 (1%) | 285 (1%) | 335 (1%) | 640 (1%) |
| Unknown | 547 (2%) | 701 (3%) | 862 (3%) | 1019 (2%) | 1933 (3%) |
| Ethnicity | | | | | |
| Hispanic or Latino | 2948 (12%) | 2946 (11%) | 3772 (11%) | 4828 (11%) | 7274 (11%) |
| Not Hispanic or Latino | 19460 (82%) | 23025 (84%) | 28388 (84%) | 37782 (85%) | 55999 (84%) |
| Declined to answer | 343 (1%) | 339 (1%) | 364 (1%) | 443 (1%) | 852 (1%) |
| Unknown | 1101 (5%) | 1136 (4%) | 1406 (4%) | 1648 (4%) | 2444 (4%) |
| T2D | 18410 (77%) | 18876 (69%) | 22096 (65%) | 25325 (57%) | 32612 (49%) |
| Obesity or Overweight | 17662 (74%) | 23343 (85%) | 29128 (86%) | 39428 (88%) | 60394 (91%) |
| FDA-labeled Use | | | | | |
| T2D | 18519 (78%) | 19058 (69%) | 22142 (65%) | 29602 (66%) | 36583 (55%) |
| Obesity | 1215 (5%) | 2782 (10%) | 3740 (11%) | 3708 (8%) | 13000 (20%) |
| Unknown | 4118 (17%) | 5606 (20%) | 8048 (24%) | 11391 (25%) | 16986 (26%) |
| Generic | | | | | |
| Semaglutide | 9935 (42%) | 15724 (57%) | 21546 (64%) | 25759 (58%) | 49053 (74%) |
| Dulaglutide | 8542 (36%) | 7793 (28%) | 8524 (25%) | 7980 (18%) | 6952 (10%) |
| Liraglutide | 4473 (19%) | 3450 (13%) | 3365 (10%) | 4126 (9%) | 2802 (4%) |
| Tirzepatide | 1 (0%) | 0 (0%) | 104 (0%) | 6531 (15%) | 7483 (11%) |
| Exenatide | 671 (3%) | 301 (1%) | 203 (1%) | 141 (0%) | 113 (0%) |
| Lixisenatide | 230 (1%) | 178 (1%) | 188 (1%) | 164 (0%) | 166 (0%) |
| Brand | | | | | |
| Ozempic | 5218 (22%) | 6776 (25%) | 8611 (25%) | 10168 (23%) | 16546 (25%) |
| Rybelsus | 2058 (9%) | 2610 (10%) | 3236 (10%) | 3349 (7%) | 3698 (6%) |
| Wegovy | 15 (0%) | 1809 (7%) | 2653 (8%) | 2229 (5%) | 12074 (18%) |
| Trulicity | 8542 (36%) | 7793 (28%) | 8524 (25%) | 7980 (18%) | 6952 (10%) |
| Victoza | 1558 (7%) | 1100 (4%) | 891 (3%) | 721 (2%) | 695 (1%) |
| Saxenda | 1200 (5%) | 972 (4%) | 1087 (3%) | 1479 (3%) | 926 (1%) |
| Mounjaro | 1 (0%) | 0 (0%) | 104 (0%) | 6531 (15%) | 7483 (11%) |
| Bydureon | 516 (2%) | 241 (1%) | 153 (0%) | 96 (0%) | 84 (0%) |
| Byetta | 47 (0%) | 18 (0%) | 16 (0%) | 15 (0%) | 11 (0%) |
| Soliqua | 209 (1%) | 168 (1%) | 171 (1%) | 150 (0%) | 146 (0%) |
| Unknown | 4488 (19%) | 5959 (22%) | 8484 (25%) | 11983 (27%) | 17954 (27%) |
| Comorbidities | | | | | |

Continued on next page

**Table2 – continued from previous page**

| Feature | 2021 Jan-Jun | 2021 Jul-Dec | 2022 Jan-Jun | 2022 Jul-Dec | 2023 Jan-Jun |
|---|---|---|---|---|---|
| Atrial Fibrilation | 1914 (8%) | 2092 (8%) | 2460 (7%) | 2993 (7%) | 4200 (6%) |
| Asthma | 4023 (17%) | 5110 (19%) | 6515 (19%) | 9060 (20%) | 13657 (21%) |
| CKD | 4202 (18%) | 4327 (16%) | 5248 (15%) | 6143 (14%) | 7853 (12%) |
| COPD | 2160 (9%) | 2284 (8%) | 2713 (8%) | 3325 (7%) | 4621 (7%) |
| Glaucoma | 646 (3%) | 649 (2%) | 850 (3%) | 1016 (2%) | 1406 (2%) |
| Heart Failure | 2406 (10%) | 2585 (9%) | 3121 (9%) | 3542 (8%) | 4819 (7%) |
| Hyperlipidemia | 15853 (66%) | 18497 (67%) | 22871 (67%) | 29511 (66%) | 42372 (64%) |
| Hypertension | 16245 (68%) | 18842 (69%) | 23262 (69%) | 29815 (67%) | 42434 (64%) |
| Ischemic Heart Disease | 2278 (10%) | 2451 (9%) | 3006 (9%) | 3294 (7%) | 4522 (7%) |
| Acute MI | 932 (4%) | 1047 (4%) | 1317 (4%) | 1413 (3%) | 1860 (3%) |
| Ischemic Stroke | 94 (0%) | 89 (0%) | 105 (0%) | 120 (0%) | 169 (0%) |
| Major Depressive Disorder | 4740 (20%) | 5904 (22%) | 7487 (22%) | 10137 (23%) | 14854 (22%) |
| Osteoporosis | 820 (3%) | 950 (3%) | 1205 (4%) | 1540 (3%) | 2343 (4%) |
| Bariatric Surgery | 518 (2%) | 805 (3%) | 1076 (3%) | 1451 (3%) | 2401 (4%) |
| ADM Use | | | | | |
| Metformin | 14250 (60%) | 14798 (54%) | 17374 (51%) | 19701 (44%) | 25673 (39%) |
| SGLT2i | 5122 (21%) | 5120 (19%) | 6230 (18%) | 7166 (16%) | 9002 (14%) |
| DPP4 | 3795 (16%) | 4014 (15%) | 4283 (13%) | 4122 (9%) | 4748 (7%) |
| Sulfonylurea | 6602 (28%) | 6530 (24%) | 7185 (21%) | 7116 (16%) | 8381 (13%) |
| Insulin | 2539 (11%) | 2811 (10%) | 3272 (10%) | 3583 (8%) | 4715 (7%) |
| ADM Use | | | | | |
| Orlistat | 46 (0%) | 64 (0%) | 69 (0%) | 85 (0%) | 118 (0%) |
| Phentermine Topiramate | 126 (1%) | 168 (1%) | 216 (1%) | 249 (1%) | 396 (1%) |
| BMI | | | | | |
| Mean (SD) | 37 (8.7) | 37 (8.7) | 38 (8.8) | 38 (8.6) | 38 (8.3) |
| Missing | 12845 (53.9%) | 11586 (42.2%) | 14288 (42.1%) | 20301 (45.4%) | 27808 (41.8%) |
| Weight (in lbs) | | | | | |
| Mean (SD) | 240 (59) | 240 (60) | 230 (58) | 230 (56) | 230 (55) |
| Missing | 11135 (46.7%) | 11334 (41.3%) | 11497 (33.9%) | 10854 (24.3%) | 17555 (26.4%) |
| HbA1c | | | | | |
| Mean (SD) | 8.2 (1.9) | 7.9 (2.0) | 7.7 (2.0) | 7.3 (1.9) | 7.0 (1.8) |
| Missing | 4885 (20.5%) | 7519 (27.4%) | 9885 (29.1%) | 14334 (32.1%) | 24376 (36.6%) |

Table 2: Characteristics of patients initiating a GLP-1 RA, by period (part 2).: ADM=anti-diabetic medication, AI = American Indian, AN = Alaska Native, AOM=anti-obesity medication, Black = Black or African American, BMI = body mass index, CKD=chronic kidney disease, COPD=chronic obstructive pulmonary disease, DPP4= dipeptidyl peptidase 4 inhibitor, FDA=Food and Drug Administration, HbA1c=hemoglobin A1C, MI=myocardial infarction, NH = Native Hawaiian, PI = Pacific Islander, SD=standard deviation, SGLT2i=sodium/glucose cotransporter-2 inhibitor, T2D=type 2 diabetes mellitus.

### 4 Patient Characteristics by Brand

#### 4.1 Patient Characteristics by Brand: Semaglutide and Liraglutide

| | Semaglutide | | | Liraglutide | | |
|---|---|---|---|---|---|---|
| | Ozempic | Rybelsus | Wegovy | Victoza | Saxenda | Unknown |
| | (N=54114) | (N=16523) | (N=18780) | (N=10300) | (N=7715) | (N=74779) |
| Age | | | | | | |
| Mean (SD) | 58 (13) | 60 (13) | 48 (12) | 58 (13) | 47 (12) | 55 (14) |
| Age Group | | | | | | |
| 0-17 | 0 (0%) | 0 (0%) | 0 (0%) | 0 (0%) | 0 (0%) | 0 (0%) |
| 18-44 | 9428 (17%) | 2036 (12%) | 7321 (39%) | 1737 (17%) | 3209 (42%) | 18561 (25%) |
| 45-64 | 27879 (52%) | 8034 (49%) | 9770 (52%) | 5233 (51%) | 4049 (52%) | 38110 (51%) |
| 65+ | 16807 (31%) | 6453 (39%) | 1689 (9%) | 3330 (32%) | 457 (6%) | 18108 (24%) |
| Sex | | | | | | |
| Female | 34044 (63%) | 9288 (56%) | 15360 (82%) | 6455 (63%) | 6597 (86%) | 50648 (68%) |
| Male | 20013 (37%) | 7220 (44%) | 3416 (18%) | 3831 (37%) | 1114 (14%) | 24043 (32%) |
| Unknown | 57 (0%) | 15 (0%) | 4 (0%) | 14 (0%) | 4 (0%) | 88 (0%) |
| Race | | | | | | |
| White | 38832 (72%) | 10881 (66%) | 13529 (72%) | 7206 (70%) | 5492 (71%) | 54457 (73%) |
| Black or African American | 9085 (17%) | 2915 (18%) | 3294 (18%) | 1454 (14%) | 1545 (20%) | 11513 (15%) |
| Asian | 1223 (2%) | 766 (5%) | 246 (1%) | 306 (3%) | 69 (1%) | 1856 (2%) |
| American Indian or Alaska Native | 330 (1%) | 98 (1%) | 88 (0%) | 133 (1%) | 43 (1%) | 656 (1%) |
| Native Hawaiian or Other Pacific Islander | 184 (0%) | 92 (1%) | 54 (0%) | 51 (0%) | 15 (0%) | 382 (1%) |
| Other Race | 2560 (5%) | 1062 (6%) | 957 (5%) | 715 (7%) | 332 (4%) | 3679 (5%) |
| Declined to answer | 523 (1%) | 178 (1%) | 184 (1%) | 115 (1%) | 70 (1%) | 511 (1%) |
| Unknown | 1377 (3%) | 531 (3%) | 428 (2%) | 320 (3%) | 149 (2%) | 1725 (2%) |
| Ethnicity | | | | | | |
| Hispanic or Latino | 5210 (10%) | 1716 (10%) | 1811 (10%) | 1023 (10%) | 753 (10%) | 7448 (10%) |
| Not Hispanic or Latino | 46054 (85%) | 13834 (84%) | 16120 (86%) | 8410 (82%) | 6617 (86%) | 63635 (85%) |
| Declined to answer | 696 (1%) | 235 (1%) | 241 (1%) | 168 (2%) | 85 (1%) | 702 (1%) |
| Unknown | 2154 (4%) | 738 (4%) | 608 (3%) | 699 (7%) | 260 (3%) | 2994 (4%) |
| T2D | 36628 (68%) | 13765 (83%) | 1270 (7%) | 7431 (72%) | 811 (11%) | 45329 (61%) |

Continued on next page

**Table3 – continued from previous page**

| | Semaglutide | | | Liraglutide | | |
|---|---|---|---|---|---|---|
| Feature | Ozempic | Rybelsus | Wegovy | Victoza | Saxenda | Unknown |
| Obesity or Overweight | 43967 (81%) | 13743 (83%) | 17913 (95%) | 7107 (69%) | 7004 (91%) | 67521 (90%) |
| Period | | | | | | |
| 2018 Jan-Jun | 44 (0%) | 0 (0%) | 0 (0%) | 712 (7%) | 145 (2%) | 3543 (5%) |
| 2018 Jul-Dec | 201 (0%) | 0 (0%) | 0 (0%) | 768 (7%) | 165 (2%) | 3989 (5%) |
| 2019 Jan-Jun | 448 (1%) | 0 (0%) | 0 (0%) | 1103 (11%) | 202 (3%) | 5436 (7%) |
| 2019 Jul-Dec | 774 (1%) | 110 (1%) | 0 (0%) | 568 (6%) | 233 (3%) | 5827 (8%) |
| 2020 Jan-Jun | 1837 (3%) | 443 (3%) | 0 (0%) | 790 (8%) | 467 (6%) | 4196 (6%) |
| 2020 Jul-Dec | 3491 (6%) | 1019 (6%) | 0 (0%) | 1394 (14%) | 839 (11%) | 3234 (4%) |
| 2021 Jan-Jun | 5218 (10%) | 2058 (12%) | 15 (0%) | 1558 (15%) | 1200 (16%) | 4359 (6%) |
| 2021 Jul-Dec | 6776 (13%) | 2610 (16%) | 1809 (10%) | 1100 (11%) | 972 (13%) | 5907 (8%) |
| 2022 Jan-Jun | 8611 (16%) | 3236 (20%) | 2653 (14%) | 891 (9%) | 1087 (14%) | 8433 (11%) |
| 2022 Jul-Dec | 10168 (19%) | 3349 (20%) | 2229 (12%) | 721 (7%) | 1479 (19%) | 11939 (16%) |
| 2023 Jan-Jun | 16546 (31%) | 3698 (22%) | 12074 (64%) | 695 (7%) | 926 (12%) | 17916 (24%) |
| Comorbidities | | | | | | |
| Atrial Fibrilation | 4244 (8%) | 1351 (8%) | 533 (3%) | 856 (8%) | 206 (3%) | 5149 (7%) |
| Asthma | 9890 (18%) | 2682 (16%) | 3942 (21%) | 1993 (19%) | 1577 (20%) | 15886 (21%) |
| CKD | 7990 (15%) | 3023 (18%) | 732 (4%) | 1754 (17%) | 284 (4%) | 10670 (14%) |
| COPD | 4357 (8%) | 1360 (8%) | 616 (3%) | 1004 (10%) | 253 (3%) | 6080 (8%) |
| Glaucoma | 1277 (2%) | 501 (3%) | 171 (1%) | 236 (2%) | 79 (1%) | 1864 (2%) |
| Heart Failure | 5065 (9%) | 1457 (9%) | 512 (3%) | 1211 (12%) | 197 (3%) | 6430 (9%) |
| Hyperlipidemia | 37022 (68%) | 12641 (77%) | 9072 (48%) | 6340 (62%) | 3245 (42%) | 50921 (68%) |
| Hypertension | 37630 (70%) | 12398 (75%) | 8940 (48%) | 6790 (66%) | 3535 (46%) | 51431 (69%) |
| Ischemic Heart Disease | 4878 (9%) | 1463 (9%) | 499 (3%) | 1107 (11%) | 206 (3%) | 6250 (8%) |
| Acute MI | 2095 (4%) | 550 (3%) | 217 (1%) | 461 (4%) | 71 (1%) | 2795 (4%) |
| Ischemic Stroke | 161 (0%) | 54 (0%) | 24 (0%) | 41 (0%) | 10 (0%) | 222 (0%) |
| Major Depressive Disorder | 10045 (19%) | 2834 (17%) | 4459 (24%) | 2057 (20%) | 1857 (24%) | 19549 (26%) |
| Osteoporosis | 1921 (4%) | 696 (4%) | 402 (2%) | 321 (3%) | 160 (2%) | 2615 (3%) |
| Bariatric Surgery | 1404 (3%) | 196 (1%) | 1065 (6%) | 264 (3%) | 589 (8%) | 2759 (4%) |
| ADM Use | | | | | | |
| Metformin | 27483 (51%) | 11015 (67%) | 1879 (10%) | 5147 (50%) | 1045 (14%) | 36288 (49%) |
| SGLT2i | 10346 (19%) | 4734 (29%) | 163 (1%) | 1615 (16%) | 89 (1%) | 10782 (14%) |
| DPP4 | 6528 (12%) | 3309 (20%) | 57 (0%) | 1068 (10%) | 54 (1%) | 8571 (11%) |
| Sulfonylurea | 10136 (19%) | 5351 (32%) | 78 (0%) | 2023 (20%) | 71 (1%) | 14633 (20%) |
| Insulin | 4716 (9%) | 1275 (8%) | 385 (2%) | 906 (9%) | 264 (3%) | 7659 (10%) |
| ADM Use | | | | | | |
| Orlistat | 64 (0%) | 17 (0%) | 47 (0%) | 17 (0%) | 37 (0%) | 237 (0%) |
| Phentermine Topiramate | 203 (0%) | 35 (0%) | 296 (2%) | 26 (0%) | 194 (3%) | 745 (1%) |

Continued on next page

**Table3 – continued from previous page**

| | Semaglutide | | | Liraglutide | | |
|---|---|---|---|---|---|---|
| Feature | Ozempic | Rybelsus | Wegovy | Victoza | Saxenda | Unknown |
| BMI | | | | | | |
| Mean (SD) | 37 (8.4) | 35 (8.1) | 39 (8.0) | 39 (9.3) | 39 (8.6) | 39 (8.6) |
| Missing | 27779 (51.3%) | 6636 (40.2%) | 9785 (52.1%) | 5735 (55.7%) | 4598 (59.6%) | 27881 (37.3%) |
| Weight (in lbs) | | | | | | |
| Mean (SD) | 240 (57) | 220 (56) | 230 (54) | 240 (60) | 240 (58) | 240 (57) |
| Missing | 18977 (35.1%) | 5746 (34.8%) | 6377 (34.0%) | 5320 (51.7%) | 3085 (40.0%) | 22359 (29.9%) |
| HbA1c | | | | | | |
| Mean (SD) | 7.6 (1.9) | 8.1 (1.8) | 5.6 (0.55) | 7.9 (1.9) | 5.7 (0.80) | 7.5 (2.0) |
| Missing | 17222 (31.8%) | 3446 (20.9%) | 9366 (49.9%) | 3568 (34.6%) | 4044 (52.4%) | 18855 (25.2%) |

Table 3: Characteristics of patients first prescribed a GLP-1 RA, by brand for Semaglutide and Liraglutide.: ADM=anti-diabetic medication, AI = American Indian, AN = Alaska Native, AOM=anti-obesity medication, Black = Black or African American, BMI = body mass index, CKD=chronic kidney disease, COPD=chronic obstructive pulmonary disease, DPP4= dipeptidyl peptidase 4 inhibitor, FDA=Food and Drug Administration, HbA1c=hemoglobin A1C, MI=myocardial infarction, NH = Native Hawaiian, PI = Pacific Islander, SD=standard deviation, SGLT2i=sodium/glucose cotransporter-2 inhibitor, T2D=type 2 diabetes mellitus.

#### 4.2 Patient Characteristics by Brand: Dulaglutide, Tirzepatide, and Other

| | Dulaglutide | Tirzepatide | Exenatide | | Lixisenatide | |
|---|---|---|---|---|---|---|
| | Trulicity | Mounjaro | Bydureon | Byetta | Soliqua | Unknown |
| | (N=68330) | (N=14120) | (N=3360) | (N=348) | (N=1612) | (N=4581) |
| Age | | | | | | |
| Mean (SD) | 60 (13) | 52 (13) | 60 (13) | 62 (12) | 65 (12) | 58 (12) |
| Age Group | | | | | | |
| 0-17 | 0 (0%) | 0 (0%) | 0 (0%) | 0 (0%) | 0 (0%) | 0 (0%) |
| 18-44 | 8338 (12%) | 4075 (29%) | 421 (13%) | 33 (9%) | 110 (7%) | 646 (14%) |
| 45-64 | 34448 (50%) | 7655 (54%) | 1655 (49%) | 179 (51%) | 666 (41%) | 2483 (54%) |
| 65+ | 25544 (37%) | 2390 (17%) | 1284 (38%) | 136 (39%) | 836 (52%) | 1452 (32%) |
| Sex | | | | | | |
| Female | 37231 (54%) | 10104 (72%) | 1841 (55%) | 208 (60%) | 830 (51%) | 2573 (56%) |
| Male | 31003 (45%) | 4009 (28%) | 1510 (45%) | 140 (40%) | 781 (48%) | 1992 (43%) |
| Unknown | 96 (0%) | 7 (0%) | 9 (0%) | 0 (0%) | 1 (0%) | 16 (0%) |
| Race | | | | | | |
| White | 47168 (69%) | 10668 (76%) | 2344 (70%) | 262 (75%) | 1050 (65%) | 3306 (72%) |
| Black | 11447 (17%) | 1812 (13%) | 482 (14%) | 31 (9%) | 380 (24%) | 796 (17%) |
| Asian | 2107 (3%) | 212 (2%) | 111 (3%) | 9 (3%) | 38 (2%) | 118 (3%) |
| AI or AN | 448 (1%) | 82 (1%) | 33 (1%) | 4 (1%) | 6 (0%) | 55 (1%) |
| NH or PI | 311 (0%) | 37 (0%) | 19 (1%) | 1 (0%) | 5 (0%) | 13 (0%) |
| Other Race | 4167 (6%) | 866 (6%) | 215 (6%) | 24 (7%) | 72 (4%) | 162 (4%) |
| Declined to answer | 676 (1%) | 108 (1%) | 40 (1%) | 2 (1%) | 17 (1%) | 19 (0%) |
| Unknown | 2006 (3%) | 335 (2%) | 116 (3%) | 15 (4%) | 44 (3%) | 112 (2%) |
| Ethnicity | | | | | | |
| Hispanic or Latino | 7929 (12%) | 1607 (11%) | 483 (14%) | 41 (12%) | 182 (11%) | 272 (6%) |
| Not Hispanic or Latino | 55843 (82%) | 11847 (84%) | 2592 (77%) | 286 (82%) | 1305 (81%) | 3974 (87%) |
| Declined to answer | 837 (1%) | 134 (1%) | 48 (1%) | 2 (1%) | 19 (1%) | 38 (1%) |
| Unknown | 3721 (5%) | 532 (4%) | 237 (7%) | 19 (5%) | 106 (7%) | 297 (6%) |
| T2D | 60949 (89%) | 6682 (47%) | 2879 (86%) | 282 (81%) | 1461 (91%) | 4373 (95%) |
| Obesity or Overweight | 52971 (78%) | 12598 (89%) | 1985 (59%) | 209 (60%) | 903 (56%) | 3794 (83%) |
| Period | | | | | | |
| 2018 Jan-Jun | 3508 (5%) | 1 (0%) | 326 (10%) | 32 (9%) | 105 (7%) | 1226 (27%) |
| 2018 Jul-Dec | 4040 (6%) | 0 (0%) | 406 (12%) | 75 (22%) | 121 (8%) | 1013 (22%) |
| 2019 Jan-Jun | 5309 (8%) | 0 (0%) | 419 (12%) | 62 (18%) | 118 (7%) | 774 (17%) |
| 2019 Jul-Dec | 4982 (7%) | 0 (0%) | 295 (9%) | 19 (5%) | 112 (7%) | 676 (15%) |
| 2020 Jan-Jun | 4797 (7%) | 0 (0%) | 312 (9%) | 17 (5%) | 127 (8%) | 426 (9%) |

Continued on next page

**Table4 – continued from previous page**

| Dulaglutide | Tirzepatide | | Exenatide | Lixisenatide | | |
|---|---|---|---|---|---|---|
| Feature | Trulicity | Mounjaro | Bydureon | Byetta | Soliqua | Unknown |
| 2020 Jul-Dec | 5903 (9%) | 0 (0%) | 512 (15%) | 36 (10%) | 185 (11%) | 152 (3%) |
| 2021 Jan-Jun | 8542 (13%) | 1 (0%) | 516 (15%) | 47 (14%) | 209 (13%) | 129 (3%) |
| 2021 Jul-Dec | 7793 (11%) | 0 (0%) | 241 (7%) | 18 (5%) | 168 (10%) | 52 (1%) |
| 2022 Jan-Jun | 8524 (12%) | 104 (1%) | 153 (5%) | 16 (5%) | 171 (11%) | 51 (1%) |
| 2022 Jul-Dec | 7980 (12%) | 6531 (46%) | 96 (3%) | 15 (4%) | 150 (9%) | 44 (1%) |
| 2023 Jan-Jun | 6952 (10%) | 7483 (53%) | 84 (2%) | 11 (3%) | 146 (9%) | 38 (1%) |
| Comorbidities | | | | | | |
| Atrial Fibrilation | 6128 (9%) | 710 (5%) | 251 (7%) | 25 (7%) | 156 (10%) | 385 (8%) |
| Asthma | 11454 (17%) | 2804 (20%) | 518 (15%) | 52 (15%) | 201 (12%) | 822 (18%) |
| CKD | 13407 (20%) | 1225 (9%) | 518 (15%) | 44 (13%) | 420 (26%) | 771 (17%) |
| COPD | 7161 (10%) | 721 (5%) | 297 (9%) | 27 (8%) | 175 (11%) | 579 (13%) |
| Glaucoma | 2080 (3%) | 202 (1%) | 106 (3%) | 9 (3%) | 67 (4%) | 136 (3%) |
| Heart Failure | 7935 (12%) | 711 (5%) | 310 (9%) | 35 (10%) | 211 (13%) | 521 (11%) |
| Hyperlipidemia | 51652 (76%) | 8477 (60%) | 2290 (68%) | 228 (66%) | 1235 (77%) | 3725 (81%) |
| Hypertension | 52303 (77%) | 8587 (61%) | 2357 (70%) | 229 (66%) | 1284 (80%) | 3812 (83%) |
| Ischemic Heart Disease | 8028 (12%) | 748 (5%) | 313 (9%) | 35 (10%) | 201 (12%) | 555 (12%) |
| Acute MI | 3353 (5%) | 294 (2%) | 135 (4%) | 13 (4%) | 82 (5%) | 228 (5%) |
| Ischemic Stroke | 304 (0%) | 29 (0%) | 8 (0%) | 2 (1%) | 8 (0%) | 21 (0%) |
| Major Depressive Disorder | 13235 (19%) | 2997 (21%) | 499 (15%) | 54 (16%) | 272 (17%) | 1115 (24%) |
| Osteoporosis | 2654 (4%) | 363 (3%) | 106 (3%) | 8 (2%) | 61 (4%) | 151 (3%) |
| Bariatric Surgery | 950 (1%) | 504 (4%) | 34 (1%) | 11 (3%) | 2 (0%) | 66 (1%) |
| ADM Use | | | | | | |
| Metformin | 45453 (67%) | 5404 (38%) | 1953 (58%) | 187 (54%) | 970 (60%) | 3403 (74%) |
| SGLT2i | 17052 (25%) | 1690 (12%) | 860 (26%) | 53 (15%) | 436 (27%) | 1173 (26%) |
| DPP4 | 15173 (22%) | 847 (6%) | 659 (20%) | 41 (12%) | 342 (21%) | 1313 (29%) |
| Sulfonylurea | 23689 (35%) | 1414 (10%) | 943 (28%) | 78 (22%) | 535 (33%) | 1950 (43%) |
| Insulin | 9069 (13%) | 760 (5%) | 356 (11%) | 25 (7%) | 276 (17%) | 675 (15%) |
| ADM Use | | | | | | |
| Orlistat | 52 (0%) | 16 (0%) | 1 (0%) | 1 (0%) | 1 (0%) | 2 (0%) |
| Phentermine Topiramate | 90 (0%) | 112 (1%) | 3 (0%) | 0 (0%) | 3 (0%) | 7 (0%) |
| BMI | | | | | | |
| Mean (SD) | 36 (8.3) | 38 (8.4) | 36 (8.7) | 38 (9.4) | 33 (7.3) | 37 (8.4) |
| Missing | 30839 (45.1%) | 6978 (49.4%) | 2221 (66.1%) | 215 (61.8%) | 1211 (75.1%) | 1764 (38.5%) |
| Weight (in lbs) | | | | | | |
| Mean (SD) | 230 (58) | 240 (56) | 230 (58) | 230 (61) | 210 (54) | 230 (58) |
| Missing | 23285 (34.1%) | 3608 (25.6%) | 1628 (48.5%) | 229 (65.8%) | 600 (37.2%) | 1639 (35.8%) |
| HbA1c | | | | | | |
| Mean (SD) | 8.5 (1.9) | 6.8 (1.6) | 8.5 (1.9) | 7.9 (1.7) | 9.3 (2.0) | 8.8 (1.9) |

Continued on next page

**Table4 – continued from previous page**

| Dulaglutide | Tirzepatide | | Exenatide | Lixisenatide | | |
|---|---|---|---|---|---|---|
| Feature | Trulicity | Mounjaro | Bydureon | Byetta | Soliqua | Unknown |
| Missing | 14665 (21.5%) | 5275 (37.4%) | 823 (24.5%) | 95 (27.3%) | 390 (24.2%) | 827 (18.1%) |

Table 4: Characteristics of patients first prescribed a GLP-1 RA, by brand for Dulaglutide, Tirzepatide, and Other.: ADM=anti-diabetic medication, AI = American Indian, AN = Alaska Native, AOM=anti-obesity medication, Black = Black or African American, BMI = body mass index, CKD=chronic kidney disease, COPD=chronic obstructive pulmonary disease, DPP4= dipeptidyl peptidase 4 inhibitor, FDA=Food and Drug Administration, HbA1c=hemoglobin A1C, MI=myocardial infarction, NH = Native Hawaiian, PI = Pacific Islander, SD=standard deviation, SGLT2i=sodium/glucose cotransporter-2 inhibitor, T2D=type 2 diabetes mellitus.

### 5 Patient Characteristics by FDA-labelled Use

| | T2D | Obesity | Unknown | Overall |
|---|---|---|---|---|
| | (N=177235) | (N=26496) | (N=70831) | (N=274562) |
| Age | | | | |
| Mean (SD) | 59 (13) | 48 (12) | 54 (14) | 57 (13) |
| Age Group | | | | |
| 0-17 | 0 (0%) | 0 (0%) | 0 (0%) | 0 (0%) |
| 18-44 | 27371 (15%) | 10530 (40%) | 18014 (25%) | 55915 (20%) |
| 45-64 | 90339 (51%) | 13819 (52%) | 36003 (51%) | 140161 (51%) |
| 65+ | 59525 (34%) | 2147 (8%) | 16814 (24%) | 78486 (29%) |
| Sex | | | | |
| Female | 104675 (59%) | 21958 (83%) | 48546 (69%) | 175179 (64%) |
| Male | 72344 (41%) | 4530 (17%) | 22198 (31%) | 99072 (36%) |
| Unknown | 216 (0%) | 8 (0%) | 87 (0%) | 311 (0%) |
| Race | | | | |
| White | 124561 (70%) | 19022 (72%) | 51612 (73%) | 195195 (71%) |
| Black | 29186 (16%) | 4839 (18%) | 10729 (15%) | 44754 (16%) |
| Asian | 5060 (3%) | 315 (1%) | 1686 (2%) | 7061 (3%) |
| AI or AN | 1208 (1%) | 131 (0%) | 637 (1%) | 1976 (1%) |
| NH or PI | 722 (0%) | 69 (0%) | 373 (1%) | 1164 (0%) |
| Other Race | 9886 (6%) | 1289 (5%) | 3636 (5%) | 14811 (5%) |
| Declined to answer | 1712 (1%) | 254 (1%) | 477 (1%) | 2443 (1%) |
| Unknown | 4900 (3%) | 577 (2%) | 1681 (2%) | 7158 (3%) |
| Ethnicity | | | | |
| Hispanic or Latino | 19132 (11%) | 2564 (10%) | 6779 (10%) | 28475 (10%) |
| Not Hispanic or Latino | 147324 (83%) | 22738 (86%) | 60455 (85%) | 230517 (84%) |
| Declined to answer | 2205 (1%) | 326 (1%) | 674 (1%) | 3205 (1%) |
| Unknown | 8574 (5%) | 868 (3%) | 2923 (4%) | 12365 (5%) |
| T2D | 137762 (78%) | 2081 (8%) | 42017 (59%) | 181860 (66%) |
| Obesity or Overweight | 139572 (79%) | 24918 (94%) | 65225 (92%) | 229715 (84%) |
| Generic | | | | |
| Semaglutide | 74586 (42%) | 18781 (71%) | 47908 (68%) | 141275 (51%) |
| Dulaglutide | 68330 (39%) | 0 (0%) | 0 (0%) | 68330 (25%) |
| Liraglutide | 10300 (6%) | 7715 (29%) | 22921 (32%) | 40936 (15%) |
| Tirzepatide | 14118 (8%) | 0 (0%) | 2 (0%) | 14120 (5%) |
| Exenatide | 7705 (4%) | 0 (0%) | 0 (0%) | 7705 (3%) |
| Lixisenatide | 2196 (1%) | 0 (0%) | 0 (0%) | 2196 (1%) |
| Brand | | | | |
| Ozempic | 54114 (31%) | 0 (0%) | 0 (0%) | 54114 (20%) |
| Rybelsus | 16523 (9%) | 0 (0%) | 0 (0%) | 16523 (6%) |
| Wegovy | 0 (0%) | 18780 (71%) | 0 (0%) | 18780 (7%) |
| Trulicity | 68330 (39%) | 0 (0%) | 0 (0%) | 68330 (25%) |
| Victoza | 10300 (6%) | 0 (0%) | 0 (0%) | 10300 (4%) |
| Saxenda | 0 (0%) | 7715 (29%) | 0 (0%) | 7715 (3%) |
| Mounjaro | 14118 (8%) | 0 (0%) | 2 (0%) | 14120 (5%) |
| Bydureon | 3360 (2%) | 0 (0%) | 0 (0%) | 3360 (1%) |
| Byetta | 348 (0%) | 0 (0%) | 0 (0%) | 348 (0%) |
| Soliqua | 1612 (1%) | 0 (0%) | 0 (0%) | 1612 (1%) |
| Unknown | 8530 (5%) | 1 (0%) | 70829 (100%) | 79360 (29%) |
| Period | | | | |
| 2018 Jan-Jun | 5977 (3%) | 145 (1%) | 3520 (5%) | 9642 (4%) |
| 2018 Jul-Dec | 6783 (4%) | 165 (1%) | 3830 (5%) | 10778 (4%) |
| 2019 Jan-Jun | 8589 (5%) | 202 (1%) | 5080 (7%) | 13871 (5%) |
| 2019 Jul-Dec | 8002 (5%) | 233 (1%) | 5361 (8%) | 13596 (5%) |

Continued on next page

**Table5 – continued from previous page**

| Feature | Type II Diabetes | Obesity | Unknown | Overall |
|---|---|---|---|---|
| 2020 Jan-Jun | 9142 (5%) | 467 (2%) | 3803 (5%) | 13412 (5%) |
| 2020 Jul-Dec | 12838 (7%) | 839 (3%) | 3088 (4%) | 16765 (6%) |
| 2021 Jan-Jun | 18519 (10%) | 1215 (5%) | 4118 (6%) | 23852 (9%) |
| 2021 Jul-Dec | 19058 (11%) | 2782 (10%) | 5606 (8%) | 27446 (10%) |
| 2022 Jan-Jun | 22142 (12%) | 3740 (14%) | 8048 (11%) | 33930 (12%) |
| 2022 Jul-Dec | 29602 (17%) | 3708 (14%) | 11391 (16%) | 44701 (16%) |
| 2023 Jan-Jun | 36583 (21%) | 13000 (49%) | 16986 (24%) | 66569 (24%) |
| Comorbidities | | | | |
| Atrial Fibrilation | 14367 (8%) | 739 (3%) | 4888 (7%) | 19994 (7%) |
| Asthma | 31008 (17%) | 5519 (21%) | 15294 (22%) | 51821 (19%) |
| CKD | 29709 (17%) | 1016 (4%) | 10113 (14%) | 40838 (15%) |
| COPD | 15919 (9%) | 869 (3%) | 5842 (8%) | 22630 (8%) |
| Glaucoma | 4740 (3%) | 250 (1%) | 1738 (2%) | 6728 (2%) |
| Heart Failure | 17742 (10%) | 709 (3%) | 6144 (9%) | 24595 (9%) |
| Hyperlipidemia | 126932 (72%) | 12318 (46%) | 47598 (67%) | 186848 (68%) |
| Hypertension | 128547 (73%) | 12476 (47%) | 48273 (68%) | 189296 (69%) |
| Ischemic Heart Disease | 17664 (10%) | 705 (3%) | 5914 (8%) | 24283 (9%) |
| Acute MI | 7365 (4%) | 288 (1%) | 2641 (4%) | 10294 (4%) |
| Ischemic Stroke | 635 (0%) | 34 (0%) | 215 (0%) | 884 (0%) |
| Major Depressive Disorder | 33656 (19%) | 6316 (24%) | 19001 (27%) | 58973 (21%) |
| Osteoporosis | 6446 (4%) | 562 (2%) | 2450 (3%) | 9458 (3%) |
| Bariatric Surgery | 3475 (2%) | 1654 (6%) | 2715 (4%) | 7844 (3%) |
| ADM Use | | | | |
| Metformin | 103666 (58%) | 2924 (11%) | 33637 (47%) | 140227 (51%) |
| SGLT2i | 39207 (22%) | 252 (1%) | 9534 (13%) | 48993 (18%) |
| DPP4 | 30225 (17%) | 111 (0%) | 7626 (11%) | 37962 (14%) |
| Sulfonylurea | 47165 (27%) | 149 (1%) | 13587 (19%) | 60901 (22%) |
| Insulin | 18256 (10%) | 649 (2%) | 7461 (11%) | 26366 (10%) |
| ADM Use | | | | |
| Orlistat | 173 (0%) | 84 (0%) | 235 (0%) | 492 (0%) |
| Phentermine Topiramate | 491 (0%) | 490 (2%) | 733 (1%) | 1714 (1%) |
| BMI | | | | |
| Mean (SD) | 37 (8.4) | 39 (8.2) | 39 (8.6) | 37 (8.5) |
| Missing | 87114 (49.2%) | 14383 (54.3%) | 24145 (34.1%) | 125642 (45.8%) |
| Weight (in lbs) | | | | |
| Mean (SD) | 230 (57) | 240 (55) | 240 (57) | 230 (57) |
| Missing | 61101 (34.5%) | 9463 (35.7%) | 22289 (31.5%) | 92853 (33.8%) |
| HbA1c | | | | |
| Mean (SD) | 8.1 (1.9) | 5.7 (0.63) | 7.4 (2.0) | 7.8 (2.0) |
| Missing | 46789 (26.4%) | 13410 (50.6%) | 18377 (25.9%) | 78576 (28.6%) |

Table 5: Characteristics of patients first prescribed a GLP-1 RA, by FDA-labeled use.: ADM=anti-diabetic medication, AI = American Indian, AN = Alaska Native, AOM=anti-obesity medication, Black = Black or African American, BMI = body mass index, CKD=chronic kidney disease, COPD=chronic obstructive pulmonary disease, DPP4= dipeptidyl peptidase 4 inhibitor, FDA=Food and Drug Administration, HbA1c=hemoglobin A1C, MI=myocardial infarction, NH = Native Hawaiian, PI = Pacific Islander, SD=standard deviation, SGLT2i=sodium/glucose cotransporter-2 inhibitor, T2D=type 2 diabetes mellitus.

### 6 Patient Characteristics by Specific Medication After May 2022

Characteristics for patients first-prescribed a GLP-1 during the post-Tirzepatide approval period.

| | Semaglutide | Dulaglutide | Liraglutide | Tirzepatide | Other | Overall |
|---|---|---|---|---|---|---|
| | (N=79189) | (N=16403) | (N=7610) | (N=14110) | (N=650) | (N=117962) |
| Age | | | | | | |
| Mean (SD) | 55 (14) | 60 (13) | 49 (14) | 52 (13) | 65 (14) | 55 (14) |
| Age Group | | | | | | |
| 0-17 | 0 (0%) | 0 (0%) | 0 (0%) | 0 (0%) | 0 (0%) | 0 (0%) |
| 18-44 | 19485 (25%) | 2200 (13%) | 2921 (38%) | 4069 (29%) | 58 (9%) | 28733 (24%) |
| 45-64 | 39919 (50%) | 7937 (48%) | 3673 (48%) | 7653 (54%) | 237 (36%) | 59419 (50%) |
| 65+ | 19785 (25%) | 6266 (38%) | 1016 (13%) | 2388 (17%) | 355 (55%) | 29810 (25%) |
| Sex | | | | | | |
| Female | 55400 (70%) | 9339 (57%) | 6176 (81%) | 10100 (72%) | 346 (53%) | 81361 (69%) |
| Male | 23748 (30%) | 7054 (43%) | 1428 (19%) | 4003 (28%) | 303 (47%) | 36536 (31%) |
| Unknown | 41 (0%) | 10 (0%) | 6 (0%) | 7 (0%) | 1 (0%) | 65 (0%) |
| Race | | | | | | |
| White | 56761 (72%) | 11015 (67%) | 5322 (70%) | 10660 (76%) | 460 (71%) | 84218 (71%) |
| Black | 12508 (16%) | 3061 (19%) | 1446 (19%) | 1811 (13%) | 103 (16%) | 18929 (16%) |
| Asian | 2007 (3%) | 495 (3%) | 110 (1%) | 211 (1%) | 14 (2%) | 2837 (2%) |
| AI or AN | 530 (1%) | 126 (1%) | 76 (1%) | 82 (1%) | 7 (1%) | 821 (1%) |
| NH or PI | 359 (0%) | 89 (1%) | 36 (0%) | 37 (0%) | 4 (1%) | 525 (0%) |
| Other Race | 4187 (5%) | 993 (6%) | 398 (5%) | 866 (6%) | 32 (5%) | 6476 (5%) |
| Declined to answer | 726 (1%) | 144 (1%) | 44 (1%) | 108 (1%) | 4 (1%) | 1026 (1%) |
| Unknown | 2111 (3%) | 480 (3%) | 178 (2%) | 335 (2%) | 26 (4%) | 3130 (3%) |
| Ethnicity | | | | | | |
| Hispanic or Latino | 8272 (10%) | 2014 (12%) | 820 (11%) | 1607 (11%) | 64 (10%) | 12777 (11%) |
| Not Hispanic or Latino | 67103 (85%) | 13508 (82%) | 6479 (85%) | 11837 (84%) | 543 (84%) | 99470 (84%) |
| Declined to answer | 955 (1%) | 196 (1%) | 64 (1%) | 134 (1%) | 7 (1%) | 1356 (1%) |
| Unknown | 2859 (4%) | 685 (4%) | 247 (3%) | 532 (4%) | 36 (6%) | 4359 (4%) |
| T2D | 38701 (49%) | 14066 (86%) | 2200 (29%) | 6677 (47%) | 559 (86%) | 62203 (53%) |
| Obesity or Overweight | 71431 (90%) | 13812 (84%) | 7113 (93%) | 12589 (89%) | 489 (75%) | 105434 (89%) |
| FDA-labeled Use | | | | | | |
| T2D | 37728 (48%) | 16403 (100%) | 1579 (21%) | 14110 (100%) | 650 (100%) | 70470 (60%) |
| Obesity | 14738 (19%) | 0 (0%) | 2628 (35%) | 0 (0%) | 0 (0%) | 17366 (15%) |
| Unknown | 26723 (34%) | 0 (0%) | 3403 (45%) | 0 (0%) | 0 (0%) | 30126 (26%) |
| Brand | | | | | | |
| Ozempic | 28500 (36%) | 0 (0%) | 0 (0%) | 0 (0%) | 0 (0%) | 28500 (24%) |
| Rybelsus | 7674 (10%) | 0 (0%) | 0 (0%) | 0 (0%) | 0 (0%) | 7674 (7%) |
| Wegovy | 14738 (19%) | 0 (0%) | 0 (0%) | 0 (0%) | 0 (0%) | 14738 (12%) |
| Trulicity | 0 (0%) | 16403 (100%) | 0 (0%) | 0 (0%) | 0 (0%) | 16403 (14%) |

Continued on next page

**Table6 – continued from previous page**

| Feature | Semaglutide | Dulaglutide | Liraglutide | Tirzepatide | Other | Overall |
|---|---|---|---|---|---|---|
| Victoza | 0 (0%) | 0 (0%) | 1579 (21%) | 0 (0%) | 0 (0%) | 1579 (1%) |
| Saxenda | 0 (0%) | 0 (0%) | 2628 (35%) | 0 (0%) | 0 (0%) | 2628 (2%) |
| Mounjaro | 0 (0%) | 0 (0%) | 0 (0%) | 14110 (100%) | 0 (0%) | 14110 (12%) |
| Bydureon | 0 (0%) | 0 (0%) | 0 (0%) | 0 (0%) | 203 (31%) | 203 (0%) |
| Byetta | 0 (0%) | 0 (0%) | 0 (0%) | 0 (0%) | 27 (4%) | 27 (0%) |
| Soliqua | 0 (0%) | 0 (0%) | 0 (0%) | 0 (0%) | 333 (51%) | 333 (0%) |
| Unknown | 28277 (36%) | 0 (0%) | 3403 (45%) | 0 (0%) | 87 (13%) | 31767 (27%) |
| Period | | | | | | |
| 2022 Jan-Jun | 4377 (6%) | 1471 (9%) | 682 (9%) | 96 (1%) | 66 (10%) | 6692 (6%) |
| 2022 Jul-Dec | 25759 (33%) | 7980 (49%) | 4126 (54%) | 6531 (46%) | 305 (47%) | 44701 (38%) |
| 2023 Jan-Jun | 49053 (62%) | 6952 (42%) | 2802 (37%) | 7483 (53%) | 279 (43%) | 66569 (56%) |
| Comorbidities | | | | | | |
| Atrial Fibrilation | 5047 (6%) | 1514 (9%) | 355 (5%) | 710 (5%) | 78 (12%) | 7704 (7%) |
| Asthma | 16162 (20%) | 3111 (19%) | 1828 (24%) | 2804 (20%) | 99 (15%) | 24004 (20%) |
| CKD | 9489 (12%) | 3448 (21%) | 723 (10%) | 1223 (9%) | 164 (25%) | 15047 (13%) |
| COPD | 5353 (7%) | 1828 (11%) | 483 (6%) | 721 (5%) | 78 (12%) | 8463 (7%) |
| Glaucoma | 1713 (2%) | 534 (3%) | 114 (1%) | 202 (1%) | 31 (5%) | 2594 (2%) |
| Heart Failure | 5649 (7%) | 2058 (13%) | 451 (6%) | 711 (5%) | 98 (15%) | 8967 (8%) |
| Hyperlipidemia | 51155 (65%) | 12529 (76%) | 3798 (50%) | 8472 (60%) | 496 (76%) | 76450 (65%) |
| Hypertension | 50813 (64%) | 12875 (78%) | 4105 (54%) | 8582 (61%) | 505 (78%) | 76880 (65%) |
| Ischemic Heart Disease | 5222 (7%) | 1931 (12%) | 384 (5%) | 746 (5%) | 85 (13%) | 8368 (7%) |
| Acute MI | 2188 (3%) | 828 (5%) | 172 (2%) | 293 (2%) | 40 (6%) | 3521 (3%) |
| Ischemic Stroke | 187 (0%) | 72 (0%) | 19 (0%) | 29 (0%) | 4 (1%) | 311 (0%) |
| Major Depressive Disorder | 17965 (23%) | 3371 (21%) | 2100 (28%) | 2994 (21%) | 122 (19%) | 26552 (23%) |
| Osteoporosis | 2821 (4%) | 732 (4%) | 195 (3%) | 363 (3%) | 28 (4%) | 4139 (4%) |
| Bariatric Surgery | 2760 (3%) | 297 (2%) | 500 (7%) | 504 (4%) | 8 (1%) | 4069 (3%) |
| ADM Use | | | | | | |
| Metformin | 30922 (39%) | 10206 (62%) | 1718 (23%) | 5400 (38%) | 380 (58%) | 48626 (41%) |
| SGLT2i | 10633 (13%) | 4363 (27%) | 493 (6%) | 1690 (12%) | 193 (30%) | 17372 (15%) |
| DPP4 | 5678 (7%) | 2743 (17%) | 197 (3%) | 847 (6%) | 91 (14%) | 9556 (8%) |
| Sulfonylurea | 10075 (13%) | 4652 (28%) | 393 (5%) | 1413 (10%) | 185 (28%) | 16718 (14%) |
| Insulin | 5265 (7%) | 2111 (13%) | 662 (9%) | 759 (5%) | 96 (15%) | 8893 (8%) |
| ADM Use | | | | | | |
| Orlistat | 146 (0%) | 14 (0%) | 38 (0%) | 16 (0%) | 1 (0%) | 215 (0%) |
| Phentermine Topiramate | 483 (1%) | 11 (0%) | 70 (1%) | 112 (1%) | 0 (0%) | 676 (1%) |
| BMI | | | | | | |
| Mean (SD) | 38 (8.4) | 36 (8.5) | 40 (9.1) | 38 (8.4) | 35 (8.8) | 38 (8.5) |
| Missing | 33342 (42.1%) | 7057 (43.0%) | 3408 (44.8%) | 6977 (49.4%) | 375 (57.7%) | 51159 (43.4%) |
| Weight (in lbs) | | | | | | |

Continued on next page

**Table6 – continued from previous page**

| Feature | Semaglutide | Dulaglutide | Liraglutide | Tirzepatide | Other | Overall |
|---|---|---|---|---|---|---|
| Mean (SD) | 230 (54) | 220 (57) | 240 (58) | 240 (56) | 210 (55) | 230 (55) |
| Missing | 20676 (26.1%) | 3536 (21.6%) | 2227 (29.3%) | 3604 (25.5%) | 160 (24.6%) | 30203 (25.6%) |
| HbA1c | | | | | | |
| Mean (SD) | 7.0 (1.7) | 8.2 (1.9) | 6.3 (1.5) | 6.8 (1.6) | 8.7 (1.9) | 7.1 (1.8) |
| Missing | 27427 (34.6%) | 4390 (26.8%) | 3520 (46.3%) | 5270 (37.3%) | 236 (36.3%) | 40843 (34.6%) |

Table 6: Characteristics of patients first prescribed a GLP-1 RA in May 2022 or later, by specific medication Other includes the two smallest groups: Exenatide and Lixisenatide.: ADM=anti-diabetic medication, AI = American Indian, AN = Alaska Native, AOM=anti-obesity medication, Black = Black or African American, BMI = body mass index, CKD=chronic kidney disease, COPD=chronic obstructive pulmonary disease, DPP4= dipeptidyl peptidase 4 inhibitor, FDA=Food and Drug Administration, HbA1c=hemoglobin A1C, MI=myocardial infarction, NH = Native Hawaiian, PI = Pacific Islander, SD=standard deviation, SGLT2i=sodium/glucose cotransporter-2 inhibitor, T2D=type 2 diabetes mellitus.

### 7 Patient Characteristics for Ozempic, Wegovy, and Mounjaro After May 2022

Characteristics of patients first-prescribed Ozempic, Wegovy, or Mounjaro during the post-Tirzepatide approval period.

| | Ozempic<br>(N=9464) | Wegovy<br>(N=4884) | Mounjaro<br>(N=4741) |
|---|---|---|---|
| Age | | | |
| Mean (SD) | 57 (13) | 48 (12) | 52 (13) |
| Age Group | | | |
| 0-17 | 0 (0%) | 0 (0%) | 0 (0%) |
| 18-44 | 1783 (19%) | 1919 (39%) | 1362 (29%) |
| 45-64 | 4811 (51%) | 2517 (52%) | 2585 (55%) |
| 65+ | 2870 (30%) | 448 (9%) | 794 (17%) |
| Sex | | | |
| Female | 6186 (65%) | 4040 (83%) | 3376 (71%) |
| Male | 3272 (35%) | 843 (17%) | 1363 (29%) |
| Unknown | 6 (0%) | 1 (0%) | 2 (0%) |
| Race | | | |
| White | 6866 (73%) | 3464 (71%) | 3587 (76%) |
| Black | 1530 (16%) | 885 (18%) | 600 (13%) |
| Asian | 208 (2%) | 65 (1%) | 70 (1%) |
| AI or AN | 58 (1%) | 30 (1%) | 22 (0%) |
| NH or PI | 29 (0%) | 16 (0%) | 8 (0%) |
| Other Race | 425 (4%) | 256 (5%) | 305 (6%) |
| Declined to answer | 96 (1%) | 38 (1%) | 43 (1%) |
| Unknown | 252 (3%) | 130 (3%) | 106 (2%) |
| Ethnicity | | | |
| Hispanic or Latino | 883 (9%) | 493 (10%) | 552 (12%) |
| Not Hispanic or Latino | 8126 (86%) | 4165 (85%) | 3969 (84%) |
| Declined to answer | 130 (1%) | 54 (1%) | 41 (1%) |
| Unknown | 325 (3%) | 172 (4%) | 179 (4%) |
| T2D | 5762 (61%) | 284 (6%) | 2274 (48%) |
| Obesity or Overweight | 8056 (85%) | 4655 (95%) | 4203 (89%) |
| Period | | | |
| 2022 Jan-Jun | 609 (6%) | 163 (3%) | 34 (1%) |
| 2022 Jul-Dec | 3272 (35%) | 739 (15%) | 2173 (46%) |
| 2023 Jan-Jun | 5583 (59%) | 3982 (82%) | 2534 (53%) |
| Comorbidities | | | |
| Atrial Fibrilation | 689 (7%) | 153 (3%) | 234 (5%) |
| Asthma | 1822 (19%) | 1021 (21%) | 933 (20%) |
| CKD | 1311 (14%) | 189 (4%) | 396 (8%) |
| COPD | 737 (8%) | 153 (3%) | 221 (5%) |
| Glaucoma | 217 (2%) | 42 (1%) | 69 (1%) |
| Heart Failure | 807 (9%) | 131 (3%) | 229 (5%) |
| Hyperlipidemia | 6422 (68%) | 2405 (49%) | 2867 (60%) |
| Hypertension | 6541 (69%) | 2347 (48%) | 2914 (61%) |
| Ischemic Heart Disease | 798 (8%) | 137 (3%) | 249 (5%) |
| Acute MI | 339 (4%) | 56 (1%) | 101 (2%) |
| Ischemic Stroke | 35 (0%) | 4 (0%) | 6 (0%) |
| Major Depressive Disorder | 1852 (20%) | 1111 (23%) | 987 (21%) |
| Osteoporosis | 327 (3%) | 107 (2%) | 131 (3%) |
| Bariatric Surgery | 262 (3%) | 272 (6%) | 160 (3%) |

Continued on next page

**Table7 – continued from previous page**

| Feature | Ozempic | Wegovy | Moujaro |
|---|---|---|---|
| ADM Use | | | |
| Metformin | 4372 (46%) | 452 (9%) | 1895 (40%) |
| SGLT2i | 1565 (17%) | 43 (1%) | 580 (12%) |
| DPP4 | 824 (9%) | 9 (0%) | 317 (7%) |
| Sulfonylurea | 1420 (15%) | 12 (0%) | 480 (10%) |
| Insulin | 679 (7%) | 89 (2%) | 273 (6%) |
| ADM Use | | | |
| Orlistat | 11 (0%) | 10 (0%) | 7 (0%) |
| Phentermine Topiramate | 28 (0%) | 64 (1%) | 43 (1%) |
| BMI | | | |
| Mean (SD) | 37 (8.4) | 39 (8.2) | 38 (8.4) |
| Missing | 4789 (50.6%) | 2588 (53.0%) | 2337 (49.3%) |
| Weight (in lbs) | | | |
| Mean (SD) | 240 (57) | 230 (53) | 240 (57) |
| Missing | 2968 (31.4%) | 1618 (33.1%) | 1239 (26.1%) |
| HbA1c | | | |
| Mean (SD) | 7.2 (1.8) | 5.6 (0.51) | 6.8 (1.6) |
| Missing | 3537 (37.4%) | 2487 (50.9%) | 1734 (36.6%) |

Table 7: Characteristics of patients first prescribed a GLP-1 RA in May 2022 or later, for Ozempic, Wegovy, and Mounjaro.: ADM=anti-diabetic medication, AI = American Indian, AN = Alaska Native, AOM=anti-obesity medication, Black = Black or African American, BMI = body mass index, CKD=chronic kidney disease, COPD=chronic obstructive pulmonary disease, DPP4= dipeptidyl peptidase 4 inhibitor, FDA=Food and Drug Administration, HbA1c=hemoglobin A1C, MI=myocardial infarction, NH = Native Hawaiian, PI = Pacific Islander, SD=standard deviation, SGLT2i=sodium/glucose cotransporter-2 inhibitor, T2D=type 2 diabetes mellitus.

### 8 Data Definitions

#### 8.1 GLP-1 Medications

The following codes were used to identify patients prescribed a GLP-1. We excluded patients if a single GLP-1 could not be established at index.

Table 8: Concept codes used to identify GLP-1 prescriptions.

| CodeSystem | ConceptCode | ConceptName |
|---|---|---|
| RxNorm | 1991302 | semaglutide |
| RxNorm | 2599362 | 3 ML semaglutide 2.68 MG/ML Pen Injector |
| RxNorm | 2599365 | 3 ML semaglutide 2.68 MG/ML Pen Injector [Ozempic] |
| RxNorm | 2599361 | semaglutide 2.68 MG/ML |
| RxNorm | 2599364 | semaglutide 2.68 MG/ML [Ozempic] |
| RxNorm | 2599363 | semaglutide 2.68 MG/ML Pen Injector |
| RxNorm | 2599366 | semaglutide 2.68 MG/ML Pen Injector [Ozempic] |
| RxNorm | 2619152 | 0.25 MG, 0.5 MG Dose 3 ML semaglutide 0.68 MG/ML Pen Injector |
| RxNorm | 2619154 | 0.25 MG, 0.5 MG Dose 3 ML semaglutide 0.68 MG/ML Pen Injector [Ozempic] |
| RxNorm | 1991303 | semaglutide 1.34 MG/ML |
| RxNorm | 1991304 | semaglutide Injectable Product |
| RxNorm | 1991305 | semaglutide Pen Injector |
| RxNorm | 1991306 | 0.25 MG, 0.5 MG Dose 1.5 ML semaglutide 1.34 MG/ML Pen Injector |
| RxNorm | 1991307 | Ozempic |
| RxNorm | 1991308 | semaglutide 1.34 MG/ML [Ozempic] |
| RxNorm | 1991309 | semaglutide Pen Injector [Ozempic] |
| RxNorm | 1991310 | Ozempic Injectable Product |
| RxNorm | 1991311 | 0.25 MG, 0.5 MG Dose 1.5 ML semaglutide 1.34 MG/ML Pen Injector [Ozempic] |
| RxNorm | 1991316 | 1 MG Dose 1.5 ML semaglutide 1.34 MG/ML Pen Injector |
| RxNorm | 1991317 | 1 MG Dose 1.5 ML semaglutide 1.34 MG/ML Pen Injector [Ozempic] |
| RxNorm | 2200654 | semaglutide 3 MG Oral Tablet [Rybelsus] |
| RxNorm | 2200655 | semaglutide 7 MG |
| RxNorm | 2200656 | semaglutide 7 MG Oral Tablet |
| RxNorm | 2200657 | semaglutide 7 MG [Rybelsus] |
| RxNorm | 2200658 | semaglutide 7 MG Oral Tablet [Rybelsus] |
| RxNorm | 2200640 | semaglutide 14 MG |
| RxNorm | 2200641 | semaglutide Oral Product |
| RxNorm | 2200642 | semaglutide Pill |
| RxNorm | 2200643 | semaglutide Oral Tablet |
| RxNorm | 2200644 | semaglutide 14 MG Oral Tablet |
| RxNorm | 2200645 | Rybelsus |

Continued on next page

**Table8 – continued from previous page**

| CodeSystem | ConceptCode | ConceptName |
|---|---|---|
| RxNorm | 2200646 | semaglutide 14 MG [Rybelsus] |
| RxNorm | 2200647 | semaglutide Oral Tablet [Rybelsus] |
| RxNorm | 2200648 | Rybelsus Oral Product |
| RxNorm | 2200649 | Rybelsus Pill |
| RxNorm | 2200650 | semaglutide 14 MG Oral Tablet [Rybelsus] |
| RxNorm | 2200651 | semaglutide 3 MG |
| RxNorm | 2200652 | semaglutide 3 MG Oral Tablet |
| RxNorm | 2200653 | semaglutide 3 MG [Rybelsus] |
| RxNorm | 2398841 | 3 ML semaglutide 1.34 MG/ML Pen Injector |
| RxNorm | 2398842 | 3 ML semaglutide 1.34 MG/ML Pen Injector [Ozempic] |
| RxNorm | 2398843 | semaglutide 1.34 MG/ML Pen Injector |
| RxNorm | 2398844 | semaglutide 1.34 MG/ML Pen Injector [Ozempic] |
| RxNorm | 2553504 | semaglutide Auto-Injector [Wegovy] |
| RxNorm | 2553506 | 0.5 ML semaglutide 0.5 MG/ML Auto-Injector [Wegovy] |
| RxNorm | 2553604 | semaglutide 1 MG/ML Auto-Injector |
| RxNorm | 2553606 | semaglutide 2 MG/ML |
| RxNorm | 2553700 | semaglutide 2 MG/ML Auto-Injector [Wegovy] |
| RxNorm | 2553900 | semaglutide 2.27 MG/ML |
| RxNorm | 2553902 | semaglutide 2.27 MG/ML [Wegovy] |
| RxNorm | 2553904 | semaglutide 2.27 MG/ML Auto-Injector |
| RxNorm | 2554103 | semaglutide 3.2 MG/ML [Wegovy] |
| RxNorm | 2553601 | 0.5 ML semaglutide 1 MG/ML Auto-Injector |
| RxNorm | 2553608 | semaglutide 2 MG/ML [Wegovy] |
| RxNorm | 2554101 | semaglutide 3.2 MG/ML |
| RxNorm | 2553501 | 0.5 ML semaglutide 0.5 MG/ML Auto-Injector |
| RxNorm | 2553905 | semaglutide 2.27 MG/ML Auto-Injector [Wegovy] |
| RxNorm | 2554102 | 0.75 ML semaglutide 3.2 MG/ML Auto-Injector |
| RxNorm | 2553400 | semaglutide 0.5 MG/ML |
| RxNorm | 2553505 | Wegovy Injectable Product |
| RxNorm | 2553507 | semaglutide 0.5 MG/ML Auto-Injector |
| RxNorm | 2553605 | semaglutide 1 MG/ML Auto-Injector [Wegovy] |
| RxNorm | 2553607 | semaglutide 2 MG/ML Auto-Injector |
| RxNorm | 2553901 | 0.75 ML semaglutide 2.27 MG/ML Auto-Injector |
| RxNorm | 2553903 | 0.75 ML semaglutide 2.27 MG/ML Auto-Injector [Wegovy] |
| RxNorm | 2554104 | 0.75 ML semaglutide 3.2 MG/ML Auto-Injector [Wegovy] |
| RxNorm | 2553502 | Wegovy |
| RxNorm | 2553803 | 0.5 ML semaglutide 2 MG/ML Auto-Injector [Wegovy] |
| RxNorm | 2554105 | semaglutide 3.2 MG/ML Auto-Injector |
| RxNorm | 2553503 | semaglutide 0.5 MG/ML [Wegovy] |
| RxNorm | 2553802 | 0.5 ML semaglutide 2 MG/ML Auto-Injector |
| RxNorm | 2554106 | semaglutide 3.2 MG/ML Auto-Injector [Wegovy] |
| RxNorm | 2553508 | semaglutide 0.5 MG/ML Auto-Injector [Wegovy] |
| RxNorm | 2553600 | semaglutide 1 MG/ML |

Continued on next page

**Table8 – continued from previous page**

| CodeSystem | ConceptCode | ConceptName |
|---|---|---|
| RxNorm | 2553602 | semaglutide 1 MG/ML [Wegovy] |
| RxNorm | 2553500 | semaglutide Auto-Injector |
| RxNorm | 2553603 | 0.5 ML semaglutide 1 MG/ML Auto-Injector [Wegovy] |
| RxNorm | 2619151 | semaglutide 0.68 MG/ML |
| RxNorm | 2619153 | semaglutide 0.68 MG/ML [Ozempic] |
| RxNorm | 2553502 | Wegovy |
| RxNorm | 2553504 | semaglutide Auto-Injector [Wegovy] |
| RxNorm | 2553506 | 0.5 ML semaglutide 0.5 MG/ML Auto-Injector [Wegovy] |
| RxNorm | 2553700 | semaglutide 2 MG/ML Auto-Injector [Wegovy] |
| RxNorm | 2553902 | semaglutide 2.27 MG/ML [Wegovy] |
| RxNorm | 2554103 | semaglutide 3.2 MG/ML [Wegovy] |
| RxNorm | 2553608 | semaglutide 2 MG/ML [Wegovy] |
| RxNorm | 2553905 | semaglutide 2.27 MG/ML Auto-Injector [Wegovy] |
| RxNorm | 2553505 | Wegovy Injectable Product |
| RxNorm | 2553605 | semaglutide 1 MG/ML Auto-Injector [Wegovy] |
| RxNorm | 2553903 | 0.75 ML semaglutide 2.27 MG/ML Auto-Injector [Wegovy] |
| RxNorm | 2554104 | 0.75 ML semaglutide 3.2 MG/ML Auto-Injector [Wegovy] |
| RxNorm | 2553803 | 0.5 ML semaglutide 2 MG/ML Auto-Injector [Wegovy] |
| RxNorm | 2553503 | semaglutide 0.5 MG/ML [Wegovy] |
| RxNorm | 2554106 | semaglutide 3.2 MG/ML Auto-Injector [Wegovy] |
| RxNorm | 2553508 | semaglutide 0.5 MG/ML Auto-Injector [Wegovy] |
| RxNorm | 2553602 | semaglutide 1 MG/ML [Wegovy] |
| RxNorm | 2553603 | 0.5 ML semaglutide 1 MG/ML Auto-Injector [Wegovy] |
| RxNorm | 1991307 | Ozempic |
| RxNorm | 2599365 | 3 ML semaglutide 2.68 MG/ML Pen Injector [Ozempic] |
| RxNorm | 2599366 | semaglutide 2.68 MG/ML Pen Injector [Ozempic] |
| RxNorm | 2619154 | 0.25 MG, 0.5 MG Dose 3 ML semaglutide 0.68 MG/ML Pen Injector [Ozempic] |
| RxNorm | 1991308 | semaglutide 1.34 MG/ML [Ozempic] |
| RxNorm | 1991309 | semaglutide Pen Injector [Ozempic] |
| RxNorm | 1991310 | Ozempic Injectable Product |
| RxNorm | 1991311 | 0.25 MG, 0.5 MG Dose 1.5 ML semaglutide 1.34 MG/ML Pen Injector [Ozempic] |
| RxNorm | 1991317 | 1 MG Dose 1.5 ML semaglutide 1.34 MG/ML Pen Injector [Ozempic] |
| RxNorm | 2398842 | 3 ML semaglutide 1.34 MG/ML Pen Injector [Ozempic] |
| RxNorm | 2398844 | semaglutide 1.34 MG/ML Pen Injector [Ozempic] |
| RxNorm | 2200645 | Rybelsus |
| RxNorm | 2200654 | semaglutide 3 MG Oral Tablet [Rybelsus] |
| RxNorm | 2200657 | semaglutide 7 MG [Rybelsus] |
| RxNorm | 2200658 | semaglutide 7 MG Oral Tablet [Rybelsus] |
| RxNorm | 2200646 | semaglutide 14 MG [Rybelsus] |
| RxNorm | 2200647 | semaglutide Oral Tablet [Rybelsus] |

Continued on next page

**Table8 – continued from previous page**

| CodeSystem | ConceptCode | ConceptName |
|---|---|---|
| RxNorm | 2200648 | Rybelsus Oral Product |
| RxNorm | 2200649 | Rybelsus Pill |
| RxNorm | 2200650 | semaglutide 14 MG Oral Tablet [Rybelsus] |
| RxNorm | 2200653 | semaglutide 3 MG [Rybelsus] |
| RxNorm | 2601723 | tirzepatide |
| RxNorm | 2601743 | 0.5 ML tirzepatide 10 MG/ML Auto-Injector |
| RxNorm | 2601746 | 0.5 ML tirzepatide 10 MG/ML Auto-Injector [Mounjaro] |
| RxNorm | 2601784 | 0.5 ML tirzepatide 15 MG/ML Auto-Injector |
| RxNorm | 2601785 | 0.5 ML tirzepatide 15 MG/ML Auto-Injector [Mounjaro] |
| RxNorm | 2601767 | 0.5 ML tirzepatide 20 MG/ML Auto-Injector |
| RxNorm | 2601770 | 0.5 ML tirzepatide 20 MG/ML Auto-Injector [Mounjaro] |
| RxNorm | 2601773 | 0.5 ML tirzepatide 25 MG/ML Auto-Injector |
| RxNorm | 2601761 | 0.5 ML tirzepatide 5 MG/ML Auto-Injector |
| RxNorm | 2601764 | 0.5 ML tirzepatide 5 MG/ML Auto-Injector [Mounjaro] |
| RxNorm | 2601742 | tirzepatide 10 MG/ML |
| RxNorm | 2601745 | tirzepatide 10 MG/ML [Mounjaro] |
| RxNorm | 2601744 | tirzepatide 10 MG/ML Auto-Injector |
| RxNorm | 2601747 | tirzepatide 10 MG/ML Auto-Injector [Mounjaro] |
| RxNorm | 2601778 | tirzepatide 15 MG/ML |
| RxNorm | 2601781 | tirzepatide 15 MG/ML [Mounjaro] |
| RxNorm | 2601780 | tirzepatide 15 MG/ML Auto-Injector |
| RxNorm | 2601783 | tirzepatide 15 MG/ML Auto-Injector [Mounjaro] |
| RxNorm | 2601736 | Mounjaro Injectable Product |
| RxNorm | 2601759 | tirzepatide 30 MG/ML Auto-Injector [Mounjaro] |
| RxNorm | 2601760 | tirzepatide 5 MG/ML |
| RxNorm | 2601763 | tirzepatide 5 MG/ML [Mounjaro] |
| RxNorm | 2601762 | tirzepatide 5 MG/ML Auto-Injector |
| RxNorm | 2601765 | tirzepatide 5 MG/ML Auto-Injector [Mounjaro] |
| RxNorm | 2601731 | tirzepatide Auto-Injector |
| RxNorm | 2601737 | tirzepatide Auto-Injector [Mounjaro] |
| RxNorm | 2601730 | tirzepatide Injectable Product |
| RxNorm | 2601755 | 0.5 ML tirzepatide 30 MG/ML Auto-Injector |
| RxNorm | 2601766 | tirzepatide 20 MG/ML |
| RxNorm | 2601757 | tirzepatide 30 MG/ML [Mounjaro] |
| RxNorm | 2601769 | tirzepatide 20 MG/ML [Mounjaro] |
| RxNorm | 2601756 | tirzepatide 30 MG/ML Auto-Injector |
| RxNorm | 2601771 | tirzepatide 20 MG/ML Auto-Injector [Mounjaro] |
| RxNorm | 2601772 | tirzepatide 25 MG/ML |
| RxNorm | 2601775 | tirzepatide 25 MG/ML [Mounjaro] |
| RxNorm | 2601774 | tirzepatide 25 MG/ML Auto-Injector |
| RxNorm | 2601768 | tirzepatide 20 MG/ML Auto-Injector |
| RxNorm | 2601777 | tirzepatide 25 MG/ML Auto-Injector [Mounjaro] |
| RxNorm | 2601754 | tirzepatide 30 MG/ML |

Continued on next page

**Table8 – continued from previous page**

| CodeSystem | ConceptCode | ConceptName |
|---|---|---|
| RxNorm | 2601758 | 0.5 ML tirzepatide 30 MG/ML Auto-Injector [Mounjaro] |
| RxNorm | 2601776 | 0.5 ML tirzepatide 25 MG/ML Auto-Injector [Mounjaro] |
| RxNorm | 2601746 | 0.5 ML tirzepatide 10 MG/ML Auto-Injector [Mounjaro] |
| RxNorm | 2601785 | 0.5 ML tirzepatide 15 MG/ML Auto-Injector [Mounjaro] |
| RxNorm | 2601770 | 0.5 ML tirzepatide 20 MG/ML Auto-Injector [Mounjaro] |
| RxNorm | 2601764 | 0.5 ML tirzepatide 5 MG/ML Auto-Injector [Mounjaro] |
| RxNorm | 2601745 | tirzepatide 10 MG/ML [Mounjaro] |
| RxNorm | 2601747 | tirzepatide 10 MG/ML Auto-Injector [Mounjaro] |
| RxNorm | 2601781 | tirzepatide 15 MG/ML [Mounjaro] |
| RxNorm | 2601783 | tirzepatide 15 MG/ML Auto-Injector [Mounjaro] |
| RxNorm | 2601734 | Mounjaro |
| RxNorm | 2601759 | tirzepatide 30 MG/ML Auto-Injector [Mounjaro] |
| RxNorm | 2601763 | tirzepatide 5 MG/ML [Mounjaro] |
| RxNorm | 2601765 | tirzepatide 5 MG/ML Auto-Injector [Mounjaro] |
| RxNorm | 2601737 | tirzepatide Auto-Injector [Mounjaro] |
| RxNorm | 2601757 | tirzepatide 30 MG/ML [Mounjaro] |
| RxNorm | 2601769 | tirzepatide 20 MG/ML [Mounjaro] |
| RxNorm | 2601771 | tirzepatide 20 MG/ML Auto-Injector [Mounjaro] |
| RxNorm | 2601777 | tirzepatide 25 MG/ML Auto-Injector [Mounjaro] |
| RxNorm | 2601758 | 0.5 ML tirzepatide 30 MG/ML Auto-Injector [Mounjaro] |
| RxNorm | 2601776 | 0.5 ML tirzepatide 25 MG/ML Auto-Injector [Mounjaro] |
| RxNorm | 1551291 | dulaglutide |
| RxNorm | 1551292 | dulaglutide 1.5 MG/ML |
| RxNorm | 1551293 | dulaglutide Injectable Product |
| RxNorm | 1551295 | 0.5 ML dulaglutide 1.5 MG/ML Auto-Injector |
| RxNorm | 1551296 | Trulicity |
| RxNorm | 1551297 | dulaglutide 1.5 MG/ML [Trulicity] |
| RxNorm | 1551299 | Trulicity Injectable Product |
| RxNorm | 1551300 | 0.5 ML dulaglutide 1.5 MG/ML Auto-Injector [Trulicity] |
| RxNorm | 1551301 | dulaglutide 1.5 MG/ML Auto-Injector |
| RxNorm | 1551302 | dulaglutide 1.5 MG/ML Auto-Injector [Trulicity] |
| RxNorm | 1551303 | dulaglutide 3 MG/ML |
| RxNorm | 1551304 | 0.5 ML dulaglutide 3 MG/ML Auto-Injector |
| RxNorm | 1551305 | dulaglutide 3 MG/ML [Trulicity] |
| RxNorm | 1551306 | 0.5 ML dulaglutide 3 MG/ML Auto-Injector [Trulicity] |
| RxNorm | 1551307 | dulaglutide 3 MG/ML Auto-Injector |
| RxNorm | 1551308 | dulaglutide 3 MG/ML Auto-Injector [Trulicity] |
| RxNorm | 1649584 | dulaglutide Auto-Injector |
| RxNorm | 1649586 | dulaglutide Auto-Injector [Trulicity] |
| RxNorm | 2395776 | dulaglutide 6 MG/ML |
| RxNorm | 2395777 | 0.5 ML dulaglutide 6 MG/ML Auto-Injector |
| RxNorm | 2395778 | dulaglutide 6 MG/ML [Trulicity] |
| RxNorm | 2395779 | 0.5 ML dulaglutide 6 MG/ML Auto-Injector [Trulicity] |

Continued on next page

**Table8 – continued from previous page**

| CodeSystem | ConceptCode | ConceptName |
|---|---|---|
| RxNorm | 2395780 | dulaglutide 6 MG/ML Auto-Injector |
| RxNorm | 2395781 | dulaglutide 6 MG/ML Auto-Injector [Trulicity] |
| RxNorm | 2395782 | dulaglutide 9 MG/ML |
| RxNorm | 2395783 | 0.5 ML dulaglutide 9 MG/ML Auto-Injector |
| RxNorm | 2395784 | dulaglutide 9 MG/ML [Trulicity] |
| RxNorm | 2395785 | 0.5 ML dulaglutide 9 MG/ML Auto-Injector [Trulicity] |
| RxNorm | 2395786 | dulaglutide 9 MG/ML Auto-Injector |
| RxNorm | 2395787 | dulaglutide 9 MG/ML Auto-Injector [Trulicity] |
| RxNorm | 1551294 | dulaglutide Prefilled Syringe |
| RxNorm | 1551298 | dulaglutide Prefilled Syringe [Trulicity] |
| RxNorm | 1551296 | Trulicity |
| RxNorm | 1551297 | dulaglutide 1.5 MG/ML [Trulicity] |
| RxNorm | 1551299 | Trulicity Injectable Product |
| RxNorm | 1551300 | 0.5 ML dulaglutide 1.5 MG/ML Auto-Injector [Trulicity] |
| RxNorm | 1551302 | dulaglutide 1.5 MG/ML Auto-Injector [Trulicity] |
| RxNorm | 1551305 | dulaglutide 3 MG/ML [Trulicity] |
| RxNorm | 1551306 | 0.5 ML dulaglutide 3 MG/ML Auto-Injector [Trulicity] |
| RxNorm | 1551308 | dulaglutide 3 MG/ML Auto-Injector [Trulicity] |
| RxNorm | 1649586 | dulaglutide Auto-Injector [Trulicity] |
| RxNorm | 2395778 | dulaglutide 6 MG/ML [Trulicity] |
| RxNorm | 2395779 | 0.5 ML dulaglutide 6 MG/ML Auto-Injector [Trulicity] |
| RxNorm | 2395781 | dulaglutide 6 MG/ML Auto-Injector [Trulicity] |
| RxNorm | 2395784 | dulaglutide 9 MG/ML [Trulicity] |
| RxNorm | 2395785 | 0.5 ML dulaglutide 9 MG/ML Auto-Injector [Trulicity] |
| RxNorm | 2395787 | dulaglutide 9 MG/ML Auto-Injector [Trulicity] |
| RxNorm | 1551298 | dulaglutide Prefilled Syringe [Trulicity] |
| RxNorm | 1440051 | lixisenatide |
| RxNorm | 1440052 | lixisenatide 0.1 MG/ML |
| RxNorm | 1440053 | lixisenatide Injectable Product |
| RxNorm | 1440056 | lixisenatide 0.05 MG/ML |
| RxNorm | 1858991 | lixisenatide 0.033 MG/ML |
| RxNorm | 1858992 | insulin glargine / lixisenatide Injectable Product |
| RxNorm | 1858993 | insulin glargine / lixisenatide Pen Injector |
| RxNorm | 1858994 | insulin glargine / lixisenatide |
| RxNorm | 1858995 | 3 ML insulin glargine 100 UNT/ML / lixisenatide 0.033 MG/ML Pen Injector |
| RxNorm | 1858996 | Soliqua |
| RxNorm | 1858997 | insulin glargine 100 UNT/ML / lixisenatide 0.033 MG/ML [Soliqua] |
| RxNorm | 1858998 | insulin glargine / lixisenatide Pen Injector [Soliqua] |
| RxNorm | 1858999 | Soliqua Injectable Product |
| RxNorm | 1859000 | 3 ML insulin glargine 100 UNT/ML / lixisenatide 0.033 MG/ML Pen Injector [Soliqua] |

Continued on next page

**Table8 – continued from previous page**

| CodeSystem | ConceptCode | ConceptName |
|---|---|---|
| RxNorm | 1859001 | insulin glargine 100 UNT/ML / lixisenatide 0.033 MG/ML Pen Injector |
| RxNorm | 1859002 | insulin glargine 100 UNT/ML / lixisenatide 0.033 MG/ML Pen Injector [Soliqua] |
| RxNorm | 1803885 | lixisenatide Pen Injector |
| RxNorm | 1803886 | lixisenatide 0.05 MG/ML Pen Injector |
| RxNorm | 1803887 | Adlyxin |
| RxNorm | 1803888 | lixisenatide 0.05 MG/ML [Adlyxin] |
| RxNorm | 1803889 | lixisenatide Pen Injector [Adlyxin] |
| RxNorm | 1803890 | Adlyxin Injectable Product |
| RxNorm | 1803891 | lixisenatide 0.05 MG/ML Pen Injector [Adlyxin] |
| RxNorm | 1803892 | 3 ML lixisenatide 0.05 MG/ML Pen Injector |
| RxNorm | 1803893 | 3 ML lixisenatide 0.05 MG/ML Pen Injector [Adlyxin] |
| RxNorm | 1803894 | 3 ML lixisenatide 0.1 MG/ML Pen Injector |
| RxNorm | 1803895 | lixisenatide 0.1 MG/ML [Adlyxin] |
| RxNorm | 1803896 | 3 ML lixisenatide 0.1 MG/ML Pen Injector [Adlyxin] |
| RxNorm | 1803897 | lixisenatide 0.1 MG/ML Pen Injector |
| RxNorm | 1803898 | lixisenatide 0.1 MG/ML Pen Injector [Adlyxin] |
| RxNorm | 1803902 | {1 (3 ML lixisenatide 0.05 MG/ML Pen Injector) / 1 (3 ML lixisenatide 0.1 MG/ML Pen Injector) } Pack |
| RxNorm | 1803903 | {1 (3 ML lixisenatide 0.05 MG/ML Pen Injector [Adlyxin]) / 1 (3 ML lixisenatide 0.1 MG/ML Pen Injector [Adlyxin]) } Pack [Adlyxin Starter Kit] |
| RxNorm | 1440054 | Lixisenatide Injectable Solution |
| RxNorm | 1440055 | Lixisenatide 0.1 MG/ML Injectable Solution |
| RxNorm | 1440057 | Lixisenatide 0.05 MG/ML Injectable Solution |
| RxNorm | 1858996 | Soliqua |
| RxNorm | 1858997 | insulin glargine 100 UNT/ML / lixisenatide 0.033 MG/ML [Soliqua] |
| RxNorm | 1858998 | insulin glargine / lixisenatide Pen Injector [Soliqua] |
| RxNorm | 1858999 | Soliqua Injectable Product |
| RxNorm | 1859000 | 3 ML insulin glargine 100 UNT/ML / lixisenatide 0.033 MG/ML Pen Injector [Soliqua] |
| RxNorm | 1859002 | insulin glargine 100 UNT/ML / lixisenatide 0.033 MG/ML Pen Injector [Soliqua] |
| RxNorm | 1803887 | Adlyxin |
| RxNorm | 1803888 | lixisenatide 0.05 MG/ML [Adlyxin] |
| RxNorm | 1803889 | lixisenatide Pen Injector [Adlyxin] |
| RxNorm | 1803890 | Adlyxin Injectable Product |
| RxNorm | 1803891 | lixisenatide 0.05 MG/ML Pen Injector [Adlyxin] |
| RxNorm | 1803893 | 3 ML lixisenatide 0.05 MG/ML Pen Injector [Adlyxin] |
| RxNorm | 1803895 | lixisenatide 0.1 MG/ML [Adlyxin] |
| RxNorm | 1803896 | 3 ML lixisenatide 0.1 MG/ML Pen Injector [Adlyxin] |

Continued on next page

**Table8 – continued from previous page**

| CodeSystem | ConceptCode | ConceptName |
|---|---|---|
| RxNorm | 1803898 | lixisenatide 0.1 MG/ML Pen Injector [Adlyxin] |
| RxNorm | 1803903 | {1 (3 ML lixisenatide 0.05 MG/ML Pen Injector [Adlyxin]) / 1 (3 ML lixisenatide 0.1 MG/ML Pen Injector [Adlyxin]) } Pack [Adlyxin Starter Kit] |
| RxNorm | 475968 | liraglutide |
| RxNorm | 1186578 | Victoza Injectable Product |
| RxNorm | 1163230 | liraglutide Injectable Product |
| RxNorm | 1360105 | liraglutide 6 MG/ML Pen Injector |
| RxNorm | 1360495 | liraglutide 6 MG/ML Pen Injector [Victoza] |
| RxNorm | 1598264 | Saxenda |
| RxNorm | 1598265 | liraglutide 6 MG/ML [Saxenda] |
| RxNorm | 1598267 | Saxenda Injectable Product |
| RxNorm | 1598268 | 3 ML liraglutide 6 MG/ML Pen Injector [Saxenda] |
| RxNorm | 1598269 | liraglutide 6 MG/ML Pen Injector [Saxenda] |
| RxNorm | 1653594 | liraglutide Pen Injector |
| RxNorm | 1653597 | liraglutide Pen Injector [Victoza] |
| RxNorm | 1653600 | liraglutide Pen Injector [Saxenda] |
| RxNorm | 1727493 | insulin degludec / liraglutide |
| RxNorm | 1860164 | liraglutide 3.6 MG/ML |
| RxNorm | 1860165 | insulin degludec / liraglutide Injectable Product |
| RxNorm | 1860166 | insulin degludec / liraglutide Pen Injector |
| RxNorm | 1860167 | 3 ML insulin degludec 100 UNT/ML / liraglutide 3.6 MG/ML Pen Injector |
| RxNorm | 1860168 | Xultophy |
| RxNorm | 1860169 | insulin degludec 100 UNT/ML / liraglutide 3.6 MG/ML [Xultophy] |
| RxNorm | 1860170 | insulin degludec / liraglutide Pen Injector [Xultophy] |
| RxNorm | 1860171 | Xultophy Injectable Product |
| RxNorm | 1860172 | 3 ML insulin degludec 100 UNT/ML / liraglutide 3.6 MG/ML Pen Injector [Xultophy] |
| RxNorm | 1860173 | insulin degludec 100 UNT/ML / liraglutide 3.6 MG/ML Pen Injector |
| RxNorm | 1860174 | insulin degludec 100 UNT/ML / liraglutide 3.6 MG/ML Pen Injector [Xultophy] |
| RxNorm | 897125 | liraglutide Prefilled Syringe [Victoza] |
| RxNorm | 897121 | liraglutide Prefilled Syringe |
| RxNorm | 1598266 | liraglutide Prefilled Syringe [Saxenda] |
| RxNorm | 897120 | liraglutide 6 MG/ML |
| RxNorm | 897122 | 3 ML liraglutide 6 MG/ML Pen Injector |
| RxNorm | 897123 | Victoza |
| RxNorm | 897124 | liraglutide 6 MG/ML [Victoza] |
| RxNorm | 897126 | 3 ML liraglutide 6 MG/ML Pen Injector [Victoza] |
| RxNorm | 897123 | Victoza |

Continued on next page

**Table8 – continued from previous page**

| CodeSystem | ConceptCode | ConceptName |
|---|---|---|
| RxNorm | 1186578 | Victoza Injectable Product |
| RxNorm | 1360495 | liraglutide 6 MG/ML Pen Injector [Victoza] |
| RxNorm | 1653597 | liraglutide Pen Injector [Victoza] |
| RxNorm | 897125 | liraglutide Prefilled Syringe [Victoza] |
| RxNorm | 897124 | liraglutide 6 MG/ML [Victoza] |
| RxNorm | 897126 | 3 ML liraglutide 6 MG/ML Pen Injector [Victoza] |
| RxNorm | 1598264 | Saxenda |
| RxNorm | 1598265 | liraglutide 6 MG/ML [Saxenda] |
| RxNorm | 1598267 | Saxenda Injectable Product |
| RxNorm | 1598268 | 3 ML liraglutide 6 MG/ML Pen Injector [Saxenda] |
| RxNorm | 1598269 | liraglutide 6 MG/ML Pen Injector [Saxenda] |
| RxNorm | 1653600 | liraglutide Pen Injector [Saxenda] |
| RxNorm | 1598266 | liraglutide Prefilled Syringe [Saxenda] |
| RxNorm | 60548 | exenatide |
| RxNorm | 1242961 | exenatide 3.08 MG/ML |
| RxNorm | 1242963 | exenatide 2 MG Injection |
| RxNorm | 1242964 | Bydureon |
| RxNorm | 1242965 | exenatide 3.08 MG/ML [Bydureon] |
| RxNorm | 1242967 | Bydureon Injectable Product |
| RxNorm | 1242968 | exenatide 2 MG Injection [Bydureon] |
| RxNorm | 1359802 | exenatide 0.005 MG/ACTUAT Pen Injector [Byetta] |
| RxNorm | 1359979 | exenatide 0.01 MG/ACTUAT Pen Injector [Byetta] |
| RxNorm | 1360454 | exenatide 0.01 MG/ACTUAT Pen Injector |
| RxNorm | 1359640 | exenatide 0.005 MG/ACTUAT Pen Injector |
| RxNorm | 1544919 | exenatide 3.08 MG/ML Pen Injector |
| RxNorm | 1544920 | exenatide 3.08 MG/ML Pen Injector [Bydureon] |
| RxNorm | 1544916 | 0.65 ML exenatide 3.08 MG/ML Pen Injector |
| RxNorm | 1544918 | 0.65 ML exenatide 3.08 MG/ML Pen Injector [Bydureon] |
| RxNorm | 1653610 | exenatide 2 MG |
| RxNorm | 1653611 | exenatide Injection |
| RxNorm | 1653613 | exenatide 2 MG [Bydureon] |
| RxNorm | 1653614 | exenatide Injection [Bydureon] |
| RxNorm | 1653616 | exenatide Pen Injector |
| RxNorm | 1653619 | exenatide Pen Injector [Bydureon] |
| RxNorm | 1653625 | exenatide Pen Injector [Byetta] |
| RxNorm | 1990864 | exenatide 2.35 MG/ML |
| RxNorm | 1990865 | exenatide Auto-Injector |
| RxNorm | 1990866 | 0.85 ML exenatide 2.35 MG/ML Auto-Injector |
| RxNorm | 1990867 | exenatide 2.35 MG/ML [Bydureon] |
| RxNorm | 1990868 | exenatide Auto-Injector [Bydureon] |
| RxNorm | 1990869 | 0.85 ML exenatide 2.35 MG/ML Auto-Injector [Bydureon] |
| RxNorm | 1990870 | exenatide 2.35 MG/ML Auto-Injector |
| RxNorm | 1990871 | exenatide 2.35 MG/ML Auto-Injector [Bydureon] |

Continued on next page

**Table8 – continued from previous page**

| CodeSystem | ConceptCode | ConceptName |
|---|---|---|
| RxNorm | 604750 | exenatide 0.250 MG/ML Injectable Solution |
| RxNorm | 604749 | exenatide Injectable Solution |
| RxNorm | 604752 | exenatide 0.250 MG/ML [Byetta] |
| RxNorm | 604748 | exenatide 0.250 MG/ML |
| RxNorm | 604753 | exenatide Injectable Solution [Byetta] |
| RxNorm | 604754 | exenatide 0.250 MG/ML Injectable Solution [Byetta] |
| RxNorm | 847909 | exenatide Prefilled Syringe |
| RxNorm | 847912 | exenatide Prefilled Syringe [Byetta] |
| RxNorm | 744863 | exenatide 0.25 MG/ML Injectable Solution |
| RxNorm | 744862 | exenatide 0.25 MG/ML |
| RxNorm | 744864 | exenatide 0.25 MG/ML [Byetta] |
| RxNorm | 744865 | exenatide 0.25 MG/ML Injectable Solution [Byetta] |
| RxNorm | 1242966 | exenatide Injectable Suspension [Bydureon] |
| RxNorm | 1242962 | exenatide Injectable Suspension |
| RxNorm | 1544917 | exenatide Prefilled Syringe [Bydureon] |
| RxNorm | 1163790 | exenatide Injectable Product |
| RxNorm | 1169415 | Byetta Injectable Product |
| RxNorm | 604751 | Byetta |
| RxNorm | 847908 | exenatide 0.01 MG/ACTUAT |
| RxNorm | 847910 | 60 ACTUAT exenatide 0.01 MG/ACTUAT Pen Injector |
| RxNorm | 847911 | exenatide 0.01 MG/ACTUAT [Byetta] |
| RxNorm | 847913 | 60 ACTUAT exenatide 0.01 MG/ACTUAT Pen Injector [Byetta] |
| RxNorm | 847914 | exenatide 0.005 MG/ACTUAT |
| RxNorm | 847915 | 60 ACTUAT exenatide 0.005 MG/ACTUAT Pen Injector |
| RxNorm | 847916 | exenatide 0.005 MG/ACTUAT [Byetta] |
| RxNorm | 847917 | 60 ACTUAT exenatide 0.005 MG/ACTUAT Pen Injector [Byetta] |
| RxNorm | 1242964 | Bydureon |
| RxNorm | 1242965 | exenatide 3.08 MG/ML [Bydureon] |
| RxNorm | 1242967 | Bydureon Injectable Product |
| RxNorm | 1242968 | exenatide 2 MG Injection [Bydureon] |
| RxNorm | 1544920 | exenatide 3.08 MG/ML Pen Injector [Bydureon] |
| RxNorm | 1544918 | 0.65 ML exenatide 3.08 MG/ML Pen Injector [Bydureon] |
| RxNorm | 1653613 | exenatide 2 MG [Bydureon] |
| RxNorm | 1653614 | exenatide Injection [Bydureon] |
| RxNorm | 1653619 | exenatide Pen Injector [Bydureon] |
| RxNorm | 1990867 | exenatide 2.35 MG/ML [Bydureon] |
| RxNorm | 1990868 | exenatide Auto-Injector [Bydureon] |
| RxNorm | 1990869 | 0.85 ML exenatide 2.35 MG/ML Auto-Injector [Bydureon] |
| RxNorm | 1990871 | exenatide 2.35 MG/ML Auto-Injector [Bydureon] |
| RxNorm | 1242966 | exenatide Injectable Suspension [Bydureon] |
| RxNorm | 1544917 | exenatide Prefilled Syringe [Bydureon] |

Continued on next page

**Table8 – continued from previous page**

| CodeSystem | ConceptCode | ConceptName |
|---|---|---|
| RxNorm | 604751 | Byetta |
| RxNorm | 1359802 | exenatide 0.005 MG/ACTUAT Pen Injector [Byetta] |
| RxNorm | 1359979 | exenatide 0.01 MG/ACTUAT Pen Injector [Byetta] |
| RxNorm | 1653625 | exenatide Pen Injector [Byetta] |
| RxNorm | 604753 | exenatide Injectable Solution [Byetta] |
| RxNorm | 604754 | exenatide 0.250 MG/ML Injectable Solution [Byetta] |
| RxNorm | 847912 | exenatide Prefilled Syringe [Byetta] |
| RxNorm | 744864 | exenatide 0.25 MG/ML [Byetta] |
| RxNorm | 744865 | exenatide 0.25 MG/ML Injectable Solution [Byetta] |
| RxNorm | 1169415 | Byetta Injectable Product |
| RxNorm | 847911 | exenatide 0.01 MG/ACTUAT [Byetta] |
| RxNorm | 847913 | 60 ACTUAT exenatide 0.01 MG/ACTUAT Pen Injector [Byetta] |
| RxNorm | 847916 | exenatide 0.005 MG/ACTUAT [Byetta] |
| RxNorm | 847917 | 60 ACTUAT exenatide 0.005 MG/ACTUAT Pen Injector [Byetta] |

#### 8.2 Type II Diabetes Evidence

Table 9: Concept codes used to identify evidence of Type II diabetes.

| CodeSystem | ConceptCode | ConceptName |
|---|---|---|
| ICD10CM | E11.610 | Type 2 diabetes mellitus with diabetic neuropathic arthropathy |
| ICD10CM | E11.00 | Type 2 diabetes mellitus with hyperosmolarity without nonketotic hyperglycemic-hyperosmolar coma (NKHHC) |
| ICD10CM | E11.01 | Type 2 diabetes mellitus with hyperosmolarity with coma |
| ICD10CM | E11.10 | Type 2 diabetes mellitus with ketoacidosis without coma |
| ICD10CM | E11.11 | Type 2 diabetes mellitus with ketoacidosis with coma |
| ICD10CM | E11.21 | Type 2 diabetes mellitus with diabetic nephropathy |
| ICD10CM | E11.22 | Type 2 diabetes mellitus with diabetic chronic kidney disease |
| ICD10CM | E11.29 | Type 2 diabetes mellitus with other diabetic kidney complication |
| ICD10CM | E11.311 | Type 2 diabetes mellitus with unspecified diabetic retinopathy with macular edema |
| ICD10CM | E11.319 | Type 2 diabetes mellitus with unspecified diabetic retinopathy without macular edema |
| ICD10CM | E11.321 | Type 2 diabetes mellitus with mild nonproliferative diabetic retinopathy with macular edema |

Continued on next page

**Table9 – continued from previous page**

| CodeSystem | ConceptCode | ConceptName |
|---|---|---|
| ICD10CM | E11.3211 | Type 2 diabetes mellitus with mild nonproliferative diabetic retinopathy with macular edema, right eye |
| ICD10CM | E11.3212 | Type 2 diabetes mellitus with mild nonproliferative diabetic retinopathy with macular edema, left eye |
| ICD10CM | E11.3213 | Type 2 diabetes mellitus with mild nonproliferative diabetic retinopathy with macular edema, bilateral |
| ICD10CM | E11.3219 | Type 2 diabetes mellitus with mild nonproliferative diabetic retinopathy with macular edema, unspecified eye |
| ICD10CM | E11.329 | Type 2 diabetes mellitus with mild nonproliferative diabetic retinopathy without macular edema |
| ICD10CM | E11.3291 | Type 2 diabetes mellitus with mild nonproliferative diabetic retinopathy without macular edema, right eye |
| ICD10CM | E11.3292 | Type 2 diabetes mellitus with mild nonproliferative diabetic retinopathy without macular edema, left eye |
| ICD10CM | E11.3293 | Type 2 diabetes mellitus with mild nonproliferative diabetic retinopathy without macular edema, bilateral |
| ICD10CM | E11.3299 | Type 2 diabetes mellitus with mild nonproliferative diabetic retinopathy without macular edema, unspecified eye |
| ICD10CM | E11.331 | Type 2 diabetes mellitus with moderate nonproliferative diabetic retinopathy with macular edema |
| ICD10CM | E11.3311 | Type 2 diabetes mellitus with moderate nonproliferative diabetic retinopathy with macular edema, right eye |
| ICD10CM | E11.3312 | Type 2 diabetes mellitus with moderate nonproliferative diabetic retinopathy with macular edema, left eye |
| ICD10CM | E11.3313 | Type 2 diabetes mellitus with moderate nonproliferative diabetic retinopathy with macular edema, bilateral |
| ICD10CM | E11.3319 | Type 2 diabetes mellitus with moderate nonproliferative diabetic retinopathy with macular edema, unspecified eye |
| ICD10CM | E11.339 | Type 2 diabetes mellitus with moderate nonproliferative diabetic retinopathy without macular edema |
| ICD10CM | E11.3391 | Type 2 diabetes mellitus with moderate nonproliferative diabetic retinopathy without macular edema, right eye |
| ICD10CM | E11.3392 | Type 2 diabetes mellitus with moderate nonproliferative diabetic retinopathy without macular edema, left eye |
| ICD10CM | E11.3393 | Type 2 diabetes mellitus with moderate nonproliferative diabetic retinopathy without macular edema, bilateral |
| ICD10CM | E11.3399 | Type 2 diabetes mellitus with moderate nonproliferative diabetic retinopathy without macular edema, unspecified eye |
| ICD10CM | E11.341 | Type 2 diabetes mellitus with severe nonproliferative diabetic retinopathy with macular edema |
| ICD10CM | E11.3411 | Type 2 diabetes mellitus with severe nonproliferative diabetic retinopathy with macular edema, right eye |

Continued on next page

**Table9 – continued from previous page**

| CodeSystem | ConceptCode | ConceptName |
|---|---|---|
| ICD10CM | E11.3412 | Type 2 diabetes mellitus with severe nonproliferative diabetic retinopathy with macular edema, left eye |
| ICD10CM | E11.3413 | Type 2 diabetes mellitus with severe nonproliferative diabetic retinopathy with macular edema, bilateral |
| ICD10CM | E11.3419 | Type 2 diabetes mellitus with severe nonproliferative diabetic retinopathy with macular edema, unspecified eye |
| ICD10CM | E11.349 | Type 2 diabetes mellitus with severe nonproliferative diabetic retinopathy without macular edema |
| ICD10CM | E11.3491 | Type 2 diabetes mellitus with severe nonproliferative diabetic retinopathy without macular edema, right eye |
| ICD10CM | E11.3492 | Type 2 diabetes mellitus with severe nonproliferative diabetic retinopathy without macular edema, left eye |
| ICD10CM | E11.3493 | Type 2 diabetes mellitus with severe nonproliferative diabetic retinopathy without macular edema, bilateral |
| ICD10CM | E11.3499 | Type 2 diabetes mellitus with severe nonproliferative diabetic retinopathy without macular edema, unspecified eye |
| ICD10CM | E11.351 | Type 2 diabetes mellitus with proliferative diabetic retinopathy with macular edema |
| ICD10CM | E11.3511 | Type 2 diabetes mellitus with proliferative diabetic retinopathy with macular edema, right eye |
| ICD10CM | E11.3512 | Type 2 diabetes mellitus with proliferative diabetic retinopathy with macular edema, left eye |
| ICD10CM | E11.3513 | Type 2 diabetes mellitus with proliferative diabetic retinopathy with macular edema, bilateral |
| ICD10CM | E11.3519 | Type 2 diabetes mellitus with proliferative diabetic retinopathy with macular edema, unspecified eye |
| ICD10CM | E11.3521 | Type 2 diabetes mellitus with proliferative diabetic retinopathy with traction retinal detachment involving the macula, right eye |
| ICD10CM | E11.3522 | Type 2 diabetes mellitus with proliferative diabetic retinopathy with traction retinal detachment involving the macula, left eye |
| ICD10CM | E11.3523 | Type 2 diabetes mellitus with proliferative diabetic retinopathy with traction retinal detachment involving the macula, bilateral |
| ICD10CM | E11.3529 | Type 2 diabetes mellitus with proliferative diabetic retinopathy with traction retinal detachment involving the macula, unspecified eye |
| ICD10CM | E11.3531 | Type 2 diabetes mellitus with proliferative diabetic retinopathy with traction retinal detachment not involving the macula, right eye |

Continued on next page

**Table9 – continued from previous page**

| CodeSystem | ConceptCode | ConceptName |
|---|---|---|
| ICD10CM | E11.3532 | Type 2 diabetes mellitus with proliferative diabetic retinopathy with traction retinal detachment not involving the macula, left eye |
| ICD10CM | E11.3533 | Type 2 diabetes mellitus with proliferative diabetic retinopathy with traction retinal detachment not involving the macula, bilateral |
| ICD10CM | E11.3539 | Type 2 diabetes mellitus with proliferative diabetic retinopathy with traction retinal detachment not involving the macula, unspecified eye |
| ICD10CM | E11.3541 | Type 2 diabetes mellitus with proliferative diabetic retinopathy with combined traction retinal detachment and rhegmatogenous retinal detachment, right eye |
| ICD10CM | E11.3542 | Type 2 diabetes mellitus with proliferative diabetic retinopathy with combined traction retinal detachment and rhegmatogenous retinal detachment, left eye |
| ICD10CM | E11.3543 | Type 2 diabetes mellitus with proliferative diabetic retinopathy with combined traction retinal detachment and rhegmatogenous retinal detachment, bilateral |
| ICD10CM | E11.3549 | Type 2 diabetes mellitus with proliferative diabetic retinopathy with combined traction retinal detachment and rhegmatogenous retinal detachment, unspecified eye |
| ICD10CM | E11.3551 | Type 2 diabetes mellitus with stable proliferative diabetic retinopathy, right eye |
| ICD10CM | E11.3552 | Type 2 diabetes mellitus with stable proliferative diabetic retinopathy, left eye |
| ICD10CM | E11.3553 | Type 2 diabetes mellitus with stable proliferative diabetic retinopathy, bilateral |
| ICD10CM | E11.3559 | Type 2 diabetes mellitus with stable proliferative diabetic retinopathy, unspecified eye |
| ICD10CM | E11.359 | Type 2 diabetes mellitus with proliferative diabetic retinopathy without macular edema |
| ICD10CM | E11.3591 | Type 2 diabetes mellitus with proliferative diabetic retinopathy without macular edema, right eye |
| ICD10CM | E11.3592 | Type 2 diabetes mellitus with proliferative diabetic retinopathy without macular edema, left eye |
| ICD10CM | E11.3593 | Type 2 diabetes mellitus with proliferative diabetic retinopathy without macular edema, bilateral |
| ICD10CM | E11.3599 | Type 2 diabetes mellitus with proliferative diabetic retinopathy without macular edema, unspecified eye |
| ICD10CM | E11.36 | Type 2 diabetes mellitus with diabetic cataract |
| ICD10CM | E11.37X1 | Type 2 diabetes mellitus with diabetic macular edema, resolved following treatment, right eye |

Continued on next page

**Table9 – continued from previous page**

| CodeSystem | ConceptCode | ConceptName |
|---|---|---|
| ICD10CM | E11.37X2 | Type 2 diabetes mellitus with diabetic macular edema, resolved following treatment, left eye |
| ICD10CM | E11.37X3 | Type 2 diabetes mellitus with diabetic macular edema, resolved following treatment, bilateral |
| ICD10CM | E11.37X9 | Type 2 diabetes mellitus with diabetic macular edema, resolved following treatment, unspecified eye |
| ICD10CM | E11.39 | Type 2 diabetes mellitus with other diabetic ophthalmic complication |
| ICD10CM | E11.40 | Type 2 diabetes mellitus with diabetic neuropathy, unspecified |
| ICD10CM | E11.41 | Type 2 diabetes mellitus with diabetic mononeuropathy |
| ICD10CM | E11.42 | Type 2 diabetes mellitus with diabetic polyneuropathy |
| ICD10CM | E11.43 | Type 2 diabetes mellitus with diabetic autonomic (poly)neuropathy |
| ICD10CM | E11.44 | Type 2 diabetes mellitus with diabetic amyotrophy |
| ICD10CM | E11.49 | Type 2 diabetes mellitus with other diabetic neurological complication |
| ICD10CM | E11.51 | Type 2 diabetes mellitus with diabetic peripheral angiopathy without gangrene |
| ICD10CM | E11.52 | Type 2 diabetes mellitus with diabetic peripheral angiopathy with gangrene |
| ICD10CM | E11.59 | Type 2 diabetes mellitus with other circulatory complications |
| ICD10CM | E11.618 | Type 2 diabetes mellitus with other diabetic arthropathy |
| ICD10CM | E11.620 | Type 2 diabetes mellitus with diabetic dermatitis |
| ICD10CM | E11.621 | Type 2 diabetes mellitus with foot ulcer |
| ICD10CM | E11.622 | Type 2 diabetes mellitus with other skin ulcer |
| ICD10CM | E11.628 | Type 2 diabetes mellitus with other skin complications |
| ICD10CM | E11.630 | Type 2 diabetes mellitus with periodontal disease |
| ICD10CM | E11.638 | Type 2 diabetes mellitus with other oral complications |
| ICD10CM | E11.641 | Type 2 diabetes mellitus with hypoglycemia with coma |
| ICD10CM | E11.649 | Type 2 diabetes mellitus with hypoglycemia without coma |
| ICD10CM | E11.65 | Type 2 diabetes mellitus with hyperglycemia |
| ICD10CM | E11.69 | Type 2 diabetes mellitus with other specified complication |
| ICD10CM | E11.8 | Type 2 diabetes mellitus with unspecified complications |
| ICD10CM | E11.9 | Type 2 diabetes mellitus without complications |
| ICD10CM | O24.111 | Pre-existing type 2 diabetes mellitus, in pregnancy, first trimester |
| ICD10CM | O24.112 | Pre-existing type 2 diabetes mellitus, in pregnancy, second trimester |
| ICD10CM | O24.113 | Pre-existing type 2 diabetes mellitus, in pregnancy, third trimester |

Continued on next page

**Table9 – continued from previous page**

| CodeSystem | ConceptCode | ConceptName |
|---|---|---|
| ICD10CM | O24.119 | Pre-existing type 2 diabetes mellitus, in pregnancy, unspecified trimester |
| ICD10CM | O24.12 | Pre-existing type 2 diabetes mellitus, in childbirth |
| ICD10CM | O24.13 | Pre-existing type 2 diabetes mellitus, in the puerperium |
| ICD9CM | 250.00 | Diabetes mellitus without mention of complication, type II or unspecified type, not stated as uncontrolled |
| ICD9CM | 250.02 | Diabetes mellitus without mention of complication, type II or unspecified type, uncontrolled |
| ICD9CM | 250.10 | Diabetes with ketoacidosis, type II or unspecified type, not stated as uncontrolled |
| ICD9CM | 250.12 | Diabetes with ketoacidosis, type II or unspecified type, uncontrolled |
| ICD9CM | 250.20 | Diabetes with hyperosmolarity, type II or unspecified type, not stated as uncontrolled |
| ICD9CM | 250.22 | Diabetes with hyperosmolarity, type II or unspecified type, uncontrolled |
| ICD9CM | 250.30 | Diabetes with other coma, type II or unspecified type, not stated as uncontrolled |
| ICD9CM | 250.32 | Diabetes with other coma, type II or unspecified type, uncontrolled |
| ICD9CM | 250.40 | Diabetes with renal manifestations, type II or unspecified type, not stated as uncontrolled |
| ICD9CM | 250.42 | Diabetes with renal manifestations, type II or unspecified type, uncontrolled |
| ICD9CM | 250.50 | Diabetes with ophthalmic manifestations, type II or unspecified type, not stated as uncontrolled |
| ICD9CM | 250.52 | Diabetes with ophthalmic manifestations, type II or unspecified type, uncontrolled |
| ICD9CM | 250.60 | Diabetes with neurological manifestations, type II or unspecified type, not stated as uncontrolled |
| ICD9CM | 250.62 | Diabetes with neurological manifestations, type II or unspecified type, uncontrolled |
| ICD9CM | 250.70 | Diabetes with peripheral circulatory disorders, type II or unspecified type, not stated as uncontrolled |
| ICD9CM | 250.72 | Diabetes with peripheral circulatory disorders, type II or unspecified type, uncontrolled |
| ICD9CM | 250.80 | Diabetes with other specified manifestations, type II or unspecified type, not stated as uncontrolled |
| ICD9CM | 250.82 | Diabetes with other specified manifestations, type II or unspecified type, uncontrolled |
| ICD9CM | 250.90 | Diabetes with unspecified complication, type II or unspecified type, not stated as uncontrolled |

Continued on next page

**Table9 – continued from previous page**

| CodeSystem | ConceptCode | ConceptName |
|---|---|---|
| ICD9CM | 250.92 | Diabetes with unspecified complication, type II or unspecified type, uncontrolled |
| SNOMED CT | 1481000119100 | Diabetes mellitus type 2 without retinopathy |
| SNOMED CT | 1501000119109 | Proliferative retinopathy due to type 2 diabetes mellitus |
| SNOMED CT | 1511000119107 | Peripheral neuropathy due to type 2 diabetes mellitus |
| SNOMED CT | 1551000119108 | Nonproliferative retinopathy due to type 2 diabetes mellitus |
| SNOMED CT | 28331000119107 | Retinal edema due to type 2 diabetes mellitus |
| SNOMED CT | 41911000119107 | Glaucoma due to type 2 diabetes mellitus |
| SNOMED CT | 60951000119105 | Blindness due to type 2 diabetes mellitus |
| SNOMED CT | 71441000119104 | Nephrotic syndrome due to type 2 diabetes mellitus |
| SNOMED CT | 82541000119100 | Traction detachment of retina due to type 2 diabetes mellitus |
| SNOMED CT | 82551000119103 | Rubeosis iridis due to type 2 diabetes mellitus |
| SNOMED CT | 97331000119101 | Macular edema and retinopathy due to type 2 diabetes mellitus |
| SNOMED CT | 104961000119108 | Ischemia of retina due to type 2 diabetes mellitus |
| SNOMED CT | 110181000119105 | Peripheral sensory neuropathy due to type 2 diabetes mellitus |
| SNOMED CT | 138911000119106 | Mild nonproliferative retinopathy due to type 2 diabetes mellitus |
| SNOMED CT | 138921000119104 | Moderate nonproliferative retinopathy due to type 2 diabetes mellitus |
| SNOMED CT | 138941000119105 | Severe nonproliferative retinopathy due to diabetes mellitus type 2 |
| SNOMED CT | 368101000119109 | Diabetes mellitus type 2 with periodontal disease |
| SNOMED CT | 368581000119106 | Neuropathy due to type 2 diabetes mellitus |
| SNOMED CT | 87921000119104 | Cranial nerve palsy with type II diabetes mellitus |
| SNOMED CT | 81531005 | Type 2 diabetes mellitus in obese |
| SNOMED CT | 313436004 | Type 2 diabetes mellitus without complication |
| SNOMED CT | 314902007 | Type 2 diabetes mellitus with peripheral angiopathy |
| SNOMED CT | 314903002 | Arthropathy due to type 2 diabetes mellitus |
| SNOMED CT | 359642000 | Type 2 diabetes mellitus in nonobese |
| SNOMED CT | 237599002 | Insulin treated type 2 diabetes mellitus |
| SNOMED CT | 237604008 | Maturity onset diabetes of the young, type 2 |
| SNOMED CT | 609567009 | Pre-existing type 2 diabetes mellitus in pregnancy |
| SNOMED CT | 712883005 | Autonomic neuropathy due to type 2 diabetes mellitus |
| SNOMED CT | 713703005 | Gastroparesis due to type 2 diabetes mellitus |
| SNOMED CT | 713706002 | Polyneuropathy due to type 2 diabetes mellitus |
| SNOMED CT | 719216001 | Hypoglycemic coma due to type 2 diabetes mellitus |
| SNOMED CT | 190331003 | Hyperosmolar coma due to type 2 diabetes mellitus |
| SNOMED CT | 190389009 | Type 2 diabetes mellitus with ulcer |
| SNOMED CT | 199230006 | Pre-existing type 2 diabetes mellitus |
| SNOMED CT | 9859006 | Acanthosis nigricans due to type 2 diabetes mellitus |

Continued on next page

**Table9 – continued from previous page**

| CodeSystem | ConceptCode | ConceptName |
|---|---|---|
| SNOMED CT | 44054006 | Type 2 diabetes mellitus |
| SNOMED CT | 420279001 | Renal disorder due to type 2 diabetes mellitus |
| SNOMED CT | 420436000 | Mononeuropathy due to type 2 diabetes mellitus |
| SNOMED CT | 421326000 | Disorder of nervous system due to type 2 diabetes mellitus |
| SNOMED CT | 421779007 | Exudative maculopathy due to type 2 diabetes mellitus |
| SNOMED CT | 421847006 | Ketoacidotic coma due to type 2 diabetes mellitus |
| SNOMED CT | 422034002 | Retinopathy due to type 2 diabetes mellitus |
| SNOMED CT | 422099009 | Disorder of eye due to type 2 diabetes mellitus |
| SNOMED CT | 422166005 | Peripheral circulatory disorder due to type 2 diabetes mellitus |
| SNOMED CT | 427027005 | Lumbosacral radiculoplexus neuropathy due to type 2 diabetes mellitus |
| SNOMED CT | 428007007 | Erectile dysfunction due to type 2 diabetes mellitus |
| SNOMED CT | 1217044000 | Ketosis-resistant diabetes mellitus |
| SNOMED CT | 711000119100 | Chronic kidney disease stage 5 due to type 2 diabetes mellitus |
| SNOMED CT | 721000119107 | Chronic kidney disease stage 4 due to type 2 diabetes mellitus |
| SNOMED CT | 731000119105 | Chronic kidney disease stage 3 due to type 2 diabetes mellitus |
| SNOMED CT | 741000119101 | Chronic kidney disease stage 2 due to type 2 diabetes mellitus |
| SNOMED CT | 751000119104 | Chronic kidney disease stage 1 due to type 2 diabetes mellitus |
| SNOMED CT | 771000119108 | Chronic kidney disease due to type 2 diabetes mellitus |
| SNOMED CT | 781000119106 | Neuropathic arthropathy due to type 2 diabetes mellitus |
| SNOMED CT | 1491000119102 | Vitreous hemorrhage due to type 2 diabetes mellitus |
| SNOMED CT | 71421000119105 | Hypertension in chronic kidney disease due to type 2 diabetes mellitus |
| SNOMED CT | 90781000119102 | Microalbuminuria due to type 2 diabetes mellitus |
| SNOMED CT | 90791000119104 | End stage renal disease on dialysis due to type 2 diabetes mellitus |
| SNOMED CT | 97341000119105 | Proliferative retinopathy with retinal edema due to type 2 diabetes mellitus |
| SNOMED CT | 127991000119101 | Hypertension concurrent and due to end stage renal disease on dialysis due to type 2 diabetes mellitus |
| SNOMED CT | 140101000119109 | Hypertension in chronic kidney disease stage 5 due to type 2 diabetes mellitus |
| SNOMED CT | 140111000119107 | Hypertension in chronic kidney disease stage 4 due to type 2 diabetes mellitus |
| SNOMED CT | 140121000119100 | Hypertension in chronic kidney disease stage 3 due to type 2 diabetes mellitus |

Continued on next page

**Table9 – continued from previous page**

| CodeSystem | ConceptCode | ConceptName |
|---|---|---|
| SNOMED CT | 140131000119102 | Hypertension in chronic kidney disease stage 2 due to type 2 diabetes mellitus |
| SNOMED CT | 157141000119108 | Proteinuria due to type 2 diabetes mellitus |
| SNOMED CT | 368591000119109 | Cheiroarthropathy due to diabetes mellitus type 2 |
| SNOMED CT | 420756003 | Cataract of eye due to diabetes mellitus type 2 |
| SNOMED CT | 769220000 | Macular edema co-occurrent and due to type 2 diabetes mellitus |
| SNOMED CT | 237627000 | Pregnancy and type 2 diabetes mellitus |
| SNOMED CT | 1003605004 | Glaucoma suspect due to diabetes mellitus type 2 |
| SNOMED CT | 16745051000119107 | Nonproliferative retinopathy of right eye due to diabetes mellitus type 2 |
| SNOMED CT | 16745131000119100 | Nonproliferative retinopathy of left eye due to diabetes mellitus type 2 |
| SNOMED CT | 16745531000119108 | Mild nonproliferative retinopathy of left eye due to diabetes mellitus type 2 |
| SNOMED CT | 16745651000119101 | Mild nonproliferative retinopathy of right eye due to diabetes mellitus type 2 |
| SNOMED CT | 16745931000119102 | Bilateral mild nonproliferative retinopathy due to diabetes mellitus type 2 |
| SNOMED CT | 16746131000119105 | Moderate nonproliferative retinopathy of left eye due to diabetes mellitus type 2 |
| SNOMED CT | 16746261000119108 | Moderate nonproliferative retinopathy of right eye due to diabetes mellitus type 2 |
| SNOMED CT | 16746341000119103 | Bilateral moderate nonproliferative retinopathy due to diabetes mellitus type 2 |
| SNOMED CT | 16746901000119101 | Severe nonproliferative retinopathy of left eye due to diabetes mellitus type 2 |
| SNOMED CT | 16747021000119109 | Bilateral severe nonproliferative retinopathy of eyes due to diabetes mellitus type 2 |
| SNOMED CT | 16747141000119104 | Severe nonproliferative retinopathy of right eye due to diabetes mellitus type 2 |
| SNOMED CT | 16747661000119109 | Bilateral nonproliferative retinopathy due to diabetes mellitus type 2 |
| SNOMED CT | 16747901000119104 | Proliferative retinopathy of left eye due to diabetes mellitus type 2 |
| SNOMED CT | 16748141000119100 | Bilateral proliferative retinopathy due to diabetes mellitus type 2 |
| SNOMED CT | 16749661000119102 | Proliferative retinopathy of right eye due to diabetes mellitus type 2 |
| SNOMED CT | 703138006 | Type II diabetes mellitus in remission |
| SNOMED CT | 420715001 | Persistent microalbuminuria due to type 2 diabetes mellitus |
| SNOMED CT | 421986006 | Persistent proteinuria due to type 2 diabetes mellitus |
| SNOMED CT | 1196922005 | Neovascular glaucoma due to diabetes mellitus type 2 |

Continued on next page

**Table9 – continued from previous page**

| CodeSystem | ConceptCode | ConceptName |
|---|---|---|
| RxNorm | 25789 | glimepiride |
| RxNorm | 10633 | tolazamide |
| RxNorm | 4815 | glyburide |
| RxNorm | 4821 | glipizide |
| RxNorm | 1175658 | Avandaryl Oral Product |
| RxNorm | 1175659 | Avandaryl Pill |
| RxNorm | 1175878 | Diabeta Oral Product |
| RxNorm | 1175879 | Diabeta Pill |
| RxNorm | 1175880 | Diabetamide Oral Product |
| RxNorm | 1175881 | Diabetamide Pill |
| RxNorm | 1168162 | Daonil Oral Product |
| RxNorm | 1168163 | Daonil Pill |
| RxNorm | 1179291 | Minidiab Oral Product |
| RxNorm | 1179292 | Minidiab Pill |
| RxNorm | 1185049 | Metaglip Oral Product |
| RxNorm | 1179740 | Malix Oral Product |
| RxNorm | 1179741 | Malix Pill |
| RxNorm | 1177973 | Tolinase Oral Product |
| RxNorm | 1177974 | Tolinase Pill |
| RxNorm | 1186169 | Libanil Oral Product |
| RxNorm | 1186170 | Libanil Pill |
| RxNorm | 1178082 | Micronase Oral Product |
| RxNorm | 1178083 | Micronase Pill |
| RxNorm | 1178368 | Semi-Daonil Oral Product |
| RxNorm | 1178369 | Semi-Daonil Pill |
| RxNorm | 1185624 | Metaglip Pill |
| RxNorm | 1361492 | glimepiride 6 MG |
| RxNorm | 1361493 | glimepiride 6 MG Oral Tablet |
| RxNorm | 1361494 | glimepiride 8 MG |
| RxNorm | 1361495 | glimepiride 8 MG Oral Tablet |
| RxNorm | 151466 | Calabren |
| RxNorm | 151615 | Diabetamide |
| RxNorm | 151725 | Euglucon |
| RxNorm | 151822 | Glibenese |
| RxNorm | 153842 | glimepiride 3 MG Oral Tablet |
| RxNorm | 153843 | glimepiride 1 MG Oral Tablet [Amaryl] |
| RxNorm | 153844 | glimepiride 3 MG Oral Tablet [Amaryl] |
| RxNorm | 153845 | glimepiride 4 MG Oral Tablet [Amaryl] |
| RxNorm | 152651 | Malix |
| RxNorm | 153591 | glimepiride 2 MG Oral Tablet [Amaryl] |
| RxNorm | 153592 | Amaryl |
| RxNorm | 197737 | glyburide 1.25 MG Oral Tablet |
| RxNorm | 199245 | glimepiride 1 MG Oral Tablet |

Continued on next page

**Table9 – continued from previous page**

| CodeSystem | ConceptCode | ConceptName |
|---|---|---|
| RxNorm | 199246 | glimepiride 2 MG Oral Tablet |
| RxNorm | 199247 | glimepiride 4 MG Oral Tablet |
| RxNorm | 198291 | tolazamide 100 MG Oral Tablet |
| RxNorm | 198292 | tolazamide 250 MG Oral Tablet |
| RxNorm | 198293 | tolazamide 500 MG Oral Tablet |
| RxNorm | 203289 | Micronase |
| RxNorm | 203295 | Diabeta |
| RxNorm | 203296 | Daonil |
| RxNorm | 201921 | glipizide 5 MG Oral Tablet [Glibenese] |
| RxNorm | 201922 | glipizide 2.5 MG Oral Tablet [Minidiab] |
| RxNorm | 203679 | Minidiab |
| RxNorm | 203680 | Glucotrol |
| RxNorm | 201056 | glyburide 2.5 MG Oral Tablet [Libanil] |
| RxNorm | 201057 | glyburide 2.5 MG Oral Tablet [Semi-Daonil] |
| RxNorm | 201058 | glyburide 2.5 MG Oral Tablet [Euglucon] |
| RxNorm | 201059 | glyburide 2.5 MG Oral Tablet [Malix] |
| RxNorm | 201060 | glyburide 2.5 MG Oral Tablet [Diabetamide] |
| RxNorm | 201061 | glyburide 5 MG Oral Tablet [Libanil] |
| RxNorm | 201062 | glyburide 5 MG Oral Tablet [Euglucon] |
| RxNorm | 201063 | glyburide 5 MG Oral Tablet [Malix] |
| RxNorm | 201064 | glyburide 5 MG Oral Tablet [Diabetamide] |
| RxNorm | 205828 | glipizide 5 MG Oral Tablet [Glucotrol] |
| RxNorm | 205830 | glipizide 10 MG Oral Tablet [Glucotrol] |
| RxNorm | 205872 | glyburide 1.25 MG Oral Tablet [Diabeta] |
| RxNorm | 205873 | glyburide 1.25 MG Oral Tablet [Micronase] |
| RxNorm | 205875 | glyburide 2.5 MG Oral Tablet [Diabeta] |
| RxNorm | 205876 | glyburide 2.5 MG Oral Tablet [Micronase] |
| RxNorm | 205879 | glyburide 5 MG Oral Tablet [Diabeta] |
| RxNorm | 205880 | glyburide 5 MG Oral Tablet [Micronase] |
| RxNorm | 207953 | tolazamide 100 MG Oral Tablet [Tolinase] |
| RxNorm | 207954 | tolazamide 250 MG Oral Tablet [Tolinase] |
| RxNorm | 207955 | tolazamide 500 MG Oral Tablet [Tolinase] |
| RxNorm | 220338 | Tolinase |
| RxNorm | 225613 | Semi-Daonil |
| RxNorm | 246391 | glyburide 2.5 MG / phenformin 25 MG Oral Tablet |
| RxNorm | 246524 | glyburide 5 MG / phenformin 50 MG Oral Tablet |
| RxNorm | 250919 | glyburide 2.5 MG / metformin 400 MG Oral Tablet |
| RxNorm | 284743 | Glucovance |
| RxNorm | 261351 | glyburide 4.5 MG Oral Tablet [Glycron] |
| RxNorm | 252960 | glyburide 4.5 MG Oral Tablet |
| RxNorm | 285129 | glyburide / metformin |
| RxNorm | 261532 | Glycron |
| RxNorm | 261974 | glyburide 3 MG Oral Tablet [Glycron] |

Continued on next page

**Table9 – continued from previous page**

| CodeSystem | ConceptCode | ConceptName |
|---|---|---|
| RxNorm | 260286 | glyburide 1.5 MG Oral Tablet [Glycron] |
| RxNorm | 260287 | glyburide 6 MG Oral Tablet [Glycron] |
| RxNorm | 310488 | glipizide 10 MG Oral Tablet |
| RxNorm | 310489 | 24 HR glipizide 2.5 MG Extended Release Oral Tablet |
| RxNorm | 310490 | glipizide 5 MG Oral Tablet |
| RxNorm | 310534 | glyburide 2.5 MG Oral Tablet |
| RxNorm | 310536 | glyburide 3 MG Oral Tablet |
| RxNorm | 310537 | glyburide 5 MG Oral Tablet |
| RxNorm | 310539 | glyburide 6 MG Oral Tablet |
| RxNorm | 313418 | tolazamide 250 MG Oral Capsule |
| RxNorm | 314000 | glyburide 1.5 MG Oral Tablet |
| RxNorm | 314006 | 24 HR glipizide 5 MG Extended Release Oral Tablet |
| RxNorm | 315107 | 24 HR glipizide 10 MG Extended Release Oral Tablet |
| RxNorm | 316832 | tolazamide 100 MG |
| RxNorm | 316833 | tolazamide 250 MG |
| RxNorm | 316834 | tolazamide 500 MG |
| RxNorm | 315978 | glimepiride 1 MG |
| RxNorm | 315979 | glimepiride 4 MG |
| RxNorm | 315980 | glipizide 5 MG |
| RxNorm | 315987 | glyburide 1.25 MG |
| RxNorm | 315988 | glyburide 1.5 MG |
| RxNorm | 315989 | glyburide 2.5 MG |
| RxNorm | 315990 | glyburide 3 MG |
| RxNorm | 315991 | glyburide 5 MG |
| RxNorm | 315992 | glyburide 6 MG |
| RxNorm | 541898 | Glyburide 1.5 MG Oral Tablet [Glyguride] |
| RxNorm | 542065 | Glyburide 6 MG [Gyburide] |
| RxNorm | 575998 | Glipizide 2.5 MG / Metformin 250 MG [metaglip] |
| RxNorm | 575996 | Glipizide 2.5 MG / Metformin 500 MG [metaglip] |
| RxNorm | 541896 | Glyburide 1.5 MG [Glyguride] |
| RxNorm | 542067 | Glyburide 6 MG Oral Tablet [Gyburide] |
| RxNorm | 542066 | Glyburide Oral Tablet [Gyburide] |
| RxNorm | 541897 | Glyburide Oral Tablet [Glyguride] |
| RxNorm | 575997 | Glipizide 5 MG / Metformin 500 MG [metaglip] |
| RxNorm | 615211 | glimepiride / rosiglitazone Oral Tablet [Avandaryl] |
| RxNorm | 668563 | glimepiride 4 MG / pioglitazone 30 MG Oral Tablet [Duetact] |
| RxNorm | 706933 | glimepiride 2 MG / rosiglitazone 8 MG Oral Tablet [Avandaryl] |
| RxNorm | 668555 | glimepiride / pioglitazone Oral Tablet [Duetact] |
| RxNorm | 615918 | glimepiride 4 MG / rosiglitazone 4 MG [Avandaryl] |
| RxNorm | 615212 | glimepiride 1 MG / rosiglitazone 4 MG Oral Tablet [Avandaryl] |

Continued on next page

**Table9 – continued from previous page**

| CodeSystem | ConceptCode | ConceptName |
|---|---|---|
| RxNorm | 668554 | glimepiride 2 MG / pioglitazone 30 MG [Duetact] |
| RxNorm | 668562 | glimepiride 4 MG / pioglitazone 30 MG [Duetact] |
| RxNorm | 700836 | Glipizide 5 MG Extended Release Tablet |
| RxNorm | 706932 | glimepiride 2 MG / rosiglitazone 8 MG [Avandaryl] |
| RxNorm | 731456 | glimepiride / pioglitazone Oral Tablet [Duetact 30/4] |
| RxNorm | 615214 | glimepiride 2 MG / rosiglitazone 4 MG Oral Tablet [Avandaryl] |
| RxNorm | 615919 | glimepiride 4 MG / rosiglitazone 4 MG Oral Tablet [Avandaryl] |
| RxNorm | 615210 | glimepiride 1 MG / rosiglitazone 4 MG [Avandaryl] |
| RxNorm | 668556 | glimepiride 2 MG / pioglitazone 30 MG Oral Tablet [Duetact] |
| RxNorm | 847723 | glimepiride / rosiglitazone Oral Tablet [Avandaryl 8/4] |
| RxNorm | 847719 | glimepiride / rosiglitazone Oral Tablet [Avandaryl 8/2] |
| RxNorm | 707181 | glimepiride 4 MG / rosiglitazone 8 MG [Avandaryl] |
| RxNorm | 615213 | glimepiride 2 MG / rosiglitazone 4 MG [Avandaryl] |
| RxNorm | 707182 | glimepiride 4 MG / rosiglitazone 8 MG Oral Tablet [Avandaryl] |
| RxNorm | 849593 | Glipizide / Metformin Oral Tablet [Metaglip 2.5 MG/500 MG] |
| RxNorm | 847715 | glimepiride / rosiglitazone Oral Tablet [Avandaryl 4/1] |
| RxNorm | 847711 | glimepiride / rosiglitazone Oral Tablet [Avandaryl 4/2] |
| RxNorm | 849589 | Glipizide / Metformin Oral Tablet [Metaglip 5 MG/500 MG] |
| RxNorm | 861746 | Glyburide / Metformin Oral Tablet [Glucovance 1.25 MG/250 MG] |
| RxNorm | 861751 | Glyburide / Metformin Oral Tablet [Glucovance 2.5 MG/500 MG] |
| RxNorm | 1185627 | Metaglip 5 MG/500 MG Oral Product |
| RxNorm | 1175663 | Avandaryl 8/2 Pill |
| RxNorm | 1175657 | Avandaryl 4/1 Pill |
| RxNorm | 1175661 | Avandaryl 4/4 Pill |
| RxNorm | 1185628 | Metaglip 5 MG/500 MG Pill |
| RxNorm | 1175664 | Avandaryl 8/4 Oral Product |
| RxNorm | 1166405 | Duetact 30/4 Oral Product |
| RxNorm | 1185625 | Metaglip 2.5 MG/500 MG Oral Product |
| RxNorm | 1171251 | Glucovance 2.5 MG/500 MG Pill |
| RxNorm | 1166406 | Duetact 30/4 Pill |
| RxNorm | 1185626 | Metaglip 2.5 MG/500 MG Pill |
| RxNorm | 1175660 | Avandaryl 4/4 Oral Product |
| RxNorm | 1175662 | Avandaryl 8/2 Oral Product |
| RxNorm | 1171253 | Glucovance 5 MG/500 MG Pill |
| RxNorm | 1171250 | Glucovance 2.5 MG/500 MG Oral Product |

Continued on next page

**Table9 – continued from previous page**

| CodeSystem | ConceptCode | ConceptName |
|---|---|---|
| RxNorm | 1175665 | Avandaryl 8/4 Pill |
| RxNorm | 1171252 | Glucovance 5 MG/500 MG Oral Product |
| RxNorm | 1175656 | Avandaryl 4/1 Oral Product |
| RxNorm | 284433 | Glyburide 5 MG / Metformin 500 MG Oral Tablet [Glucovance] |
| RxNorm | 284432 | Glyburide 2.5 MG / Metformin 500 MG Oral Tablet [Glucovance] |
| RxNorm | 351273 | Glipizide 2.5 MG / Metformin 500 MG Oral Tablet |
| RxNorm | 351275 | Glipizide 2.5 MG / Metformin 250 MG Oral Tablet |
| RxNorm | 261311 | Glipizide 2.5 MG 24 Hour Extended Release Tablet [Glucotrol XL] |
| RxNorm | 205829 | Glipizide 5 MG 24 Hour Extended Release Tablet [Glucotrol XL] |
| RxNorm | 284431 | Glyburide 1.25 MG / Metformin 250 MG Oral Tablet [Glucovance] |
| RxNorm | 205881 | Glyburide 6 MG Oral Tablet [Glynase Pres-Tab] |
| RxNorm | 205877 | Glyburide 3 MG Oral Tablet [Glynase Pres-Tab] |
| RxNorm | 205831 | Glipizide 10 MG 24 Hour Extended Release Tablet [Glucotrol XL] |
| RxNorm | 205874 | Glyburide 1.5 MG Oral Tablet [Glynase Pres-Tab] |
| RxNorm | 351274 | Glipizide 5 MG / Metformin 500 MG Oral Tablet |
| RxNorm | 566719 | Glipizide 5 MG [Glucotrol XL] |
| RxNorm | 566721 | Glipizide 10 MG [Glucotrol XL] |
| RxNorm | 574533 | Glipizide 2.5 MG [Glucotrol XL] |
| RxNorm | 566763 | Glyburide 1.5 MG [Glynase Pres-Tab] |
| RxNorm | 574869 | Glyburide 1.25 MG / Metformin 250 MG [Glucovance] |
| RxNorm | 566770 | Glyburide 6 MG [Glynase Pres-Tab] |
| RxNorm | 574871 | Glyburide 5 MG / Metformin 500 MG [Glucovance] |
| RxNorm | 566766 | Glyburide 3 MG [Glynase Pres-Tab] |
| RxNorm | 574870 | Glyburide 2.5 MG / Metformin 500 MG [Glucovance] |
| RxNorm | 310538 | Glyburide 5 MG / Metformin 500 MG Oral Tablet |
| RxNorm | 310535 | Glyburide 2.5 MG / Metformin 500 MG Oral Tablet |
| RxNorm | 310533 | Glyburide 1.25 MG / Metformin 250 MG Oral Tablet |
| RxNorm | 368567 | Glipizide / Metformin Oral Tablet [metaglip] |
| RxNorm | 352301 | Glipizide 5 MG / Metformin 500 MG Oral Tablet [metaglip] |
| RxNorm | 352302 | Glipizide 2.5 MG / Metformin 250 MG Oral Tablet [metaglip] |
| RxNorm | 379801 | Glipizide 24 Hour Extended Release Tablet [Glucotrol XL] |
| RxNorm | 369372 | Glyburide Oral Tablet [Glynase Pres-Tab] |
| RxNorm | 379805 | Glipizide 24 Hour Extended Release Tablet |
| RxNorm | 368169 | Glyburide / Metformin Oral Tablet [Glucovance] |
| RxNorm | 352300 | Glipizide 2.5 MG / Metformin 500 MG Oral Tablet [metaglip] |

Continued on next page

**Table9 – continued from previous page**

| CodeSystem | ConceptCode | ConceptName |
|---|---|---|
| RxNorm | 1008873 | glyburide / phenformin |
| RxNorm | 102845 | glyburide 5 MG Oral Tablet [Daonil] |
| RxNorm | 105371 | glyburide 2.5 MG Oral Tablet [Calabren] |
| RxNorm | 105372 | glyburide 5 MG Oral Tablet [Calabren] |
| RxNorm | 105373 | glipizide 5 MG Oral Tablet [Minidiab] |
| RxNorm | 1157121 | tolazamide Oral Product |
| RxNorm | 1157122 | tolazamide Pill |
| RxNorm | 1156197 | glyburide / metformin Pill |
| RxNorm | 1156198 | glyburide / phenformin Oral Product |
| RxNorm | 1156199 | glyburide / phenformin Pill |
| RxNorm | 1156200 | glyburide Oral Product |
| RxNorm | 1156201 | glyburide Pill |
| RxNorm | 1157240 | glimepiride / pioglitazone Oral Product |
| RxNorm | 1157241 | glimepiride / pioglitazone Pill |
| RxNorm | 1157242 | glimepiride / rosiglitazone Oral Product |
| RxNorm | 1157243 | glimepiride / rosiglitazone Pill |
| RxNorm | 1157244 | glimepiride Oral Product |
| RxNorm | 1157245 | glimepiride Pill |
| RxNorm | 1166403 | Duetact Oral Product |
| RxNorm | 1166404 | Duetact Pill |
| RxNorm | 1165205 | glipizide / metformin Oral Product |
| RxNorm | 1165206 | glipizide / metformin Pill |
| RxNorm | 1165207 | glipizide Oral Product |
| RxNorm | 1165208 | glipizide Pill |
| RxNorm | 1165845 | glyburide / metformin Oral Product |
| RxNorm | 1170663 | Amaryl Oral Product |
| RxNorm | 1170664 | Amaryl Pill |
| RxNorm | 1170866 | Calabren Oral Product |
| RxNorm | 1170867 | Calabren Pill |
| RxNorm | 1171148 | Euglucon Oral Product |
| RxNorm | 1171149 | Euglucon Pill |
| RxNorm | 1171233 | Glibenese Oral Product |
| RxNorm | 1171234 | Glibenese Pill |
| RxNorm | 1171246 | Glucotrol Oral Product |
| RxNorm | 1171247 | Glucotrol Pill |
| RxNorm | 1171248 | Glucovance Oral Product |
| RxNorm | 1171249 | Glucovance Pill |
| RxNorm | 1171929 | Glycron Oral Product |
| RxNorm | 1171930 | Glycron Pill |
| RxNorm | 1171933 | Glynase Oral Product |
| RxNorm | 1171934 | Glynase Pill |
| RxNorm | 1171919 | Glyburase Oral Product |
| RxNorm | 1171920 | Glyburase Pill |

Continued on next page

**Table9 – continued from previous page**

| CodeSystem | ConceptCode | ConceptName |
|---|---|---|
| RxNorm | 317379 | glipizide 10 MG |
| RxNorm | 317637 | glimepiride 2 MG |
| RxNorm | 331496 | glyburide 4.5 MG |
| RxNorm | 330349 | glipizide 2.5 MG |
| RxNorm | 352381 | glipizide / metformin |
| RxNorm | 352764 | Metaglip |
| RxNorm | 367762 | glimepiride Oral Tablet [Amaryl] |
| RxNorm | 368204 | glyburide Oral Tablet [Glycron] |
| RxNorm | 369304 | tolazamide Oral Tablet [Tolinase] |
| RxNorm | 369373 | glyburide Oral Tablet [Diabeta] |
| RxNorm | 369500 | glipizide Oral Tablet [Glucotrol] |
| RxNorm | 369557 | glyburide Oral Tablet [Semi-Daonil] |
| RxNorm | 369562 | glyburide Oral Tablet [Daonil] |
| RxNorm | 374635 | glyburide / metformin Oral Tablet |
| RxNorm | 375952 | tolazamide Oral Capsule |
| RxNorm | 379803 | glipizide Oral Tablet [Glibenese] |
| RxNorm | 379804 | glipizide 2.5 MG Oral Tablet |
| RxNorm | 380849 | glimepiride 3 MG |
| RxNorm | 372319 | glimepiride Oral Tablet |
| RxNorm | 372320 | glipizide Oral Tablet |
| RxNorm | 372333 | glyburide Oral Tablet |
| RxNorm | 372334 | glyburide / phenformin Oral Tablet |
| RxNorm | 378730 | glipizide / metformin Oral Tablet |
| RxNorm | 374149 | tolazamide Oral Tablet |
| RxNorm | 379559 | glyburide Oral Tablet [Euglucon] |
| RxNorm | 379565 | glyburide Oral Tablet [Diabetamide] |
| RxNorm | 379568 | glyburide Oral Tablet [Malix] |
| RxNorm | 379570 | glyburide Oral Tablet [Calabren] |
| RxNorm | 379571 | Libanil |
| RxNorm | 379572 | glyburide Oral Tablet [Libanil] |
| RxNorm | 379802 | glipizide Oral Tablet [Minidiab] |
| RxNorm | 429841 | glyburide 5 MG / metformin 1000 MG Oral Tablet |
| RxNorm | 430102 | glyburide 3.5 MG Oral Tablet |
| RxNorm | 430103 | glyburide 1.75 MG Oral Tablet |
| RxNorm | 430104 | glyburide 1 MG Oral Tablet |
| RxNorm | 440285 | glyburide 1.75 MG |
| RxNorm | 440286 | glyburide 1 MG |
| RxNorm | 440287 | glyburide 3.5 MG |
| RxNorm | 563154 | glyburide 5 MG [Daonil] |
| RxNorm | 566056 | glipizide 5 MG [Glibenese] |
| RxNorm | 566057 | glipizide 2.5 MG [Minidiab] |
| RxNorm | 565327 | glimepiride 2 MG [Amaryl] |
| RxNorm | 565408 | glimepiride 1 MG [Amaryl] |

Continued on next page

**Table9 – continued from previous page**

| CodeSystem | ConceptCode | ConceptName |
|---|---|---|
| RxNorm | 565409 | glimepiride 3 MG [Amaryl] |
| RxNorm | 565410 | glimepiride 4 MG [Amaryl] |
| RxNorm | 568684 | tolazamide 100 MG [Tolinase] |
| RxNorm | 568685 | tolazamide 250 MG [Tolinase] |
| RxNorm | 568686 | tolazamide 500 MG [Tolinase] |
| RxNorm | 564036 | glyburide 2.5 MG [Calabren] |
| RxNorm | 564037 | glyburide 5 MG [Calabren] |
| RxNorm | 564038 | glipizide 5 MG [Minidiab] |
| RxNorm | 566718 | glipizide 5 MG [Glucotrol] |
| RxNorm | 566720 | glipizide 10 MG [Glucotrol] |
| RxNorm | 566761 | glyburide 1.25 MG [Diabeta] |
| RxNorm | 566762 | glyburide 1.25 MG [Micronase] |
| RxNorm | 566764 | glyburide 2.5 MG [Diabeta] |
| RxNorm | 566765 | glyburide 2.5 MG [Micronase] |
| RxNorm | 566768 | glyburide 5 MG [Diabeta] |
| RxNorm | 566769 | glyburide 5 MG [Micronase] |
| RxNorm | 565667 | glyburide 2.5 MG [Libanil] |
| RxNorm | 565668 | glyburide 2.5 MG [Semi-Daonil] |
| RxNorm | 565669 | glyburide 2.5 MG [Euglucon] |
| RxNorm | 565670 | glyburide 2.5 MG [Malix] |
| RxNorm | 565671 | glyburide 2.5 MG [Diabetamide] |
| RxNorm | 565672 | glyburide 5 MG [Libanil] |
| RxNorm | 565673 | glyburide 5 MG [Euglucon] |
| RxNorm | 565674 | glyburide 5 MG [Malix] |
| RxNorm | 565675 | glyburide 5 MG [Diabetamide] |
| RxNorm | 574612 | glyburide 3 MG [Glycron] |
| RxNorm | 606253 | glimepiride / rosiglitazone |
| RxNorm | 602543 | glimepiride / rosiglitazone Oral Tablet |
| RxNorm | 602544 | glimepiride 1 MG / rosiglitazone 4 MG Oral Tablet |
| RxNorm | 602549 | glimepiride 2 MG / rosiglitazone 4 MG Oral Tablet |
| RxNorm | 602550 | glimepiride 4 MG / rosiglitazone 4 MG Oral Tablet |
| RxNorm | 607816 | Avandaryl |
| RxNorm | 574089 | glyburide 1.5 MG [Glycron] |
| RxNorm | 574090 | glyburide 6 MG [Glycron] |
| RxNorm | 574571 | glyburide 4.5 MG [Glycron] |
| RxNorm | 669980 | Glyburase |
| RxNorm | 669981 | glyburide 1.25 MG [Glyburase] |
| RxNorm | 669982 | glyburide Oral Tablet [Glyburase] |
| RxNorm | 669983 | glyburide 1.25 MG Oral Tablet [Glyburase] |
| RxNorm | 669984 | glyburide 2.5 MG [Glyburase] |
| RxNorm | 669985 | glyburide 2.5 MG Oral Tablet [Glyburase] |
| RxNorm | 669986 | glyburide 5 MG [Glyburase] |
| RxNorm | 669987 | glyburide 5 MG Oral Tablet [Glyburase] |

Continued on next page

**Table9 – continued from previous page**

| CodeSystem | ConceptCode | ConceptName |
|---|---|---|
| RxNorm | 647208 | Duetact |
| RxNorm | 647235 | glimepiride / pioglitazone |
| RxNorm | 647236 | glimepiride / pioglitazone Oral Tablet |
| RxNorm | 647237 | glimepiride 2 MG / pioglitazone 30 MG Oral Tablet |
| RxNorm | 647239 | glimepiride 4 MG / pioglitazone 30 MG Oral Tablet |
| RxNorm | 700835 | glipizide Extended Release Oral Tablet |
| RxNorm | 706895 | glimepiride 2 MG / rosiglitazone 8 MG Oral Tablet |
| RxNorm | 706896 | glimepiride 4 MG / rosiglitazone 8 MG Oral Tablet |
| RxNorm | 731455 | glimepiride 4 MG / pioglitazone 30 MG [Duetact] |
| RxNorm | 731457 | glimepiride 4 MG / pioglitazone 30 MG Oral Tablet [Duetact] |
| RxNorm | 731461 | glimepiride 2 MG / pioglitazone 30 MG [Duetact] |
| RxNorm | 731462 | glimepiride / pioglitazone Oral Tablet [Duetact] |
| RxNorm | 731463 | glimepiride 2 MG / pioglitazone 30 MG Oral Tablet [Duetact] |
| RxNorm | 844809 | glipizide 2.5 MG Extended Release Oral Tablet |
| RxNorm | 844824 | glipizide 5 MG Extended Release Oral Tablet |
| RxNorm | 844827 | glipizide 10 MG Extended Release Oral Tablet |
| RxNorm | 849585 | glipizide / metformin Oral Tablet [Metaglip] |
| RxNorm | 847708 | glimepiride 4 MG / rosiglitazone 4 MG Oral Tablet [Avandaryl] |
| RxNorm | 847706 | glimepiride 4 MG / rosiglitazone 4 MG [Avandaryl] |
| RxNorm | 847707 | glimepiride / rosiglitazone Oral Tablet [Avandaryl] |
| RxNorm | 847710 | glimepiride 2 MG / rosiglitazone 4 MG [Avandaryl] |
| RxNorm | 847712 | glimepiride 2 MG / rosiglitazone 4 MG Oral Tablet [Avandaryl] |
| RxNorm | 847714 | glimepiride 1 MG / rosiglitazone 4 MG [Avandaryl] |
| RxNorm | 847716 | glimepiride 1 MG / rosiglitazone 4 MG Oral Tablet [Avandaryl] |
| RxNorm | 847718 | glimepiride 2 MG / rosiglitazone 8 MG [Avandaryl] |
| RxNorm | 847720 | glimepiride 2 MG / rosiglitazone 8 MG Oral Tablet [Avandaryl] |
| RxNorm | 847722 | glimepiride 4 MG / rosiglitazone 8 MG [Avandaryl] |
| RxNorm | 847724 | glimepiride 4 MG / rosiglitazone 8 MG Oral Tablet [Avandaryl] |
| RxNorm | 861731 | glipizide 2.5 MG / metformin hydrochloride 250 MG Oral Tablet |
| RxNorm | 861732 | glipizide 2.5 MG / metformin hydrochloride 250 MG [Metaglip] |
| RxNorm | 861733 | glipizide 2.5 MG / metformin hydrochloride 250 MG Oral Tablet [Metaglip] |
| RxNorm | 861736 | glipizide 2.5 MG / metformin hydrochloride 500 MG Oral Tablet |

Continued on next page

**Table9 – continued from previous page**

| CodeSystem | ConceptCode | ConceptName |
|---|---|---|
| RxNorm | 861737 | glipizide 2.5 MG / metformin hydrochloride 500 MG [Metaglip] |
| RxNorm | 861738 | glipizide 2.5 MG / metformin hydrochloride 500 MG Oral Tablet [Metaglip] |
| RxNorm | 861740 | glipizide 5 MG / metformin hydrochloride 500 MG Oral Tablet |
| RxNorm | 861741 | glipizide 5 MG / metformin hydrochloride 500 MG [Metaglip] |
| RxNorm | 861742 | glipizide 5 MG / metformin hydrochloride 500 MG Oral Tablet [Metaglip] |
| RxNorm | 861743 | glyburide 1.25 MG / metformin hydrochloride 250 MG Oral Tablet |
| RxNorm | 861745 | glyburide 1.25 MG / metformin hydrochloride 250 MG [Glucovance] |
| RxNorm | 861747 | glyburide 1.25 MG / metformin hydrochloride 250 MG Oral Tablet [Glucovance] |
| RxNorm | 861748 | glyburide 2.5 MG / metformin hydrochloride 500 MG Oral Tablet |
| RxNorm | 861750 | glyburide 2.5 MG / metformin hydrochloride 500 MG [Glucovance] |
| RxNorm | 861752 | glyburide 2.5 MG / metformin hydrochloride 500 MG Oral Tablet [Glucovance] |
| RxNorm | 861753 | glyburide 5 MG / metformin hydrochloride 500 MG Oral Tablet |
| RxNorm | 861755 | glyburide 5 MG / metformin hydrochloride 500 MG [Glucovance] |
| RxNorm | 861756 | glyburide / metformin Oral Tablet [Glucovance] |
| RxNorm | 861757 | glyburide 5 MG / metformin hydrochloride 500 MG Oral Tablet [Glucovance] |
| RxNorm | 881404 | Glynase |
| RxNorm | 881405 | glyburide 1.5 MG [Glynase] |
| RxNorm | 881406 | glyburide Oral Tablet [Glynase] |
| RxNorm | 881407 | glyburide 1.5 MG Oral Tablet [Glynase] |
| RxNorm | 881408 | glyburide 3 MG [Glynase] |
| RxNorm | 881409 | glyburide 3 MG Oral Tablet [Glynase] |
| RxNorm | 881410 | glyburide 6 MG [Glynase] |
| RxNorm | 881411 | glyburide 6 MG Oral Tablet [Glynase] |
| RxNorm | 865567 | glipizide Extended Release Oral Tablet [Glucotrol] |
| RxNorm | 865568 | 24 HR glipizide 10 MG Extended Release Oral Tablet [Glucotrol] |
| RxNorm | 865569 | glipizide 10 MG Extended Release Oral Tablet [Glucotrol] |
| RxNorm | 865570 | glipizide 2.5 MG [Glucotrol] |

Continued on next page

**Table9 – continued from previous page**

| CodeSystem | ConceptCode | ConceptName |
|---|---|---|
| RxNorm | 865571 | 24 HR glipizide 2.5 MG Extended Release Oral Tablet [Glucotrol] |
| RxNorm | 865572 | glipizide 2.5 MG Extended Release Oral Tablet [Glucotrol] |
| RxNorm | 865573 | 24 HR glipizide 5 MG Extended Release Oral Tablet [Glucotrol] |
| RxNorm | 865574 | glipizide 5 MG Extended Release Oral Tablet [Glucotrol] |
| RxNorm | 93312 | glyburide Oral Tablet [Micronase] |
| RxNorm | 1368001 | alogliptin |
| RxNorm | 1546030 | saxagliptin anhydrous |
| RxNorm | 1100699 | linagliptin |
| RxNorm | 593411 | sitagliptin |
| RxNorm | 1172860 | Kombiglyze Pill |
| RxNorm | 1172861 | Kombiglyze Oral Product |
| RxNorm | 1167810 | Janumet Oral Product |
| RxNorm | 1167811 | Janumet Pill |
| RxNorm | 1167814 | Januvia Oral Product |
| RxNorm | 1167815 | Januvia Pill |
| RxNorm | 1179163 | Tradjenta Oral Product |
| RxNorm | 1179164 | Tradjenta Pill |
| RxNorm | 1189800 | simvastatin / sitagliptin Oral Product |
| RxNorm | 1189801 | simvastatin / sitagliptin Pill |
| RxNorm | 1189802 | simvastatin / sitagliptin Oral Tablet |
| RxNorm | 1189803 | simvastatin / sitagliptin |
| RxNorm | 1189804 | simvastatin 10 MG / sitagliptin 100 MG Oral Tablet |
| RxNorm | 1189806 | simvastatin 10 MG / sitagliptin 100 MG [Juvisync] |
| RxNorm | 1189808 | simvastatin 20 MG / sitagliptin 100 MG Oral Tablet |
| RxNorm | 1189810 | simvastatin 20 MG / sitagliptin 100 MG [Juvisync] |
| RxNorm | 1189811 | simvastatin / sitagliptin Oral Tablet [Juvisync] |
| RxNorm | 1189812 | Juvisync Oral Product |
| RxNorm | 1189813 | Juvisync Pill |
| RxNorm | 1189814 | simvastatin 20 MG / sitagliptin 100 MG Oral Tablet [Juvisync] |
| RxNorm | 1189818 | simvastatin 10 MG / sitagliptin 100 MG Oral Tablet [Juvisync] |
| RxNorm | 1189821 | simvastatin 40 MG / sitagliptin 100 MG Oral Tablet |
| RxNorm | 1189823 | simvastatin 40 MG / sitagliptin 100 MG [Juvisync] |
| RxNorm | 1189827 | simvastatin 40 MG / sitagliptin 100 MG Oral Tablet [Juvisync] |
| RxNorm | 1243015 | linagliptin 2.5 MG |
| RxNorm | 1243016 | linagliptin / metformin Oral Product |
| RxNorm | 1243017 | linagliptin / metformin Pill |
| RxNorm | 1243018 | linagliptin / metformin Oral Tablet |
| RxNorm | 1243019 | linagliptin / metformin |

Continued on next page

**Table9 – continued from previous page**

| CodeSystem | ConceptCode | ConceptName |
|---|---|---|
| RxNorm | 1243020 | linagliptin 2.5 MG / metformin hydrochloride 1000 MG Oral Tablet |
| RxNorm | 1243022 | linagliptin 2.5 MG / metformin hydrochloride 1000 MG [Jentadueto] |
| RxNorm | 1243026 | linagliptin 2.5 MG / metformin hydrochloride 1000 MG Oral Tablet [Jentadueto] |
| RxNorm | 1243027 | linagliptin 2.5 MG / metformin hydrochloride 500 MG Oral Tablet |
| RxNorm | 1243029 | linagliptin 2.5 MG / metformin hydrochloride 500 MG [Jentadueto] |
| RxNorm | 1243033 | linagliptin 2.5 MG / metformin hydrochloride 500 MG Oral Tablet [Jentadueto] |
| RxNorm | 1243034 | linagliptin 2.5 MG / metformin hydrochloride 850 MG Oral Tablet |
| RxNorm | 1243036 | linagliptin 2.5 MG / metformin hydrochloride 850 MG [Jentadueto] |
| RxNorm | 1243037 | linagliptin / metformin Oral Tablet [Jentadueto] |
| RxNorm | 1243038 | Jentadueto Oral Product |
| RxNorm | 1243039 | Jentadueto Pill |
| RxNorm | 1243040 | linagliptin 2.5 MG / metformin hydrochloride 850 MG Oral Tablet [Jentadueto] |
| RxNorm | 1243826 | metformin / sitagliptin Extended Release Oral Tablet |
| RxNorm | 1243827 | 24 HR metformin hydrochloride 1000 MG / sitagliptin 100 MG Extended Release Oral Tablet |
| RxNorm | 1243829 | metformin hydrochloride 1000 MG / sitagliptin 100 MG [Janumet] |
| RxNorm | 1243833 | 24 HR metformin hydrochloride 1000 MG / sitagliptin 100 MG Extended Release Oral Tablet [Janumet] |
| RxNorm | 1243834 | metformin hydrochloride 1000 MG / sitagliptin 100 MG Extended Release Oral Tablet |
| RxNorm | 1243835 | metformin hydrochloride 1000 MG / sitagliptin 100 MG Extended Release Oral Tablet [Janumet] |
| RxNorm | 1243839 | metformin / sitagliptin Extended Release Oral Tablet [Janumet] |
| RxNorm | 1243842 | 24 HR metformin hydrochloride 1000 MG / sitagliptin 50 MG Extended Release Oral Tablet |
| RxNorm | 1243843 | 24 HR metformin hydrochloride 1000 MG / sitagliptin 50 MG Extended Release Oral Tablet [Janumet] |
| RxNorm | 1243844 | metformin hydrochloride 1000 MG / sitagliptin 50 MG Extended Release Oral Tablet |
| RxNorm | 1243845 | metformin hydrochloride 1000 MG / sitagliptin 50 MG Extended Release Oral Tablet [Janumet] |

Continued on next page

**Table9 – continued from previous page**

| CodeSystem | ConceptCode | ConceptName |
|---|---|---|
| RxNorm | 1243846 | 24 HR metformin hydrochloride 500 MG / sitagliptin 50 MG Extended Release Oral Tablet |
| RxNorm | 1243848 | 24 HR metformin hydrochloride 500 MG / sitagliptin 50 MG Extended Release Oral Tablet [Janumet] |
| RxNorm | 1243849 | metformin hydrochloride 500 MG / sitagliptin 50 MG Extended Release Oral Tablet |
| RxNorm | 1243850 | metformin hydrochloride 500 MG / sitagliptin 50 MG Extended Release Oral Tablet [Janumet] |
| RxNorm | 1312409 | simvastatin 10 MG / sitagliptin 50 MG Oral Tablet |
| RxNorm | 1312411 | simvastatin 10 MG / sitagliptin 50 MG [Juvisync] |
| RxNorm | 1312415 | simvastatin 10 MG / sitagliptin 50 MG Oral Tablet [Juvisync] |
| RxNorm | 1312416 | simvastatin 20 MG / sitagliptin 50 MG Oral Tablet |
| RxNorm | 1312418 | simvastatin 20 MG / sitagliptin 50 MG [Juvisync] |
| RxNorm | 1312422 | simvastatin 20 MG / sitagliptin 50 MG Oral Tablet [Juvisync] |
| RxNorm | 1312423 | simvastatin 40 MG / sitagliptin 50 MG Oral Tablet |
| RxNorm | 1312425 | simvastatin 40 MG / sitagliptin 50 MG [Juvisync] |
| RxNorm | 1312429 | simvastatin 40 MG / sitagliptin 50 MG Oral Tablet [Juvisync] |
| RxNorm | 1368002 | alogliptin 25 MG |
| RxNorm | 1368003 | alogliptin Oral Product |
| RxNorm | 1368004 | alogliptin Pill |
| RxNorm | 1368005 | alogliptin Oral Tablet |
| RxNorm | 1368006 | alogliptin 25 MG Oral Tablet |
| RxNorm | 1368007 | Nesina |
| RxNorm | 1368008 | alogliptin 25 MG [Nesina] |
| RxNorm | 1368009 | alogliptin Oral Tablet [Nesina] |
| RxNorm | 1368010 | Nesina Oral Product |
| RxNorm | 1368011 | Nesina Pill |
| RxNorm | 1368012 | alogliptin 25 MG Oral Tablet [Nesina] |
| RxNorm | 1368017 | alogliptin 6.25 MG |
| RxNorm | 1368018 | alogliptin 6.25 MG Oral Tablet |
| RxNorm | 1368019 | alogliptin 6.25 MG [Nesina] |
| RxNorm | 1368020 | alogliptin 6.25 MG Oral Tablet [Nesina] |
| RxNorm | 1368033 | alogliptin 12.5 MG |
| RxNorm | 1368034 | alogliptin 12.5 MG Oral Tablet |
| RxNorm | 1368035 | alogliptin 12.5 MG [Nesina] |
| RxNorm | 1368036 | alogliptin 12.5 MG Oral Tablet [Nesina] |
| RxNorm | 1368381 | alogliptin / metformin Oral Product |
| RxNorm | 1368382 | alogliptin / metformin Pill |
| RxNorm | 1368383 | alogliptin / metformin Oral Tablet |
| RxNorm | 1368384 | alogliptin / metformin |

Continued on next page

**Table9 – continued from previous page**

| CodeSystem | ConceptCode | ConceptName |
|---|---|---|
| RxNorm | 1368385 | alogliptin 12.5 MG / metformin hydrochloride 1000 MG Oral Tablet |
| RxNorm | 1368387 | alogliptin 12.5 MG / metformin hydrochloride 1000 MG [Kazano] |
| RxNorm | 1368391 | alogliptin 12.5 MG / metformin hydrochloride 1000 MG Oral Tablet [Kazano] |
| RxNorm | 1368392 | alogliptin 12.5 MG / metformin hydrochloride 500 MG Oral Tablet |
| RxNorm | 1368394 | alogliptin 12.5 MG / metformin hydrochloride 500 MG [Kazano] |
| RxNorm | 1368395 | alogliptin / metformin Oral Tablet [Kazano] |
| RxNorm | 1368396 | Kazano Oral Product |
| RxNorm | 1368397 | Kazano Pill |
| RxNorm | 1368398 | alogliptin 12.5 MG / metformin hydrochloride 500 MG Oral Tablet [Kazano] |
| RxNorm | 1368399 | alogliptin / pioglitazone Oral Product |
| RxNorm | 1368400 | alogliptin / pioglitazone Pill |
| RxNorm | 1368401 | alogliptin / pioglitazone Oral Tablet |
| RxNorm | 1368402 | alogliptin / pioglitazone |
| RxNorm | 1368403 | alogliptin 12.5 MG / pioglitazone 15 MG Oral Tablet |
| RxNorm | 1368405 | alogliptin 12.5 MG / pioglitazone 15 MG [Oseni] |
| RxNorm | 1368409 | alogliptin 12.5 MG / pioglitazone 15 MG Oral Tablet [Oseni] |
| RxNorm | 1368410 | alogliptin 12.5 MG / pioglitazone 30 MG Oral Tablet |
| RxNorm | 1368412 | alogliptin 12.5 MG / pioglitazone 30 MG [Oseni] |
| RxNorm | 1368416 | alogliptin 12.5 MG / pioglitazone 30 MG Oral Tablet [Oseni] |
| RxNorm | 1368417 | alogliptin 12.5 MG / pioglitazone 45 MG Oral Tablet |
| RxNorm | 1368419 | alogliptin 12.5 MG / pioglitazone 45 MG [Oseni] |
| RxNorm | 1368423 | alogliptin 12.5 MG / pioglitazone 45 MG Oral Tablet [Oseni] |
| RxNorm | 1368424 | alogliptin 25 MG / pioglitazone 15 MG Oral Tablet |
| RxNorm | 1368426 | alogliptin 25 MG / pioglitazone 15 MG [Oseni] |
| RxNorm | 1368430 | alogliptin 25 MG / pioglitazone 15 MG Oral Tablet [Oseni] |
| RxNorm | 1368431 | alogliptin 25 MG / pioglitazone 30 MG Oral Tablet |
| RxNorm | 1368433 | alogliptin 25 MG / pioglitazone 30 MG [Oseni] |
| RxNorm | 1368434 | alogliptin / pioglitazone Oral Tablet [Oseni] |
| RxNorm | 1368435 | Oseni Oral Product |
| RxNorm | 1368436 | Oseni Pill |
| RxNorm | 1368437 | alogliptin 25 MG / pioglitazone 30 MG Oral Tablet [Oseni] |
| RxNorm | 1368438 | alogliptin 25 MG / pioglitazone 45 MG Oral Tablet |
| RxNorm | 1368440 | alogliptin 25 MG / pioglitazone 45 MG [Oseni] |
| RxNorm | 1368444 | alogliptin 25 MG / pioglitazone 45 MG Oral Tablet [Oseni] |
| RxNorm | 1372692 | Kazano |
| RxNorm | 1372706 | Jentadueto |
| RxNorm | 1372717 | Oseni |

Continued on next page

**Table9 – continued from previous page**

| CodeSystem | ConceptCode | ConceptName |
|---|---|---|
| RxNorm | 1372730 | Kombiglyze |
| RxNorm | 1372738 | Janumet |
| RxNorm | 1372754 | Juvisync |
| RxNorm | 1598392 | empagliflozin / linagliptin |
| RxNorm | 1602106 | empagliflozin / linagliptin Oral Product |
| RxNorm | 1602107 | empagliflozin / linagliptin Pill |
| RxNorm | 1602108 | empagliflozin / linagliptin Oral Tablet |
| RxNorm | 1602109 | empagliflozin 10 MG / linagliptin 5 MG Oral Tablet |
| RxNorm | 1602110 | Glyxambi |
| RxNorm | 1602111 | empagliflozin 10 MG / linagliptin 5 MG [Glyxambi] |
| RxNorm | 1602112 | empagliflozin / linagliptin Oral Tablet [Glyxambi] |
| RxNorm | 1602113 | Glyxambi Oral Product |
| RxNorm | 1602114 | Glyxambi Pill |
| RxNorm | 1602115 | empagliflozin 10 MG / linagliptin 5 MG Oral Tablet [Glyxambi] |
| RxNorm | 1602118 | empagliflozin 25 MG / linagliptin 5 MG Oral Tablet |
| RxNorm | 1602119 | empagliflozin 25 MG / linagliptin 5 MG [Glyxambi] |
| RxNorm | 1602120 | empagliflozin 25 MG / linagliptin 5 MG Oral Tablet [Glyxambi] |
| RxNorm | 1727500 | dapagliflozin / saxagliptin |
| RxNorm | 1796088 | linagliptin / metformin Extended Release Oral Tablet |
| RxNorm | 1796089 | 24 HR linagliptin 2.5 MG / metformin hydrochloride 1000 MG Extended Release Oral Tablet |
| RxNorm | 1796090 | linagliptin / metformin Extended Release Oral Tablet [Jentadueto] |
| RxNorm | 1796091 | 24 HR linagliptin 2.5 MG / metformin hydrochloride 1000 MG Extended Release Oral Tablet [Jentadueto] |
| RxNorm | 1796092 | linagliptin 2.5 MG / metformin hydrochloride 1000 MG Extended Release Oral Tablet |
| RxNorm | 1796093 | linagliptin 2.5 MG / metformin hydrochloride 1000 MG Extended Release Oral Tablet [Jentadueto] |
| RxNorm | 1796094 | 24 HR linagliptin 5 MG / metformin hydrochloride 1000 MG Extended Release Oral Tablet |
| RxNorm | 1796095 | linagliptin 5 MG / metformin hydrochloride 1000 MG [Jentadueto] |
| RxNorm | 1796096 | 24 HR linagliptin 5 MG / metformin hydrochloride 1000 MG Extended Release Oral Tablet [Jentadueto] |
| RxNorm | 1796097 | linagliptin 5 MG / metformin hydrochloride 1000 MG Extended Release Oral Tablet |
| RxNorm | 1796098 | linagliptin 5 MG / metformin hydrochloride 1000 MG Extended Release Oral Tablet [Jentadueto] |
| RxNorm | 1925495 | dapagliflozin / saxagliptin Oral Product |
| RxNorm | 1925496 | dapagliflozin / saxagliptin Pill |

Continued on next page

**Table9 – continued from previous page**

| CodeSystem | ConceptCode | ConceptName |
|---|---|---|
| RxNorm | 1925497 | dapagliflozin / saxagliptin Oral Tablet |
| RxNorm | 1925498 | dapagliflozin 10 MG / saxagliptin 5 MG Oral Tablet |
| RxNorm | 1925499 | Qtern |
| RxNorm | 1925500 | dapagliflozin 10 MG / saxagliptin 5 MG [Qtern] |
| RxNorm | 1925501 | dapagliflozin / saxagliptin Oral Tablet [Qtern] |
| RxNorm | 1925502 | Qtern Oral Product |
| RxNorm | 1925503 | Qtern Pill |
| RxNorm | 1925504 | dapagliflozin 10 MG / saxagliptin 5 MG Oral Tablet [Qtern] |
| RxNorm | 1992822 | ertugliflozin / sitagliptin Oral Product |
| RxNorm | 1992823 | ertugliflozin / sitagliptin Pill |
| RxNorm | 1992824 | ertugliflozin / sitagliptin Oral Tablet |
| RxNorm | 1992825 | ertugliflozin / sitagliptin |
| RxNorm | 1992826 | ertugliflozin 15 MG / sitagliptin 100 MG Oral Tablet |
| RxNorm | 1992827 | Steglujan |
| RxNorm | 1992828 | ertugliflozin 15 MG / sitagliptin 100 MG [Steglujan] |
| RxNorm | 1992829 | ertugliflozin / sitagliptin Oral Tablet [Steglujan] |
| RxNorm | 1992830 | Steglujan Oral Product |
| RxNorm | 1992831 | Steglujan Pill |
| RxNorm | 1992832 | ertugliflozin 15 MG / sitagliptin 100 MG Oral Tablet [Steglujan] |
| RxNorm | 1992835 | ertugliflozin 5 MG / sitagliptin 100 MG Oral Tablet |
| RxNorm | 1992836 | ertugliflozin 5 MG / sitagliptin 100 MG [Steglujan] |
| RxNorm | 1992837 | ertugliflozin 5 MG / sitagliptin 100 MG Oral Tablet [Steglujan] |
| RxNorm | 2117292 | dapagliflozin / metformin / saxagliptin |
| RxNorm | 2169274 | dapagliflozin 5 MG / saxagliptin 5 MG Oral Tablet |
| RxNorm | 2169275 | dapagliflozin 5 MG / saxagliptin 5 MG [Qtern] |
| RxNorm | 2169276 | dapagliflozin 5 MG / saxagliptin 5 MG Oral Tablet [Qtern] |
| RxNorm | 2281864 | empagliflozin / linagliptin / metformin |
| RxNorm | 2371722 | dapagliflozin / metformin / saxagliptin Oral Product |
| RxNorm | 2371723 | dapagliflozin / metformin / saxagliptin Pill |
| RxNorm | 2371724 | dapagliflozin / metformin / saxagliptin Extended Release Oral Tablet |
| RxNorm | 2371725 | 24 HR dapagliflozin 10 MG / metformin hydrochloride 1000 MG / saxagliptin 5 MG Extended Release Oral Tablet |
| RxNorm | 2371726 | Qternmet |
| RxNorm | 2371727 | dapagliflozin 10 MG / metformin hydrochloride 1000 MG / saxagliptin 5 MG [Qternmet] |
| RxNorm | 2371728 | dapagliflozin / metformin / saxagliptin Extended Release Oral Tablet [Qternmet] |
| RxNorm | 2371729 | Qternmet Oral Product |
| RxNorm | 2371730 | Qternmet Pill |

Continued on next page

**Table9 – continued from previous page**

| CodeSystem | ConceptCode | ConceptName |
|---|---|---|
| RxNorm | 2371731 | 24 HR dapagliflozin 10 MG / metformin hydrochloride 1000 MG / saxagliptin 5 MG Extended Release Oral Tablet [Qternmet] |
| RxNorm | 2371732 | dapagliflozin 10 MG / metformin hydrochloride 1000 MG / saxagliptin 5 MG Extended Release Oral Tablet |
| RxNorm | 2371733 | dapagliflozin 10 MG / metformin hydrochloride 1000 MG / saxagliptin 5 MG Extended Release Oral Tablet [Qternmet] |
| RxNorm | 2371734 | 24 HR dapagliflozin 2.5 MG / metformin hydrochloride 1000 MG / saxagliptin 2.5 MG Extended Release Oral Tablet |
| RxNorm | 2371735 | dapagliflozin 2.5 MG / metformin hydrochloride 1000 MG / saxagliptin 2.5 MG [Qternmet] |
| RxNorm | 2371736 | 24 HR dapagliflozin 2.5 MG / metformin hydrochloride 1000 MG / saxagliptin 2.5 MG Extended Release Oral Tablet [Qternmet] |
| RxNorm | 2371737 | dapagliflozin 2.5 MG / metformin hydrochloride 1000 MG / saxagliptin 2.5 MG Extended Release Oral Tablet |
| RxNorm | 2371738 | dapagliflozin 2.5 MG / metformin hydrochloride 1000 MG / saxagliptin 2.5 MG Extended Release Oral Tablet [Qternmet] |
| RxNorm | 2371740 | 24 HR dapagliflozin 5 MG / metformin hydrochloride 1000 MG / saxagliptin 2.5 MG Extended Release Oral Tablet |
| RxNorm | 2371741 | dapagliflozin 5 MG / metformin hydrochloride 1000 MG / saxagliptin 2.5 MG [Qternmet] |
| RxNorm | 2371742 | 24 HR dapagliflozin 5 MG / metformin hydrochloride 1000 MG / saxagliptin 2.5 MG Extended Release Oral Tablet [Qternmet] |
| RxNorm | 2371743 | dapagliflozin 5 MG / metformin hydrochloride 1000 MG / saxagliptin 2.5 MG Extended Release Oral Tablet |
| RxNorm | 2371744 | dapagliflozin 5 MG / metformin hydrochloride 1000 MG / saxagliptin 2.5 MG Extended Release Oral Tablet [Qternmet] |
| RxNorm | 2371745 | 24 HR dapagliflozin 5 MG / metformin hydrochloride 1000 MG / saxagliptin 5 MG Extended Release Oral Tablet |
| RxNorm | 2371746 | dapagliflozin 5 MG / metformin hydrochloride 1000 MG / saxagliptin 5 MG [Qternmet] |
| RxNorm | 2371747 | 24 HR dapagliflozin 5 MG / metformin hydrochloride 1000 MG / saxagliptin 5 MG Extended Release Oral Tablet [Qternmet] |
| RxNorm | 2371748 | dapagliflozin 5 MG / metformin hydrochloride 1000 MG / saxagliptin 5 MG Extended Release Oral Tablet |
| RxNorm | 2371749 | dapagliflozin 5 MG / metformin hydrochloride 1000 MG / saxagliptin 5 MG Extended Release Oral Tablet [Qternmet] |
| RxNorm | 2359276 | empagliflozin / linagliptin / metformin Oral Product |

Continued on next page

**Table9 – continued from previous page**

| CodeSystem | ConceptCode | ConceptName |
|---|---|---|
| RxNorm | 2359277 | empagliflozin / linagliptin / metformin Pill |
| RxNorm | 2359278 | empagliflozin / linagliptin / metformin Extended Release Oral Tablet |
| RxNorm | 2359279 | 24 HR empagliflozin 10 MG / linagliptin 5 MG / metformin hydrochloride 1000 MG Extended Release Oral Tablet |
| RxNorm | 2359280 | Trijardy |
| RxNorm | 2359281 | empagliflozin 10 MG / linagliptin 5 MG / metformin hydrochloride 1000 MG [Trijardy] |
| RxNorm | 2359282 | empagliflozin / linagliptin / metformin Extended Release Oral Tablet [Trijardy] |
| RxNorm | 2359283 | Trijardy Oral Product |
| RxNorm | 2359284 | Trijardy Pill |
| RxNorm | 2359285 | 24 HR empagliflozin 10 MG / linagliptin 5 MG / metformin hydrochloride 1000 MG Extended Release Oral Tablet [Trijardy] |
| RxNorm | 2359286 | empagliflozin 10 MG / linagliptin 5 MG / metformin hydrochloride 1000 MG Extended Release Oral Tablet |
| RxNorm | 2359287 | empagliflozin 10 MG / linagliptin 5 MG / metformin hydrochloride 1000 MG Extended Release Oral Tablet [Trijardy] |
| RxNorm | 2359288 | 24 HR empagliflozin 12.5 MG / linagliptin 2.5 MG / metformin hydrochloride 1000 MG Extended Release Oral Tablet |
| RxNorm | 2359289 | empagliflozin 12.5 MG / linagliptin 2.5 MG / metformin hydrochloride 1000 MG [Trijardy] |
| RxNorm | 2359290 | 24 HR empagliflozin 12.5 MG / linagliptin 2.5 MG / metformin hydrochloride 1000 MG Extended Release Oral Tablet [Trijardy] |
| RxNorm | 2359291 | empagliflozin 12.5 MG / linagliptin 2.5 MG / metformin hydrochloride 1000 MG Extended Release Oral Tablet |
| RxNorm | 2359292 | empagliflozin 12.5 MG / linagliptin 2.5 MG / metformin hydrochloride 1000 MG Extended Release Oral Tablet [Trijardy] |
| RxNorm | 2359351 | 24 HR empagliflozin 25 MG / linagliptin 5 MG / metformin hydrochloride 1000 MG Extended Release Oral Tablet |
| RxNorm | 2359352 | empagliflozin 25 MG / linagliptin 5 MG / metformin hydrochloride 1000 MG [Trijardy] |
| RxNorm | 2359353 | 24 HR empagliflozin 25 MG / linagliptin 5 MG / metformin hydrochloride 1000 MG Extended Release Oral Tablet [Trijardy] |
| RxNorm | 2359354 | empagliflozin 25 MG / linagliptin 5 MG / metformin hydrochloride 1000 MG Extended Release Oral Tablet |

Continued on next page

**Table9 – continued from previous page**

| CodeSystem | ConceptCode | ConceptName |
|---|---|---|
| RxNorm | 2359355 | empagliflozin 25 MG / linagliptin 5 MG / metformin hydrochloride 1000 MG Extended Release Oral Tablet [Trijardy] |
| RxNorm | 2359356 | 24 HR empagliflozin 5 MG / linagliptin 2.5 MG / metformin hydrochloride 1000 MG Extended Release Oral Tablet |
| RxNorm | 2359357 | empagliflozin 5 MG / linagliptin 2.5 MG / metformin hydrochloride 1000 MG [Trijardy] |
| RxNorm | 2359358 | 24 HR empagliflozin 5 MG / linagliptin 2.5 MG / metformin hydrochloride 1000 MG Extended Release Oral Tablet [Trijardy] |
| RxNorm | 2359359 | empagliflozin 5 MG / linagliptin 2.5 MG / metformin hydrochloride 1000 MG Extended Release Oral Tablet |
| RxNorm | 2359360 | empagliflozin 5 MG / linagliptin 2.5 MG / metformin hydrochloride 1000 MG Extended Release Oral Tablet [Trijardy] |
| RxNorm | 705136 | Metformin 1000 MG / sitagliptin 50 MG Oral Tablet [Janumet] |
| RxNorm | 705134 | Metformin 1000 MG / sitagliptin 50 MG [Janumet] |
| RxNorm | 705135 | Metformin / sitagliptin Oral Tablet [Janumet] |
| RxNorm | 705138 | Metformin 500 MG / sitagliptin 50 MG Oral Tablet [Janumet] |
| RxNorm | 748763 | Metformin 500 MG / sitagliptin 64.3 MG Oral Tablet |
| RxNorm | 757607 | Metformin / sitagliptin Oral Tablet [Janumet 50 mg/500 mg] |
| RxNorm | 748762 | sitagliptin 64.3 MG |
| RxNorm | 748765 | Metformin 500 MG / sitagliptin 64.3 MG Oral Tablet [Janumet] |
| RxNorm | 705137 | Metformin 500 MG / sitagliptin 50 MG [Janumet] |
| RxNorm | 748764 | Metformin 500 MG / sitagliptin 64.3 MG [Janumet] |
| RxNorm | 1043581 | Metformin / saxagliptin Extended Release Tablet [Kombiglyze 5/500] |
| RxNorm | 1043573 | Metformin / saxagliptin Extended Release Tablet [Kombiglyze 5/1000] |
| RxNorm | 1189807 | Simvastatin / sitagliptin Oral Tablet [Juvisync 100/10] |
| RxNorm | 1189824 | Simvastatin / sitagliptin Oral Tablet [Juvisync 100/40] |
| RxNorm | 1243031 | Jentadueto 2.5/500 Oral Product |
| RxNorm | 1243025 | Jentadueto 2.5/1000 Pill |
| RxNorm | 1173547 | Kombiglyze 5/500 Oral Product |
| RxNorm | 1189825 | Juvisync 100/40 Oral Product |
| RxNorm | 1172859 | Kombiglyze 2.5/1000 Oral Product |
| RxNorm | 1173546 | Kombiglyze 5/1000 Pill |
| RxNorm | 1243832 | Janumet 100/1000 Pill |
| RxNorm | 1189816 | Juvisync 100/10 Oral Product |

Continued on next page

**Table9 – continued from previous page**

| CodeSystem | ConceptCode | ConceptName |
|---|---|---|
| RxNorm | 1189826 | Juvisync 100/40 Pill |
| RxNorm | 1189817 | Juvisync 100/10 Pill |
| RxNorm | 1167812 | Janumet 50/500 Oral Product |
| RxNorm | 1243032 | Jentadueto 2.5/500 Pill |
| RxNorm | 1173548 | Kombiglyze 5/500 Pill |
| RxNorm | 1167813 | Janumet 50/500 Pill |
| RxNorm | 1243024 | Jentadueto 2.5/1000 Oral Product |
| RxNorm | 1243831 | Janumet 100/1000 Oral Product |
| RxNorm | 700518 | Metformin 1000 MG / sitagliptin 50 MG Oral Tablet |
| RxNorm | 700517 | Metformin 500 MG / sitagliptin 50 MG Oral Tablet |
| RxNorm | 757606 | Metformin 500 MG / sitagliptin 50 MG [Janumet 50 mg/500 mg] |
| RxNorm | 757602 | Metformin 1000 MG / sitagliptin 50 MG [Janumet 50 mg/1000 mg] |
| RxNorm | 757608 | Metformin 500 MG / sitagliptin 50 MG Oral Tablet [Janumet 50 mg/500 mg] |
| RxNorm | 757604 | Metformin 1000 MG / sitagliptin 50 MG Oral Tablet [Janumet 50 mg/1000 mg] |
| RxNorm | 1243830 | Metformin / sitagliptin Extended Release Tablet [Janumet 100/1000] |
| RxNorm | 1243847 | Metformin / sitagliptin Extended Release Tablet [Janumet 50/500] |
| RxNorm | 1243023 | Linagliptin / Metformin Oral Tablet [Jentadueto 2.5/1000] |
| RxNorm | 1243030 | Linagliptin / Metformin Oral Tablet [Jentadueto 2.5/500] |
| RxNorm | 1043561 | metformin / saxagliptin Extended Release Oral Tablet |
| RxNorm | 1043562 | metformin / saxagliptin |
| RxNorm | 1043563 | 24 HR metformin hydrochloride 1000 MG / saxagliptin 2.5 MG Extended Release Oral Tablet |
| RxNorm | 1043565 | metformin hydrochloride 1000 MG / saxagliptin 2.5 MG [Kombiglyze] |
| RxNorm | 1043566 | metformin / saxagliptin Extended Release Oral Tablet [Kombiglyze] |
| RxNorm | 1043567 | 24 HR metformin hydrochloride 1000 MG / saxagliptin 2.5 MG Extended Release Oral Tablet [Kombiglyze] |
| RxNorm | 1043568 | metformin hydrochloride 1000 MG / saxagliptin 2.5 MG Extended Release Oral Tablet |
| RxNorm | 1043569 | metformin hydrochloride 1000 MG / saxagliptin 2.5 MG Extended Release Oral Tablet [Kombiglyze] |
| RxNorm | 1043570 | 24 HR metformin hydrochloride 1000 MG / saxagliptin 5 MG Extended Release Oral Tablet |
| RxNorm | 1043572 | metformin hydrochloride 1000 MG / saxagliptin 5 MG [Kombiglyze] |

Continued on next page

**Table9 – continued from previous page**

| CodeSystem | ConceptCode | ConceptName |
|---|---|---|
| RxNorm | 1043574 | 24 HR metformin hydrochloride 1000 MG / saxagliptin 5 MG Extended Release Oral Tablet [Kombiglyze] |
| RxNorm | 1043575 | metformin hydrochloride 1000 MG / saxagliptin 5 MG Extended Release Oral Tablet |
| RxNorm | 1043576 | metformin hydrochloride 1000 MG / saxagliptin 5 MG Extended Release Oral Tablet [Kombiglyze] |
| RxNorm | 1043578 | 24 HR metformin hydrochloride 500 MG / saxagliptin 5 MG Extended Release Oral Tablet |
| RxNorm | 1043580 | metformin hydrochloride 500 MG / saxagliptin 5 MG [Kombiglyze] |
| RxNorm | 1043582 | 24 HR metformin hydrochloride 500 MG / saxagliptin 5 MG Extended Release Oral Tablet [Kombiglyze] |
| RxNorm | 1043583 | metformin hydrochloride 500 MG / saxagliptin 5 MG Extended Release Oral Tablet |
| RxNorm | 1043584 | metformin hydrochloride 500 MG / saxagliptin 5 MG Extended Release Oral Tablet [Kombiglyze] |
| RxNorm | 1100700 | linagliptin 5 MG |
| RxNorm | 1100701 | linagliptin Oral Tablet |
| RxNorm | 1100702 | linagliptin 5 MG Oral Tablet |
| RxNorm | 1100703 | Tradjenta |
| RxNorm | 1100704 | linagliptin 5 MG [Tradjenta] |
| RxNorm | 1100705 | linagliptin Oral Tablet [Tradjenta] |
| RxNorm | 1100706 | linagliptin 5 MG Oral Tablet [Tradjenta] |
| RxNorm | 1158518 | saxagliptin Oral Product |
| RxNorm | 1158519 | saxagliptin Pill |
| RxNorm | 1159662 | sitagliptin Oral Product |
| RxNorm | 1159663 | sitagliptin Pill |
| RxNorm | 1161605 | metformin / saxagliptin Oral Product |
| RxNorm | 1161606 | metformin / saxagliptin Pill |
| RxNorm | 1161607 | metformin / sitagliptin Oral Product |
| RxNorm | 1161608 | metformin / sitagliptin Pill |
| RxNorm | 1164670 | linagliptin Oral Product |
| RxNorm | 1164671 | linagliptin Pill |
| RxNorm | 1181729 | Onglyza Oral Product |
| RxNorm | 1181730 | Onglyza Pill |
| RxNorm | 665031 | sitagliptin 100 MG |
| RxNorm | 665032 | sitagliptin Oral Tablet |
| RxNorm | 665033 | sitagliptin 100 MG Oral Tablet |
| RxNorm | 665034 | sitagliptin 100 MG [Januvia] |
| RxNorm | 665035 | sitagliptin Oral Tablet [Januvia] |
| RxNorm | 665036 | sitagliptin 100 MG Oral Tablet [Januvia] |
| RxNorm | 665037 | sitagliptin 25 MG |
| RxNorm | 665038 | sitagliptin 25 MG Oral Tablet |

Continued on next page

**Table9 – continued from previous page**

| CodeSystem | ConceptCode | ConceptName |
|---|---|---|
| RxNorm | 665039 | sitagliptin 25 MG [Januvia] |
| RxNorm | 665040 | sitagliptin 25 MG Oral Tablet [Januvia] |
| RxNorm | 665041 | sitagliptin 50 MG |
| RxNorm | 665042 | sitagliptin 50 MG Oral Tablet |
| RxNorm | 665043 | sitagliptin 50 MG [Januvia] |
| RxNorm | 665044 | sitagliptin 50 MG Oral Tablet [Januvia] |
| RxNorm | 638596 | Januvia |
| RxNorm | 700516 | metformin / sitagliptin Oral Tablet |
| RxNorm | 729717 | metformin / sitagliptin |
| RxNorm | 757603 | metformin / sitagliptin Oral Tablet [Janumet] |
| RxNorm | 857974 | saxagliptin |
| RxNorm | 858034 | saxagliptin 5 MG |
| RxNorm | 858035 | saxagliptin Oral Tablet |
| RxNorm | 858036 | saxagliptin 5 MG Oral Tablet |
| RxNorm | 858037 | Onglyza |
| RxNorm | 858038 | saxagliptin 5 MG [Onglyza] |
| RxNorm | 858039 | saxagliptin Oral Tablet [Onglyza] |
| RxNorm | 858040 | saxagliptin 5 MG Oral Tablet [Onglyza] |
| RxNorm | 858041 | saxagliptin 2.5 MG |
| RxNorm | 858042 | saxagliptin 2.5 MG Oral Tablet |
| RxNorm | 858043 | saxagliptin 2.5 MG [Onglyza] |
| RxNorm | 858044 | saxagliptin 2.5 MG Oral Tablet [Onglyza] |
| RxNorm | 861769 | metformin hydrochloride 1000 MG / sitagliptin 50 MG Oral Tablet |
| RxNorm | 861770 | metformin hydrochloride 1000 MG / sitagliptin 50 MG [Janumet] |
| RxNorm | 861771 | metformin hydrochloride 1000 MG / sitagliptin 50 MG Oral Tablet [Janumet] |
| RxNorm | 861819 | metformin hydrochloride 500 MG / sitagliptin 50 MG Oral Tablet |
| RxNorm | 861820 | metformin hydrochloride 500 MG / sitagliptin 50 MG [Janumet] |
| RxNorm | 861821 | metformin hydrochloride 500 MG / sitagliptin 50 MG Oral Tablet [Janumet] |
| RxNorm | 253182 | insulin, regular, human |
| RxNorm | 1183566 | Pur-In Mix Injectable Product |
| RxNorm | 1183568 | Pur-In Neutral Injectable Product |
| RxNorm | 1178125 | Novolin Injectable Product |
| RxNorm | 1178128 | Novolin R Injectable Product |
| RxNorm | 1187762 | Velosulin Injectable Product |
| RxNorm | 1359720 | insulin isophane, human 70 UNT/ML / insulin, regular, human 30 UNT/ML Prefilled Syringe [Humulin] |

Continued on next page

**Table9 – continued from previous page**

| CodeSystem | ConceptCode | ConceptName |
|---|---|---|
| RxNorm | 1360172 | insulin isophane, human 70 UNT/ML / insulin, regular, human 30 UNT/ML Prefilled Syringe |
| RxNorm | 1360226 | insulin isophane, human 70 UNT/ML / insulin, regular, human 30 UNT/ML Prefilled Syringe [Novolin] |
| RxNorm | 1360482 | insulin, regular, human 100 UNT/ML Prefilled Syringe |
| RxNorm | 1360435 | insulin, regular, human 100 UNT/ML Prefilled Syringe [Novolin R] |
| RxNorm | 1372685 | Human Mixtard |
| RxNorm | 1372723 | Novolin |
| RxNorm | 1372744 | Humulin |
| RxNorm | 1372761 | Pur-In Mix |
| RxNorm | 150659 | insulin, regular, human 100 UNT/ML Injectable Solution [Pur-In Neutral] |
| RxNorm | 150831 | insulin, regular, human 100 UNT/ML Injectable Solution [Human Actrapid Penfill] |
| RxNorm | 150973 | insulin, regular, human 100 UNT/ML Injectable Solution [Human Actrapid] |
| RxNorm | 150974 | insulin, regular, human 100 UNT/ML Injectable Solution [Humulin S] |
| RxNorm | 1544488 | insulin, regular, human 8 UNT Inhalation Powder |
| RxNorm | 1544490 | insulin, regular, human 8 UNT Inhalation Powder [Afrezza] |
| RxNorm | 1544568 | {60 (insulin, regular, human 4 UNT Inhalation Powder) / 30 (insulin, regular, human 8 UNT Inhalation Powder) } Pack |
| RxNorm | 1544569 | {60 (insulin, regular, human 4 UNT Inhalation Powder [Afrezza]) / 30 (insulin, regular, human 8 UNT Inhalation Powder [Afrezza]) } Pack [Afrezza 90 - 60 (4 UNT), 30 (8 UNT)] |
| RxNorm | 1544570 | {30 (insulin, regular, human 4 UNT Inhalation Powder) / 60 (insulin, regular, human 8 UNT Inhalation Powder) } Pack |
| RxNorm | 1544571 | {30 (insulin, regular, human 4 UNT Inhalation Powder [Afrezza]) / 60 (insulin, regular, human 8 UNT Inhalation Powder [Afrezza]) } Pack [Afrezza 90 - 30 (4 UNT), 60 (8 UNT)] |
| RxNorm | 1543200 | insulin, regular, human Inhalant Product |
| RxNorm | 1543202 | insulin, regular, human 4 UNT Inhalation Powder |
| RxNorm | 1543203 | Afrezza |
| RxNorm | 1543206 | Afrezza Inhalant Product |
| RxNorm | 1543207 | insulin, regular, human 4 UNT Inhalation Powder [Afrezza] |
| RxNorm | 152644 | Human Actrapid |
| RxNorm | 1650256 | insulin, regular, human 4 UNT |
| RxNorm | 1650260 | insulin, regular, human 4 UNT [Afrezza] |

Continued on next page

**Table9 – continued from previous page**

| CodeSystem | ConceptCode | ConceptName |
|---|---|---|
| RxNorm | 1650262 | insulin, regular, human 8 UNT |
| RxNorm | 1650264 | insulin, regular, human 8 UNT [Afrezza] |
| RxNorm | 1653899 | insulin isophane / insulin, regular, human Injectable Product |
| RxNorm | 1654651 | insulin isophane / insulin, regular, human Pen Injector |
| RxNorm | 1654855 | insulin isophane / insulin, regular, human Pen Injector [Humulin] |
| RxNorm | 1654857 | insulin isophane, human 70 UNT/ML / insulin, regular, human 30 UNT/ML Pen Injector |
| RxNorm | 1654858 | insulin isophane, human 70 UNT/ML / insulin, regular, human 30 UNT/ML Pen Injector [Humulin] |
| RxNorm | 1654909 | insulin, regular, human 12 UNT |
| RxNorm | 1654910 | insulin, regular, human 12 UNT Inhalation Powder |
| RxNorm | 1654911 | insulin, regular, human 12 UNT [Afrezza] |
| RxNorm | 1654912 | insulin, regular, human 12 UNT Inhalation Powder [Afrezza] |
| RxNorm | 1656705 | {30 (insulin, regular, human 12 UNT Inhalation Powder) / 60 (insulin, regular, human 8 UNT Inhalation Powder) } Pack |
| RxNorm | 1656706 | {30 (insulin, regular, human 12 UNT Inhalation Powder [Afrezza]) / 60 (insulin, regular, human 8 UNT Inhalation Powder [Afrezza]) } Pack [Afrezza 90 - 60 (8 UNT), 30 (12 UNT)] |
| RxNorm | 1731314 | insulin, regular, human Pen Injector |
| RxNorm | 1731315 | 3 ML insulin, regular, human 500 UNT/ML Pen Injector |
| RxNorm | 1731316 | insulin, regular, human Pen Injector [Humulin R] |
| RxNorm | 1731317 | 3 ML insulin, regular, human 500 UNT/ML Pen Injector [Humulin R] |
| RxNorm | 1731318 | insulin, regular, human 500 UNT/ML Pen Injector |
| RxNorm | 1731319 | insulin, regular, human 500 UNT/ML Pen Injector [Humulin R] |
| RxNorm | 1798387 | {90 (insulin, regular, human 4 UNT Inhalation Powder) / 90 (insulin, regular, human 8 UNT Inhalation Powder) } Pack |
| RxNorm | 1798388 | {90 (insulin, regular, human 4 UNT Inhalation Powder [Afrezza]) / 90 (insulin, regular, human 8 UNT Inhalation Powder [Afrezza]) } Pack [Afrezza Titration Pack] |
| RxNorm | 1862101 | {60 (insulin, regular, human 12 UNT Inhalation Powder) / 60 (insulin, regular, human 4 UNT Inhalation Powder) / 60 (insulin, regular, human 8 UNT Inhalation Powder) } Pack |

Continued on next page

**Table9 – continued from previous page**

| CodeSystem | ConceptCode | ConceptName |
|---|---|---|
| RxNorm | 1862102 | {60 (insulin, regular, human 12 UNT Inhalation Powder [Afrezza]) / 60 (insulin, regular, human 4 UNT Inhalation Powder [Afrezza]) / 60 (insulin, regular, human 8 UNT Inhalation Powder [Afrezza]) } Pack [Afrezza 180 Titration Pack- 60 (4 UNT), 60 (8 UNT), 60 (12 UNT)] |
| RxNorm | 203209 | Human Actrapid Penfill |
| RxNorm | 2049379 | insulin isophane / insulin, regular, human Pen Injector [Novolin] |
| RxNorm | 2049380 | 3 ML insulin isophane, human 70 UNT/ML / insulin, regular, human 30 UNT/ML Pen Injector [Novolin] |
| RxNorm | 2049381 | insulin isophane, human 70 UNT/ML / insulin, regular, human 30 UNT/ML Pen Injector [Novolin] |
| RxNorm | 2100028 | {90 (insulin, regular, human 12 UNT Inhalation Powder) / 90 (insulin, regular, human 8 UNT Inhalation Powder) } Pack |
| RxNorm | 2100029 | {90 (insulin, regular, human 12 UNT Inhalation Powder [Afrezza]) / 90 (insulin, regular, human 8 UNT Inhalation Powder [Afrezza]) } Pack [Afrezza 180 - 90 (8 UNT), 90 (12 UNT)] |
| RxNorm | 2108525 | insulin, regular, human Inhalation Powder |
| RxNorm | 2108527 | insulin, regular, human Inhalation Powder [Afrezza] |
| RxNorm | 213442 | insulin isophane, human 70 UNT/ML / insulin, regular, human 30 UNT/ML Injectable Suspension [Novolin] |
| RxNorm | 2179742 | insulin, regular, human 1 UNT/ML |
| RxNorm | 2179743 | insulin, regular, human Injection |
| RxNorm | 2179744 | 100 ML insulin, regular, human 1 UNT/ML Injection |
| RxNorm | 2179745 | Myxredlin |
| RxNorm | 2179746 | insulin, regular, human 1 UNT/ML [Myxredlin] |
| RxNorm | 2179747 | insulin, regular, human Injection [Myxredlin] |
| RxNorm | 2179748 | Myxredlin Injectable Product |
| RxNorm | 2179749 | 100 ML insulin, regular, human 1 UNT/ML Injection [Myxredlin] |
| RxNorm | 2179750 | insulin, regular, human 1 UNT/ML Injection |
| RxNorm | 2179751 | insulin, regular, human 1 UNT/ML Injection [Myxredlin] |
| RxNorm | 2206090 | 3 ML insulin, regular, human 100 UNT/ML Pen Injector |
| RxNorm | 2206091 | insulin, regular, human Pen Injector [Novolin R] |
| RxNorm | 2206092 | 3 ML insulin, regular, human 100 UNT/ML Pen Injector [Novolin R] |
| RxNorm | 2206093 | insulin, regular, human 100 UNT/ML Pen Injector |
| RxNorm | 2206094 | insulin, regular, human 100 UNT/ML Pen Injector [Novolin R] |
| RxNorm | 247511 | insulin isophane, human 90 UNT/ML / insulin, regular, human 10 UNT/ML Injectable Suspension |

Continued on next page

**Table9 – continued from previous page**

| CodeSystem | ConceptCode | ConceptName |
|---|---|---|
| RxNorm | 247512 | insulin isophane, human 80 UNT/ML / insulin, regular, human 20 UNT/ML Injectable Suspension |
| RxNorm | 247513 | insulin isophane, human 60 UNT/ML / insulin, regular, human 40 UNT/ML Injectable Suspension |
| RxNorm | 249026 | insulin, regular, human 40 UNT/ML Injectable Solution |
| RxNorm | 245264 | insulin isophane, human 85 UNT/ML / insulin, regular, human 15 UNT/ML Injectable Suspension |
| RxNorm | 245265 | insulin isophane, human 50 UNT/ML / insulin, regular, human 50 UNT/ML Injectable Suspension |
| RxNorm | 249220 | insulin, regular, human 500 UNT/ML Injectable Solution |
| RxNorm | 379742 | Insulin, Isophane, Human / Insulin, Regular, Human Injectable Suspension [Human Mixtard 30 ge] |
| RxNorm | 365582 | Insulin, Isophane, Human / Insulin, Regular, Human Injectable Suspension [Humulin M3] |
| RxNorm | 365576 | Insulin, Isophane, Human / Insulin, Regular, Human Injectable Suspension [Humulin M4] |
| RxNorm | 365684 | Insulin, Isophane, Human / Insulin, Regular, Human Injectable Suspension [Pur-In Mix 25/75] |
| RxNorm | 575139 | Insulin, Isophane, Human 70 UNT/ML / Insulin, Regular, Human 30 UNT/ML [Novolin 70/30 PenFill] |
| RxNorm | 724341 | Insulin, Isophane, Human / Insulin, Regular, Human Injectable Suspension [ReliOn/Novolin 70/30] |
| RxNorm | 727909 | 1.5 ML Insulin, Isophane, Human 70 UNT/ML / Insulin, Regular, Human 30 UNT/ML Prefilled Syringe |
| RxNorm | 727917 | 3 ML Insulin, Regular, Human 100 UNT/ML Prefilled Syringe |
| RxNorm | 727770 | 3 ML Insulin, Isophane, Human 50 UNT/ML / Insulin, Regular, Human 50 UNT/ML Prefilled Syringe |
| RxNorm | 727766 | 3 ML Insulin, Isophane, Human 85 UNT/ML / Insulin, Regular, Human 15 UNT/ML Prefilled Syringe |
| RxNorm | 727626 | Insulin, Isophane, Human / Insulin, Regular, Human Prefilled Syringe |
| RxNorm | 723552 | Insulin, Regular, Human Injectable Solution [ReliOn R/Novolin] |
| RxNorm | 727895 | Insulin, Isophane, Human / Insulin, Regular, Human Prefilled Syringe [Humulin 70/30] |
| RxNorm | 727916 | Insulin, Regular, Human Prefilled Syringe |
| RxNorm | 727769 | 3 ML Insulin, Isophane, Human 60 UNT/ML / Insulin, Regular, Human 40 UNT/ML Prefilled Syringe |
| RxNorm | 727918 | 1.5 ML Insulin, Regular, Human 100 UNT/ML Prefilled Syringe |
| RxNorm | 727627 | 3 ML Insulin, Isophane, Human 70 UNT/ML / Insulin, Regular, Human 30 UNT/ML Prefilled Syringe |

Continued on next page

**Table9 – continued from previous page**

| CodeSystem | ConceptCode | ConceptName |
|---|---|---|
| RxNorm | 727755 | 3 ML Insulin, Isophane, Human 90 UNT/ML / Insulin, Regular, Human 10 UNT/ML Prefilled Syringe |
| RxNorm | 727628 | Insulin, Isophane, Human / Insulin, Regular, Human Prefilled Syringe [Novolin 70/30] |
| RxNorm | 727896 | 3 ML Insulin, Isophane, Human 70 UNT/ML / Insulin, Regular, Human 30 UNT/ML Prefilled Syringe [Humulin 70/30] |
| RxNorm | 724342 | Insulin, Isophane, Human 70 UNT/ML / Insulin, Regular, Human 30 UNT/ML Injectable Suspension [ReliOn/Novolin 70/30] |
| RxNorm | 727629 | 3 ML Insulin, Isophane, Human 70 UNT/ML / Insulin, Regular, Human 30 UNT/ML Prefilled Syringe [Novolin 70/30] |
| RxNorm | 727768 | 3 ML Insulin, Isophane, Human 75 UNT/ML / Insulin, Regular, Human 25 UNT/ML Prefilled Syringe |
| RxNorm | 723553 | Insulin, Regular, Human 100 UNT/ML Injectable Solution [ReliOn R/Novolin] |
| RxNorm | 727910 | 1.5 ML Insulin, Isophane, Human 70 UNT/ML / Insulin, Regular, Human 30 UNT/ML Prefilled Syringe [Novolin 70/30] |
| RxNorm | 723551 | Insulin, Regular, Human 100 UNT/ML [ReliOn R/Novolin] |
| RxNorm | 727767 | 3 ML Insulin, Isophane, Human 80 UNT/ML / Insulin, Regular, Human 20 UNT/ML Prefilled Syringe |
| RxNorm | 724340 | Insulin, Isophane, Human 70 UNT/ML / Insulin, Regular, Human 30 UNT/ML [ReliOn/Novolin 70/30] |
| RxNorm | 1172687 | Humulin M2 Injectable Product |
| RxNorm | 1172688 | Humulin M3 Injectable Product |
| RxNorm | 1156396 | NPH Insulin, Human / Regular Insulin, Human Injectable Product |
| RxNorm | 1172689 | Humulin M4 Injectable Product |
| RxNorm | 1171298 | Human Mixtard 50 Injectable Product |
| RxNorm | 1172690 | Humulin M5 Injectable Product |
| RxNorm | 1183567 | Pur-In Mix 50/50 Injectable Product |
| RxNorm | 1183565 | Pur-In Mix 15/85 Injectable Product |
| RxNorm | 1172683 | Humulin 70/30 Injectable Product |
| RxNorm | 1172686 | Humulin M1 Injectable Product |
| RxNorm | 1171299 | Human Mixtard Penfill Injectable Product |
| RxNorm | 1171297 | Human Mixtard 30 ge Injectable Product |
| RxNorm | 575133 | Insulin, Isophane, Human 50 UNT/ML / Insulin, Regular, Human 50 UNT/ML [Humulin 50/50] |
| RxNorm | 106893 | Insulin, Isophane, Human 70 UNT/ML / Insulin, Regular, Human 30 UNT/ML Injectable Suspension [Human Mixtard 30] |

Continued on next page

**Table9 – continued from previous page**

| CodeSystem | ConceptCode | ConceptName |
|---|---|---|
| RxNorm | 108812 | Insulin, Isophane, Human 70 UNT/ML / Insulin, Regular, Human 30 UNT/ML Injectable Suspension [Human Mixtard 30 ge] |
| RxNorm | 847195 | 3 ML insulin human, isophane 70 UNT/ML / Regular Insulin, Human 30 UNT/ML Prefilled Syringe [Novolin] |
| RxNorm | 213441 | Insulin, Isophane, Human 70 UNT/ML / Insulin, Regular, Human 30 UNT/ML Injectable Suspension [Humulin 70/30] |
| RxNorm | 153122 | Insulin, Isophane, Human 50 UNT/ML / Insulin, Regular, Human 50 UNT/ML Injectable Suspension [Human Mixtard 50] |
| RxNorm | 311016 | Insulin, Isophane, Human 50 UNT/ML / Insulin, Regular, Human 50 UNT/ML Injectable Suspension [Humulin 50/50] |
| RxNorm | 564604 | Insulin, Isophane, Human 50 UNT/ML / Insulin, Regular, Human 50 UNT/ML [Human Mixtard Penfill] |
| RxNorm | 564394 | Insulin, Isophane, Human 70 UNT/ML / Insulin, Regular, Human 30 UNT/ML [Human Mixtard 30] |
| RxNorm | 564600 | Insulin, Isophane, Human 70 UNT/ML / Insulin, Regular, Human 30 UNT/ML [Human Mixtard 30 ge] |
| RxNorm | 564393 | Insulin, Isophane, Human 70 UNT/ML / Insulin, Regular, Human 30 UNT/ML [Humulin M3] |
| RxNorm | 1544487 | Regular Insulin, Human 0.7 MG/ACTUAT |
| RxNorm | 1544489 | Regular Insulin, Human 0.7 MG/ACTUAT [Afrezza] |
| RxNorm | 1543204 | Regular Insulin, Human 0.35 MG/ACTUAT [Afrezza] |
| RxNorm | 1543201 | Regular Insulin, Human Inhalant Powder |
| RxNorm | 1543205 | Regular Insulin, Human Inhalant Powder [Afrezza] |
| RxNorm | 1543199 | Regular Insulin, Human 0.35 MG/ACTUAT |
| RxNorm | 380934 | Insulin, Isophane, Human / Insulin, Regular, Human Injectable Suspension [Human Mixtard Penfill] |
| RxNorm | 379754 | Insulin, Isophane, Human / Insulin, Regular, Human Injectable Suspension [Novolin 70/30 PenFill] |
| RxNorm | 365578 | Insulin, Isophane, Human / Insulin, Regular, Human Injectable Suspension [Humulin M1] |
| RxNorm | 365580 | Insulin, Isophane, Human / Insulin, Regular, Human Injectable Suspension [Pur-In Mix 50/50] |
| RxNorm | 365577 | Insulin, Isophane, Human / Insulin, Regular, Human Injectable Suspension [Humulin M2] |
| RxNorm | 365575 | Insulin, Isophane, Human / Insulin, Regular, Human Injectable Suspension [Humulin M5] |
| RxNorm | 379741 | Insulin, Isophane, Human / Insulin, Regular, Human Injectable Suspension [Human Mixtard 50] |
| RxNorm | 365673 | Insulin, Isophane, Human / Insulin, Regular, Human Injectable Suspension [Humulin 70/30] |

Continued on next page

**Table9 – continued from previous page**

| CodeSystem | ConceptCode | ConceptName |
|---|---|---|
| RxNorm | 311023 | Insulin, Isophane, Human 70 UNT/ML / Insulin, Regular, Human 30 UNT/ML Injectable Suspension [Novolin 70/30 PenFill] |
| RxNorm | 1008501 | insulin isophane / insulin, regular, human |
| RxNorm | 108813 | insulin isophane, human 90 UNT/ML / insulin, regular, human 10 UNT/ML Injectable Suspension [Human Mixtard] |
| RxNorm | 108814 | insulin isophane, human 80 UNT/ML / insulin, regular, human 20 UNT/ML Injectable Suspension [Human Mixtard] |
| RxNorm | 108815 | insulin isophane, human 70 UNT/ML / insulin, regular, human 30 UNT/ML Injectable Suspension [Human Mixtard] |
| RxNorm | 108816 | insulin isophane, human 50 UNT/ML / insulin, regular, human 50 UNT/ML Injectable Suspension [Human Mixtard] |
| RxNorm | 108822 | insulin isophane, human 60 UNT/ML / insulin, regular, human 40 UNT/ML Injectable Suspension [Human Mixtard] |
| RxNorm | 11160 | Velosulin |
| RxNorm | 106892 | insulin isophane, human 70 UNT/ML / insulin, regular, human 30 UNT/ML Injectable Suspension [Humulin] |
| RxNorm | 106894 | insulin isophane, human 85 UNT/ML / insulin, regular, human 15 UNT/ML Injectable Suspension [Pur-In Mix] |
| RxNorm | 106895 | insulin isophane, human 75 UNT/ML / insulin, regular, human 25 UNT/ML Injectable Suspension [Pur-In Mix] |
| RxNorm | 106896 | insulin isophane, human 50 UNT/ML / insulin, regular, human 50 UNT/ML Injectable Suspension [Pur-In Mix] |
| RxNorm | 106899 | insulin isophane, human 90 UNT/ML / insulin, regular, human 10 UNT/ML Injectable Suspension [Humulin] |
| RxNorm | 106900 | insulin isophane, human 80 UNT/ML / insulin, regular, human 20 UNT/ML Injectable Suspension [Humulin] |
| RxNorm | 106901 | insulin isophane, human 60 UNT/ML / insulin, regular, human 40 UNT/ML Injectable Suspension [Humulin] |
| RxNorm | 108407 | insulin isophane, human 50 UNT/ML / insulin, regular, human 50 UNT/ML Injectable Suspension [Humulin] |
| RxNorm | 1164824 | insulin, regular, human Injectable Product |
| RxNorm | 1171292 | Human Actrapid Injectable Product |
| RxNorm | 1171293 | Human Actrapid Penfill Injectable Product |
| RxNorm | 1171296 | Human Mixtard Injectable Product |
| RxNorm | 1171997 | Humulin Injectable Product |
| RxNorm | 1172692 | Humulin R Injectable Product |
| RxNorm | 1172693 | Humulin S Injectable Product |
| RxNorm | 311033 | insulin, regular, human 100 UNT/ML Injectable Solution [Novolin R] |
| RxNorm | 311034 | insulin, regular, human 100 UNT/ML Injectable Solution |
| RxNorm | 311035 | insulin, regular, human 100 UNT/ML Injectable Solution [Velosulin] |

Continued on next page

**Table9 – continued from previous page**

| CodeSystem | ConceptCode | ConceptName |
|---|---|---|
| RxNorm | 311036 | insulin, regular, human 100 UNT/ML Injectable Solution [Humulin R] |
| RxNorm | 311048 | insulin isophane, human 70 UNT/ML / insulin, regular, human 30 UNT/ML Injectable Suspension |
| RxNorm | 340325 | insulin, regular, human 100 UNT/ML |
| RxNorm | 340326 | insulin, regular, human 40 UNT/ML |
| RxNorm | 340327 | insulin, regular, human 500 UNT/ML |
| RxNorm | 351859 | insulin, regular, human 500 UNT/ML Injectable Solution [Humulin R] |
| RxNorm | 362622 | insulin, regular, human Injectable Solution [Humulin R] |
| RxNorm | 363120 | insulin, regular, human Injectable Solution [Velosulin] |
| RxNorm | 363221 | insulin, regular, human Injectable Solution [Novolin R] |
| RxNorm | 343083 | insulin, regular, human 30 UNT/ML |
| RxNorm | 343258 | insulin, regular, human 50 UNT/ML |
| RxNorm | 360894 | insulin, regular, human 25 UNT/ML |
| RxNorm | 359125 | insulin, regular, human 15 UNT/ML |
| RxNorm | 359126 | insulin, regular, human 10 UNT/ML |
| RxNorm | 359127 | insulin, regular, human 20 UNT/ML |
| RxNorm | 365668 | insulin isophane / insulin, regular, human Injectable Suspension [Pur-In Mix] |
| RxNorm | 365672 | insulin isophane / insulin, regular, human Injectable Suspension [Novolin] |
| RxNorm | 365679 | insulin isophane / insulin, regular, human Injectable Suspension [Humulin] |
| RxNorm | 392660 | insulin isophane, human 75 UNT/ML / insulin, regular, human 25 UNT/ML Injectable Suspension |
| RxNorm | 378857 | insulin isophane / insulin, regular, human Injectable Suspension |
| RxNorm | 376915 | insulin, regular, human Injectable Solution |
| RxNorm | 412978 | insulin isophane, human 16 UNT/ML / insulin, regular, human 24 UNT/ML Injectable Suspension |
| RxNorm | 379734 | insulin, regular, human Injectable Solution [Humulin S] |
| RxNorm | 379740 | insulin isophane / insulin, regular, human Injectable Suspension [Human Mixtard] |
| RxNorm | 379744 | insulin, regular, human Injectable Solution [Human Actrapid] |
| RxNorm | 379747 | insulin, regular, human Injectable Solution [Human Actrapid Penfill] |
| RxNorm | 385895 | Pur-In Neutral |
| RxNorm | 385896 | insulin, regular, human Injectable Solution [Pur-In Neutral] |
| RxNorm | 412453 | insulin, regular, human 5 UNT/ML Injectable Solution |
| RxNorm | 415088 | insulin isophane, human 28 UNT/ML / insulin, regular, human 12 UNT/ML Injectable Suspension |

Continued on next page

**Table9 – continued from previous page**

| CodeSystem | ConceptCode | ConceptName |
|---|---|---|
| RxNorm | 415089 | insulin isophane, human 30 UNT/ML / insulin, regular, human 10 UNT/ML Injectable Suspension |
| RxNorm | 415090 | insulin isophane, human 34 UNT/ML / insulin, regular, human 6 UNT/ML Injectable Suspension |
| RxNorm | 415184 | insulin isophane, human 32 UNT/ML / insulin, regular, human 8 UNT/ML Injectable Suspension |
| RxNorm | 415185 | insulin isophane, human 36 UNT/ML / insulin, regular, human 4 UNT/ML Injectable Suspension |
| RxNorm | 440399 | insulin, regular, human 24 UNT/ML |
| RxNorm | 440650 | insulin, regular, human 12 UNT/ML |
| RxNorm | 440653 | insulin, regular, human 6 UNT/ML |
| RxNorm | 440654 | insulin, regular, human 5 UNT/ML |
| RxNorm | 451437 | insulin, regular, human 8 UNT/ML |
| RxNorm | 451439 | insulin, regular, human 4 UNT/ML |
| RxNorm | 564395 | insulin isophane, human 85 UNT/ML / insulin, regular, human 15 UNT/ML [Pur-In Mix] |
| RxNorm | 564396 | insulin isophane, human 75 UNT/ML / insulin, regular, human 25 UNT/ML [Pur-In Mix] |
| RxNorm | 564397 | insulin isophane, human 50 UNT/ML / insulin, regular, human 50 UNT/ML [Pur-In Mix] |
| RxNorm | 564399 | insulin isophane, human 90 UNT/ML / insulin, regular, human 10 UNT/ML [Humulin] |
| RxNorm | 564400 | insulin isophane, human 80 UNT/ML / insulin, regular, human 20 UNT/ML [Humulin] |
| RxNorm | 564401 | insulin isophane, human 60 UNT/ML / insulin, regular, human 40 UNT/ML [Humulin] |
| RxNorm | 5459 | Humulin S |
| RxNorm | 564531 | insulin isophane, human 50 UNT/ML / insulin, regular, human 50 UNT/ML [Humulin] |
| RxNorm | 564601 | insulin isophane, human 90 UNT/ML / insulin, regular, human 10 UNT/ML [Human Mixtard] |
| RxNorm | 564602 | insulin isophane, human 80 UNT/ML / insulin, regular, human 20 UNT/ML [Human Mixtard] |
| RxNorm | 564603 | insulin isophane, human 70 UNT/ML / insulin, regular, human 30 UNT/ML [Human Mixtard] |
| RxNorm | 564605 | insulin isophane, human 60 UNT/ML / insulin, regular, human 40 UNT/ML [Human Mixtard] |
| RxNorm | 564766 | insulin, regular, human 100 UNT/ML [Pur-In Neutral] |
| RxNorm | 564820 | insulin, regular, human 100 UNT/ML [Human Actrapid Penfill] |
| RxNorm | 564881 | insulin, regular, human 100 UNT/ML [Human Actrapid] |
| RxNorm | 564882 | insulin, regular, human 100 UNT/ML [Humulin S] |

Continued on next page

**Table9 – continued from previous page**

| CodeSystem | ConceptCode | ConceptName |
|---|---|---|
| RxNorm | 565176 | insulin isophane, human 50 UNT/ML / insulin, regular, human 50 UNT/ML [Human Mixtard] |
| RxNorm | 575146 | insulin, regular, human 100 UNT/ML [Novolin R] |
| RxNorm | 575147 | insulin, regular, human 100 UNT/ML [Velosulin] |
| RxNorm | 575148 | insulin, regular, human 100 UNT/ML [Humulin R] |
| RxNorm | 575628 | insulin, regular, human 500 UNT/ML [Humulin R] |
| RxNorm | 573330 | insulin isophane, human 70 UNT/ML / insulin, regular, human 30 UNT/ML [Humulin] |
| RxNorm | 573331 | insulin isophane, human 70 UNT/ML / insulin, regular, human 30 UNT/ML [Novolin] |
| RxNorm | 847186 | insulin isophane / insulin, regular, human Prefilled Syringe |
| RxNorm | 847187 | 3 ML insulin isophane, human 70 UNT/ML / insulin, regular, human 30 UNT/ML Pen Injector |
| RxNorm | 847188 | insulin isophane / insulin, regular, human Prefilled Syringe [Humulin] |
| RxNorm | 847189 | 3 ML insulin isophane, human 70 UNT/ML / insulin, regular, human 30 UNT/ML Pen Injector [Humulin] |
| RxNorm | 847194 | insulin isophane / insulin, regular, human Prefilled Syringe [Novolin] |
| RxNorm | 847202 | insulin, regular, human Prefilled Syringe |
| RxNorm | 847203 | 3 ML insulin, regular, human 100 UNT/ML Prefilled Syringe |
| RxNorm | 847204 | insulin, regular, human Prefilled Syringe [Novolin R] |
| RxNorm | 847205 | 3 ML insulin, regular, human 100 UNT/ML Prefilled Syringe [Novolin R] |
| RxNorm | 847256 | 1.5 ML insulin isophane, human 70 UNT/ML / insulin, regular, human 30 UNT/ML Prefilled Syringe |
| RxNorm | 847257 | 1.5 ML insulin isophane, human 70 UNT/ML / insulin, regular, human 30 UNT/ML Prefilled Syringe [Novolin] |
| RxNorm | 847343 | 1.5 ML insulin isophane, human 70 UNT/ML / insulin, regular, human 30 UNT/ML Prefilled Syringe [Humulin] |
| RxNorm | 847417 | 1.5 ML insulin, regular, human 100 UNT/ML Prefilled Syringe |
| RxNorm | 92881 | Humulin R |
| RxNorm | 93560 | Novolin R |
| LOINC | 4548-4 | Hemoglobin A1c/Hemoglobin.total in Blood |
| LOINC | 4549-2 | Hemoglobin A1c/Hemoglobin.total in Blood by Electrophoresis |
| LOINC | 17856-6 | Hemoglobin A1c/Hemoglobin.total in Blood by HPLC |
| LOINC | 59261-8 | Hemoglobin A1c/Hemoglobin.total in Blood by IFCC protocol |
| LOINC | 62388-4 | Hemoglobin A1c/Hemoglobin.total in Blood by JDS/JSCC protocol |

Continued on next page

**Table9 – continued from previous page**

| CodeSystem | ConceptCode | ConceptName |
|---|---|---|
| LOINC | 96595-4 | Hemoglobin A1c/Hemoglobin.total in DBS |

#### 8.3 Overweight and Obesity

Table 10: Concept codes used to identify overweight or obesity according to BMI >=27. Identification based on BMI observations, rather than diagnoses, required a numeric value of 27 or higher.

| CodeSystem | ConceptCode | ConceptName |
|---|---|---|
| ICD10CM | E66.01 | Morbid (severe) obesity due to excess calories |
| ICD10CM | E66.09 | Other obesity due to excess calories |
| ICD10CM | E66.1 | Drug-induced obesity |
| ICD10CM | E66.2 | Morbid (severe) obesity with alveolar hypoventilation |
| ICD10CM | E66.3 | Overweight |
| ICD10CM | E66.8 | Other obesity |
| ICD10CM | E66.9 | Obesity, unspecified |
| ICD10CM | Z68.30 | Body mass index (BMI) 30.0-30.9, adult |
| ICD10CM | Z68.31 | Body mass index (BMI) 31.0-31.9, adult |
| ICD10CM | Z68.32 | Body mass index (BMI) 32.0-32.9, adult |
| ICD10CM | Z68.33 | Body mass index (BMI) 33.0-33.9, adult |
| ICD10CM | Z68.34 | Body mass index (BMI) 34.0-34.9, adult |
| ICD10CM | Z68.35 | Body mass index (BMI) 35.0-35.9, adult |
| ICD10CM | Z68.36 | Body mass index (BMI) 36.0-36.9, adult |
| ICD10CM | Z68.37 | Body mass index (BMI) 37.0-37.9, adult |
| ICD10CM | Z68.38 | Body mass index (BMI) 38.0-38.9, adult |
| ICD10CM | Z68.39 | Body mass index (BMI) 39.0-39.9, adult |
| ICD10CM | Z68.41 | Body mass index (BMI) 40.0-44.9, adult |
| ICD10CM | Z68.42 | Body mass index (BMI) 45.0-49.9, adult |
| ICD10CM | Z68.43 | Body mass index (BMI) 50.0-59.9, adult |
| ICD10CM | Z68.44 | Body mass index (BMI) 60.0-69.9, adult |
| ICD10CM | Z68.45 | Body mass index (BMI) 70 or greater, adult |
| SNOMED CT | 83911000119104 | Severe obesity |
| SNOMED CT | 238131007 | Overweight |
| SNOMED CT | 238136002 | Morbid obesity |
| SNOMED CT | 162864005 | Body mass index 30+ - obesity |
| SNOMED CT | 190965006 | Drug-induced obesity |
| SNOMED CT | 190966007 | Extreme obesity with alveolar hypoventilation |
| SNOMED CT | 408512008 | Body mass index 40+ - severely obese |
| SNOMED CT | 414915002 | Obese |
| SNOMED CT | 414916001 | Obesity |
| SNOMED CT | 415530009 | Simple obesity |

Continued on next page

**Table10 – continued from previous page**

| CodeSystem | ConceptCode | ConceptName |
|---|---|---|
| SNOMED CT | 1229946007 | MAGEL2-related Prader-Willi-like syndrome |
| SNOMED CT | 1208987006 | PHIP-related behavioral problems, intellectual disability, obesity, dysmorphic features syndrome |
| SNOMED CT | 1229943004 | SIM1-related Prader-Willi-like syndrome |
| SNOMED CT | 1187531009 | Obesity due to pituitary disease |
| SNOMED CT | 788996008 | Obesity in adolescence |
| SNOMED CT | 819948005 | Obese class III |
| SNOMED CT | 171000119107 | Maternal obesity complicating pregnancy, childbirth and the puerperium, antepartum |
| SNOMED CT | 443371000124107 | Obese class I |
| SNOMED CT | 443381000124105 | Obese class II |
| SNOMED CT | 461341000124106 | Lower body obesity |
| SNOMED CT | 1076701000119104 | Hypertrophy of fat pad of right knee |
| SNOMED CT | 1076711000119101 | Hypertrophy of fat pad of left knee |
| SNOMED CT | 10750551000119100 | Obesity in mother complicating childbirth |
| SNOMED CT | 15750121000119108 | Severe obesity complicating pregnancy |
| SNOMED CT | 722037004 | Intellectual disability, epileptic seizures, hypogonadism and hypogenitalism, microcephaly, obesity syndrome |
| SNOMED CT | 80660001 | Mauriac's syndrome |
| SNOMED CT | 82793005 | Hypothalamic obesity |
| SNOMED CT | 111036000 | Hyperinsulinar obesity |
| SNOMED CT | 360566006 | Buffalo obesity |
| SNOMED CT | 238132000 | Android obesity |
| SNOMED CT | 238133005 | Gynecoid obesity |
| SNOMED CT | 238134004 | Generalized obesity |
| SNOMED CT | 238135003 | Fat pad syndrome |
| SNOMED CT | 248311001 | Central obesity |
| SNOMED CT | 248312008 | Peripheral obesity |
| SNOMED CT | 268915006 | O/E - weight 10-20% over ideal |
| SNOMED CT | 268916007 | O/E - weight greater than 20% over ideal |
| SNOMED CT | 270486005 | Localized adiposity |
| SNOMED CT | 275947003 | O/E - overweight |
| SNOMED CT | 290439001 | Familial obesity |
| SNOMED CT | 292464007 | Constitutional obesity |
| SNOMED CT | 293481008 | Hyperplastic-hypertrophic obesity |
| SNOMED CT | 294493008 | Lifelong obesity |
| SNOMED CT | 295509007 | Hypertrophic obesity |
| SNOMED CT | 296526005 | Adult-onset obesity |
| SNOMED CT | 297500005 | Endogenous obesity |
| SNOMED CT | 298464002 | Obesity of endocrine origin |
| SNOMED CT | 1003380001 | 6q16 microdeletion syndrome |
| SNOMED CT | 444862003 | Childhood obesity |
| SNOMED CT | 450451007 | Overweight in childhood |

Continued on next page

**Table10 – continued from previous page**

| CodeSystem | ConceptCode | ConceptName |
|---|---|---|
| SNOMED CT | 702949005 | Proopiomelanocortin deficiency syndrome |
| SNOMED CT | 715628009 | MORM syndrome |
| SNOMED CT | 717269008 | Obesity due to melanocortin 4 receptor deficiency |
| SNOMED CT | 717761005 | Choroideremia with deafness and obesity syndrome |
| SNOMED CT | 719160009 | Syndromic X-linked intellectual disability type 7 |
| SNOMED CT | 719834005 | Wilson Turner syndrome |
| SNOMED CT | 721231007 | Hydrocephalus with obesity and hypogonadism syndrome |
| SNOMED CT | 722051004 | Obesity, colitis, hypothyroidism, cardiac hypertrophy, developmental delay syndrome |
| SNOMED CT | 722053001 | Obesity due to prohormone convertase I deficiency |
| SNOMED CT | 722595002 | Overweight in adulthood with body mass index of 25 or more but less than 30 |
| SNOMED CT | 722596001 | Obesity caused by energy imbalance |
| SNOMED CT | 724137002 | MOMO syndrome |
| SNOMED CT | 763350002 | Intellectual disability, obesity, brain malformation, facial dysmorphism syndrome |
| SNOMED CT | 770680004 | Prader-Willi-like syndrome |
| SNOMED CT | 770750002 | Intellectual disability, seizures, macrocephaly, obesity syndrome |
| SNOMED CT | 773663004 | Rapid-onset childhood obesity, hypothalamic dysfunction, hypoventilation, autonomic dysregulation syndrome |
| SNOMED CT | 774102003 | Intellectual disability, obesity, prognathism, eye and skin anomalies syndrome |
| SNOMED CT | 776204008 | Colobomatous microphthalmia, obesity, hypogenitalism, intellectual disability syndrome |
| SNOMED CT | 783549006 | Obesity due to CEP19 deficiency |
| SNOMED CT | 783556000 | Severe early-onset obesity insulin resistance syndrome due to SH2B1 deficiency |
| SNOMED CT | 783719006 | Obesity due to SIM1 deficiency |
| SNOMED CT | 785722006 | Obesity due to leptin receptor gene deficiency |
| SNOMED CT | 162690006 | O/E - obese |
| SNOMED CT | 162863004 | Body mass index 25-29 - overweight |
| SNOMED CT | 5036006 | Hypogonadal obesity |
| SNOMED CT | 44772007 | Maternal obesity syndrome |
| SNOMED CT | 53146006 | Hypothyroid obesity |
| SNOMED CT | 57337005 | Steatopygia |
| SNOMED CT | 62999006 | Adiposogenital dystrophy |
| SNOMED CT | 63702009 | Alstrom syndrome |
| SNOMED CT | 72894001 | Hypertrophy of fat pad of knee |
| SNOMED CT | 414438005 | Hyperplastic obesity |
| SNOMED CT | 414917005 | Obesity by adipocyte growth pattern |
| SNOMED CT | 414918000 | Obesity by age of onset |
| SNOMED CT | 414919008 | Obesity by contributing factors |

Continued on next page

**Table10 – continued from previous page**

| CodeSystem | ConceptCode | ConceptName |
|---|---|---|
| SNOMED CT | 414920002 | Obesity by fat distribution pattern |
| ICD10CM | E66.3 | Overweight |
| ICD10CM | Z68.27 | Body mass index (BMI) 27.0-27.9, adult |
| ICD10CM | Z68.28 | Body mass index (BMI) 28.0-28.9, adult |
| ICD10CM | Z68.29 | Body mass index (BMI) 29.0-29.9, adult |
| SNOMED CT | 238131007 | Overweight |
| SNOMED CT | 268915006 | O/E - weight 10-20% over ideal |
| SNOMED CT | 268916007 | O/E - weight greater than 20% over ideal |
| SNOMED CT | 275947003 | O/E - overweight |
| SNOMED CT | 450451007 | Overweight in childhood |
| SNOMED CT | 722595002 | Overweight in adulthood with body mass index of 25 or more but less than 30 |
| SNOMED CT | 162863004 | Body mass index 25-29 - overweight |
| LOINC | 39156-5 | Body mass index (BMI) [Ratio] |

#### 8.4 Comborbidities

##### 8.4.1 Asthma

Table 11: Concept codes used to identify if a patient was diagnosed with asthma.

| CodeSystem | ConceptCode | ConceptName |
|---|---|---|
| ICD10CM | J45 | Asthma |
| ICD10CM | J45.2 | Mild intermittent asthma |
| ICD10CM | J45.20 | Mild intermittent asthma, uncomplicated |
| ICD10CM | J45.21 | Mild intermittent asthma with (acute) exacerbation |
| ICD10CM | J45.22 | Mild intermittent asthma with status asthmaticus |
| ICD10CM | J45.3 | Mild persistent asthma |
| ICD10CM | J45.30 | Mild persistent asthma, uncomplicated |
| ICD10CM | J45.31 | Mild persistent asthma with (acute) exacerbation |
| ICD10CM | J45.32 | Mild persistent asthma with status asthmaticus |
| ICD10CM | J45.4 | Moderate persistent asthma |
| ICD10CM | J45.40 | Moderate persistent asthma, uncomplicated |
| ICD10CM | J45.41 | Moderate persistent asthma with (acute) exacerbation |
| ICD10CM | J45.42 | Moderate persistent asthma with status asthmaticus |
| ICD10CM | J45.5 | Severe persistent asthma |
| ICD10CM | J45.50 | Severe persistent asthma, uncomplicated |
| ICD10CM | J45.51 | Severe persistent asthma with (acute) exacerbation |
| ICD10CM | J45.52 | Severe persistent asthma with status asthmaticus |
| ICD10CM | J45.9 | Other and unspecified asthma |
| ICD10CM | J45.90 | Unspecified asthma |

Continued on next page

**Table11 – continued from previous page**

| CodeSystem | ConceptCode | ConceptName |
|---|---|---|
| ICD10CM | J45.901 | Unspecified asthma with (acute) exacerbation |
| ICD10CM | J45.902 | Unspecified asthma with status asthmaticus |
| ICD10CM | J45.909 | Unspecified asthma, uncomplicated |
| ICD10CM | J45.99 | Other asthma |
| ICD10CM | J45.990 | Exercise induced bronchospasm |
| ICD10CM | J45.991 | Cough variant asthma |
| ICD10CM | J45.998 | Other asthma |
| SNOMED CT | 707446004 | Exacerbation of moderate persistent asthma |
| SNOMED CT | 707447008 | Exacerbation of severe persistent asthma |
| SNOMED CT | 707512002 | Uncomplicated moderate persistent asthma |
| SNOMED CT | 707513007 | Uncomplicated severe persistent asthma |
| SNOMED CT | 707979007 | Acute severe exacerbation of severe persistent asthma |
| SNOMED CT | 707980005 | Acute severe exacerbation of moderate persistent asthma |
| SNOMED CT | 707981009 | Acute severe exacerbation of mild persistent asthma |
| SNOMED CT | 708090002 | Acute severe exacerbation of asthma |
| SNOMED CT | 708093000 | Acute exacerbation of allergic asthma |
| SNOMED CT | 708094006 | Acute exacerbation of intrinsic asthma |
| SNOMED CT | 708095007 | Acute severe exacerbation of immunoglobin E-mediated allergic asthma |
| SNOMED CT | 708096008 | Acute severe exacerbation of intrinsic asthma |
| SNOMED CT | 1751000119100 | Acute exacerbation of chronic obstructive airways disease with asthma |
| SNOMED CT | 99031000119107 | Acute exacerbation of asthma co-occurrent with allergic rhinitis |
| SNOMED CT | 124991000119109 | Severe persistent asthma co-occurrent with allergic rhinitis |
| SNOMED CT | 125001000119103 | Moderate persistent asthma co-occurrent with allergic rhinitis |
| SNOMED CT | 10674711000119105 | Acute severe exacerbation of asthma co-occurrent with allergic rhinitis |
| SNOMED CT | 10675431000119106 | Severe persistent allergic asthma |
| SNOMED CT | 10675471000119109 | Acute severe exacerbation of severe persistent allergic asthma |
| SNOMED CT | 10675551000119104 | Acute severe exacerbation of severe persistent asthma co-occurrent with allergic rhinitis |
| SNOMED CT | 10675911000119109 | Acute severe exacerbation of mild persistent allergic asthma |
| SNOMED CT | 10675991000119100 | Acute severe exacerbation of mild persistent allergic asthma co-occurrent with allergic rhinitis |
| SNOMED CT | 10676391000119108 | Moderate persistent allergic asthma |
| SNOMED CT | 10676431000119103 | Acute severe exacerbation of moderate persistent allergic asthma |
| SNOMED CT | 10676511000119109 | Acute severe exacerbation of moderate persistent asthma co-occurrent with allergic rhinitis |

Continued on next page

**Table11 – continued from previous page**

| CodeSystem | ConceptCode | ConceptName |
|---|---|---|
| SNOMED CT | 10692721000119102 | Chronic obstructive asthma co-occurrent with acute exacerbation of asthma |
| SNOMED CT | 426656000 | Severe persistent asthma |
| SNOMED CT | 427295004 | Moderate persistent asthma |
| SNOMED CT | 442025000 | Acute exacerbation of chronic asthmatic bronchitis |
| SNOMED CT | 370219009 | Moderate asthma |
| SNOMED CT | 370221004 | Severe asthma |
| SNOMED CT | 786836003 | Near fatal asthma |
| SNOMED CT | 829976001 | Thunderstorm asthma |
| SNOMED CT | 135171000119106 | Acute exacerbation of moderate persistent asthma |
| SNOMED CT | 10675391000119101 | Severe controlled persistent asthma |
| SNOMED CT | 10675751000119107 | Severe uncontrolled persistent asthma |
| SNOMED CT | 733858005 | Acute severe refractory exacerbation of asthma |
| SNOMED CT | 734904007 | Life threatening acute exacerbation of asthma |
| SNOMED CT | 782513000 | Acute severe exacerbation of allergic asthma |

##### 8.4.2 Atrial Fibrillation

Table 12: Concept codes used to identify if a patient was diagnosed with atrial-fibrillation-flutter.

| CodeSystem | ConceptCode | ConceptName |
|---|---|---|
| ICD10CM | I48.0 | Paroxysmal atrial fibrillation |
| ICD10CM | I48.11 | Longstanding persistent atrial fibrillation |
| ICD10CM | I48.19 | Other persistent atrial fibrillation |
| ICD10CM | I48.20 | Chronic atrial fibrillation, unspecified |
| ICD10CM | I48.21 | Permanent atrial fibrillation |
| ICD10CM | I48.3 | Typical atrial flutter |
| ICD10CM | I48.4 | Atypical atrial flutter |
| ICD10CM | I48.91 | Unspecified atrial fibrillation |
| ICD10CM | I48.92 | Unspecified atrial flutter |
| ICD9CM | 427.31 | Atrial fibrillation |
| ICD9CM | 427.32 | Atrial flutter |
| SNOMED CT | 120041000119109 | Atrial fibrillation with rapid ventricular response |
| SNOMED CT | 15964901000119107 | Atypical atrial flutter |
| SNOMED CT | 5370000 | Atrial flutter |
| SNOMED CT | 706923002 | Longstanding persistent atrial fibrillation |
| SNOMED CT | 715395008 | Familial atrial fibrillation |
| SNOMED CT | 720448006 | Typical atrial flutter |
| SNOMED CT | 195080001 | Atrial fibrillation and flutter |
| SNOMED CT | 314208002 | Rapid atrial fibrillation |
| SNOMED CT | 233910005 | Lone atrial fibrillation |

Continued on next page

**Table12 – continued from previous page**

| CodeSystem | ConceptCode | ConceptName |
|---|---|---|
| SNOMED CT | 233911009 | Non-rheumatic atrial fibrillation |
| SNOMED CT | 49436004 | Atrial fibrillation |
| SNOMED CT | 425615007 | Chronic atrial flutter |
| SNOMED CT | 426749004 | Chronic atrial fibrillation |
| SNOMED CT | 426814001 | Transient cerebral ischemia due to atrial fibrillation |
| SNOMED CT | 427665004 | Paroxysmal atrial flutter |
| SNOMED CT | 440028005 | Permanent atrial fibrillation |
| SNOMED CT | 440059007 | Persistent atrial fibrillation |
| SNOMED CT | 282825002 | Paroxysmal atrial fibrillation |
| SNOMED CT | 300996004 | Controlled atrial fibrillation |
| SNOMED CT | 715560009 | Idiopathic neonatal atrial flutter |
| SNOMED CT | 762247006 | Preexcited atrial fibrillation |
| SNOMED CT | 1010405004 | Paroxysmal atrial fibrillation with rapid ventricular response |

##### 8.4.3 Bariatric Surgery

Table 13: Concept codes used to identify if a patient was diagnosed with bariatric-surgery.

| CodeSystem | ConceptCode | ConceptName |
|---|---|---|
| ICD10CM | Z98.84 | Bariatric surgery status |
| SNOMED CT | 329281000119107 | History of sleeve gastrectomy |
| SNOMED CT | 608848006 | History of bariatric surgical procedure |
| SNOMED CT | 427074001 | Laparoscopic sleeve gastrectomy |
| SNOMED CT | 430715008 | Bariatric operative procedure |
| SNOMED CT | 442338001 | Bypass of stomach |
| SNOMED CT | 18692006 | Bypass gastroenterostomy |
| SNOMED CT | 30803004 | Printen and Mason operation, high gastric bypass |
| SNOMED CT | 1172524007 | Primary obesity surgery endoluminal 2 |
| SNOMED CT | 708629005 | Laparoscopic bypass gastrojejunostomy |
| SNOMED CT | 708983005 | Laparoscopic bypass of stomach |
| SNOMED CT | 173587003 | Bypass of esophagus using stomach |
| SNOMED CT | 173748000 | Bypass of stomach by anastomosis of stomach to transposed jejunum |
| SNOMED CT | 173750008 | Conversion to anastomosis of stomach to transposed jejunum |
| SNOMED CT | 173751007 | Conversion from previous anastomosis of stomach to transposed jejunum |
| SNOMED CT | 173857001 | Bypass of duodenum by anastomosis of stomach to jejunum |

Continued on next page

**Table13 – continued from previous page**

| CodeSystem | ConceptCode | ConceptName |
|---|---|---|
| ICD10PCS | 0D11076 | Bypass Upper Esophagus to Stomach with Autologous Tissue Substitute, Open Approach |
| ICD10PCS | 0D110J6 | Bypass Upper Esophagus to Stomach with Synthetic Substitute, Open Approach |
| ICD10PCS | 0D110K6 | Bypass Upper Esophagus to Stomach with Nonautologous Tissue Substitute, Open Approach |
| ICD10PCS | 0D110Z6 | Bypass Upper Esophagus to Stomach, Open Approach |
| ICD10PCS | 0D11476 | Bypass Upper Esophagus to Stomach with Autologous Tissue Substitute, Percutaneous Endoscopic Approach |
| ICD10PCS | 0D114J6 | Bypass Upper Esophagus to Stomach with Synthetic Substitute, Percutaneous Endoscopic Approach |
| ICD10PCS | 0D114K6 | Bypass Upper Esophagus to Stomach with Nonautologous Tissue Substitute, Percutaneous Endoscopic Approach |
| ICD10PCS | 0D114Z6 | Bypass Upper Esophagus to Stomach, Percutaneous Endoscopic Approach |
| ICD10PCS | 0D11876 | Bypass Upper Esophagus to Stomach with Autologous Tissue Substitute, Via Natural or Artificial Opening Endoscopic |
| ICD10PCS | 0D118J6 | Bypass Upper Esophagus to Stomach with Synthetic Substitute, Via Natural or Artificial Opening Endoscopic |
| ICD10PCS | 0D118K6 | Bypass Upper Esophagus to Stomach with Nonautologous Tissue Substitute, Via Natural or Artificial Opening Endoscopic |
| ICD10PCS | 0D118Z6 | Bypass Upper Esophagus to Stomach, Via Natural or Artificial Opening Endoscopic |
| ICD10PCS | 0D12076 | Bypass Middle Esophagus to Stomach with Autologous Tissue Substitute, Open Approach |
| ICD10PCS | 0D120J6 | Bypass Middle Esophagus to Stomach with Synthetic Substitute, Open Approach |
| ICD10PCS | 0D120K6 | Bypass Middle Esophagus to Stomach with Nonautologous Tissue Substitute, Open Approach |
| ICD10PCS | 0D120Z6 | Bypass Middle Esophagus to Stomach, Open Approach |
| ICD10PCS | 0D12476 | Bypass Middle Esophagus to Stomach with Autologous Tissue Substitute, Percutaneous Endoscopic Approach |
| ICD10PCS | 0D124J6 | Bypass Middle Esophagus to Stomach with Synthetic Substitute, Percutaneous Endoscopic Approach |
| ICD10PCS | 0D124K6 | Bypass Middle Esophagus to Stomach with Nonautologous Tissue Substitute, Percutaneous Endoscopic Approach |
| ICD10PCS | 0D124Z6 | Bypass Middle Esophagus to Stomach, Percutaneous Endoscopic Approach |
| ICD10PCS | 0D12876 | Bypass Middle Esophagus to Stomach with Autologous Tissue Substitute, Via Natural or Artificial Opening Endoscopic |

Continued on next page

**Table13 – continued from previous page**

| CodeSystem | ConceptCode | ConceptName |
|---|---|---|
| ICD10PCS | 0D128J6 | Bypass Middle Esophagus to Stomach with Synthetic Substitute, Via Natural or Artificial Opening Endoscopic |
| ICD10PCS | 0D128K6 | Bypass Middle Esophagus to Stomach with Nonautologous Tissue Substitute, Via Natural or Artificial Opening Endoscopic |
| ICD10PCS | 0D128Z6 | Bypass Middle Esophagus to Stomach, Via Natural or Artificial Opening Endoscopic |
| ICD10PCS | 0D13076 | Bypass Lower Esophagus to Stomach with Autologous Tissue Substitute, Open Approach |
| ICD10PCS | 0D130J6 | Bypass Lower Esophagus to Stomach with Synthetic Substitute, Open Approach |
| ICD10PCS | 0D130K6 | Bypass Lower Esophagus to Stomach with Nonautologous Tissue Substitute, Open Approach |
| ICD10PCS | 0D130Z6 | Bypass Lower Esophagus to Stomach, Open Approach |
| ICD10PCS | 0D13476 | Bypass Lower Esophagus to Stomach with Autologous Tissue Substitute, Percutaneous Endoscopic Approach |
| ICD10PCS | 0D134J6 | Bypass Lower Esophagus to Stomach with Synthetic Substitute, Percutaneous Endoscopic Approach |
| ICD10PCS | 0D134K6 | Bypass Lower Esophagus to Stomach with Nonautologous Tissue Substitute, Percutaneous Endoscopic Approach |
| ICD10PCS | 0D134Z6 | Bypass Lower Esophagus to Stomach, Percutaneous Endoscopic Approach |
| ICD10PCS | 0D13876 | Bypass Lower Esophagus to Stomach with Autologous Tissue Substitute, Via Natural or Artificial Opening Endoscopic |
| ICD10PCS | 0D138J6 | Bypass Lower Esophagus to Stomach with Synthetic Substitute, Via Natural or Artificial Opening Endoscopic |
| ICD10PCS | 0D138K6 | Bypass Lower Esophagus to Stomach with Nonautologous Tissue Substitute, Via Natural or Artificial Opening Endoscopic |
| ICD10PCS | 0D138Z6 | Bypass Lower Esophagus to Stomach, Via Natural or Artificial Opening Endoscopic |
| ICD10PCS | 0D15076 | Bypass Esophagus to Stomach with Autologous Tissue Substitute, Open Approach |
| ICD10PCS | 0D150J6 | Bypass Esophagus to Stomach with Synthetic Substitute, Open Approach |
| ICD10PCS | 0D150K6 | Bypass Esophagus to Stomach with Nonautologous Tissue Substitute, Open Approach |
| ICD10PCS | 0D150Z6 | Bypass Esophagus to Stomach, Open Approach |
| ICD10PCS | 0D15476 | Bypass Esophagus to Stomach with Autologous Tissue Substitute, Percutaneous Endoscopic Approach |
| ICD10PCS | 0D154J6 | Bypass Esophagus to Stomach with Synthetic Substitute, Percutaneous Endoscopic Approach |

Continued on next page

**Table13 – continued from previous page**

| CodeSystem | ConceptCode | ConceptName |
|---|---|---|
| ICD10PCS | 0D154K6 | Bypass Esophagus to Stomach with Nonautologous Tissue Substitute, Percutaneous Endoscopic Approach |
| ICD10PCS | 0D154Z6 | Bypass Esophagus to Stomach, Percutaneous Endoscopic Approach |
| ICD10PCS | 0D15876 | Bypass Esophagus to Stomach with Autologous Tissue Substitute, Via Natural or Artificial Opening Endoscopic |
| ICD10PCS | 0D158J6 | Bypass Esophagus to Stomach with Synthetic Substitute, Via Natural or Artificial Opening Endoscopic |
| ICD10PCS | 0D158K6 | Bypass Esophagus to Stomach with Nonautologous Tissue Substitute, Via Natural or Artificial Opening Endoscopic |
| ICD10PCS | 0D158Z6 | Bypass Esophagus to Stomach, Via Natural or Artificial Opening Endoscopic |
| ICD10PCS | 0D16074 | Bypass Stomach to Cutaneous with Autologous Tissue Substitute, Open Approach |
| ICD10PCS | 0D16079 | Bypass Stomach to Duodenum with Autologous Tissue Substitute, Open Approach |
| ICD10PCS | 0D1607A | Bypass Stomach to Jejunum with Autologous Tissue Substitute, Open Approach |
| ICD10PCS | 0D1607B | Bypass Stomach to Ileum with Autologous Tissue Substitute, Open Approach |
| ICD10PCS | 0D1607L | Bypass Stomach to Transverse Colon with Autologous Tissue Substitute, Open Approach |
| ICD10PCS | 0D160J4 | Bypass Stomach to Cutaneous with Synthetic Substitute, Open Approach |
| ICD10PCS | 0D160J9 | Bypass Stomach to Duodenum with Synthetic Substitute, Open Approach |
| ICD10PCS | 0D160JA | Bypass Stomach to Jejunum with Synthetic Substitute, Open Approach |
| ICD10PCS | 0D160JB | Bypass Stomach to Ileum with Synthetic Substitute, Open Approach |
| ICD10PCS | 0D160JL | Bypass Stomach to Transverse Colon with Synthetic Substitute, Open Approach |
| ICD10PCS | 0D160K4 | Bypass Stomach to Cutaneous with Nonautologous Tissue Substitute, Open Approach |
| ICD10PCS | 0D160K9 | Bypass Stomach to Duodenum with Nonautologous Tissue Substitute, Open Approach |
| ICD10PCS | 0D160KA | Bypass Stomach to Jejunum with Nonautologous Tissue Substitute, Open Approach |
| ICD10PCS | 0D160KB | Bypass Stomach to Ileum with Nonautologous Tissue Substitute, Open Approach |
| ICD10PCS | 0D160KL | Bypass Stomach to Transverse Colon with Nonautologous Tissue Substitute, Open Approach |
| ICD10PCS | 0D160Z4 | Bypass Stomach to Cutaneous, Open Approach |

Continued on next page

**Table13 – continued from previous page**

| CodeSystem | ConceptCode | ConceptName |
|---|---|---|
| ICD10PCS | 0D160Z9 | Bypass Stomach to Duodenum, Open Approach |
| ICD10PCS | 0D160ZA | Bypass Stomach to Jejunum, Open Approach |
| ICD10PCS | 0D160ZB | Bypass Stomach to Ileum, Open Approach |
| ICD10PCS | 0D160ZL | Bypass Stomach to Transverse Colon, Open Approach |
| ICD10PCS | 0D163J4 | Bypass Stomach to Cutaneous with Synthetic Substitute, Percutaneous Approach |
| ICD10PCS | 0D16474 | Bypass Stomach to Cutaneous with Autologous Tissue Substitute, Percutaneous Endoscopic Approach |
| ICD10PCS | 0D16479 | Bypass Stomach to Duodenum with Autologous Tissue Substitute, Percutaneous Endoscopic Approach |
| ICD10PCS | 0D1647A | Bypass Stomach to Jejunum with Autologous Tissue Substitute, Percutaneous Endoscopic Approach |
| ICD10PCS | 0D1647B | Bypass Stomach to Ileum with Autologous Tissue Substitute, Percutaneous Endoscopic Approach |
| ICD10PCS | 0D1647L | Bypass Stomach to Transverse Colon with Autologous Tissue Substitute, Percutaneous Endoscopic Approach |
| ICD10PCS | 0D164J4 | Bypass Stomach to Cutaneous with Synthetic Substitute, Percutaneous Endoscopic Approach |
| ICD10PCS | 0D164J9 | Bypass Stomach to Duodenum with Synthetic Substitute, Percutaneous Endoscopic Approach |
| ICD10PCS | 0D164JA | Bypass Stomach to Jejunum with Synthetic Substitute, Percutaneous Endoscopic Approach |
| ICD10PCS | 0D164JB | Bypass Stomach to Ileum with Synthetic Substitute, Percutaneous Endoscopic Approach |
| ICD10PCS | 0D164JL | Bypass Stomach to Transverse Colon with Synthetic Substitute, Percutaneous Endoscopic Approach |
| ICD10PCS | 0D164K4 | Bypass Stomach to Cutaneous with Nonautologous Tissue Substitute, Percutaneous Endoscopic Approach |
| ICD10PCS | 0D164K9 | Bypass Stomach to Duodenum with Nonautologous Tissue Substitute, Percutaneous Endoscopic Approach |
| ICD10PCS | 0D164KA | Bypass Stomach to Jejunum with Nonautologous Tissue Substitute, Percutaneous Endoscopic Approach |
| ICD10PCS | 0D164KB | Bypass Stomach to Ileum with Nonautologous Tissue Substitute, Percutaneous Endoscopic Approach |
| ICD10PCS | 0D164KL | Bypass Stomach to Transverse Colon with Nonautologous Tissue Substitute, Percutaneous Endoscopic Approach |
| ICD10PCS | 0D164Z4 | Bypass Stomach to Cutaneous, Percutaneous Endoscopic Approach |
| ICD10PCS | 0D164Z9 | Bypass Stomach to Duodenum, Percutaneous Endoscopic Approach |
| ICD10PCS | 0D164ZA | Bypass Stomach to Jejunum, Percutaneous Endoscopic Approach |

Continued on next page

**Table13 – continued from previous page**

| CodeSystem | ConceptCode | ConceptName |
|---|---|---|
| ICD10PCS | 0D164ZB | Bypass Stomach to Ileum, Percutaneous Endoscopic Approach |
| ICD10PCS | 0D164ZL | Bypass Stomach to Transverse Colon, Percutaneous Endoscopic Approach |
| ICD10PCS | 0D16874 | Bypass Stomach to Cutaneous with Autologous Tissue Substitute, Via Natural or Artificial Opening Endoscopic |
| ICD10PCS | 0D16879 | Bypass Stomach to Duodenum with Autologous Tissue Substitute, Via Natural or Artificial Opening Endoscopic |
| ICD10PCS | 0D1687A | Bypass Stomach to Jejunum with Autologous Tissue Substitute, Via Natural or Artificial Opening Endoscopic |
| ICD10PCS | 0D1687B | Bypass Stomach to Ileum with Autologous Tissue Substitute, Via Natural or Artificial Opening Endoscopic |
| ICD10PCS | 0D1687L | Bypass Stomach to Transverse Colon with Autologous Tissue Substitute, Via Natural or Artificial Opening Endoscopic |
| ICD10PCS | 0D168J4 | Bypass Stomach to Cutaneous with Synthetic Substitute, Via Natural or Artificial Opening Endoscopic |
| ICD10PCS | 0D168J9 | Bypass Stomach to Duodenum with Synthetic Substitute, Via Natural or Artificial Opening Endoscopic |
| ICD10PCS | 0D168JA | Bypass Stomach to Jejunum with Synthetic Substitute, Via Natural or Artificial Opening Endoscopic |
| ICD10PCS | 0D168JB | Bypass Stomach to Ileum with Synthetic Substitute, Via Natural or Artificial Opening Endoscopic |
| ICD10PCS | 0D168JL | Bypass Stomach to Transverse Colon with Synthetic Substitute, Via Natural or Artificial Opening Endoscopic |
| ICD10PCS | 0D168K4 | Bypass Stomach to Cutaneous with Nonautologous Tissue Substitute, Via Natural or Artificial Opening Endoscopic |
| ICD10PCS | 0D168K9 | Bypass Stomach to Duodenum with Nonautologous Tissue Substitute, Via Natural or Artificial Opening Endoscopic |
| ICD10PCS | 0D168KA | Bypass Stomach to Jejunum with Nonautologous Tissue Substitute, Via Natural or Artificial Opening Endoscopic |
| ICD10PCS | 0D168KB | Bypass Stomach to Ileum with Nonautologous Tissue Substitute, Via Natural or Artificial Opening Endoscopic |
| ICD10PCS | 0D168KL | Bypass Stomach to Transverse Colon with Nonautologous Tissue Substitute, Via Natural or Artificial Opening Endoscopic |
| ICD10PCS | 0D168Z4 | Bypass Stomach to Cutaneous, Via Natural or Artificial Opening Endoscopic |
| ICD10PCS | 0D168Z9 | Bypass Stomach to Duodenum, Via Natural or Artificial Opening Endoscopic |
| ICD10PCS | 0D168ZA | Bypass Stomach to Jejunum, Via Natural or Artificial Opening Endoscopic |

Continued on next page

**Table13 – continued from previous page**

| CodeSystem | ConceptCode | ConceptName |
|---|---|---|
| ICD10PCS | 0D168ZB | Bypass Stomach to Ileum, Via Natural or Artificial Opening Endoscopic |
| ICD10PCS | 0D168ZL | Bypass Stomach to Transverse Colon, Via Natural or Artificial Opening Endoscopic |
| ICD10PCS | 0F170Z4 | Bypass Common Hepatic Duct to Stomach, Open Approach |
| SNOMED CT | 870378000 | Endoscopic sleeve gastroplasty |
| SNOMED CT | 173747005 | Connection of stomach to transposed jejunum |
| SNOMED CT | 314593003 | Posterior gastrojejunostomy |
| SNOMED CT | 358575006 | Gastroduodenostomy |
| SNOMED CT | 359529008 | Jaboulay operation, gastroduodenostomy |
| SNOMED CT | 235281005 | Anterior gastrojejunostomy |
| SNOMED CT | 235282003 | Anastomosis of stomach to ileum |
| SNOMED CT | 45753003 | Cholecystenterostomy with gastroenterostomy |
| SNOMED CT | 49245001 | Bypass gastrojejunostomy |
| SNOMED CT | 57245002 | Pylorostomy |
| SNOMED CT | 433024005 | Gastrojejunostomy using fluoroscopic guidance |
| SNOMED CT | 443906008 | Bypass of stomach with short limb Roux-en-Y gastroenterostomy |
| SNOMED CT | 265365007 | Bypass of stomach by anastomosis of stomach to duodenum |
| SNOMED CT | 287834004 | Gastroenterostomy - no gastrectomy |
| ICD10PCS | 0F140D4 | Bypass Gallbladder to Stomach with Intraluminal Device, Open Approach |
| ICD10PCS | 0F140Z4 | Bypass Gallbladder to Stomach, Open Approach |
| ICD10PCS | 0F144D4 | Bypass Gallbladder to Stomach with Intraluminal Device, Percutaneous Endoscopic Approach |
| ICD10PCS | 0F144Z4 | Bypass Gallbladder to Stomach, Percutaneous Endoscopic Approach |
| ICD10PCS | 0F150D4 | Bypass Right Hepatic Duct to Stomach with Intraluminal Device, Open Approach |
| ICD10PCS | 0F150Z4 | Bypass Right Hepatic Duct to Stomach, Open Approach |
| ICD10PCS | 0F160Z4 | Bypass Left Hepatic Duct to Stomach, Open Approach |
| ICD10PCS | 0F154D4 | Bypass Right Hepatic Duct to Stomach with Intraluminal Device, Percutaneous Endoscopic Approach |
| ICD10PCS | 0F154Z4 | Bypass Right Hepatic Duct to Stomach, Percutaneous Endoscopic Approach |
| ICD10PCS | 0F160D4 | Bypass Left Hepatic Duct to Stomach with Intraluminal Device, Open Approach |
| ICD10PCS | 0F164D4 | Bypass Left Hepatic Duct to Stomach with Intraluminal Device, Percutaneous Endoscopic Approach |
| ICD10PCS | 0F164Z4 | Bypass Left Hepatic Duct to Stomach, Percutaneous Endoscopic Approach |
| ICD10PCS | 0F170D4 | Bypass Common Hepatic Duct to Stomach with Intraluminal Device, Open Approach |

Continued on next page

**Table13 – continued from previous page**

| CodeSystem | ConceptCode | ConceptName |
|---|---|---|
| ICD10PCS | 0F174D4 | Bypass Common Hepatic Duct to Stomach with Intraluminal Device, Percutaneous Endoscopic Approach |
| ICD10PCS | 0F174Z4 | Bypass Common Hepatic Duct to Stomach, Percutaneous Endoscopic Approach |
| ICD10PCS | 0F180D4 | Bypass Cystic Duct to Stomach with Intraluminal Device, Open Approach |
| ICD10PCS | 0F180Z4 | Bypass Cystic Duct to Stomach, Open Approach |
| ICD10PCS | 0F184D4 | Bypass Cystic Duct to Stomach with Intraluminal Device, Percutaneous Endoscopic Approach |
| ICD10PCS | 0F184Z4 | Bypass Cystic Duct to Stomach, Percutaneous Endoscopic Approach |
| ICD10PCS | 0F190D4 | Bypass Common Bile Duct to Stomach with Intraluminal Device, Open Approach |
| ICD10PCS | 0F190Z4 | Bypass Common Bile Duct to Stomach, Open Approach |
| ICD10PCS | 0F194D4 | Bypass Common Bile Duct to Stomach with Intraluminal Device, Percutaneous Endoscopic Approach |
| ICD10PCS | 0F194Z4 | Bypass Common Bile Duct to Stomach, Percutaneous Endoscopic Approach |
| ICD10PCS | 0F1D0D4 | Bypass Pancreatic Duct to Stomach with Intraluminal Device, Open Approach |
| ICD10PCS | 0F1D0Z4 | Bypass Pancreatic Duct to Stomach, Open Approach |
| ICD10PCS | 0F1D4D4 | Bypass Pancreatic Duct to Stomach with Intraluminal Device, Percutaneous Endoscopic Approach |
| ICD10PCS | 0F1D4Z4 | Bypass Pancreatic Duct to Stomach, Percutaneous Endoscopic Approach |
| ICD9CM | V45.86 | Bariatric surgery status |

###### 8.4.4 Chronic Kidney Disease

Table 14: Concept codes used to identify if a patient was diagnosed with chronic-kidney-disease.

| CodeSystem | ConceptCode | ConceptName |
|---|---|---|
| ICD10CM | D63.1 | Anemia in chronic kidney disease |
| ICD10CM | E08.22 | Diabetes mellitus due to underlying condition with diabetic chronic kidney disease |
| ICD10CM | E09.22 | Drug or chemical induced diabetes mellitus with diabetic chronic kidney disease |
| ICD10CM | E13.22 | Other specified diabetes mellitus with diabetic chronic kidney disease |
| ICD10CM | I12 | Hypertensive chronic kidney disease |

Continued on next page

**Table14 – continued from previous page**

| CodeSystem | ConceptCode | ConceptName |
|---|---|---|
| ICD10CM | I12.0 | Hypertensive chronic kidney disease with stage 5 chronic kidney disease or end stage renal disease |
| ICD10CM | I12.9 | Hypertensive chronic kidney disease with stage 1 through stage 4 chronic kidney disease, or unspecified chronic kidney disease |
| ICD10CM | I13 | Hypertensive heart and chronic kidney disease |
| ICD10CM | I13.0 | Hypertensive heart and chronic kidney disease with heart failure and stage 1 through stage 4 chronic kidney disease, or unspecified chronic kidney disease |
| ICD10CM | I13.1 | Hypertensive heart and chronic kidney disease without heart failure |
| ICD10CM | I13.10 | Hypertensive heart and chronic kidney disease without heart failure, with stage 1 through stage 4 chronic kidney disease, or unspecified chronic kidney disease |
| ICD10CM | I13.11 | Hypertensive heart and chronic kidney disease without heart failure, with stage 5 chronic kidney disease, or end stage renal disease |
| ICD10CM | I13.2 | Hypertensive heart and chronic kidney disease with heart failure and with stage 5 chronic kidney disease, or end stage renal disease |
| ICD10CM | N18 | Chronic kidney disease (CKD) |
| ICD10CM | N18.1 | Chronic kidney disease, stage 1 |
| ICD10CM | N18.2 | Chronic kidney disease, stage 2 (mild) |
| ICD10CM | N18.3 | Chronic kidney disease, stage 3 (moderate) |
| ICD10CM | N18.4 | Chronic kidney disease, stage 4 (severe) |
| ICD10CM | N18.5 | Chronic kidney disease, stage 5 |
| ICD10CM | N18.6 | End stage renal disease |
| ICD10CM | N18.9 | Chronic kidney disease, unspecified |
| ICD10CM | O10.211 | Pre-existing hypertensive chronic kidney disease complicating pregnancy, first trimester |
| ICD10CM | O10.212 | Pre-existing hypertensive chronic kidney disease complicating pregnancy, second trimester |
| ICD10CM | O10.213 | Pre-existing hypertensive chronic kidney disease complicating pregnancy, third trimester |
| ICD10CM | O10.219 | Pre-existing hypertensive chronic kidney disease complicating pregnancy, unspecified trimester |
| ICD10CM | O10.22 | Pre-existing hypertensive chronic kidney disease complicating childbirth |
| ICD10CM | O10.31 | Pre-existing hypertensive heart and chronic kidney disease complicating pregnancy |
| ICD10CM | O10.311 | Pre-existing hypertensive heart and chronic kidney disease complicating pregnancy, first trimester |

Continued on next page

**Table14 – continued from previous page**

| CodeSystem | ConceptCode | ConceptName |
|---|---|---|
| ICD10CM | O10.312 | Pre-existing hypertensive heart and chronic kidney disease complicating pregnancy, second trimester |
| ICD10CM | O10.313 | Pre-existing hypertensive heart and chronic kidney disease complicating pregnancy, third trimester |
| ICD10CM | O10.319 | Pre-existing hypertensive heart and chronic kidney disease complicating pregnancy, unspecified trimester |
| ICD10CM | O10.32 | Pre-existing hypertensive heart and chronic kidney disease complicating childbirth |
| ICD10CM | O10.33 | Pre-existing hypertensive heart and chronic kidney disease complicating the puerperium |
| ICD10CM | Q61.2 | Polycystic kidney, adult type |
| ICD10CM | Q61.3 | Polycystic kidney, unspecified |
| ICD10CM | Q61.8 | Other cystic kidney diseases |
| ICD10CM | N18.30 | Chronic kidney disease, stage 3 unspecified |
| ICD10CM | N18.31 | Chronic kidney disease, stage 3a |
| ICD10CM | N18.32 | Chronic kidney disease, stage 3b |
| ICD10CM | Z94.0 | Kidney transplant status |
| SNOMED CT | 709044004 | Chronic kidney disease |
| SNOMED CT | 1208934006 | Sagliker syndrome |
| SNOMED CT | 90688005 | Chronic renal failure |
| SNOMED CT | 711000119100 | Chronic kidney disease stage 5 due to type 2 diabetes mellitus |
| SNOMED CT | 721000119107 | Chronic kidney disease stage 4 due to type 2 diabetes mellitus |
| SNOMED CT | 731000119105 | Chronic kidney disease stage 3 due to type 2 diabetes mellitus |
| SNOMED CT | 741000119101 | Chronic kidney disease stage 2 due to type 2 diabetes mellitus |
| SNOMED CT | 751000119104 | Chronic kidney disease stage 1 due to type 2 diabetes mellitus |
| SNOMED CT | 771000119108 | Chronic kidney disease due to type 2 diabetes mellitus |
| SNOMED CT | 1801000119106 | Anemia, pre-end stage renal disease on erythropoietin protocol |
| SNOMED CT | 8501000119104 | Hypertensive heart and chronic kidney disease |
| SNOMED CT | 71421000119105 | Hypertension in chronic kidney disease due to type 2 diabetes mellitus |
| SNOMED CT | 71701000119105 | Hypertension in chronic kidney disease due to type 1 diabetes mellitus |
| SNOMED CT | 90721000119101 | Chronic kidney disease stage 1 due to type 1 diabetes mellitus |
| SNOMED CT | 90731000119103 | Chronic kidney disease stage 2 due to type 1 diabetes mellitus |

Continued on next page

**Table14 – continued from previous page**

| CodeSystem | ConceptCode | ConceptName |
|---|---|---|
| SNOMED CT | 90741000119107 | Chronic kidney disease stage 3 due to type 1 diabetes mellitus |
| SNOMED CT | 90751000119109 | Chronic kidney disease stage 4 due to type 1 diabetes mellitus |
| SNOMED CT | 90761000119106 | Chronic kidney disease stage 5 due to type 1 diabetes mellitus |
| SNOMED CT | 90771000119100 | End stage renal disease on dialysis due to type 1 diabetes mellitus |
| SNOMED CT | 90791000119104 | End stage renal disease on dialysis due to type 2 diabetes mellitus |
| SNOMED CT | 96441000119101 | Chronic kidney disease due to type 1 diabetes mellitus |
| SNOMED CT | 96701000119107 | Hypertensive heart AND chronic kidney disease on dialysis |
| SNOMED CT | 96711000119105 | Hypertensive heart AND chronic kidney disease stage 5 |
| SNOMED CT | 96721000119103 | Hypertensive heart AND chronic kidney disease stage 4 |
| SNOMED CT | 96731000119100 | Hypertensive heart AND chronic kidney disease stage 3 |
| SNOMED CT | 96741000119109 | Hypertensive heart AND chronic kidney disease stage 2 |
| SNOMED CT | 96751000119106 | Hypertensive heart AND chronic kidney disease stage 1 |
| SNOMED CT | 104931000119100 | Chronic kidney disease due to hypertension |
| SNOMED CT | 111411000119103 | End stage renal disease due to hypertension |
| SNOMED CT | 117681000119102 | Chronic kidney disease stage 1 due to hypertension |
| SNOMED CT | 118781000119108 | Pre-existing hypertensive chronic kidney disease in mother complicating pregnancy |
| SNOMED CT | 120261000119101 | Benign hypertensive heart disease and chronic renal disease stage 5 |
| SNOMED CT | 127991000119101 | Hypertension concurrent and due to end stage renal disease on dialysis due to type 2 diabetes mellitus |
| SNOMED CT | 128001000119105 | Hypertension concurrent and due to end stage renal disease on dialysis due to type 1 diabetes mellitus |
| SNOMED CT | 129151000119102 | Chronic kidney disease stage 4 due to hypertension |
| SNOMED CT | 129161000119100 | Chronic kidney disease stage 5 due to hypertension |
| SNOMED CT | 129171000119106 | Chronic kidney disease stage 3 due to hypertension |
| SNOMED CT | 129181000119109 | Chronic kidney disease stage 2 due to hypertension |
| SNOMED CT | 140101000119109 | Hypertension in chronic kidney disease stage 5 due to type 2 diabetes mellitus |
| SNOMED CT | 140111000119107 | Hypertension in chronic kidney disease stage 4 due to type 2 diabetes mellitus |
| SNOMED CT | 140121000119100 | Hypertension in chronic kidney disease stage 3 due to type 2 diabetes mellitus |
| SNOMED CT | 140131000119102 | Hypertension in chronic kidney disease stage 2 due to type 2 diabetes mellitus |
| SNOMED CT | 153851000119106 | Malignant hypertensive chronic kidney disease stage 5 |
| SNOMED CT | 153891000119101 | End stage renal disease on dialysis due to hypertension |
| SNOMED CT | 284961000119106 | Chronic kidney disease due to benign hypertension |

Continued on next page

**Table14 – continued from previous page**

| CodeSystem | ConceptCode | ConceptName |
|---|---|---|
| SNOMED CT | 284971000119100 | Chronic kidney disease stage 1 due to benign hypertension |
| SNOMED CT | 284981000119102 | Chronic kidney disease stage 2 due to benign hypertension |
| SNOMED CT | 284991000119104 | Chronic kidney disease stage 3 due to benign hypertension |
| SNOMED CT | 285001000119105 | Chronic kidney disease stage 4 due to benign hypertension |
| SNOMED CT | 285011000119108 | Chronic kidney disease stage 5 due to benign hypertension |
| SNOMED CT | 285041000119107 | Benign hypertensive heart disease and chronic renal disease stage 1 |
| SNOMED CT | 285061000119106 | Benign hypertensive heart disease and chronic renal disease stage 2 |
| SNOMED CT | 285081000119102 | Benign hypertensive heart disease and chronic renal disease stage 3 |
| SNOMED CT | 285101000119109 | Benign hypertensive heart disease and chronic renal disease stage 4 |
| SNOMED CT | 285831000119108 | Malignant hypertensive chronic kidney disease |
| SNOMED CT | 285841000119104 | Malignant hypertensive end stage renal disease |
| SNOMED CT | 285851000119102 | Malignant hypertensive chronic kidney disease stage 1 |
| SNOMED CT | 285861000119100 | Malignant hypertensive chronic kidney disease stage 2 |
| SNOMED CT | 285871000119106 | Malignant hypertensive chronic kidney disease stage 3 |
| SNOMED CT | 285881000119109 | Malignant hypertensive chronic kidney disease stage 4 |
| SNOMED CT | 285911000119109 | Malignant hypertensive heart disease and chronic renal disease stage 3 |
| SNOMED CT | 285921000119102 | Malignant hypertensive heart disease and chronic renal disease stage 4 |
| SNOMED CT | 286371000119107 | Malignant hypertensive end stage renal disease on dialysis |
| SNOMED CT | 434431000124103 | Hypertensive end stage renal disease |
| SNOMED CT | 449631000124102 | Benign hypertensive heart disease and chronic renal disease |
| SNOMED CT | 691421000119108 | Anemia co-occurrent and due to chronic kidney disease stage 3 |
| SNOMED CT | 10757401000119104 | Pre-existing hypertensive heart and chronic kidney disease in mother complicating childbirth |
| SNOMED CT | 10757481000119107 | Pre-existing hypertensive heart and chronic kidney disease in mother complicating pregnancy |
| SNOMED CT | 16726004 | Renal osteodystrophy |
| SNOMED CT | 700378005 | Chronic kidney disease stage 3A |
| SNOMED CT | 700379002 | Chronic kidney disease stage 3B |
| SNOMED CT | 704667004 | Hypertension concurrent and due to end stage renal disease on dialysis |
| SNOMED CT | 707324008 | Anemia in end stage renal disease |
| SNOMED CT | 708975004 | Mixed renal osteodystrophy |
| SNOMED CT | 712487000 | End stage renal disease due to benign hypertension |
| SNOMED CT | 713313000 | Chronic kidney disease mineral and bone disorder |
| SNOMED CT | 714152005 | Chronic kidney disease stage 5 on dialysis |
| SNOMED CT | 714153000 | Chronic kidney disease stage 5 with transplant |

Continued on next page

**Table14 – continued from previous page**

| CodeSystem | ConceptCode | ConceptName |
|---|---|---|
| SNOMED CT | 722098007 | Chronic kidney disease following donor nephrectomy |
| SNOMED CT | 722149000 | Chronic kidney disease due to and following excision of neoplasm of kidney |
| SNOMED CT | 722150000 | Chronic kidney disease due to systemic infection |
| SNOMED CT | 722467000 | Chronic kidney disease due to traumatic loss of kidney |
| SNOMED CT | 723190009 | Chronic renal insufficiency |
| SNOMED CT | 776416004 | Hyperuricemia, pulmonary hypertension, renal failure, alkalosis syndrome |
| SNOMED CT | 236433006 | Acute-on-chronic renal failure |
| SNOMED CT | 236434000 | End stage renal failure untreated by renal replacement therapy |
| SNOMED CT | 236435004 | End stage renal failure on dialysis |
| SNOMED CT | 236436003 | End stage renal failure with renal transplant |
| SNOMED CT | 236552002 | Adynamic bone disease |
| SNOMED CT | 368421000119108 | Chronic kidney disease stage 1 due to drug induced diabetes mellitus |
| SNOMED CT | 368431000119106 | Chronic kidney disease stage 2 due to drug induced diabetes mellitus |
| SNOMED CT | 368441000119102 | Chronic kidney disease stage 3 due to drug induced diabetes mellitus |
| SNOMED CT | 368451000119100 | Chronic kidney disease stage 4 due to drug induced diabetes mellitus |
| SNOMED CT | 368461000119103 | Chronic kidney disease stage 5 due to drug induced diabetes mellitus |
| SNOMED CT | 368471000119109 | End stage renal disease on dialysis due to drug induced diabetes mellitus |
| SNOMED CT | 897308007 | Chronic kidney disease with osteoporosis |
| SNOMED CT | 897310009 | Renal osteodystrophy with high bone turnover |
| SNOMED CT | 897311008 | Renal osteodystrophy with normal bone turnover |
| SNOMED CT | 897312001 | Renal osteodystrophy with low bone turnover |
| SNOMED CT | 46177005 | End-stage renal disease |
| SNOMED CT | 49708008 | Anemia of chronic renal failure |
| SNOMED CT | 57557005 | Chronic milk alkali syndrome |
| SNOMED CT | 425369003 | Chronic progressive renal failure |
| SNOMED CT | 431855005 | Chronic kidney disease stage 1 |
| SNOMED CT | 431856006 | Chronic kidney disease stage 2 |
| SNOMED CT | 431857002 | Chronic kidney disease stage 4 |
| SNOMED CT | 433144002 | Chronic kidney disease stage 3 |
| SNOMED CT | 433146000 | Chronic kidney disease stage 5 |
| SNOMED CT | 1217070004 | Renal osteodystrophy due to hyperparathyroidism |

###### 8.4.5 COPD

Table 15: Concept codes used to identify if a patient was diagnosed with chronic-obstructive-pulmonary-disease.

| CodeSystem | ConceptCode | ConceptName |
|---|---|---|
| ICD10CM | J41 | Simple and mucopurulent chronic bronchitis |
| ICD10CM | J42 | Unspecified chronic bronchitis |
| ICD10CM | J43 | Emphysema |
| ICD10CM | J44.0 | Chronic obstructive pulmonary disease with (acute) lower respiratory infection |
| ICD10CM | J44.1 | Chronic obstructive pulmonary disease with (acute) exacerbation |
| ICD10CM | J44.9 | Chronic obstructive pulmonary disease, unspecified |
| ICD10CM | J41.0 | Simple chronic bronchitis |
| ICD10CM | J41.1 | Mucopurulent chronic bronchitis |
| ICD10CM | J41.8 | Mixed simple and mucopurulent chronic bronchitis |
| ICD10CM | J43.0 | Unilateral pulmonary emphysema [MacLeod's syndrome] |
| ICD10CM | J43.1 | Panlobular emphysema |
| ICD10CM | J43.2 | Centrilobular emphysema |
| ICD10CM | J43.8 | Other emphysema |
| ICD10CM | J43.9 | Emphysema, unspecified |
| SNOMED CT | 13645005 | Chronic obstructive lung disease |
| SNOMED CT | 86680006 | Ruptured emphysematous bleb of lung |
| SNOMED CT | 87433001 | Pulmonary emphysema |
| SNOMED CT | 135836000 | End stage chronic obstructive airways disease |
| SNOMED CT | 708030004 | Pulmonary emphysema co-occurrent with fibrosis of lung |
| SNOMED CT | 195951007 | Acute exacerbation of chronic obstructive airways disease |
| SNOMED CT | 196001008 | Chronic obstructive pulmonary disease with acute lower respiratory infection |
| SNOMED CT | 313296004 | Mild chronic obstructive pulmonary disease |
| SNOMED CT | 313297008 | Moderate chronic obstructive pulmonary disease |
| SNOMED CT | 313299006 | Severe chronic obstructive pulmonary disease |
| SNOMED CT | 285381006 | Acute infective exacerbation of chronic obstructive airways disease |
| SNOMED CT | 1177120001 | Bronchiolitis obliterans syndrome due to and following allogeneic stem cell transplant |
| SNOMED CT | 836477007 | Chronic emphysema due to vapor |
| SNOMED CT | 840350008 | Chronic obliterative bronchiolitis due to chemical fumes |
| SNOMED CT | 840351007 | Chronic obliterative bronchiolitis due to vapor |
| SNOMED CT | 1751000119100 | Acute exacerbation of chronic obstructive airways disease with asthma |
| SNOMED CT | 106001000119101 | Chronic obstructive lung disease co-occurrent with acute bronchitis |
| SNOMED CT | 10692761000119107 | Asthma-chronic obstructive pulmonary disease overlap syndrome |

Continued on next page

**Table15 – continued from previous page**

| CodeSystem | ConceptCode | ConceptName |
|---|---|---|
| SNOMED CT | 233674008 | Pulmonary emphysema in alpha-1 primary immunodeficiency deficiency |
| SNOMED CT | 762618008 | Bronchiolitis obliterans syndrome due to and after lung transplantation |
| SNOMED CT | 195957006 | Chronic bullous emphysema |
| SNOMED CT | 195958001 | Segmental bullous emphysema |
| SNOMED CT | 195959009 | Zonal bullous emphysema |
| SNOMED CT | 196026004 | Chronic emphysema due to chemical fumes |
| SNOMED CT | 4981000 | Panacinar emphysema |
| SNOMED CT | 16003001 | Giant bullous emphysema |
| SNOMED CT | 16846004 | Obstructive emphysema |
| SNOMED CT | 23958009 | Vanishing lung |
| SNOMED CT | 31898008 | Paraseptal emphysema |
| SNOMED CT | 33325001 | Compensatory emphysema |
| SNOMED CT | 233675009 | Toxic emphysema |
| SNOMED CT | 233677001 | Scar emphysema |
| SNOMED CT | 47895001 | Congenital emphysema |
| SNOMED CT | 47938003 | Chronic obliterative bronchiolitis |
| SNOMED CT | 57686001 | Emphysematous bleb of lung |
| SNOMED CT | 60805002 | Hemolytic anemia with emphysema AND cutis laxa |
| SNOMED CT | 66987001 | Congenital lobar emphysema |
| SNOMED CT | 68328006 | Centriacinar emphysema |
| SNOMED CT | 70756004 | Bronchial atresia with segmental pulmonary emphysema |
| SNOMED CT | 77690003 | Interstitial emphysema of lung |
| SNOMED CT | 266355005 | Bullous emphysema with collapse |
| SNOMED CT | 266356006 | Atrophic (senile) emphysema |
| SNOMED CT | 1010333003 | Emphysema of left lung |
| SNOMED CT | 1010334009 | Emphysema of right lung |

##### 8.4.6 Glaucoma

Table 16: Concept codes used to identify if a patient was diagnosed with glaucoma.

| CodeSystem | ConceptCode | ConceptName |
|---|---|---|
| ICD10CM | H40.10X0 | Unspecified open-angle glaucoma, stage unspecified |
| ICD10CM | H40.10X1 | Unspecified open-angle glaucoma, mild stage |
| ICD10CM | H40.10X2 | Unspecified open-angle glaucoma, moderate stage |
| ICD10CM | H40.10X3 | Unspecified open-angle glaucoma, severe stage |
| ICD10CM | H40.10X4 | Unspecified open-angle glaucoma, indeterminate stage |
| ICD10CM | H40.1110 | Primary open-angle glaucoma, right eye, stage unspecified |

Continued on next page

**Table16 – continued from previous page**

| CodeSystem | ConceptCode | ConceptName |
|---|---|---|
| ICD10CM | H40.1111 | Primary open-angle glaucoma, right eye, mild stage |
| ICD10CM | H40.1112 | Primary open-angle glaucoma, right eye, moderate stage |
| ICD10CM | H40.1113 | Primary open-angle glaucoma, right eye, severe stage |
| ICD10CM | H40.1114 | Primary open-angle glaucoma, right eye, indeterminate stage |
| ICD10CM | H40.1120 | Primary open-angle glaucoma, left eye, stage unspecified |
| ICD10CM | H40.1121 | Primary open-angle glaucoma, left eye, mild stage |
| ICD10CM | H40.1122 | Primary open-angle glaucoma, left eye, moderate stage |
| ICD10CM | H40.1123 | Primary open-angle glaucoma, left eye, severe stage |
| ICD10CM | H40.1124 | Primary open-angle glaucoma, left eye, indeterminate stage |
| ICD10CM | H40.1130 | Primary open-angle glaucoma, bilateral, stage unspecified |
| ICD10CM | H40.1131 | Primary open-angle glaucoma, bilateral, mild stage |
| ICD10CM | H40.1132 | Primary open-angle glaucoma, bilateral, moderate stage |
| ICD10CM | H40.1133 | Primary open-angle glaucoma, bilateral, severe stage |
| ICD10CM | H40.1134 | Primary open-angle glaucoma, bilateral, indeterminate stage |
| ICD10CM | H40.1210 | Low-tension glaucoma, right eye, stage unspecified |
| ICD10CM | H40.1211 | Low-tension glaucoma, right eye, mild stage |
| ICD10CM | H40.1212 | Low-tension glaucoma, right eye, moderate stage |
| ICD10CM | H40.1213 | Low-tension glaucoma, right eye, severe stage |
| ICD10CM | H40.1214 | Low-tension glaucoma, right eye, indeterminate stage |
| ICD10CM | H40.1220 | Low-tension glaucoma, left eye, stage unspecified |
| ICD10CM | H40.1221 | Low-tension glaucoma, left eye, mild stage |
| ICD10CM | H40.1222 | Low-tension glaucoma, left eye, moderate stage |
| ICD10CM | H40.1223 | Low-tension glaucoma, left eye, severe stage |
| ICD10CM | H40.1224 | Low-tension glaucoma, left eye, indeterminate stage |
| ICD10CM | H40.1230 | Low-tension glaucoma, bilateral, stage unspecified |
| ICD10CM | H40.1231 | Low-tension glaucoma, bilateral, mild stage |
| ICD10CM | H40.1232 | Low-tension glaucoma, bilateral, moderate stage |
| ICD10CM | H40.1233 | Low-tension glaucoma, bilateral, severe stage |
| ICD10CM | H40.1234 | Low-tension glaucoma, bilateral, indeterminate stage |
| ICD10CM | H40.1310 | Pigmentary glaucoma, right eye, stage unspecified |
| ICD10CM | H40.1311 | Pigmentary glaucoma, right eye, mild stage |
| ICD10CM | H40.1312 | Pigmentary glaucoma, right eye, moderate stage |
| ICD10CM | H40.1313 | Pigmentary glaucoma, right eye, severe stage |
| ICD10CM | H40.1314 | Pigmentary glaucoma, right eye, indeterminate stage |
| ICD10CM | H40.1320 | Pigmentary glaucoma, left eye, stage unspecified |
| ICD10CM | H40.1321 | Pigmentary glaucoma, left eye, mild stage |
| ICD10CM | H40.1322 | Pigmentary glaucoma, left eye, moderate stage |
| ICD10CM | H40.1323 | Pigmentary glaucoma, left eye, severe stage |
| ICD10CM | H40.1324 | Pigmentary glaucoma, left eye, indeterminate stage |
| ICD10CM | H40.1330 | Pigmentary glaucoma, bilateral, stage unspecified |
| ICD10CM | H40.1331 | Pigmentary glaucoma, bilateral, mild stage |

Continued on next page

**Table16 – continued from previous page**

| CodeSystem | ConceptCode | ConceptName |
|---|---|---|
| ICD10CM | H40.1332 | Pigmentary glaucoma, bilateral, moderate stage |
| ICD10CM | H40.1333 | Pigmentary glaucoma, bilateral, severe stage |
| ICD10CM | H40.1334 | Pigmentary glaucoma, bilateral, indeterminate stage |
| ICD10CM | H40.1410 | Capsular glaucoma with pseudoexfoliation of lens, right eye, stage unspecified |
| ICD10CM | H40.1411 | Capsular glaucoma with pseudoexfoliation of lens, right eye, mild stage |
| ICD10CM | H40.1412 | Capsular glaucoma with pseudoexfoliation of lens, right eye, moderate stage |
| ICD10CM | H40.1413 | Capsular glaucoma with pseudoexfoliation of lens, right eye, severe stage |
| ICD10CM | H40.1414 | Capsular glaucoma with pseudoexfoliation of lens, right eye, indeterminate stage |
| ICD10CM | H40.1420 | Capsular glaucoma with pseudoexfoliation of lens, left eye, stage unspecified |
| ICD10CM | H40.1421 | Capsular glaucoma with pseudoexfoliation of lens, left eye, mild stage |
| ICD10CM | H40.1422 | Capsular glaucoma with pseudoexfoliation of lens, left eye, moderate stage |
| ICD10CM | H40.1423 | Capsular glaucoma with pseudoexfoliation of lens, left eye, severe stage |
| ICD10CM | H40.1424 | Capsular glaucoma with pseudoexfoliation of lens, left eye, indeterminate stage |
| ICD10CM | H40.1430 | Capsular glaucoma with pseudoexfoliation of lens, bilateral, stage unspecified |
| ICD10CM | H40.1431 | Capsular glaucoma with pseudoexfoliation of lens, bilateral, mild stage |
| ICD10CM | H40.1432 | Capsular glaucoma with pseudoexfoliation of lens, bilateral, moderate stage |
| ICD10CM | H40.1433 | Capsular glaucoma with pseudoexfoliation of lens, bilateral, severe stage |
| ICD10CM | H40.1434 | Capsular glaucoma with pseudoexfoliation of lens, bilateral, indeterminate stage |
| ICD10CM | H40.151 | Residual stage of open-angle glaucoma, right eye |
| ICD10CM | H40.152 | Residual stage of open-angle glaucoma, left eye |
| ICD10CM | H40.153 | Residual stage of open-angle glaucoma, bilateral |
| ICD10CM | H40.20X0 | Unspecified primary angle-closure glaucoma, stage unspecified |
| ICD10CM | H40.20X1 | Unspecified primary angle-closure glaucoma, mild stage |
| ICD10CM | H40.20X2 | Unspecified primary angle-closure glaucoma, moderate stage |
| ICD10CM | H40.20X3 | Unspecified primary angle-closure glaucoma, severe stage |

Continued on next page

**Table16 – continued from previous page**

| CodeSystem | ConceptCode | ConceptName |
|---|---|---|
| ICD10CM | H40.20X4 | Unspecified primary angle-closure glaucoma, indeterminate stage |
| ICD10CM | H40.211 | Acute angle-closure glaucoma, right eye |
| ICD10CM | H40.212 | Acute angle-closure glaucoma, left eye |
| ICD10CM | H40.213 | Acute angle-closure glaucoma, bilateral |
| ICD10CM | H40.2210 | Chronic angle-closure glaucoma, right eye, stage unspecified |
| ICD10CM | H40.2211 | Chronic angle-closure glaucoma, right eye, mild stage |
| ICD10CM | H40.2212 | Chronic angle-closure glaucoma, right eye, moderate stage |
| ICD10CM | H40.2213 | Chronic angle-closure glaucoma, right eye, severe stage |
| ICD10CM | H40.2214 | Chronic angle-closure glaucoma, right eye, indeterminate stage |
| ICD10CM | H40.2220 | Chronic angle-closure glaucoma, left eye, stage unspecified |
| ICD10CM | H40.2221 | Chronic angle-closure glaucoma, left eye, mild stage |
| ICD10CM | H40.2222 | Chronic angle-closure glaucoma, left eye, moderate stage |
| ICD10CM | H40.2223 | Chronic angle-closure glaucoma, left eye, severe stage |
| ICD10CM | H40.2224 | Chronic angle-closure glaucoma, left eye, indeterminate stage |
| ICD10CM | H40.2230 | Chronic angle-closure glaucoma, bilateral, stage unspecified |
| ICD10CM | H40.2231 | Chronic angle-closure glaucoma, bilateral, mild stage |
| ICD10CM | H40.2232 | Chronic angle-closure glaucoma, bilateral, moderate stage |
| ICD10CM | H40.2233 | Chronic angle-closure glaucoma, bilateral, severe stage |
| ICD10CM | H40.2234 | Chronic angle-closure glaucoma, bilateral, indeterminate stage |
| ICD10CM | H40.231 | Intermittent angle-closure glaucoma, right eye |
| ICD10CM | H40.232 | Intermittent angle-closure glaucoma, left eye |
| ICD10CM | H40.233 | Intermittent angle-closure glaucoma, bilateral |
| ICD10CM | H40.241 | Residual stage of angle-closure glaucoma, right eye |
| ICD10CM | H40.242 | Residual stage of angle-closure glaucoma, left eye |
| ICD10CM | H40.243 | Residual stage of angle-closure glaucoma, bilateral |
| ICD10CM | H40.31X0 | Glaucoma secondary to eye trauma, right eye, stage unspecified |
| ICD10CM | H40.31X1 | Glaucoma secondary to eye trauma, right eye, mild stage |
| ICD10CM | H40.31X2 | Glaucoma secondary to eye trauma, right eye, moderate stage |
| ICD10CM | H40.31X3 | Glaucoma secondary to eye trauma, right eye, severe stage |
| ICD10CM | H40.31X4 | Glaucoma secondary to eye trauma, right eye, indeterminate stage |
| ICD10CM | H40.32X0 | Glaucoma secondary to eye trauma, left eye, stage unspecified |
| ICD10CM | H40.32X1 | Glaucoma secondary to eye trauma, left eye, mild stage |
| ICD10CM | H40.32X2 | Glaucoma secondary to eye trauma, left eye, moderate stage |
| ICD10CM | H40.32X3 | Glaucoma secondary to eye trauma, left eye, severe stage |

Continued on next page

**Table16 – continued from previous page**

| CodeSystem | ConceptCode | ConceptName |
|---|---|---|
| ICD10CM | H40.32X4 | Glaucoma secondary to eye trauma, left eye, indeterminate stage |
| ICD10CM | H40.33X0 | Glaucoma secondary to eye trauma, bilateral, stage unspecified |
| ICD10CM | H40.33X1 | Glaucoma secondary to eye trauma, bilateral, mild stage |
| ICD10CM | H40.33X2 | Glaucoma secondary to eye trauma, bilateral, moderate stage |
| ICD10CM | H40.33X3 | Glaucoma secondary to eye trauma, bilateral, severe stage |
| ICD10CM | H40.33X4 | Glaucoma secondary to eye trauma, bilateral, indeterminate stage |
| ICD10CM | H40.41X0 | Glaucoma secondary to eye inflammation, right eye, stage unspecified |
| ICD10CM | H40.41X1 | Glaucoma secondary to eye inflammation, right eye, mild stage |
| ICD10CM | H40.41X2 | Glaucoma secondary to eye inflammation, right eye, moderate stage |
| ICD10CM | H40.41X3 | Glaucoma secondary to eye inflammation, right eye, severe stage |
| ICD10CM | H40.41X4 | Glaucoma secondary to eye inflammation, right eye, indeterminate stage |
| ICD10CM | H40.42X0 | Glaucoma secondary to eye inflammation, left eye, stage unspecified |
| ICD10CM | H40.42X1 | Glaucoma secondary to eye inflammation, left eye, mild stage |
| ICD10CM | H40.42X2 | Glaucoma secondary to eye inflammation, left eye, moderate stage |
| ICD10CM | H40.42X3 | Glaucoma secondary to eye inflammation, left eye, severe stage |
| ICD10CM | H40.42X4 | Glaucoma secondary to eye inflammation, left eye, indeterminate stage |
| ICD10CM | H40.43X0 | Glaucoma secondary to eye inflammation, bilateral, stage unspecified |
| ICD10CM | H40.43X1 | Glaucoma secondary to eye inflammation, bilateral, mild stage |
| ICD10CM | H40.43X2 | Glaucoma secondary to eye inflammation, bilateral, moderate stage |
| ICD10CM | H40.43X3 | Glaucoma secondary to eye inflammation, bilateral, severe stage |
| ICD10CM | H40.43X4 | Glaucoma secondary to eye inflammation, bilateral, indeterminate stage |
| ICD10CM | H40.51X0 | Glaucoma secondary to other eye disorders, right eye, stage unspecified |

Continued on next page

**Table16 – continued from previous page**

| CodeSystem | ConceptCode | ConceptName |
|---|---|---|
| ICD10CM | H40.51X1 | Glaucoma secondary to other eye disorders, right eye, mild stage |
| ICD10CM | H40.51X2 | Glaucoma secondary to other eye disorders, right eye, moderate stage |
| ICD10CM | H40.51X3 | Glaucoma secondary to other eye disorders, right eye, severe stage |
| ICD10CM | H40.51X4 | Glaucoma secondary to other eye disorders, right eye, indeterminate stage |
| ICD10CM | H40.52X0 | Glaucoma secondary to other eye disorders, left eye, stage unspecified |
| ICD10CM | H40.52X1 | Glaucoma secondary to other eye disorders, left eye, mild stage |
| ICD10CM | H40.52X2 | Glaucoma secondary to other eye disorders, left eye, moderate stage |
| ICD10CM | H40.52X3 | Glaucoma secondary to other eye disorders, left eye, severe stage |
| ICD10CM | H40.52X4 | Glaucoma secondary to other eye disorders, left eye, indeterminate stage |
| ICD10CM | H40.53X0 | Glaucoma secondary to other eye disorders, bilateral, stage unspecified |
| ICD10CM | H40.53X1 | Glaucoma secondary to other eye disorders, bilateral, mild stage |
| ICD10CM | H40.53X2 | Glaucoma secondary to other eye disorders, bilateral, moderate stage |
| ICD10CM | H40.53X3 | Glaucoma secondary to other eye disorders, bilateral, severe stage |
| ICD10CM | H40.53X4 | Glaucoma secondary to other eye disorders, bilateral, indeterminate stage |
| ICD10CM | H40.61X0 | Glaucoma secondary to drugs, right eye, stage unspecified |
| ICD10CM | H40.61X1 | Glaucoma secondary to drugs, right eye, mild stage |
| ICD10CM | H40.61X2 | Glaucoma secondary to drugs, right eye, moderate stage |
| ICD10CM | H40.61X3 | Glaucoma secondary to drugs, right eye, severe stage |
| ICD10CM | H40.61X4 | Glaucoma secondary to drugs, right eye, indeterminate stage |
| ICD10CM | H40.62X0 | Glaucoma secondary to drugs, left eye, stage unspecified |
| ICD10CM | H40.62X1 | Glaucoma secondary to drugs, left eye, mild stage |
| ICD10CM | H40.62X2 | Glaucoma secondary to drugs, left eye, moderate stage |
| ICD10CM | H40.62X3 | Glaucoma secondary to drugs, left eye, severe stage |
| ICD10CM | H40.62X4 | Glaucoma secondary to drugs, left eye, indeterminate stage |
| ICD10CM | H40.63X0 | Glaucoma secondary to drugs, bilateral, stage unspecified |
| ICD10CM | H40.63X1 | Glaucoma secondary to drugs, bilateral, mild stage |
| ICD10CM | H40.63X2 | Glaucoma secondary to drugs, bilateral, moderate stage |
| ICD10CM | H40.63X3 | Glaucoma secondary to drugs, bilateral, severe stage |

Continued on next page

**Table16 – continued from previous page**

| CodeSystem | ConceptCode | ConceptName |
|---|---|---|
| ICD10CM | H40.63X4 | Glaucoma secondary to drugs, bilateral, indeterminate stage |
| ICD10CM | H40.811 | Glaucoma with increased episcleral venous pressure, right eye |
| ICD10CM | H40.812 | Glaucoma with increased episcleral venous pressure, left eye |
| ICD10CM | H40.813 | Glaucoma with increased episcleral venous pressure, bilateral |
| ICD10CM | H40.821 | Hypersecretion glaucoma, right eye |
| ICD10CM | H40.822 | Hypersecretion glaucoma, left eye |
| ICD10CM | H40.823 | Hypersecretion glaucoma, bilateral |
| ICD10CM | H40.831 | Aqueous misdirection, right eye |
| ICD10CM | H40.832 | Aqueous misdirection, left eye |
| ICD10CM | H40.833 | Aqueous misdirection, bilateral |
| ICD10CM | H40.89 | Other specified glaucoma |
| ICD10CM | Q15.0 | Congenital glaucoma |
| SNOMED CT | 1654001 | Corticosteroid-induced glaucoma |
| SNOMED CT | 15374009 | Aphakic glaucoma |
| SNOMED CT | 19144002 | Absolute glaucoma |
| SNOMED CT | 19309007 | Glaucoma associated with vascular disorder |
| SNOMED CT | 21571006 | Secondary angle-closure glaucoma |
| SNOMED CT | 21928008 | Secondary open-angle glaucoma |
| SNOMED CT | 23986001 | Glaucoma |
| SNOMED CT | 27735002 | Glaucoma associated with tumors AND/OR cysts |
| SNOMED CT | 29369005 | Hypersecretion glaucoma |
| SNOMED CT | 30041005 | Acute angle-closure glaucoma |
| SNOMED CT | 32893002 | Phacolytic glaucoma |
| SNOMED CT | 33647009 | Chronic angle-closure glaucoma |
| SNOMED CT | 84333006 | Phacogenic glaucoma |
| SNOMED CT | 84494001 | Open-angle glaucoma |
| SNOMED CT | 89215000 | Postoperative angle-closure glaucoma |
| SNOMED CT | 92829008 | Glaucoma due to combination of mechanisms |
| SNOMED CT | 95213001 | Primary glaucoma due to combination of mechanisms |
| SNOMED CT | 95250000 | Secondary glaucoma due to combination mechanisms |
| SNOMED CT | 95717004 | Secondary glaucoma |
| SNOMED CT | 111513000 | Advanced open-angle glaucoma |
| SNOMED CT | 111514006 | Pseudoexfoliation glaucoma |
| SNOMED CT | 787051000 | Open-angle glaucoma of left eye |
| SNOMED CT | 787052007 | Open-angle glaucoma of right eye |
| SNOMED CT | 24151000119106 | Steroid-induced open angle glaucoma |
| SNOMED CT | 41911000119107 | Glaucoma due to type 2 diabetes mellitus |
| SNOMED CT | 60981000119103 | Glaucoma due to diabetes mellitus type 1 |
| SNOMED CT | 336611000119109 | Acute angle-closure glaucoma of right eye |
| SNOMED CT | 336631000119104 | Absolute glaucoma right eye |

Continued on next page

**Table16 – continued from previous page**

| CodeSystem | ConceptCode | ConceptName |
|---|---|---|
| SNOMED CT | 342221000119104 | Acute angle-closure glaucoma of left eye |
| SNOMED CT | 342241000119105 | Absolute glaucoma left eye |
| SNOMED CT | 347381000119106 | Bilateral acute angle-closure glaucoma |
| SNOMED CT | 347401000119106 | Bilateral absolute glaucoma |
| SNOMED CT | 12239301000119102 | Bilateral open-angle glaucoma |
| SNOMED CT | 12239421000119101 | Bilateral glaucoma |
| SNOMED CT | 12239461000119106 | Glaucoma of left eye |
| SNOMED CT | 12239501000119106 | Glaucoma of right eye |
| SNOMED CT | 15633281000119103 | Bilateral primary open angle glaucoma |
| SNOMED CT | 15633321000119108 | Primary open angle glaucoma of left eye |
| SNOMED CT | 15640441000119104 | Primary open angle glaucoma of right eye |
| SNOMED CT | 15673001000119103 | Congenital glaucoma of bilateral eyes |
| SNOMED CT | 15679801000119105 | Bilateral uveitis-glaucoma-hyphema syndrome of eyes |
| SNOMED CT | 15736441000119108 | Primary angle-closure glaucoma of bilateral eyes |
| SNOMED CT | 15736481000119103 | Primary angle-closure glaucoma of right eye |
| SNOMED CT | 15736521000119103 | Primary angle-closure glaucoma of left eye |
| SNOMED CT | 15736561000119108 | Neovascular glaucoma of bilateral eyes |
| SNOMED CT | 15736601000119108 | Neovascular glaucoma of right eye |
| SNOMED CT | 15736641000119105 | Neovascular glaucoma of left eye |
| SNOMED CT | 15736681000119100 | Narrow-angle glaucoma of right eye |
| SNOMED CT | 15736721000119106 | Bilateral angle-closure glaucoma |
| SNOMED CT | 15736761000119101 | Narrow angle glaucoma of left eye |
| SNOMED CT | 15738841000119105 | Glaucoma of bilateral eyes due to combination of mechanisms |
| SNOMED CT | 15738881000119100 | Glaucoma of left eye due to combination of mechanisms |
| SNOMED CT | 15738921000119107 | Glaucoma of right eye due to combination of mechanisms |
| SNOMED CT | 15739041000119106 | Glaucoma of bilateral eyes due to iris anomaly |
| SNOMED CT | 15739081000119101 | Glaucoma of bilateral eyes due to anterior segment anomaly |
| SNOMED CT | 15739121000119104 | Glaucoma of right eye due to anterior segment anomaly |
| SNOMED CT | 15739161000119109 | Glaucoma of left eye due to anterior segment anomaly |
| SNOMED CT | 15739201000119104 | Glaucoma of left eye due to ocular vascular disorder |
| SNOMED CT | 15739241000119102 | Glaucoma of right eye due to ocular vascular disorder |
| SNOMED CT | 15739281000119107 | Glaucoma of bilateral eyes due to ocular vascular disorder |
| SNOMED CT | 15739321000119102 | Glaucoma of right eye due to chamber angle anomaly |
| SNOMED CT | 15739361000119107 | Glaucoma of bilateral eyes due to chamber angle anomaly |
| SNOMED CT | 15739401000119103 | Glaucoma of left eye due to chamber angle anomaly |
| SNOMED CT | 15739441000119101 | Glaucoma of right eye due to trauma |
| SNOMED CT | 15739481000119106 | Glaucoma of left eye due to trauma |
| SNOMED CT | 15739561000119101 | Glaucoma of right eye due to lens disorder |
| SNOMED CT | 15739641000119104 | Glaucoma of left eye due to lens disorder |
| SNOMED CT | 15739681000119109 | Glaucoma of right eye due to systemic disorder |
| SNOMED CT | 15739721000119103 | Glaucoma of left eye due to systemic disorder |
| SNOMED CT | 15739761000119108 | Glaucoma of bilateral eyes due to systemic disorder |

Continued on next page

**Table16 – continued from previous page**

| CodeSystem | ConceptCode | ConceptName |
|---|---|---|
| SNOMED CT | 15993671000119108 | Open-angle glaucoma of right eye caused by steroid |
| SNOMED CT | 15993711000119107 | Open-angle glaucoma of left eye caused by steroid |
| SNOMED CT | 15993751000119108 | Open-angle glaucoma of bilateral eyes caused by steroid |
| SNOMED CT | 15996831000119101 | Glaucoma caused by Onchocerca volvulus |
| SNOMED CT | 444863008 | Anatomical narrow angle glaucoma with borderline intraocular pressure |
| SNOMED CT | 698840003 | Neovascular glaucoma due to hyphema |
| SNOMED CT | 713457002 | Neovascular glaucoma due to diabetes mellitus |
| SNOMED CT | 715144004 | Glaucoma caused by silicone oil |
| SNOMED CT | 716166002 | Microcornea with glaucoma and absent frontal sinus syndrome |
| SNOMED CT | 721898008 | Corticosteroid-induced glaucoma of bilateral eyes |
| SNOMED CT | 722321001 | Corticosteroid-induced glaucoma of left eye |
| SNOMED CT | 722329004 | Corticosteroid-induced glaucoma of right eye |
| SNOMED CT | 193548006 | Steroid-induced glaucoma glaucomatous stage |
| SNOMED CT | 193552006 | Glaucoma due to chamber angle anomaly |
| SNOMED CT | 193553001 | Glaucoma due to iris anomaly |
| SNOMED CT | 193555008 | Glaucoma due to systemic syndrome |
| SNOMED CT | 193561006 | Secondary angle-closure glaucoma with pupillary block |
| SNOMED CT | 193562004 | Glaucoma due to ocular vascular disorder |
| SNOMED CT | 204113001 | Congenital glaucoma |
| SNOMED CT | 206248004 | Traumatic glaucoma due to birth trauma |
| SNOMED CT | 314017009 | Acute-on-chronic glaucoma |
| SNOMED CT | 314033007 | Uveitic glaucoma |
| SNOMED CT | 314784002 | Secondary angle-closure glaucoma - synechial |
| SNOMED CT | 370504007 | Aqueous humor misdirect |
| SNOMED CT | 392030001 | Hemolytic glaucoma |
| SNOMED CT | 392288006 | Primary angle-closure glaucoma |
| SNOMED CT | 392291006 | Angle-closure glaucoma |
| SNOMED CT | 392300000 | Phacomorphic glaucoma |
| SNOMED CT | 392352004 | Angle recession glaucoma |
| SNOMED CT | 232081005 | Iatrogenic glaucoma |
| SNOMED CT | 232082003 | Iatrogenic angle-closure glaucoma |
| SNOMED CT | 232083008 | Glaucoma and corneal anomaly |
| SNOMED CT | 232086000 | Neovascular glaucoma |
| SNOMED CT | 232087009 | Glaucoma with intraocular hemorrhage |
| SNOMED CT | 232088004 | Ghost cell glaucoma |
| SNOMED CT | 232090003 | Glaucoma following surgery |
| SNOMED CT | 34623005 | Glaucoma with increased episcleral venous pressure |
| SNOMED CT | 37155002 | Glaucoma associated with ocular inflammation |
| SNOMED CT | 45623002 | Glaucoma associated with anterior segment anomaly |
| SNOMED CT | 46168003 | Pigmentary glaucoma |
| SNOMED CT | 50485007 | Low tension glaucoma |

Continued on next page

**Table16 – continued from previous page**

| CodeSystem | ConceptCode | ConceptName |
|---|---|---|
| SNOMED CT | 53667005 | Glaucoma associated with systemic syndromes |
| SNOMED CT | 65460003 | Intermittent angle-closure glaucoma |
| SNOMED CT | 66725002 | Glaucoma due to perforating injury |
| SNOMED CT | 66747002 | Glaucoma associated with ocular disorder |
| SNOMED CT | 68241007 | Glaucoma associated with ocular trauma |
| SNOMED CT | 77075001 | Primary open angle glaucoma |
| SNOMED CT | 404648005 | Lens particle glaucoma |
| SNOMED CT | 415176004 | Primary congenital glaucoma |
| SNOMED CT | 267625001 | Glaucoma due to ocular tumor or cyst |
| SNOMED CT | 275477002 | Glaucoma due to ocular cyst |
| SNOMED CT | 1207009 | Glaucomatous atrophy of optic disc |
| SNOMED CT | 29538005 | Glaucomatocyclitic crisis |
| SNOMED CT | 718851007 | Cataract glaucoma syndrome |
| SNOMED CT | 719976001 | Glaucoma and sleep apnea syndrome |
| SNOMED CT | 721974000 | Lowry MacLean syndrome |
| SNOMED CT | 722450007 | GEMSS syndrome |
| SNOMED CT | 723503006 | Retinal degeneration, nanophthalmos, glaucoma syndrome |
| SNOMED CT | 733086003 | Pseudoprogeria syndrome |
| SNOMED CT | 733116005 | Aniridia, renal agenesis, psychomotor retardation syndrome |
| SNOMED CT | 733455003 | Spastic paraplegia, glaucoma, intellectual disability syndrome |
| SNOMED CT | 736838001 | Cupping of optic disc co-occurrent and due to open angle glaucoma |
| SNOMED CT | 737006004 | Cupping of optic discs of bilateral eyes co-occurrent and due to open-angle glaucoma |
| SNOMED CT | 770776002 | Borderline angle-closure glaucoma of right eye |
| SNOMED CT | 770777006 | Borderline angle-closure glaucoma of left eye |
| SNOMED CT | 783246000 | Megalocornea, spherophakia, secondary glaucoma syndrome |
| SNOMED CT | 193531003 | Borderline glaucoma |
| SNOMED CT | 193533000 | Open-angle glaucoma - borderline |
| SNOMED CT | 193534006 | Angle-closure glaucoma - borderline |
| SNOMED CT | 193556009 | Glaucoma in endocrine, nutritional and metabolic diseases |
| SNOMED CT | 1231271005 | Glaucoma due to and following retinopathy of prematurity |
| SNOMED CT | 1231270006 | Glaucoma following ocular onchocerciasis |
| SNOMED CT | 334961000119105 | Right secondary angle-closure glaucoma due to pupil block |
| SNOMED CT | 349231000119108 | Bilateral aqueous humor misdirection of eyes |
| SNOMED CT | 1231243004 | High pressure primary open-angle glaucoma |
| SNOMED CT | 1137633000 | Secondary angle-closure glaucoma due to iridocorneal endothelial syndrome |
| SNOMED CT | 788945006 | Glaucoma caused by contact lens |
| SNOMED CT | 788946007 | Glaucoma due to eye inflammation |
| SNOMED CT | 788947003 | Glaucoma due to intraocular neoplasm |

Continued on next page

**Table16 – continued from previous page**

| CodeSystem | ConceptCode | ConceptName |
|---|---|---|
| SNOMED CT | 788948008 | Glaucoma due to retinal detachment |
| SNOMED CT | 788995007 | Normal pressure primary open-angle glaucoma |
| SNOMED CT | 833278008 | Glaucoma suspect |
| SNOMED CT | 860798008 | Glaucoma due to diabetes mellitus |
| SNOMED CT | 331471000119105 | Uveitic glaucoma of right eye |
| SNOMED CT | 332871000119103 | Pseudoexfoliation glaucoma of right eye |
| SNOMED CT | 333111000119107 | Pigmentary glaucoma of right eye |
| SNOMED CT | 333151000119108 | Phacolytic glaucoma of right eye |
| SNOMED CT | 334321000119101 | Low tension glaucoma of right eye |
| SNOMED CT | 336031000119105 | Anatomic narrow angle glaucoma of right eye |
| SNOMED CT | 337081000119109 | Uveitic glaucoma of left eye |
| SNOMED CT | 338481000119100 | Pseudoexfoliation glaucoma of left eye |
| SNOMED CT | 338721000119108 | Pigmentary glaucoma of left eye |
| SNOMED CT | 338761000119103 | Phacolytic glaucoma of left eye |
| SNOMED CT | 339921000119104 | Low tension glaucoma of left eye |
| SNOMED CT | 341641000119100 | Anatomic narrow angle glaucoma of left eye |
| SNOMED CT | 343161000119109 | Bilateral uveitic glaucoma of eyes |
| SNOMED CT | 344251000119103 | Bilateral pseudoexfoliation glaucoma of eyes |
| SNOMED CT | 344491000119107 | Bilateral pigmentary glaucoma of eyes |
| SNOMED CT | 345291000119109 | Bilateral low tension glaucoma of eyes |
| SNOMED CT | 346861000119107 | Bilateral eye anatomic narrow angle glaucoma |
| SNOMED CT | 348361000119103 | Aphakic glaucoma of right eye |
| SNOMED CT | 348811000119108 | Aphakic glaucoma of left eye |
| SNOMED CT | 349311000119106 | Bilateral aphakic glaucoma |
| SNOMED CT | 678401000119102 | Bilateral glaucoma of eyes caused by drug |
| SNOMED CT | 678411000119104 | Glaucoma of left eye caused by drug |
| SNOMED CT | 678421000119106 | Glaucoma of right eye caused by drug |
| SNOMED CT | 678471000119107 | Aqueous humor misdirection of left eye |
| SNOMED CT | 678481000119105 | Aqueous humor misdirection of right eye |
| SNOMED CT | 15673041000119101 | Congenital glaucoma of right eye |
| SNOMED CT | 15673081000119106 | Congenital glaucoma of left eye |
| SNOMED CT | 15679721000119107 | Uveitis-hyphema-glaucoma syndrome of right eye |
| SNOMED CT | 15679761000119102 | Uveitis-hyphema-glaucoma syndrome of left eye |
| SNOMED CT | 15738761000119100 | Bilateral eye borderline angle-closure glaucoma |
| SNOMED CT | 15739801000119100 | Cupping of left optic disc due to open-angle glaucoma of left eye |
| SNOMED CT | 15739881000119108 | Cupping of right optic disc due to open-angle glaucoma of right eye |
| SNOMED CT | 392029006 | Uveitis-glaucoma-hyphema syndrome |
| SNOMED CT | 340571000119101 | Left secondary angle-closure glaucoma due to pupil block |
| SNOMED CT | 232080006 | Plateau iris |
| SNOMED CT | 35472004 | Open angle with borderline intraocular pressure |
| SNOMED CT | 71111008 | Glaucoma of childhood |

Continued on next page

**Table16 – continued from previous page**

| CodeSystem | ConceptCode | ConceptName |
|---|---|---|
| SNOMED CT | 404634005 | Glaucoma associated with iridocorneal endothelial syndrome |
| SNOMED CT | 415501002 | Schwartz ocular syndrome |
| SNOMED CT | 302895007 | Steroid-induced glaucoma - borderline |
| SNOMED CT | 93435005 | Drug-induced glaucoma |
| SNOMED CT | 1003425007 | Glaucoma due to Lowe syndrome |
| SNOMED CT | 1003428009 | Glaucoma due to congenital anomaly of eye |
| SNOMED CT | 1003657004 | Congenital glaucoma of left eye |
| SNOMED CT | 15995551000119104 | Bilateral glaucoma of eyes due to plateau iris |
| SNOMED CT | 334761000119107 | Intermittent angle-closure glaucoma of right eye |
| SNOMED CT | 340361000119101 | Intermittent angle-closure glaucoma of left eye |
| SNOMED CT | 345701000119106 | Bilateral intermittent angle-closure glaucoma |
| SNOMED CT | 678431000119109 | Bilateral glaucoma of eyes due to increased episcleral venous pressure |
| SNOMED CT | 678441000119100 | Glaucoma of left eye due to increased episcleral venous pressure |
| SNOMED CT | 678451000119103 | Glaucoma of right eye due to increased episcleral venous pressure |
| SNOMED CT | 890423003 | Secondary childhood glaucoma following congenital cataract surgery |
| SNOMED CT | 897585008 | Bilateral primary congenital glaucoma |
| SNOMED CT | 345881000119107 | Bilateral secondary angle-closure glaucoma due to pupil block |
| SNOMED CT | 1196922005 | Neovascular glaucoma due to diabetes mellitus type 2 |
| SNOMED CT | 1196923000 | Neovascular glaucoma due to diabetes mellitus type 1 |

###### 8.4.7 Heart Failure

Table 17: Concept codes used to identify if a patient was diagnosed with heart-failure.

| CodeSystem | ConceptCode | ConceptName |
|---|---|---|
| ICD10CM | I11.0 | Hypertensive heart disease with heart failure |
| ICD10CM | I13.0 | Hypertensive heart and chronic kidney disease with heart failure and stage 1 through stage 4 chronic kidney disease, or unspecified chronic kidney disease |
| ICD10CM | I13.2 | Hypertensive heart and chronic kidney disease with heart failure and with stage 5 chronic kidney disease, or end stage renal disease |
| ICD10CM | I50.1 | Left ventricular failure, unspecified |
| ICD10CM | I50.20 | Unspecified systolic (congestive) heart failure |
| ICD10CM | I50.21 | Acute systolic (congestive) heart failure |

Continued on next page

**Table17 – continued from previous page**

| CodeSystem | ConceptCode | ConceptName |
|---|---|---|
| ICD10CM | I50.22 | Chronic systolic (congestive) heart failure |
| ICD10CM | I50.23 | Acute on chronic systolic (congestive) heart failure |
| ICD10CM | I50.30 | Unspecified diastolic (congestive) heart failure |
| ICD10CM | I50.31 | Acute diastolic (congestive) heart failure |
| ICD10CM | I50.32 | Chronic diastolic (congestive) heart failure |
| ICD10CM | I50.33 | Acute on chronic diastolic (congestive) heart failure |
| ICD10CM | I50.40 | Unspecified combined systolic (congestive) and diastolic (congestive) heart failure |
| ICD10CM | I50.41 | Acute combined systolic (congestive) and diastolic (congestive) heart failure |
| ICD10CM | I50.42 | Chronic combined systolic (congestive) and diastolic (congestive) heart failure |
| ICD10CM | I50.43 | Acute on chronic combined systolic (congestive) and diastolic (congestive) heart failure |
| ICD10CM | I50.81 | Right heart failure |
| ICD10CM | I50.82 | Biventricular heart failure |
| ICD10CM | I50.83 | High output heart failure |
| ICD10CM | I50.84 | End stage heart failure |
| ICD10CM | I50.89 | Other heart failure |
| ICD10CM | I50.9 | Heart failure, unspecified |
| ICD10CM | I50.810 | Right heart failure, unspecified |
| ICD10CM | I50.811 | Acute right heart failure |
| ICD10CM | I50.812 | Chronic right heart failure |
| ICD10CM | I50.813 | Acute on chronic right heart failure |
| ICD10CM | I50.814 | Right heart failure due to left heart failure |
| SNOMED CT | 871617000 | Low output heart failure due to and following Fontan operation |
| SNOMED CT | 7411000175102 | Chronic heart failure co-occurrent with normal ejection fraction |
| SNOMED CT | 15781000119107 | Hypertensive heart AND chronic kidney disease with congestive heart failure |
| SNOMED CT | 23341000119109 | Congestive heart failure with right heart failure |
| SNOMED CT | 67431000119105 | Congestive heart failure stage D |
| SNOMED CT | 67441000119101 | Congestive heart failure stage C |
| SNOMED CT | 72481000119103 | Congestive heart failure as early postoperative complication |
| SNOMED CT | 101281000119107 | Congestive heart failure due to cardiomyopathy |
| SNOMED CT | 120851000119104 | Systolic heart failure stage D |
| SNOMED CT | 120861000119102 | Systolic heart failure stage C |
| SNOMED CT | 120871000119108 | Systolic heart failure stage B |
| SNOMED CT | 120881000119106 | Diastolic heart failure stage D |
| SNOMED CT | 120891000119109 | Diastolic heart failure stage C |
| SNOMED CT | 120901000119108 | Diastolic heart failure stage B |
| SNOMED CT | 153931000119109 | Acute combined systolic and diastolic heart failure |

Continued on next page

**Table17 – continued from previous page**

| CodeSystem | ConceptCode | ConceptName |
|---|---|---|
| SNOMED CT | 153941000119100 | Chronic combined systolic and diastolic heart failure |
| SNOMED CT | 153951000119103 | Acute on chronic combined systolic and diastolic heart failure |
| SNOMED CT | 15629541000119106 | Congestive heart failure stage C due to ischemic cardiomyopathy |
| SNOMED CT | 15629591000119103 | Congestive heart failure stage B due to ischaemic cardiomyopathy |
| SNOMED CT | 15629641000119107 | Systolic heart failure stage B due to ischemic cardiomyopathy |
| SNOMED CT | 15629741000119102 | Systolic heart failure stage C due to ischemic cardiomyopathy |
| SNOMED CT | 15964701000119109 | Saddle embolus of pulmonary artery with acute cor pulmonale |
| SNOMED CT | 42343007 | Congestive heart failure |
| SNOMED CT | 43736008 | Rheumatic left ventricular failure |
| SNOMED CT | 44088000 | Low cardiac output syndrome |
| SNOMED CT | 44313006 | Right heart failure secondary to left heart failure |
| SNOMED CT | 46113002 | Hypertensive heart failure |
| SNOMED CT | 48447003 | Chronic heart failure |
| SNOMED CT | 56675007 | Acute heart failure |
| SNOMED CT | 74960003 | Acute left-sided congestive heart failure |
| SNOMED CT | 410431009 | Cardiorespiratory failure |
| SNOMED CT | 417996009 | Systolic heart failure |
| SNOMED CT | 418304008 | Diastolic heart failure |
| SNOMED CT | 424404003 | Decompensated chronic heart failure |
| SNOMED CT | 426263006 | Congestive heart failure due to left ventricular systolic dysfunction |
| SNOMED CT | 426611007 | Congestive heart failure due to valvular disease |
| SNOMED CT | 441481004 | Chronic systolic heart failure |
| SNOMED CT | 441530006 | Chronic diastolic heart failure |
| SNOMED CT | 443253003 | Acute on chronic systolic heart failure |
| SNOMED CT | 443254009 | Acute systolic heart failure |
| SNOMED CT | 443343001 | Acute diastolic heart failure |
| SNOMED CT | 443344007 | Acute on chronic diastolic heart failure |
| SNOMED CT | 364006 | Acute left-sided heart failure |
| SNOMED CT | 5148006 | Hypertensive heart disease with congestive heart failure |
| SNOMED CT | 5375005 | Chronic left-sided congestive heart failure |
| SNOMED CT | 10091002 | High output heart failure |
| SNOMED CT | 10633002 | Acute congestive heart failure |
| SNOMED CT | 25544003 | Low output heart failure |
| SNOMED CT | 314206003 | Refractory heart failure |
| SNOMED CT | 233924009 | Heart failure as a complication of care |
| SNOMED CT | 82523003 | Congestive rheumatic heart failure |

Continued on next page

**Table17 – continued from previous page**

| CodeSystem | ConceptCode | ConceptName |
|---|---|---|
| SNOMED CT | 83105008 | Malignant hypertensive heart disease with congestive heart failure |
| SNOMED CT | 84114007 | Heart failure |
| SNOMED CT | 85232009 | Left heart failure |
| SNOMED CT | 88805009 | Chronic congestive heart failure |
| SNOMED CT | 90727007 | Pleural effusion due to congestive heart failure |
| SNOMED CT | 92506005 | Biventricular congestive heart failure |
| SNOMED CT | 111283005 | Chronic left-sided heart failure |
| SNOMED CT | 446221000 | Heart failure with normal ejection fraction |
| SNOMED CT | 471880001 | Heart failure due to end stage congenital heart disease |
| SNOMED CT | 698296002 | Acute exacerbation of chronic congestive heart failure |
| SNOMED CT | 698594003 | Symptomatic congestive heart failure |
| SNOMED CT | 703272007 | Heart failure with reduced ejection fraction |
| SNOMED CT | 703273002 | Heart failure with reduced ejection fraction due to coronary artery disease |
| SNOMED CT | 703274008 | Heart failure with reduced ejection fraction due to myocarditis |
| SNOMED CT | 703275009 | Heart failure with reduced ejection fraction due to cardiomyopathy |
| SNOMED CT | 703276005 | Heart failure with reduced ejection fraction due to heart valve disease |
| SNOMED CT | 717840005 | Congestive heart failure stage B |
| SNOMED CT | 194767001 | Benign hypertensive heart disease with congestive cardiac failure |
| SNOMED CT | 194779001 | Hypertensive heart and renal disease with (congestive) heart failure |
| SNOMED CT | 194781004 | Hypertensive heart and renal disease with both (congestive) heart failure and renal failure |
| SNOMED CT | 195111005 | Decompensated cardiac failure |
| SNOMED CT | 195112003 | Compensated cardiac failure |
| SNOMED CT | 195114002 | Acute left ventricular failure |
| SNOMED CT | 206586007 | Congenital cardiac failure |
| SNOMED CT | 788950000 | Heart failure with mid range ejection fraction |
| SNOMED CT | 7371000175103 | Reduced ejection fraction co-occurrent and due to acute on chronic heart failure |
| SNOMED CT | 7381000175100 | Reduced ejection fraction co-occurrent and due to chronic heart failure |
| SNOMED CT | 7391000175102 | Reduced ejection fraction co-occurrent and due to acute heart failure |
| SNOMED CT | 7401000175100 | Acute on chronic heart failure co-occurrent with normal ejection fraction |
| SNOMED CT | 7421000175106 | Acute heart failure co-occurrent with normal ejection fraction |

Continued on next page

**Table17 – continued from previous page**

| CodeSystem | ConceptCode | ConceptName |
|---|---|---|
| SNOMED CT | 96311000119109 | Exacerbation of congestive heart failure |
| SNOMED CT | 285211000119102 | Congestive heart failure as post-operative complication of cardiac surgery |
| SNOMED CT | 285221000119109 | Congestive heart failure as post-operative complication of non-cardiac surgery |
| SNOMED CT | 16838951000119100 | Acute on chronic right-sided congestive heart failure |
| SNOMED CT | 13839000 | Right ventricular obstruction - failure syndrome |
| SNOMED CT | 78862003 | Cardiopathia nigra |
| SNOMED CT | 5053004 | Cardiac insufficiency due to prosthesis |
| SNOMED CT | 10335000 | Chronic right-sided heart failure |
| SNOMED CT | 16253001 | Dilated peripartum cardiomyopathy |
| SNOMED CT | 24841007 | Cardiorespiratory failure during AND/OR resulting from a procedure |
| SNOMED CT | 33644002 | Postvalvulotomy syndrome |
| SNOMED CT | 45650007 | Kyphoscoliotic heart disease |
| SNOMED CT | 49584005 | Acute cor pulmonale |
| SNOMED CT | 55565007 | Cardiac failure after obstetrical surgery AND/OR other procedure including delivery |
| SNOMED CT | 60856006 | Cardiac insufficiency following cardiac surgery |
| SNOMED CT | 62377009 | Peripartum cardiomyopathy in puerperium |
| SNOMED CT | 66989003 | Chronic right-sided congestive heart failure |
| SNOMED CT | 71892000 | Cardiac asthma |
| SNOMED CT | 79955004 | Chronic cardiopulmonary disease |
| SNOMED CT | 80479009 | Acute right-sided congestive heart failure |
| SNOMED CT | 426012001 | Right heart failure due to pulmonary hypertension |
| SNOMED CT | 276514007 | Neonatal cardiac failure |
| SNOMED CT | 277638005 | Sepsis-associated left ventricular failure |
| SNOMED CT | 277639002 | Sepsis-associated right ventricular failure |
| SNOMED CT | 1010444007 | Legal abortion complicated by cardiac failure |
| SNOMED CT | 1010447000 | Illegal abortion complicated by cardiac failure |
| SNOMED CT | 15964661000119102 | Acute cor pulmonale due to septic pulmonary embolism |
| SNOMED CT | 898208007 | Heart failure due to thyrotoxicosis |
| SNOMED CT | 1204206001 | Left ventricular failure with normal ejection fraction due to cardiomyopathy |
| SNOMED CT | 1204203009 | Left ventricular failure with normal ejection fraction due to coronary artery disease |
| SNOMED CT | 1204204003 | Left ventricular failure with normal ejection fraction due to myocarditis |
| SNOMED CT | 1204200007 | Left ventricular failure with normal ejection fraction due to valvular heart disease |
| SNOMED CT | 1204462004 | Left ventricular failure with sepsis |
| SNOMED CT | 1208846006 | Right ventricular failure due to pulmonary disease |
| SNOMED CT | 1208848007 | Right ventricular failure due to pulmonary vascular disease |

Continued on next page

**Table17 – continued from previous page**

| CodeSystem | ConceptCode | ConceptName |
|---|---|---|
| SNOMED CT | 1208850004 | Right ventricular failure due to right ventricular infarction |
| SNOMED CT | 1208843003 | Right ventricular failure due to valvular heart disease |
| SNOMED CT | 1204468000 | Right ventricular failure with sepsis |
| SNOMED CT | 1204460007 | Myocardial dysfunction with sepsis |
| SNOMED CT | 83291003 | Right heart failure due to pulmonary hypertension |
| SNOMED CT | 89819002 | Cardiac insufficiency during AND/OR resulting from a procedure |
| SNOMED CT | 359617009 | Acute right-sided heart failure |
| SNOMED CT | 367363000 | Right ventricular failure |
| SNOMED CT | 445236007 | Cardiorenal syndrome |
| SNOMED CT | 462172006 | Fetal heart failure |
| SNOMED CT | 462174007 | Fetal heart failure with myocardial hypertrophy |
| SNOMED CT | 462175008 | Fetal heart failure with redistribution of cardiac output |
| SNOMED CT | 609507007 | Induced termination of pregnancy complicated by cardiac failure |
| SNOMED CT | 704242009 | Fetal heart failure due to extracardiac disease |
| SNOMED CT | 722095005 | Acute kidney injury due to circulatory failure |
| SNOMED CT | 722919003 | Neonatal cardiac failure due to decreased left ventricular output |
| SNOMED CT | 724550005 | Neonatal cardiac failure due to pulmonary overperfusion |

###### 8.4.8 Hyperlipidemia

Table 18: Concept codes used to identify if a patient was diagnosed with hyperlipidemia.

| CodeSystem | ConceptCode | ConceptName |
|---|---|---|
| ICD10CM | E78.0 | Pure hypercholesterolemia |
| ICD10CM | E78.00 | Pure hypercholesterolemia, unspecified |
| ICD10CM | E78.01 | Familial hypercholesterolemia |
| ICD10CM | E78.1 | Pure hyperglyceridemia |
| ICD10CM | E78.2 | Mixed hyperlipidemia |
| ICD10CM | E78.3 | Hyperchylomicronemia |
| ICD10CM | E78.4 | Other hyperlipidemia |
| ICD10CM | E78.41 | Elevated Lipoprotein(a) |
| ICD10CM | E78.49 | Other hyperlipidemia |
| ICD10CM | E78.5 | Hyperlipidemia, unspecified |
| SNOMED CT | 701000119103 | Mixed hyperlipidemia due to type 2 diabetes mellitus |
| SNOMED CT | 1571000119104 | Mixed hyperlipidemia due to type 1 diabetes mellitus |
| SNOMED CT | 15771000119109 | Familial hyperalphalipoproteinemia |
| SNOMED CT | 114831000119107 | Hyperlipidemia caused by steroid |

Continued on next page

**Table18 – continued from previous page**

| CodeSystem | ConceptCode | ConceptName |
|---|---|---|
| SNOMED CT | 137931000119102 | Hyperlipidemia due to type 2 diabetes mellitus |
| SNOMED CT | 137941000119106 | Hyperlipidemia due to type 1 diabetes mellitus |
| SNOMED CT | 403829002 | Familial hypercholesterolemia due to heterozygous low density lipoprotein receptor mutation |
| SNOMED CT | 403830007 | Familial hypercholesterolemia due to homozygous low density lipoprotein receptor mutation |
| SNOMED CT | 767133009 | Compound heterozygous familial hypercholesterolemia |
| SNOMED CT | 238040008 | Familial combined hyperlipidemia |
| SNOMED CT | 238076009 | Primary hypercholesterolemia |
| SNOMED CT | 238077000 | Polygenic hypercholesterolemia |
| SNOMED CT | 238078005 | Familial hypercholesterolemia - homozygous |
| SNOMED CT | 238079002 | Familial hypercholesterolemia - heterozygous |
| SNOMED CT | 238080004 | Hyperalphalipoproteinemia |
| SNOMED CT | 238081000 | Familial defective apolipoprotein B-100 |
| SNOMED CT | 238082007 | Secondary hypercholesterolemia |
| SNOMED CT | 238083002 | Primary hypertriglyceridemia |
| SNOMED CT | 238084008 | Very low density lipoprotinemia |
| SNOMED CT | 238085009 | Fredrickson type IV hyperlipoproteinemia |
| SNOMED CT | 238087001 | Secondary hypertriglyceridemia |
| SNOMED CT | 238088006 | Primary combined hyperlipidemia |
| SNOMED CT | 238089003 | Secondary combined hyperlipidemia |
| SNOMED CT | 445261005 | Posttransplant hyperlipidemia |
| SNOMED CT | 773649005 | Transient infantile hypertriglyceridemia and hepatosteatosis |
| SNOMED CT | 773726000 | Hypercholesterolemia due to cholesterol 7alpha-hydroxylase deficiency |
| SNOMED CT | 190774002 | Hyperlipidemia, group A |
| SNOMED CT | 129589009 | Endogenous hyperlipidemia |
| SNOMED CT | 129590000 | Exogenous hyperlipidemia |
| SNOMED CT | 129591001 | Mixed hypercholesterolemia and hypertriglyceridemia |
| SNOMED CT | 13644009 | Hypercholesterolemia |
| SNOMED CT | 33513003 | Familial apoC-II deficiency |
| SNOMED CT | 34349009 | Familial type 5 hyperlipoproteinemia |
| SNOMED CT | 34528009 | Familial hypertriglyceridemia |
| SNOMED CT | 55822004 | Hyperlipidemia |
| SNOMED CT | 397915002 | Fredrickson type IIa hyperlipoproteinemia |
| SNOMED CT | 398036000 | Familial hypercholesterolemia |
| SNOMED CT | 402473001 | Sporadic primary hypertriglyceridemia |
| SNOMED CT | 402474007 | Primary polygenic type IIb combined hyperlipidemia |
| SNOMED CT | 402475008 | Primary acquired chylomicronemia |
| SNOMED CT | 402725005 | Hyperlipidemia with lipid deposition in skin |
| SNOMED CT | 402726006 | Primary chylomicronemia |
| SNOMED CT | 402727002 | Secondary hyperlipidemia |

Continued on next page

**Table18 – continued from previous page**

| CodeSystem | ConceptCode | ConceptName |
|---|---|---|
| SNOMED CT | 402785008 | Primary genetic hyperlipidemia |
| SNOMED CT | 402786009 | Chylomicronemia syndrome |
| SNOMED CT | 402787000 | Primary genetic mixed hyperlipidemia |
| SNOMED CT | 403827000 | Familial lipoprotein lipase deficiency with type I phenotype |
| SNOMED CT | 403828005 | Familial lipoprotein lipase deficiency with type V phenotype |
| SNOMED CT | 403831006 | Familial Combined Hypercholesterolaemia |
| SNOMED CT | 426161002 | Chemically induced hyperlipidemia |
| SNOMED CT | 267432004 | Pure hypercholesterolemia |
| SNOMED CT | 267433009 | Pure hyperglyceridemia |
| SNOMED CT | 267434003 | Mixed hyperlipidemia |
| SNOMED CT | 267435002 | Familial hyperchylomicronemia |
| SNOMED CT | 299465007 | Familial multiple lipoprotein-type hyperlipidemia |
| SNOMED CT | 302870006 | Hypertriglyceridemia |
| SNOMED CT | 275598004 | Familial hyperlipoproteinemia, type I |
| SNOMED CT | 1197489003 | Familial chylomicronemia syndrome |
| SNOMED CT | 1208738002 | TMEM199 congenital disorder of glycosylation |

###### 8.4.9 Hypertension

Table 19: Concept codes used to identify if a patient was diagnosed with hypertension.

| CodeSystem | ConceptCode | ConceptName |
|---|---|---|
| ICD10CM | I10 | Essential (primary) hypertension |
| ICD10CM | I11.0 | Hypertensive heart disease with heart failure |
| ICD10CM | I11.9 | Hypertensive heart disease without heart failure |
| ICD10CM | I12.0 | Hypertensive chronic kidney disease with stage 5 chronic kidney disease or end stage renal disease |
| ICD10CM | I12.9 | Hypertensive chronic kidney disease with stage 1 through stage 4 chronic kidney disease, or unspecified chronic kidney disease |
| ICD10CM | I13.0 | Hypertensive heart and chronic kidney disease with heart failure and stage 1 through stage 4 chronic kidney disease, or unspecified chronic kidney disease |
| ICD10CM | I13.1 | Hypertensive heart and chronic kidney disease without heart failure |
| ICD10CM | I13.10 | Hypertensive heart and chronic kidney disease without heart failure, with stage 1 through stage 4 chronic kidney disease, or unspecified chronic kidney disease |

Continued on next page

**Table19 – continued from previous page**

| CodeSystem | ConceptCode | ConceptName |
|---|---|---|
| ICD10CM | I13.11 | Hypertensive heart and chronic kidney disease without heart failure, with stage 5 chronic kidney disease, or end stage renal disease |
| ICD10CM | I13.2 | Hypertensive heart and chronic kidney disease with heart failure and with stage 5 chronic kidney disease, or end stage renal disease |
| ICD10CM | I15 | Secondary hypertension |
| ICD10CM | I15.0 | Renovascular hypertension |
| ICD10CM | I15.1 | Hypertension secondary to other renal disorders |
| ICD10CM | I15.2 | Hypertension secondary to endocrine disorders |
| ICD10CM | I15.8 | Other secondary hypertension |
| ICD10CM | I15.9 | Secondary hypertension, unspecified |
| ICD10CM | I16 | Hypertensive crisis |
| ICD10CM | I16.0 | Hypertensive urgency |
| ICD10CM | I16.1 | Hypertensive emergency |
| ICD10CM | I16.9 | Hypertensive crisis, unspecified |
| ICD9CM | 401.0 | Malignant essential hypertension |
| ICD9CM | 401.1 | Benign essential hypertension |
| ICD9CM | 401.9 | Unspecified essential hypertension |
| ICD9CM | 402.00 | Malignant hypertensive heart disease without heart failure |
| ICD9CM | 402.01 | Malignant hypertensive heart disease with heart failure |
| ICD9CM | 402.10 | Benign hypertensive heart disease without heart failure |
| ICD9CM | 402.11 | Benign hypertensive heart disease with heart failure |
| ICD9CM | 402.90 | Unspecified hypertensive heart disease without heart failure |
| ICD9CM | 402.91 | Unspecified hypertensive heart disease with heart failure |
| ICD9CM | 403.00 | Hypertensive chronic kidney disease, malignant, with chronic kidney disease stage I through stage IV, or unspecified |
| ICD9CM | 403.01 | Hypertensive chronic kidney disease, malignant, with chronic kidney disease stage V or end stage renal disease |
| ICD9CM | 403.10 | Hypertensive chronic kidney disease, benign, with chronic kidney disease stage I through stage IV, or unspecified |
| ICD9CM | 403.11 | Hypertensive chronic kidney disease, benign, with chronic kidney disease stage V or end stage renal disease |
| ICD9CM | 403.90 | Hypertensive chronic kidney disease, unspecified, with chronic kidney disease stage I through stage IV, or unspecified |
| ICD9CM | 403.91 | Hypertensive chronic kidney disease, unspecified, with chronic kidney disease stage V or end stage renal disease |
| ICD9CM | 404.00 | Hypertensive heart and chronic kidney disease, malignant, without heart failure and with chronic kidney disease stage I through stage IV, or unspecified |

Continued on next page

**Table19 – continued from previous page**

| CodeSystem | ConceptCode | ConceptName |
|---|---|---|
| ICD9CM | 404.01 | Hypertensive heart and chronic kidney disease, malignant, with heart failure and with chronic kidney disease stage I through stage IV, or unspecified |
| ICD9CM | 404.02 | Hypertensive heart and chronic kidney disease, malignant, without heart failure and with chronic kidney disease stage V or end stage renal disease |
| ICD9CM | 404.03 | Hypertensive heart and chronic kidney disease, malignant, with heart failure and with chronic kidney disease stage V or end stage renal disease |
| ICD9CM | 404.10 | Hypertensive heart and chronic kidney disease, benign, without heart failure and with chronic kidney disease stage I through stage IV, or unspecified |
| ICD9CM | 404.11 | Hypertensive heart and chronic kidney disease, benign, with heart failure and with chronic kidney disease stage I through stage IV, or unspecified |
| ICD9CM | 404.12 | Hypertensive heart and chronic kidney disease, benign, without heart failure and with chronic kidney disease stage V or end stage renal disease |
| ICD9CM | 404.13 | Hypertensive heart and chronic kidney disease, benign, with heart failure and chronic kidney disease stage V or end stage renal disease |
| ICD9CM | 404.90 | Hypertensive heart and chronic kidney disease, unspecified, without heart failure and with chronic kidney disease stage I through stage IV, or unspecified |
| ICD9CM | 404.91 | Hypertensive heart and chronic kidney disease, unspecified, with heart failure and with chronic kidney disease stage I through stage IV, or unspecified |
| ICD9CM | 404.92 | Hypertensive heart and chronic kidney disease, unspecified, without heart failure and with chronic kidney disease stage V or end stage renal disease |
| ICD9CM | 404.93 | Hypertensive heart and chronic kidney disease, unspecified, with heart failure and chronic kidney disease stage V or end stage renal disease |
| ICD9CM | 405.01 | Malignant renovascular hypertension |
| ICD9CM | 405.09 | Other malignant secondary hypertension |
| ICD9CM | 405.11 | Benign renovascular hypertension |
| ICD9CM | 405.19 | Other benign secondary hypertension |
| ICD9CM | 405.91 | Unspecified renovascular hypertension |
| ICD9CM | 405.99 | Other unspecified secondary hypertension |
| SNOMED CT | 1201005 | Benign essential hypertension |
| SNOMED CT | 10725009 | Benign hypertension |
| SNOMED CT | 14973001 | Renal sclerosis with hypertension |

Continued on next page

**Table19 – continued from previous page**

| CodeSystem | ConceptCode | ConceptName |
|---|---|---|
| SNOMED CT | 26078007 | Hypertension secondary to renal disease complicating AND/OR reason for care during childbirth |
| SNOMED CT | 28119000 | Renal hypertension |
| SNOMED CT | 31992008 | Secondary hypertension |
| SNOMED CT | 284981000119102 | Chronic kidney disease stage 2 due to benign hypertension |
| SNOMED CT | 284991000119104 | Chronic kidney disease stage 3 due to benign hypertension |
| SNOMED CT | 461301000124109 | Resistant hypertensive disorder |
| SNOMED CT | 1078301000112109 | Multiple drug intolerant hypertension |
| SNOMED CT | 89242004 | Malignant secondary hypertension |
| SNOMED CT | 111438007 | Hypertension secondary to renal disease in obstetric context |
| SNOMED CT | 123799005 | Renovascular hypertension |
| SNOMED CT | 123800009 | Goldblatt hypertension |
| SNOMED CT | 371125006 | Labile essential hypertension |
| SNOMED CT | 762463000 | Diastolic hypertension and systolic hypertension |
| SNOMED CT | 169465000 | Hypertension induced by oral contraceptive pill |
| SNOMED CT | 194783001 | Malignant secondary renovascular hypertension |
| SNOMED CT | 194785008 | Benign secondary hypertension |
| SNOMED CT | 194788005 | Hypertension secondary to endocrine disorder |
| SNOMED CT | 194791005 | Hypertension secondary to drug |
| SNOMED CT | 199008003 | Pre-existing secondary hypertension complicating pregnancy, childbirth and puerperium |
| SNOMED CT | 38341003 | Hypertensive disorder |
| SNOMED CT | 39018007 | Renal arterial hypertension |
| SNOMED CT | 39727004 | Hypertension secondary to renal disease complicating AND/OR reason for care during puerperium |
| SNOMED CT | 46481004 | Low-renin essential hypertension |
| SNOMED CT | 48146000 | Diastolic hypertension |
| SNOMED CT | 48552006 | Hypertension secondary to renal disease complicating AND/OR reason for care during pregnancy |
| SNOMED CT | 56218007 | Systolic hypertension |
| SNOMED CT | 57684003 | Parenchymal renal hypertension |
| SNOMED CT | 59621000 | Essential hypertension |
| SNOMED CT | 59720008 | Sustained diastolic hypertension |
| SNOMED CT | 65518004 | Labile diastolic hypertension |
| SNOMED CT | 73410007 | Benign secondary renovascular hypertension |
| SNOMED CT | 74451002 | Secondary diastolic hypertension |
| SNOMED CT | 78975002 | Malignant essential hypertension |
| SNOMED CT | 427889009 | Hypertension associated with transplantation |
| SNOMED CT | 428575007 | Hypertension secondary to kidney transplant |
| SNOMED CT | 429457004 | Systolic essential hypertension |
| SNOMED CT | 1204139007 | Hypertension due to congenital adrenal hyperplasia |
| SNOMED CT | 1208845005 | Secondary hypertension due to congenital heart disorder |
| SNOMED CT | 1208839002 | Secondary hypertension due to renal tubular disorder |

Continued on next page

**Table19 – continued from previous page**

| CodeSystem | ConceptCode | ConceptName |
|---|---|---|
| SNOMED CT | 8218002 | Chronic hypertension complicating AND/OR reason for care during childbirth |
| SNOMED CT | 8762007 | Chronic hypertension in obstetric context |
| SNOMED CT | 9901000 | Essential hypertension complicating AND/OR reason for care during puerperium |
| SNOMED CT | 10562009 | Malignant hypertension complicating AND/OR reason for care during childbirth |
| SNOMED CT | 15394000 | Toxemia of pregnancy |
| SNOMED CT | 18416000 | Essential hypertension complicating AND/OR reason for care during childbirth |
| SNOMED CT | 19769006 | High-renin essential hypertension |
| SNOMED CT | 23130000 | Paroxysmal hypertension |
| SNOMED CT | 23717007 | Benign essential hypertension complicating AND/OR reason for care during pregnancy |
| SNOMED CT | 23786008 | Malignant hypertension complicating AND/OR reason for care during puerperium |
| SNOMED CT | 24042004 | Chronic hypertension complicating AND/OR reason for care during puerperium |
| SNOMED CT | 29259002 | Malignant hypertension complicating AND/OR reason for care during pregnancy |
| SNOMED CT | 31407004 | Pre-existing hypertension complicating AND/OR reason for care during puerperium |
| SNOMED CT | 81626002 | Malignant hypertension in obstetric context |
| SNOMED CT | 84094009 | Rebound hypertension |
| SNOMED CT | 86041002 | Pre-existing hypertension in obstetric context |
| SNOMED CT | 697929007 | Intermittent hypertension |
| SNOMED CT | 697930002 | Labile hypertension due to being in a clinical environment |
| SNOMED CT | 698638005 | Pregnancy induced hypertension with pulmonary edema |
| SNOMED CT | 698640000 | Hypertension in the puerperium with pulmonary edema |
| SNOMED CT | 704667004 | Hypertension concurrent and due to end stage renal disease on dialysis |
| SNOMED CT | 706882009 | Hypertensive crisis |
| SNOMED CT | 712832005 | Supine hypertension |
| SNOMED CT | 720568003 | Brachydactyly and arterial hypertension syndrome |
| SNOMED CT | 765182005 | Postpartum pre-eclampsia |
| SNOMED CT | 766937004 | Hypertension due to gain-of-function mutation in mineralocorticoid receptor |
| SNOMED CT | 198941007 | Hypertension complicating pregnancy, childbirth and the puerperium |
| SNOMED CT | 198942000 | Benign essential hypertension complicating pregnancy, childbirth and the puerperium |
| SNOMED CT | 198944004 | Benign essential hypertension complicating pregnancy, childbirth and the puerperium - delivered |

Continued on next page

**Table19 – continued from previous page**

| CodeSystem | ConceptCode | ConceptName |
|---|---|---|
| SNOMED CT | 198945003 | Benign essential hypertension complicating pregnancy, childbirth and the puerperium - delivered with postnatal complication |
| SNOMED CT | 198946002 | Benign essential hypertension complicating pregnancy, childbirth and the puerperium - not delivered |
| SNOMED CT | 198947006 | Benign essential hypertension complicating pregnancy, childbirth and the puerperium with postnatal complication |
| SNOMED CT | 198949009 | Renal hypertension complicating pregnancy, childbirth and the puerperium |
| SNOMED CT | 198951008 | Renal hypertension complicating pregnancy, childbirth and the puerperium - delivered |
| SNOMED CT | 198952001 | Renal hypertension complicating pregnancy, childbirth and the puerperium - delivered with postnatal complication |
| SNOMED CT | 198953006 | Renal hypertension complicating pregnancy, childbirth and the puerperium - not delivered |
| SNOMED CT | 198965005 | Transient hypertension of pregnancy - delivered |
| SNOMED CT | 198966006 | Transient hypertension of pregnancy - delivered with postnatal complication |
| SNOMED CT | 198967002 | Transient hypertension of pregnancy - not delivered |
| SNOMED CT | 198968007 | Transient hypertension of pregnancy with postnatal complication |
| SNOMED CT | 198983002 | Severe pre-eclampsia - delivered |
| SNOMED CT | 198984008 | Severe pre-eclampsia - delivered with postnatal complication |
| SNOMED CT | 198985009 | Severe pre-eclampsia - not delivered |
| SNOMED CT | 198986005 | Severe pre-eclampsia with postnatal complication |
| SNOMED CT | 198997005 | Pre-eclampsia or eclampsia with pre-existing hypertension |
| SNOMED CT | 198999008 | Pre-eclampsia or eclampsia with pre-existing hypertension - delivered |
| SNOMED CT | 199000005 | Pre-eclampsia or eclampsia with pre-existing hypertension - delivered with postnatal complication |
| SNOMED CT | 199002002 | Pre-eclampsia or eclampsia with pre-existing hypertension - not delivered |
| SNOMED CT | 199003007 | Pre-eclampsia or eclampsia with pre-existing hypertension with postnatal complication |
| SNOMED CT | 199005000 | Pre-existing hypertension complicating pregnancy, childbirth and puerperium |
| SNOMED CT | 199007008 | Pre-existing hypertensive heart and renal disease complicating pregnancy, childbirth and the puerperium |
| SNOMED CT | 206596003 | Neonatal hypertension |
| SNOMED CT | 307632004 | Non-proteinuric hypertension of pregnancy |
| SNOMED CT | 367390009 | Hypertension in the obstetric context |
| SNOMED CT | 237279007 | Transient hypertension of pregnancy |

Continued on next page

**Table19 – continued from previous page**

| CodeSystem | ConceptCode | ConceptName |
|---|---|---|
| SNOMED CT | 237281009 | Moderate proteinuric hypertension of pregnancy |
| SNOMED CT | 237282002 | Impending eclampsia |
| SNOMED CT | 871642009 | Hypertension due to aortic arch obstruction |
| SNOMED CT | 541000119105 | Hypertension complicating pregnancy, childbirth and the puerperium, antepartum |
| SNOMED CT | 5501000119106 | Postoperative hypertension |
| SNOMED CT | 40511000119107 | Postpartum pre-existing essential hypertension |
| SNOMED CT | 40521000119100 | Postpartum pregnancy-induced hypertension |
| SNOMED CT | 71421000119105 | Hypertension in chronic kidney disease due to type 2 diabetes mellitus |
| SNOMED CT | 71701000119105 | Hypertension in chronic kidney disease due to type 1 diabetes mellitus |
| SNOMED CT | 82771000119102 | Hypertension complicating pregnancy |
| SNOMED CT | 118781000119108 | Pre-existing hypertensive chronic kidney disease in mother complicating pregnancy |
| SNOMED CT | 127991000119101 | Hypertension concurrent and due to end stage renal disease on dialysis due to type 2 diabetes mellitus |
| SNOMED CT | 128001000119105 | Hypertension concurrent and due to end stage renal disease on dialysis due to type 1 diabetes mellitus |
| SNOMED CT | 132721000119104 | Hypertensive emergency |
| SNOMED CT | 140101000119109 | Hypertension in chronic kidney disease stage 5 due to type 2 diabetes mellitus |
| SNOMED CT | 140111000119107 | Hypertension in chronic kidney disease stage 4 due to type 2 diabetes mellitus |
| SNOMED CT | 140121000119100 | Hypertension in chronic kidney disease stage 3 due to type 2 diabetes mellitus |
| SNOMED CT | 140131000119102 | Hypertension in chronic kidney disease stage 2 due to type 2 diabetes mellitus |
| SNOMED CT | 367821000119106 | Page kidney |
| SNOMED CT | 434711000124103 | Perioperative hypertension |
| SNOMED CT | 10752641000119102 | Eclampsia with pre-existing hypertension in childbirth |
| SNOMED CT | 10757401000119104 | Pre-existing hypertensive heart and chronic kidney disease in mother complicating childbirth |
| SNOMED CT | 16229371000119106 | Labile systemic arterial hypertension |
| SNOMED CT | 48194001 | GH - Gestational hypertension |
| SNOMED CT | 95605009 | Hemolysis-elevated liver enzymes-low platelet count syndrome |
| SNOMED CT | 34694006 | Pre-existing hypertension complicating AND/OR reason for care during childbirth |
| SNOMED CT | 35303009 | Benign essential hypertension complicating AND/OR reason for care during puerperium |
| SNOMED CT | 37618003 | Chronic hypertension complicating AND/OR reason for care during pregnancy |

Continued on next page

**Table19 – continued from previous page**

| CodeSystem | ConceptCode | ConceptName |
|---|---|---|
| SNOMED CT | 41114007 | Mild pre-eclampsia |
| SNOMED CT | 46764007 | Severe pre-eclampsia |
| SNOMED CT | 52698002 | Transient hypertension |
| SNOMED CT | 63287004 | Benign essential hypertension in obstetric context |
| SNOMED CT | 65402008 | Pre-existing hypertension complicating AND/OR reason for care during pregnancy |
| SNOMED CT | 67359005 | Pre-eclampsia added to pre-existing hypertension |
| SNOMED CT | 69909000 | Eclampsia added to pre-existing hypertension |
| SNOMED CT | 70272006 | Malignant hypertension |
| SNOMED CT | 71874008 | Benign essential hypertension complicating AND/OR reason for care during childbirth |
| SNOMED CT | 72022006 | Essential hypertension in obstetric context |
| SNOMED CT | 78808002 | Essential hypertension complicating AND/OR reason for care during pregnancy |
| SNOMED CT | 397748008 | Hypertension with albuminuria |
| SNOMED CT | 398254007 | Toxemia of pregnancy |
| SNOMED CT | 429198000 | Exertional hypertension |
| SNOMED CT | 443482000 | Hypertensive urgency |
| SNOMED CT | 288250001 | Maternal hypertension |

###### 8.4.10 Ischemic Stroke

Table 20: Concept codes used to identify if a patient was diagnosed with ischemic-stroke.

| CodeSystem | ConceptCode | ConceptName |
|---|---|---|
| ICD10CM | I63.10 | Cerebral infarction due to embolism of unspecified precerebral artery |
| ICD10CM | I63.111 | Cerebral infarction due to embolism of right vertebral artery |
| ICD10CM | I63.112 | Cerebral infarction due to embolism of left vertebral artery |
| ICD10CM | I63.113 | Cerebral infarction due to embolism of bilateral vertebral arteries |
| ICD10CM | I63.119 | Cerebral infarction due to embolism of unspecified vertebral artery |
| ICD10CM | I63.12 | Cerebral infarction due to embolism of basilar artery |
| ICD10CM | I63.131 | Cerebral infarction due to embolism of right carotid artery |
| ICD10CM | I63.132 | Cerebral infarction due to embolism of left carotid artery |
| ICD10CM | I63.133 | Cerebral infarction due to embolism of bilateral carotid arteries |
| ICD10CM | I63.139 | Cerebral infarction due to embolism of unspecified carotid artery |

Continued on next page

**Table20 – continued from previous page**

| CodeSystem | ConceptCode | ConceptName |
|---|---|---|
| ICD10CM | I63.19 | Cerebral infarction due to embolism of other precerebral artery |
| ICD10CM | I63.40 | Cerebral infarction due to embolism of unspecified cerebral artery |
| ICD10CM | I63.411 | Cerebral infarction due to embolism of right middle cerebral artery |
| ICD10CM | I63.412 | Cerebral infarction due to embolism of left middle cerebral artery |
| ICD10CM | I63.413 | Cerebral infarction due to embolism of bilateral middle cerebral arteries |
| ICD10CM | I63.419 | Cerebral infarction due to embolism of unspecified middle cerebral artery |
| ICD10CM | I63.421 | Cerebral infarction due to embolism of right anterior cerebral artery |
| ICD10CM | I63.422 | Cerebral infarction due to embolism of left anterior cerebral artery |
| ICD10CM | I63.423 | Cerebral infarction due to embolism of bilateral anterior cerebral arteries |
| ICD10CM | I63.429 | Cerebral infarction due to embolism of unspecified anterior cerebral artery |
| ICD10CM | I63.431 | Cerebral infarction due to embolism of right posterior cerebral artery |
| ICD10CM | I63.432 | Cerebral infarction due to embolism of left posterior cerebral artery |
| ICD10CM | I63.433 | Cerebral infarction due to embolism of bilateral posterior cerebral arteries |
| ICD10CM | I63.439 | Cerebral infarction due to embolism of unspecified posterior cerebral artery |
| ICD10CM | I63.441 | Cerebral infarction due to embolism of right cerebellar artery |
| ICD10CM | I63.442 | Cerebral infarction due to embolism of left cerebellar artery |
| ICD10CM | I63.443 | Cerebral infarction due to embolism of bilateral cerebellar arteries |
| ICD10CM | I63.449 | Cerebral infarction due to embolism of unspecified cerebellar artery |
| ICD10CM | I63.49 | Cerebral infarction due to embolism of other cerebral artery |
| ICD9CM | 433.01 | Occlusion and stenosis of basilar artery with cerebral infarction |
| ICD9CM | 433.10 | Occlusion and stenosis of carotid artery without mention of cerebral infarction |
| ICD9CM | 433.11 | Occlusion and stenosis of carotid artery with cerebral infarction |

Continued on next page

**Table20 – continued from previous page**

| CodeSystem | ConceptCode | ConceptName |
|---|---|---|
| ICD9CM | 433.21 | Occlusion and stenosis of vertebral artery with cerebral infarction |
| ICD9CM | 433.31 | Occlusion and stenosis of multiple and bilateral precerebral arteries with cerebral infarction |
| ICD9CM | 433.81 | Occlusion and stenosis of other specified precerebral artery with cerebral infarction |
| ICD9CM | 433.91 | Occlusion and stenosis of unspecified precerebral artery with cerebral infarction |
| ICD9CM | 434.00 | Cerebral thrombosis without mention of cerebral infarction |
| ICD9CM | 434.01 | Cerebral thrombosis with cerebral infarction |
| ICD9CM | 434.11 | Cerebral embolism with cerebral infarction |
| ICD9CM | 434.91 | Cerebral artery occlusion, unspecified with cerebral infarction |
| ICD9CM | 436 | Acute, but ill-defined, cerebrovascular disease |
| SNOMED CT | 195186005 | Cerebral infarction due to embolism of precerebral arteries |
| SNOMED CT | 195190007 | Cerebral infarction due to embolism of cerebral arteries |

###### 8.4.11 Major Depressive Disorder

Table 21: Concept codes used to identify if a patient was diagnosed with major-depression.

| CodeSystem | ConceptCode | ConceptName |
|---|---|---|
| ICD10CM | F32.0 | Major depressive disorder, single episode, mild |
| ICD10CM | F32.1 | Major depressive disorder, single episode, moderate |
| ICD10CM | F32.2 | Major depressive disorder, single episode, severe without psychotic features |
| ICD10CM | F32.3 | Major depressive disorder, single episode, severe with psychotic features |
| ICD10CM | F32.4 | Major depressive disorder, single episode, in partial remission |
| ICD10CM | F32.9 | Major depressive disorder, single episode, unspecified |
| ICD10CM | F33.0 | Major depressive disorder, recurrent, mild |
| ICD10CM | F33.1 | Major depressive disorder, recurrent, moderate |
| ICD10CM | F33.2 | Major depressive disorder, recurrent severe without psychotic features |
| ICD10CM | F33.3 | Major depressive disorder, recurrent, severe with psychotic symptoms |
| ICD10CM | F33.41 | Major depressive disorder, recurrent, in partial remission |
| ICD10CM | F33.9 | Major depressive disorder, recurrent, unspecified |
| SNOMED CT | 832007 | Moderate major depression |

Continued on next page

**Table21 – continued from previous page**

| CodeSystem | ConceptCode | ConceptName |
|---|---|---|
| SNOMED CT | 2618002 | Chronic recurrent major depressive disorder |
| SNOMED CT | 14183003 | Chronic major depressive disorder, single episode |
| SNOMED CT | 15193003 | Severe recurrent major depression with psychotic features, mood-incongruent |
| SNOMED CT | 15639000 | Moderate major depression, single episode |
| SNOMED CT | 18818009 | Moderate recurrent major depression |
| SNOMED CT | 20250007 | Severe major depression, single episode, with psychotic features, mood-incongruent |
| SNOMED CT | 25922000 | Major depressive disorder, single episode with postpartum onset |
| SNOMED CT | 28475009 | Severe recurrent major depression with psychotic features |
| SNOMED CT | 33078009 | Severe recurrent major depression with psychotic features, mood-congruent |
| SNOMED CT | 33736005 | Severe major depression with psychotic features, mood-congruent |
| SNOMED CT | 87512008 | Mild major depression |
| SNOMED CT | 251000119105 | Severe major depression, single episode |
| SNOMED CT | 281000119103 | Severe recurrent major depression |
| SNOMED CT | 10811121000119102 | Major depressive disorder in mother complicating childbirth |
| SNOMED CT | 10811161000119107 | Major depressive disorder in mother complicating pregnancy |
| SNOMED CT | 16264621000119109 | Recurrent mild major depressive disorder co-occurrent with anxiety |
| SNOMED CT | 16264901000119109 | Recurrent moderate major depressive disorder co-occurrent with anxiety |
| SNOMED CT | 16265951000119109 | Mild major depressive disorder co-occurrent with anxiety single episode |
| SNOMED CT | 16266831000119100 | Moderate major depressive disorder co-occurrent with anxiety single episode |
| SNOMED CT | 16266991000119108 | Severe major depressive disorder co-occurrent with anxiety single episode |
| SNOMED CT | 36474008 | Severe recurrent major depression without psychotic features |
| SNOMED CT | 36923009 | Major depression, single episode |
| SNOMED CT | 38694004 | Recurrent major depressive disorder with atypical features |
| SNOMED CT | 39809009 | Recurrent major depressive disorder with catatonic features |
| SNOMED CT | 40379007 | Mild recurrent major depression |
| SNOMED CT | 42925002 | Major depressive disorder, single episode with atypical features |
| SNOMED CT | 60099002 | Severe major depression with psychotic features, mood-incongruent |
| SNOMED CT | 63778009 | Major depressive disorder, single episode with melancholic features |

Continued on next page

**Table21 – continued from previous page**

| CodeSystem | ConceptCode | ConceptName |
|---|---|---|
| SNOMED CT | 66344007 | Recurrent major depression |
| SNOMED CT | 69392006 | Major depressive disorder, single episode with catatonic features |
| SNOMED CT | 71336009 | Recurrent major depressive disorder with postpartum onset |
| SNOMED CT | 73867007 | Severe major depression with psychotic features |
| SNOMED CT | 75084000 | Severe major depression without psychotic features |
| SNOMED CT | 76441001 | Severe major depression, single episode, without psychotic features |
| SNOMED CT | 77911002 | Severe major depression, single episode, with psychotic features, mood-congruent |
| SNOMED CT | 79298009 | Mild major depression, single episode |
| SNOMED CT | 430852001 | Severe major depression, single episode, with psychotic features |
| SNOMED CT | 268621008 | Recurrent major depressive episodes |
| SNOMED CT | 450714000 | Severe major depression |
| SNOMED CT | 719592004 | Moderately severe major depression |
| SNOMED CT | 720451004 | Minimal recurrent major depression |
| SNOMED CT | 720452006 | Moderately severe recurrent major depression |
| SNOMED CT | 720453001 | Moderately severe major depression single episode |
| SNOMED CT | 720454007 | Minimal major depression single episode |
| SNOMED CT | 720455008 | Minimal major depression |
| SNOMED CT | 726772006 | Major depression with psychotic features |
| SNOMED CT | 191604000 | Single major depressive episode, severe, with psychosis |
| SNOMED CT | 191610000 | Recurrent major depressive episodes, mild |
| SNOMED CT | 191611001 | Recurrent major depressive episodes, moderate |
| SNOMED CT | 191613003 | Recurrent major depressive episodes, severe, with psychosis |
| SNOMED CT | 319768000 | Recurrent major depressive disorder with melancholic features |
| SNOMED CT | 320751009 | Major depression, melancholic type |
| SNOMED CT | 370143000 | Major depressive disorder |
| SNOMED CT | 19527009 | Single episode of major depression in full remission |
| SNOMED CT | 30605009 | Major depression in partial remission |
| SNOMED CT | 33135002 | Recurrent major depression in partial remission |
| SNOMED CT | 42810003 | Major depression in remission |
| SNOMED CT | 46244001 | Recurrent major depression in full remission |
| SNOMED CT | 63412003 | Major depression in full remission |
| SNOMED CT | 68019004 | Recurrent major depression in remission |
| SNOMED CT | 70747007 | Major depression single episode, in partial remission |
| SNOMED CT | 104851000119103 | Postpartum major depression in remission |
| SNOMED CT | 16264821000119108 | Recurrent severe major depressive disorder co-occurrent with anxiety |
| SNOMED CT | 16265061000119105 | Recurrent major depressive disorder co-occurrent with anxiety in full remission |
| Continued on next page | | |

**Table21 – continued from previous page**

| CodeSystem | ConceptCode | ConceptName |
|---|---|---|
| SNOMED CT | 16265301000119106 | Recurrent major depressive disorder in partial remission co-occurrent with anxiety |

###### 8.4.12 Myocardial Infarction

Table 22: Concept codes used to identify if a patient was diagnosed with myocardial-infarction.

| CodeSystem | ConceptCode | ConceptName |
|---|---|---|
| ICD10CM | I21.01 | ST elevation (STEMI) myocardial infarction involving left main coronary artery |
| ICD10CM | I21.02 | ST elevation (STEMI) myocardial infarction involving left anterior descending coronary artery |
| ICD10CM | I21.09 | ST elevation (STEMI) myocardial infarction involving other coronary artery of anterior wall |
| ICD10CM | I21.11 | ST elevation (STEMI) myocardial infarction involving right coronary artery |
| ICD10CM | I21.19 | ST elevation (STEMI) myocardial infarction involving other coronary artery of inferior wall |
| ICD10CM | I21.21 | ST elevation (STEMI) myocardial infarction involving left circumflex coronary artery |
| ICD10CM | I21.29 | ST elevation (STEMI) myocardial infarction involving other sites |
| ICD10CM | I21.3 | ST elevation (STEMI) myocardial infarction of unspecified site |
| ICD10CM | I21.4 | Non-ST elevation (NSTEMI) myocardial infarction |
| ICD10CM | I21.9 | Acute myocardial infarction, unspecified |
| ICD10CM | I21.A1 | Myocardial infarction type 2 |
| ICD10CM | I21.A9 | Other myocardial infarction type |
| ICD10CM | I22.0 | Subsequent ST elevation (STEMI) myocardial infarction of anterior wall |
| ICD10CM | I22.1 | Subsequent ST elevation (STEMI) myocardial infarction of inferior wall |
| ICD10CM | I22.2 | Subsequent non-ST elevation (NSTEMI) myocardial infarction |
| ICD10CM | I22.8 | Subsequent ST elevation (STEMI) myocardial infarction of other sites |
| ICD10CM | I22.9 | Subsequent ST elevation (STEMI) myocardial infarction of unspecified site |
| ICD9CM | 410.00 | Acute myocardial infarction of anterolateral wall, episode of care unspecified |

Continued on next page

**Table22 – continued from previous page**

| CodeSystem | ConceptCode | ConceptName |
|---|---|---|
| ICD9CM | 410.01 | Acute myocardial infarction of anterolateral wall, initial episode of care |
| ICD9CM | 410.02 | Acute myocardial infarction of anterolateral wall, subsequent episode of care |
| ICD9CM | 410.10 | Acute myocardial infarction of other anterior wall, episode of care unspecified |
| ICD9CM | 410.11 | Acute myocardial infarction of other anterior wall, initial episode of care |
| ICD9CM | 410.12 | Acute myocardial infarction of other anterior wall, subsequent episode of care |
| ICD9CM | 410.20 | Acute myocardial infarction of inferolateral wall, episode of care unspecified |
| ICD9CM | 410.21 | Acute myocardial infarction of inferolateral wall, initial episode of care |
| ICD9CM | 410.22 | Acute myocardial infarction of inferolateral wall, subsequent episode of care |
| ICD9CM | 410.30 | Acute myocardial infarction of inferoposterior wall, episode of care unspecified |
| ICD9CM | 410.31 | Acute myocardial infarction of inferoposterior wall, initial episode of care |
| ICD9CM | 410.32 | Acute myocardial infarction of inferoposterior wall, subsequent episode of care |
| ICD9CM | 410.40 | Acute myocardial infarction of other inferior wall, episode of care unspecified |
| ICD9CM | 410.41 | Acute myocardial infarction of other inferior wall, initial episode of care |
| ICD9CM | 410.42 | Acute myocardial infarction of other inferior wall, subsequent episode of care |
| ICD9CM | 410.50 | Acute myocardial infarction of other lateral wall, episode of care unspecified |
| ICD9CM | 410.51 | Acute myocardial infarction of other lateral wall, initial episode of care |
| ICD9CM | 410.52 | Acute myocardial infarction of other lateral wall, subsequent episode of care |
| ICD9CM | 410.60 | True posterior wall infarction, episode of care unspecified |
| ICD9CM | 410.61 | True posterior wall infarction, initial episode of care |
| ICD9CM | 410.62 | True posterior wall infarction, subsequent episode of care |
| ICD9CM | 410.70 | Subendocardial infarction, episode of care unspecified |
| ICD9CM | 410.71 | Subendocardial infarction, initial episode of care |
| ICD9CM | 410.72 | Subendocardial infarction, subsequent episode of care |
| ICD9CM | 410.80 | Acute myocardial infarction of other specified sites, episode of care unspecified |

Continued on next page

**Table22 – continued from previous page**

| CodeSystem | ConceptCode | ConceptName |
|---|---|---|
| ICD9CM | 410.81 | Acute myocardial infarction of other specified sites, initial episode of care |
| ICD9CM | 410.82 | Acute myocardial infarction of other specified sites, subsequent episode of care |
| ICD9CM | 410.90 | Acute myocardial infarction of unspecified site, episode of care unspecified |
| ICD9CM | 410.91 | Acute myocardial infarction of unspecified site, initial episode of care |
| ICD9CM | 410.92 | Acute myocardial infarction of unspecified site, subsequent episode of care |
| SNOMED CT | 10273003 | Acute infarction of papillary muscle |
| SNOMED CT | 15990001 | Acute myocardial infarction of posterolateral wall |
| SNOMED CT | 22298006 | Myocardial infarction |
| SNOMED CT | 30277009 | Rupture of ventricle due to acute myocardial infarction |
| SNOMED CT | 32574007 | Past myocardial infarction diagnosed on electrocardiogram AND/OR other special investigation, but currently presenting no symptoms |
| SNOMED CT | 194802003 | True posterior myocardial infarction |
| SNOMED CT | 194809007 | Acute myocardial infarction of atrium |
| SNOMED CT | 194856005 | Subsequent myocardial infarction |
| SNOMED CT | 304914007 | Acute Q wave myocardial infarction |
| SNOMED CT | 307140009 | Acute non-Q wave infarction |
| SNOMED CT | 314207007 | Non-Q wave myocardial infarction |
| SNOMED CT | 394710008 | First myocardial infarction |
| SNOMED CT | 233835003 | Acute widespread myocardial infarction |
| SNOMED CT | 233838001 | Acute posterior myocardial infarction |
| SNOMED CT | 233843008 | Silent myocardial infarction |
| SNOMED CT | 42531007 | Microinfarct of heart |
| SNOMED CT | 52035003 | Acute anteroapical myocardial infarction |
| SNOMED CT | 54329005 | Acute myocardial infarction of anterior wall |
| SNOMED CT | 57054005 | Acute myocardial infarction |
| SNOMED CT | 58612006 | Acute myocardial infarction of lateral wall |
| SNOMED CT | 62695002 | Acute anteroseptal myocardial infarction |
| SNOMED CT | 65547006 | Acute myocardial infarction of inferolateral wall |
| SNOMED CT | 70211005 | Acute myocardial infarction of anterolateral wall |
| SNOMED CT | 70422006 | Acute subendocardial infarction |
| SNOMED CT | 73795002 | Acute myocardial infarction of inferior wall |
| SNOMED CT | 79009004 | Acute myocardial infarction of septum |
| SNOMED CT | 401303003 | Acute ST segment elevation myocardial infarction |
| SNOMED CT | 401314000 | Acute non-ST segment elevation myocardial infarction |
| SNOMED CT | 418044006 | Myocardial infarction in recovery phase |
| SNOMED CT | 428196007 | Mixed myocardial ischemia and infarction |
| SNOMED CT | 428752002 | Recent myocardial infarction |

Continued on next page

**Table22 – continued from previous page**

| CodeSystem | ConceptCode | ConceptName |
|---|---|---|
| SNOMED CT | 129574000 | Postoperative myocardial infarction |
| SNOMED CT | 1208872002 | Subsequent anterior non-ST segment elevation myocardial infarction |
| SNOMED CT | 1208873007 | Subsequent inferior non-ST segment elevation myocardial infarction |
| SNOMED CT | 1163440003 | Postoperative acute myocardial infarction |
| SNOMED CT | 1755008 | Old myocardial infarction |
| SNOMED CT | 282006 | Acute myocardial infarction of basal-lateral wall |
| SNOMED CT | 836293000 | Acute myocardial infarction of right ventricle |
| SNOMED CT | 836294006 | Acute myocardial infarction of apex of heart |
| SNOMED CT | 836295007 | Acute myocardial infarction of inferolateral wall with posterior extension |
| SNOMED CT | 840309000 | Acute ST segment elevation myocardial infarction due to proximal left anterior descending coronary artery occlusion |
| SNOMED CT | 840312002 | Acute ST segment elevation myocardial infarction due to mid left anterior descending coronary artery occlusion |
| SNOMED CT | 840316004 | Acute ST segment elevation myocardial infarction due to distal left anterior descending coronary artery occlusion |
| SNOMED CT | 840609007 | Acute ST segment elevation myocardial infarction due to occlusion of anterior descending branch of left coronary artery |
| SNOMED CT | 840680009 | Acute ST segment elevation myocardial infarction due to occlusion of septal branch of anterior descending branch of left coronary artery |
| SNOMED CT | 846668006 | Acute ST segment elevation myocardial infarction due to occlusion of diagonal branch of anterior descending branch of left coronary artery |
| SNOMED CT | 846683001 | Acute ST segment elevation myocardial infarction due to occlusion of intermediate artery |
| SNOMED CT | 868214006 | Acute ST segment elevation myocardial infarction due to occlusion of proximal portion of right coronary artery |
| SNOMED CT | 868217004 | Acute ST segment elevation myocardial infarction due to occlusion of distal portion of right coronary artery |
| SNOMED CT | 868220007 | Acute ST segment elevation myocardial infarction due to occlusion of mid portion of right coronary artery |
| SNOMED CT | 868224003 | Acute ST segment elevation myocardial infarction due to occlusion of marginal branch of right coronary artery |
| SNOMED CT | 868225002 | Acute ST segment elevation myocardial infarction due to occlusion of posterior descending branch of right coronary artery |
| SNOMED CT | 868226001 | Acute ST segment elevation myocardial infarction due to occlusion of posterior lateral branch of right coronary artery |
| SNOMED CT | 17531000119105 | Acute myocardial infarction due to left coronary artery occlusion |

Continued on next page

**Table22 – continued from previous page**

| CodeSystem | ConceptCode | ConceptName |
|---|---|---|
| SNOMED CT | 23311000119105 | Acute myocardial infarction due to right coronary artery occlusion |
| SNOMED CT | 285981000119103 | Acute ST segment elevation myocardial infarction involving left anterior descending coronary artery |
| SNOMED CT | 380001000004106 | Subendocardial myocardial infarction |
| SNOMED CT | 12238111000119106 | Acute ST segment elevation myocardial infarction of inferolateral wall |
| SNOMED CT | 12238151000119107 | Acute ST segment elevation myocardial infarction of inferoposterior wall |
| SNOMED CT | 15712841000119100 | Acute ST segment elevation myocardial infarction of posterolateral wall |
| SNOMED CT | 15712881000119105 | Acute ST segment elevation myocardial infarction of anterolateral wall |
| SNOMED CT | 15712921000119103 | Acute ST segment elevation myocardial infarction of lateral wall |
| SNOMED CT | 15712961000119108 | Acute ST segment elevation myocardial infarction of anteroseptal wall |
| SNOMED CT | 15713041000119103 | Acute ST segment elevation myocardial infarction of posterior wall |
| SNOMED CT | 15713161000119100 | Acute ST segment elevation myocardial infarction of septum |
| SNOMED CT | 15713201000119105 | Acute ST segment elevation myocardial infarction of posterobasal wall |
| SNOMED CT | 15962541000119106 | Acute ST segment elevation myocardial infarction of anteroapical wall |
| SNOMED CT | 15963181000119104 | Acute ST segment elevation myocardial infarction due to occlusion of circumflex coronary artery |
| SNOMED CT | 16837681000119104 | Myocardial infarction due to demand ischemia |
| SNOMED CT | 15713081000119108 | Acute ST segment elevation myocardial infarction involving left main coronary artery |
| SNOMED CT | 15713121000119105 | Acute STEMI (ST elevation myocardial infarction) due to RCA (right coronary artery) occlusion |
| SNOMED CT | 703164000 | Acute anterior ST segment elevation myocardial infarction |
| SNOMED CT | 703165004 | Acute ST segment elevation myocardial infarction of anterior wall involving right ventricle |
| SNOMED CT | 703209002 | Subsequent ST segment elevation myocardial infarction of inferior wall |
| SNOMED CT | 703210007 | Subsequent ST segment elevation myocardial infarction of anterior wall |
| SNOMED CT | 703211006 | Subsequent ST segment elevation myocardial infarction |
| SNOMED CT | 703212004 | Acute myocardial infarction during procedure |
| SNOMED CT | 703213009 | Acute ST segment elevation myocardial infarction of inferior wall |

Continued on next page

**Table22 – continued from previous page**

| CodeSystem | ConceptCode | ConceptName |
|---|---|---|
| SNOMED CT | 703251009 | Acute myocardial infarction of inferior wall involving right ventricle |
| SNOMED CT | 703252002 | Acute myocardial infarction of anterior wall involving right ventricle |
| SNOMED CT | 703253007 | Acute ST segment elevation myocardial infarction of inferior wall involving right ventricle |
| SNOMED CT | 703360004 | Subsequent non-ST segment elevation myocardial infarction |
| SNOMED CT | 194857001 | Subsequent myocardial infarction of anterior wall |
| SNOMED CT | 194858006 | Subsequent myocardial infarction of inferior wall |
| SNOMED CT | 59063002 | Acute myocardial infarction of apical-lateral wall |
| SNOMED CT | 64627002 | Acute myocardial infarction of high lateral wall |
| SNOMED CT | 70998009 | Acute myocardial infarction of posterobasal wall |
| SNOMED CT | 76593002 | Acute myocardial infarction of inferoposterior wall |
| SNOMED CT | 311792005 | Postoperative transmural myocardial infarction of anterior wall |
| SNOMED CT | 311793000 | Postoperative transmural myocardial infarction of inferior wall |
| SNOMED CT | 311796008 | Postoperative subendocardial myocardial infarction |
| SNOMED CT | 233825009 | Acute Q wave infarction - anteroseptal |
| SNOMED CT | 233826005 | Acute non-Q wave infarction - anteroseptal |
| SNOMED CT | 233827001 | Acute Q wave infarction - anterolateral |
| SNOMED CT | 233828006 | Acute non-Q wave infarction - anterolateral |
| SNOMED CT | 233829003 | Acute Q wave infarction - inferior |
| SNOMED CT | 233830008 | Acute non-Q wave infarction - inferior |
| SNOMED CT | 233831007 | Acute Q wave infarction - inferolateral |
| SNOMED CT | 233832000 | Acute non-Q wave infarction - inferolateral |
| SNOMED CT | 233833005 | Acute Q wave infarction - lateral |
| SNOMED CT | 233834004 | Acute non-Q wave infarction - lateral |
| SNOMED CT | 233836002 | Acute Q wave infarction - widespread |
| SNOMED CT | 233837006 | Acute non-Q wave infarction - widespread |
| SNOMED CT | 233839009 | Old anterior myocardial infarction |
| SNOMED CT | 233840006 | Old inferior myocardial infarction |
| SNOMED CT | 233841005 | Old lateral myocardial infarction |
| SNOMED CT | 233842003 | Old posterior myocardial infarction |
| SNOMED CT | 726499301000119105 | Myocardial infarction due to atherothrombotic coronary artery disease |
| SNOMED CT | 879955009 | Myocardial infarction with non-obstructive coronary artery |
| SNOMED CT | 896689003 | Acute myocardial infarction due to occlusion of circumflex branch of left coronary artery |
| SNOMED CT | 896691006 | Acute ST segment elevation myocardial infarction due to occlusion of circumflex branch of left coronary artery |
| SNOMED CT | 896696001 | Acute ST segment elevation myocardial infarction of apex of heart |

Continued on next page

**Table22 – continued from previous page**

| CodeSystem | ConceptCode | ConceptName |
|---|---|---|
| SNOMED CT | 896697005 | Acute ST segment elevation myocardial infarction of right ventricle |
| SNOMED CT | 1204151009 | Acute inferior non-ST segment elevation myocardial infarction of right ventricle |
| SNOMED CT | 1204154001 | Acute anterior non-ST segment elevation myocardial infarction with right ventricular involvement |
| SNOMED CT | 1204155000 | Acute anterior non-ST segment elevation myocardial infarction |
| SNOMED CT | 1204152002 | Acute inferior non-ST segment elevation myocardial infarction |
| SNOMED CT | 1204222000 | Acute non-ST segment elevation myocardial infarction of right ventricle |

###### 8.4.13 Osteoporosis

Table 23: Concept codes used to identify if a patient was diagnosed with osteoporosis.

| CodeSystem | ConceptCode | ConceptName |
|---|---|---|
| ICD10CM | M81.6 | Localized osteoporosis [Lequesne] |
| ICD10CM | M81.0 | Age-related osteoporosis without current pathological fracture |
| ICD10CM | M81.8 | Other osteoporosis without current pathological fracture |
| ICD9CM | 733.01 | Senile osteoporosis |
| ICD9CM | 733.02 | Idiopathic osteoporosis |
| ICD9CM | 733.03 | Disuse osteoporosis |
| ICD9CM | 733.09 | Other osteoporosis |
| SNOMED CT | 1515008 | Massive osteolysis |
| SNOMED CT | 18040001 | Type II osteoporosis |
| SNOMED CT | 102447009 | Postmenopausal osteoporosis |
| SNOMED CT | 109346000 | Osteoporotic bone marrow defect |
| SNOMED CT | 3345002 | Idiopathic osteoporosis |
| SNOMED CT | 14651005 | Drug-induced osteoporosis |
| SNOMED CT | 32369003 | Menopausal osteoporosis |
| SNOMED CT | 699528002 | Transient osteoporosis |
| SNOMED CT | 703264005 | Secondary osteoporosis |
| SNOMED CT | 735617003 | Premenopausal idiopathic osteoporosis |
| SNOMED CT | 735618008 | Osteoporosis due to malabsorption |
| SNOMED CT | 739301006 | Osteoporosis co-occurrent and due to multiple myeloma |
| SNOMED CT | 203429007 | Idiopathic generalized osteoporosis |
| SNOMED CT | 203433000 | Postoophorectomy osteoporosis |

Continued on next page

**Table23 – continued from previous page**

| CodeSystem | ConceptCode | ConceptName |
|---|---|---|
| SNOMED CT | 203434006 | Post-surgical malabsorption osteoporosis |
| SNOMED CT | 203435007 | Localized osteoporosis - Lequesne |
| SNOMED CT | 203437004 | Osteoporosis in endocrine disorders |
| SNOMED CT | 203438009 | Vertebral osteoporosis |
| SNOMED CT | 203444008 | Postoophorectomy osteoporosis with pathological fracture |
| SNOMED CT | 203446005 | Post-surgical malabsorption osteoporosis with pathological fracture |
| SNOMED CT | 203657009 | Osteoporotic kyphosis |
| SNOMED CT | 53174001 | Disuse osteoporosis |
| SNOMED CT | 64859006 | Osteoporosis |
| SNOMED CT | 268028001 | Localized disuse osteoporosis |
| SNOMED CT | 276661002 | Primary osteoporosis |
| SNOMED CT | 281387004 | Regional osteoporosis |
| SNOMED CT | 311806008 | Osteoporotic collapse of cervical vertebra |
| SNOMED CT | 390833005 | Osteoporosis due to corticosteroid |
| SNOMED CT | 240155001 | Adult idiopathic generalized osteoporosis |
| SNOMED CT | 240157009 | Secondary generalized osteoporosis |
| SNOMED CT | 240158004 | Regional migrating osteoporosis |
| SNOMED CT | 240159007 | Transient osteoporosis of hip |
| SNOMED CT | 240161003 | Disappearing bone disease |
| SNOMED CT | 240162005 | Secondary localized osteoporosis |
| SNOMED CT | 240164006 | Post-irradiation osteoporosis |
| SNOMED CT | 240198002 | Osteoporotic vertebral collapse |
| SNOMED CT | 454201000124109 | Pathological fracture of left ulna due to osteoporosis |
| SNOMED CT | 11304761000119103 | Pathological fracture of right tibia due to osteoporosis |
| SNOMED CT | 11305081000119109 | Pathological fracture of right fibula due to osteoporosis |
| SNOMED CT | 11305121000119106 | Pathological fracture of left fibula due to osteoporosis |
| SNOMED CT | 11305721000119107 | Pathological fracture of right humerus due to secondary osteoporosis |
| SNOMED CT | 11305761000119102 | Pathological fracture of left humerus due to secondary osteoporosis |
| SNOMED CT | 11305801000119105 | Pathological fracture of right clavicle due to osteoporosis |
| SNOMED CT | 11305841000119107 | Pathological fracture of left clavicle due to osteoporosis |
| SNOMED CT | 11306041000119109 | Pathological fracture of right clavicle due to secondary osteoporosis |
| SNOMED CT | 11309321000119105 | Pathological fracture of right foot due to osteoporosis |
| SNOMED CT | 11309361000119100 | Pathological fracture of left foot due to osteoporosis |
| SNOMED CT | 11309641000119108 | Pathological fracture of right foot due to secondary osteoporosis |
| SNOMED CT | 11309681000119103 | Pathological fracture of left foot due to secondary osteoporosis |
| SNOMED CT | 11309801000119107 | Pathological fracture of right femur due to osteoporosis |
| SNOMED CT | 11309841000119109 | Pathological fracture of left femur due to osteoporosis |

Continued on next page

**Table23 – continued from previous page**

| CodeSystem | ConceptCode | ConceptName |
|---|---|---|
| SNOMED CT | 11310121000119102 | Pathological fracture of right femur due to secondary osteoporosis |
| SNOMED CT | 11310161000119107 | Pathological fracture of left femur due to secondary osteoporosis |
| SNOMED CT | 11310201000119102 | Pathological fracture of right ankle due to osteoporosis |
| SNOMED CT | 11310241000119100 | Pathological fracture of left ankle due to osteoporosis |
| SNOMED CT | 11311361000119103 | Secondary osteoporotic fracture of thoracic vertebra |
| SNOMED CT | 11311441000119109 | Secondary osteoporotic fracture of lumbar vertebra |
| SNOMED CT | 11311521000119104 | Secondary osteoporotic fracture of cervical vertebra |
| SNOMED CT | 11311601000119109 | Osteoporotic fracture of vertebra |
| SNOMED CT | 11311641000119106 | Osteoporotic fracture of sacral vertebra |
| SNOMED CT | 11311801000119105 | Secondary osteoporotic fracture of vertebra |
| SNOMED CT | 11311841000119107 | Secondary osteoporotic fracture of sacral vertebra |
| SNOMED CT | 11312241000119101 | Pathological fracture of right hand due to osteoporosis |
| SNOMED CT | 11312281000119106 | Pathological fracture of left hand due to osteoporosis |
| SNOMED CT | 11312721000119102 | Pathological fracture of right radius due to osteoporosis |
| SNOMED CT | 11312761000119107 | Pathological fracture of left radius due to osteoporosis |
| SNOMED CT | 11313281000119104 | Pathological fracture of right scapula due to osteoporosis |
| SNOMED CT | 11313321000119109 | Pathological fracture of left scapula due to osteoporosis |
| SNOMED CT | 11314841000119105 | Secondary osteoporotic fracture |
| SNOMED CT | 11314881000119100 | Pathological fracture of right hip due to osteoporosis |
| SNOMED CT | 11314921000119107 | Pathological fracture of left hip due to osteoporosis |
| SNOMED CT | 11315121000119108 | Pathological fracture of proximal end of right femur due to secondary osteoporosis |
| SNOMED CT | 11315161000119103 | Pathological fracture of left hip due to secondary osteoporosis |
| SNOMED CT | 11874341000119106 | Osteoporotic fracture of left rib |
| SNOMED CT | 11874381000119101 | Osteoporotic fracture of right rib |
| SNOMED CT | 16280471000119103 | Pathological fracture of left tibia due to osteoporosis |
| SNOMED CT | 11305481000119104 | Osteoporotic fracture of right humerus |
| SNOMED CT | 11305521000119104 | Osteoporotic fracture of left humerus |
| SNOMED CT | 11312681000119108 | Osteoporotic fracture of right ulna |
| SNOMED CT | 704282002 | Secondary osteoporotic fracture of ulna |
| SNOMED CT | 704283007 | Secondary osteoporotic fracture of tibia |
| SNOMED CT | 704284001 | Secondary osteoporotic fracture of scapula |
| SNOMED CT | 704285000 | Secondary osteoporotic fracture of radius |
| SNOMED CT | 704286004 | Secondary osteoporotic fracture of humerus |
| SNOMED CT | 704287008 | Secondary osteoporotic fracture of proximal femur |
| SNOMED CT | 704288003 | Secondary osteoporotic fracture of hand |
| SNOMED CT | 704289006 | Secondary osteoporotic fracture of foot |
| SNOMED CT | 704290002 | Secondary osteoporotic fracture of fibula |
| SNOMED CT | 704291003 | Secondary osteoporotic fracture of femur |
| SNOMED CT | 704292005 | Secondary osteoporotic fracture of clavicle |

Continued on next page

**Table23 – continued from previous page**

| CodeSystem | ConceptCode | ConceptName |
|---|---|---|
| SNOMED CT | 704293000 | Secondary osteoporotic fracture of ankle |
| SNOMED CT | 704328003 | Osteoporotic fracture of ankle |
| SNOMED CT | 704329006 | Osteoporotic fracture of clavicle |
| SNOMED CT | 704330001 | Osteoporotic fracture of femur |
| SNOMED CT | 704331002 | Osteoporotic fracture of fibula |
| SNOMED CT | 704332009 | Osteoporotic fracture of bone of foot |
| SNOMED CT | 704333004 | Osteoporotic fracture of hand |
| SNOMED CT | 704334005 | Osteoporotic fracture of proximal femur |
| SNOMED CT | 704335006 | Osteoporotic fracture of humerus |
| SNOMED CT | 704336007 | Osteoporotic fracture of radius |
| SNOMED CT | 704337003 | Osteoporotic fracture of scapula |
| SNOMED CT | 704338008 | Osteoporotic fracture of tibia |
| SNOMED CT | 704339000 | Osteoporotic fracture of ulna |
| SNOMED CT | 707419009 | Osteoporosis due to cystic fibrosis |
| SNOMED CT | 203445009 | Osteoporosis of disuse with pathological fracture |
| SNOMED CT | 203447001 | Drug-induced osteoporosis with pathological fracture |
| SNOMED CT | 203448006 | Idiopathic osteoporosis with pathological fracture |
| SNOMED CT | 203450003 | Osteoporotic fracture of lumbar vertebra |
| SNOMED CT | 203451004 | Osteoporotic fracture of thoracic vertebra |
| SNOMED CT | 203452006 | Osteoporotic fracture of cervical vertebra |
| SNOMED CT | 203453001 | Postmenopausal osteoporosis with pathological fracture |
| SNOMED CT | 311890007 | Osteoporotic collapse of lumbar vertebra |
| SNOMED CT | 311891006 | Osteoporotic collapse of thoracic vertebra |
| SNOMED CT | 240154002 | Idiopathic osteoporosis in pregnancy |
| SNOMED CT | 240156000 | Juvenile idiopathic generalized osteoporosis |
| SNOMED CT | 240160002 | Transient osteoporosis of hip in pregnancy |
| SNOMED CT | 15743005 | Posttraumatic osteoporosis |
| SNOMED CT | 897308007 | Chronic kidney disease with osteoporosis |
| SNOMED CT | 431031001 | Osteoporotic fracture of neck of femur |
| SNOMED CT | 443165006 | Osteoporotic fracture |

#### 8.5 Anti-Diabetic Medications (ADMs)

##### 8.5.1 Metformin

Table 24: Concept codes used to identify if a patient was prescribed metformin.

| CodeSystem | ConceptCode | ConceptName |
|---|---|---|
| RxNorm | 6809 | metformin |
| RxNorm | 351269 | Metformin 500 MG / rosiglitazone 2 MG Oral Tablet |
| RxNorm | 349554 | Metformin 500 MG / rosiglitazone 4 MG Oral Tablet |

Continued on next page

**Table24 – continued from previous page**

| CodeSystem | ConceptCode | ConceptName |
|---|---|---|
| RxNorm | 351268 | Metformin 500 MG / rosiglitazone 1 MG Oral Tablet |
| RxNorm | 541765 | Metformin 1000 MG Extended Release Tablet |
| RxNorm | 284433 | Glyburide 5 MG / Metformin 500 MG Oral Tablet [Glucovance] |
| RxNorm | 311572 | Metformin 850 MG Oral Tablet |
| RxNorm | 311571 | Metformin 500 MG Extended Release Tablet |
| RxNorm | 213454 | Metformin 1000 MG Oral Tablet [Glucophage] |
| RxNorm | 284432 | Glyburide 2.5 MG / Metformin 500 MG Oral Tablet [Glucovance] |
| RxNorm | 647242 | Metformin 500 MG Extended Release Tablet [Glumetza] |
| RxNorm | 541811 | Metformin 500 MG Extended Release Tablet [Fortamet] |
| RxNorm | 828382 | Metformin 750 MG Extended Release Tablet [Glucophage] |
| RxNorm | 316257 | Metformin 850 MG |
| RxNorm | 541769 | Metformin 1000 MG Extended Release Tablet [Fortamet] |
| RxNorm | 351273 | Glipizide 2.5 MG / Metformin 500 MG Oral Tablet |
| RxNorm | 311569 | Metformin 1000 MG Oral Tablet |
| RxNorm | 358336 | Metformin 500 MG 24 Hour Extended Release Tablet |
| RxNorm | 351275 | Glipizide 2.5 MG / Metformin 250 MG Oral Tablet |
| RxNorm | 284431 | Glyburide 1.25 MG / Metformin 250 MG Oral Tablet [Glucovance] |
| RxNorm | 731426 | Metformin 850 MG / pioglitazone 15 MG Oral Tablet [Actoplus Met 15/850] |
| RxNorm | 351274 | Glipizide 5 MG / Metformin 500 MG Oral Tablet |
| RxNorm | 731443 | Metformin 500 MG / pioglitazone 15 MG Oral Tablet [Actoplus Met 15/500] |
| RxNorm | 574869 | Glyburide 1.25 MG / Metformin 250 MG [Glucovance] |
| RxNorm | 566228 | Metformin 500 MG [Orabet] |
| RxNorm | 566230 | Metformin 850 MG [Orabet] |
| RxNorm | 574871 | Glyburide 5 MG / Metformin 500 MG [Glucovance] |
| RxNorm | 403819 | Metformin 750 MG Extended Release Tablet |
| RxNorm | 403968 | Metformin 100 MG/ML Oral Solution |
| RxNorm | 403915 | Metformin 1000 MG / rosiglitazone 2 MG Oral Tablet |
| RxNorm | 541767 | Metformin 1000 MG [Fortamet] |
| RxNorm | 828303 | Metformin 500 MG Extended Release Tablet [Glucophage] |
| RxNorm | 404727 | Metformin 100 MG/ML Oral Solution [Riomet] |
| RxNorm | 574870 | Glyburide 2.5 MG / Metformin 500 MG [Glucovance] |
| RxNorm | 406061 | Metformin 750 MG |
| RxNorm | 403916 | Metformin 1000 MG / rosiglitazone 4 MG Oral Tablet |
| RxNorm | 406081 | Metformin 100 MG/ML |
| RxNorm | 577095 | Metformin 850 MG / pioglitazone 15 MG Oral Tablet |
| RxNorm | 577094 | Metformin 500 MG / pioglitazone 15 MG Oral Tablet |
| RxNorm | 583193 | Metformin 850 MG [Metforming] |
| RxNorm | 576612 | Metformin 100 MG/ML [Riomet] |

Continued on next page

**Table24 – continued from previous page**

| CodeSystem | ConceptCode | ConceptName |
|---|---|---|
| RxNorm | 654546 | Metformin 625 MG Oral Tablet |
| RxNorm | 731441 | Metformin 500 MG / pioglitazone 15 MG [Actoplus Met 15/500] |
| RxNorm | 654545 | Metformin 625 MG |
| RxNorm | 310538 | Glyburide 5 MG / Metformin 500 MG Oral Tablet |
| RxNorm | 700518 | Metformin 1000 MG / sitagliptin 50 MG Oral Tablet |
| RxNorm | 645110 | Metformin 500 MG [Glumetza] |
| RxNorm | 731424 | Metformin 850 MG / pioglitazone 15 MG [Actoplus Met 15/850] |
| RxNorm | 700517 | Metformin 500 MG / sitagliptin 50 MG Oral Tablet |
| RxNorm | 310535 | Glyburide 2.5 MG / Metformin 500 MG Oral Tablet |
| RxNorm | 310533 | Glyburide 1.25 MG / Metformin 250 MG Oral Tablet |
| RxNorm | 566229 | Metformin 500 MG [Glucophage] |
| RxNorm | 311570 | Metformin 500 MG Oral Tablet |
| RxNorm | 573342 | Metformin 1000 MG [Glucophage] |
| RxNorm | 566231 | Metformin 850 MG [Glucophage] |
| RxNorm | 757606 | Metformin 500 MG / sitagliptin 50 MG [Janumet 50 mg/500 mg] |
| RxNorm | 204046 | Metformin 500 MG Oral Tablet [Glucophage] |
| RxNorm | 204048 | Metformin 850 MG Oral Tablet [Glucophage] |
| RxNorm | 476506 | Metformin 1000 MG 24 Hour Extended Release Tablet |
| RxNorm | 757602 | Metformin 1000 MG / sitagliptin 50 MG [Janumet 50 mg/1000 mg] |
| RxNorm | 757608 | Metformin 500 MG / sitagliptin 50 MG Oral Tablet [Janumet 50 mg/500 mg] |
| RxNorm | 757604 | Metformin 1000 MG / sitagliptin 50 MG Oral Tablet [Janumet 50 mg/1000 mg] |
| RxNorm | 541810 | Metformin 500 MG [Fortamet] |
| RxNorm | 802743 | Metformin 500 MG / repaglinide 1 MG Oral Tablet |
| RxNorm | 802050 | Metformin 750 MG [Glucophage] |
| RxNorm | 801708 | Metformin 1000 MG Extended Release Tablet [Glumetza] |
| RxNorm | 564040 | Metformin 850 MG [Glucamet] |
| RxNorm | 805669 | Metformin 500 MG / repaglinide 1 MG [PrandiMet 1/500] |
| RxNorm | 564039 | Metformin 500 MG [Glucamet] |
| RxNorm | 801707 | Metformin 1000 MG [Glumetza] |
| RxNorm | 805673 | Metformin 500 MG / repaglinide 2 MG [PrandiMet 2/500] |
| RxNorm | 806290 | Metformin 500 MG / rosiglitazone 4 MG [Avandamet 4/500] |
| RxNorm | 805671 | Metformin 500 MG / repaglinide 1 MG Oral Tablet [PrandiMet 1/500] |
| RxNorm | 806288 | Metformin 1000 MG / rosiglitazone 2 MG Oral Tablet [Avandamet 2/1000] |
| RxNorm | 806296 | Metformin 500 MG / rosiglitazone 2 MG Oral Tablet [Avandamet 2/500] |

Continued on next page

**Table24 – continued from previous page**

| CodeSystem | ConceptCode | ConceptName |
|---|---|---|
| RxNorm | 806276 | Metformin 500 MG / rosiglitazone 1 MG Oral Tablet [Avandamet 1/500] |
| RxNorm | 806284 | Metformin 1000 MG / rosiglitazone 4 MG Oral Tablet [Avandamet 4/1000] |
| RxNorm | 806294 | Metformin 500 MG / rosiglitazone 2 MG [Avandamet 2/500] |
| RxNorm | 805675 | Metformin 500 MG / repaglinide 2 MG Oral Tablet [PrandiMet 2/500] |
| RxNorm | 806292 | Metformin 500 MG / rosiglitazone 4 MG Oral Tablet [Avandamet 4/500] |
| RxNorm | 806282 | Metformin 1000 MG / rosiglitazone 4 MG [Avandamet 4/1000] |
| RxNorm | 806286 | Metformin 1000 MG / rosiglitazone 2 MG [Avandamet 2/1000] |
| RxNorm | 802744 | Metformin 500 MG / repaglinide 2 MG Oral Tablet |
| RxNorm | 806274 | Metformin 500 MG / rosiglitazone 1 MG [Avandamet 1/500] |
| RxNorm | 1243830 | Metformin / sitagliptin Extended Release Tablet [Janumet 100/1000] |
| RxNorm | 1243847 | Metformin / sitagliptin Extended Release Tablet [Janumet 50/500] |
| RxNorm | 1243023 | Linagliptin / Metformin Oral Tablet [Jentadueto 2.5/1000] |
| RxNorm | 1243030 | Linagliptin / Metformin Oral Tablet [Jentadueto 2.5/500] |
| RxNorm | 340678 | Metformin 850 MG Extended Release Tablet |
| RxNorm | 368567 | Glipizide / Metformin Oral Tablet [metaglip] |
| RxNorm | 352301 | Glipizide 5 MG / Metformin 500 MG Oral Tablet [metaglip] |
| RxNorm | 352302 | Glipizide 2.5 MG / Metformin 250 MG Oral Tablet [metaglip] |
| RxNorm | 368169 | Glyburide / Metformin Oral Tablet [Glucovance] |
| RxNorm | 352300 | Glipizide 2.5 MG / Metformin 500 MG Oral Tablet [metaglip] |
| RxNorm | 368568 | Metformin / rosiglitazone Oral Tablet [avandamet] |
| RxNorm | 446358 | Metformin 12 hour Extended Release Tablet |
| RxNorm | 404646 | Metformin 1000 MG / rosiglitazone 2 MG Oral Tablet [avandamet] |
| RxNorm | 352296 | Metformin 500 MG / rosiglitazone 4 MG Oral Tablet [avandamet] |
| RxNorm | 404647 | Metformin 1000 MG / rosiglitazone 4 MG Oral Tablet [avandamet] |
| RxNorm | 352294 | Metformin 500 MG / rosiglitazone 1 MG Oral Tablet [avandamet] |
| RxNorm | 375561 | Metformin 24 Hour Extended Release Tablet |
| RxNorm | 369995 | Metformin Extended Release Tablet [Glucophage XR] |
| RxNorm | 284467 | Metformin 500 MG Extended Release Tablet [Glucophage XR] |

Continued on next page

**Table24 – continued from previous page**

| CodeSystem | ConceptCode | ConceptName |
|---|---|---|
| RxNorm | 433928 | Metformin 750 MG 24 Hour Extended Release Tablet |
| RxNorm | 404477 | Metformin 750 MG Extended Release Tablet [Glucophage XR] |
| RxNorm | 1008476 | metformin / ropinirole |
| RxNorm | 1043561 | metformin / saxagliptin Extended Release Oral Tablet |
| RxNorm | 1043562 | metformin / saxagliptin |
| RxNorm | 1043563 | 24 HR metformin hydrochloride 1000 MG / saxagliptin 2.5 MG Extended Release Oral Tablet |
| RxNorm | 1043565 | metformin hydrochloride 1000 MG / saxagliptin 2.5 MG [Kombiglyze] |
| RxNorm | 1043566 | metformin / saxagliptin Extended Release Oral Tablet [Kombiglyze] |
| RxNorm | 1043567 | 24 HR metformin hydrochloride 1000 MG / saxagliptin 2.5 MG Extended Release Oral Tablet [Kombiglyze] |
| RxNorm | 1043568 | metformin hydrochloride 1000 MG / saxagliptin 2.5 MG Extended Release Oral Tablet |
| RxNorm | 1043569 | metformin hydrochloride 1000 MG / saxagliptin 2.5 MG Extended Release Oral Tablet [Kombiglyze] |
| RxNorm | 1043570 | 24 HR metformin hydrochloride 1000 MG / saxagliptin 5 MG Extended Release Oral Tablet |
| RxNorm | 1043572 | metformin hydrochloride 1000 MG / saxagliptin 5 MG [Kombiglyze] |
| RxNorm | 1043574 | 24 HR metformin hydrochloride 1000 MG / saxagliptin 5 MG Extended Release Oral Tablet [Kombiglyze] |
| RxNorm | 1043575 | metformin hydrochloride 1000 MG / saxagliptin 5 MG Extended Release Oral Tablet |
| RxNorm | 1043576 | metformin hydrochloride 1000 MG / saxagliptin 5 MG Extended Release Oral Tablet [Kombiglyze] |
| RxNorm | 1043578 | 24 HR metformin hydrochloride 500 MG / saxagliptin 5 MG Extended Release Oral Tablet |
| RxNorm | 1043580 | metformin hydrochloride 500 MG / saxagliptin 5 MG [Kombiglyze] |
| RxNorm | 1043582 | 24 HR metformin hydrochloride 500 MG / saxagliptin 5 MG Extended Release Oral Tablet [Kombiglyze] |
| RxNorm | 1043583 | metformin hydrochloride 500 MG / saxagliptin 5 MG Extended Release Oral Tablet |
| RxNorm | 1043584 | metformin hydrochloride 500 MG / saxagliptin 5 MG Extended Release Oral Tablet [Kombiglyze] |
| RxNorm | 105376 | metformin hydrochloride 500 MG Oral Tablet [Glucamet] |
| RxNorm | 105377 | metformin hydrochloride 850 MG Oral Tablet [Glucamet] |
| RxNorm | 1156197 | glyburide / metformin Pill |
| RxNorm | 1155467 | chlorpropamide / metformin Oral Product |
| RxNorm | 1155468 | chlorpropamide / metformin Pill |

Continued on next page

**Table24 – continued from previous page**

| CodeSystem | ConceptCode | ConceptName |
|---|---|---|
| RxNorm | 1161597 | metformin / pioglitazone Oral Product |
| RxNorm | 1161598 | metformin / pioglitazone Pill |
| RxNorm | 1161599 | metformin / repaglinide Oral Product |
| RxNorm | 1161600 | metformin / repaglinide Pill |
| RxNorm | 1161601 | metformin / ropinirole Oral Product |
| RxNorm | 1161602 | metformin / ropinirole Pill |
| RxNorm | 1161603 | metformin / rosiglitazone Oral Product |
| RxNorm | 1161604 | metformin / rosiglitazone Pill |
| RxNorm | 1161605 | metformin / saxagliptin Oral Product |
| RxNorm | 1161606 | metformin / saxagliptin Pill |
| RxNorm | 1161607 | metformin / sitagliptin Oral Product |
| RxNorm | 1161608 | metformin / sitagliptin Pill |
| RxNorm | 1161609 | metformin Oral Liquid Product |
| RxNorm | 1161610 | metformin Oral Product |
| RxNorm | 1161611 | metformin Pill |
| RxNorm | 1165205 | glipizide / metformin Oral Product |
| RxNorm | 1165206 | glipizide / metformin Pill |
| RxNorm | 1165845 | glyburide / metformin Oral Product |
| RxNorm | 1171242 | Glucamet Oral Product |
| RxNorm | 1171243 | Glucamet Pill |
| RxNorm | 1171244 | Glucophage Oral Product |
| RxNorm | 1171245 | Glucophage Pill |
| RxNorm | 1171248 | Glucovance Oral Product |
| RxNorm | 1171249 | Glucovance Pill |
| RxNorm | 1171254 | Glumetza Oral Product |
| RxNorm | 1171255 | Glumetza Pill |
| RxNorm | 1169920 | Actoplus Met Oral Product |
| RxNorm | 1169923 | Actoplus Met Pill |
| RxNorm | 1175016 | Avandamet Oral Product |
| RxNorm | 1175021 | Avandamet Pill |
| RxNorm | 1172629 | Fortamet Oral Product |
| RxNorm | 1172630 | Fortamet Pill |
| RxNorm | 352295 | Metformin 500 MG / rosiglitazone 2 MG Oral Tablet [avandamet] |
| RxNorm | 575998 | Glipizide 2.5 MG / Metformin 250 MG [metaglip] |
| RxNorm | 574900 | Metformin 500 MG [Glucophage XR] |
| RxNorm | 575996 | Glipizide 2.5 MG / Metformin 500 MG [metaglip] |
| RxNorm | 541776 | Metformin 500 MG / ropinirole 2 MG [avandamet] |
| RxNorm | 575991 | Metformin 500 MG / rosiglitazone 1 MG [avandamet] |
| RxNorm | 575993 | Metformin 500 MG / rosiglitazone 4 MG [avandamet] |
| RxNorm | 575992 | Metformin 500 MG / rosiglitazone 2 MG [avandamet] |
| RxNorm | 576550 | Metformin 1000 MG / rosiglitazone 4 MG [avandamet] |
| RxNorm | 576428 | Metformin 750 MG [Glucophage XR] |

Continued on next page

**Table24 – continued from previous page**

| CodeSystem | ConceptCode | ConceptName |
|---|---|---|
| RxNorm | 541778 | Metformin 500 MG / ropinirole 2 MG Oral Tablet [avandamet] |
| RxNorm | 575997 | Glipizide 5 MG / Metformin 500 MG [metaglip] |
| RxNorm | 576549 | Metformin 1000 MG / rosiglitazone 2 MG [avandamet] |
| RxNorm | 602413 | Metformin / pioglitazone Oral Tablet [Actoplus Met] |
| RxNorm | 705136 | Metformin 1000 MG / sitagliptin 50 MG Oral Tablet [Janumet] |
| RxNorm | 602416 | Metformin 850 MG / pioglitazone 15 MG Oral Tablet [Actoplus Met] |
| RxNorm | 645111 | Metformin Oral Tablet [Glumetza] |
| RxNorm | 705134 | Metformin 1000 MG / sitagliptin 50 MG [Janumet] |
| RxNorm | 705135 | Metformin / sitagliptin Oral Tablet [Janumet] |
| RxNorm | 706114 | Metformin / rosiglitazone Oral Tablet [Avandamet] |
| RxNorm | 705138 | Metformin 500 MG / sitagliptin 50 MG Oral Tablet [Janumet] |
| RxNorm | 731425 | Metformin / pioglitazone Oral Tablet [Actoplus Met 15/850] |
| RxNorm | 748763 | Metformin 500 MG / sitagliptin 64.3 MG Oral Tablet |
| RxNorm | 602414 | Metformin 500 MG / pioglitazone 15 MG Oral Tablet [Actoplus Met] |
| RxNorm | 757607 | Metformin / sitagliptin Oral Tablet [Janumet 50 mg/500 mg] |
| RxNorm | 602412 | Metformin 500 MG / pioglitazone 15 MG [Actoplus Met] |
| RxNorm | 759753 | Metformin / rosiglitazone Oral Tablet [Adandamet 2/500] |
| RxNorm | 759754 | Metformin 500 MG / rosiglitazone 2 MG Oral Tablet [Adandamet 2/500] |
| RxNorm | 806291 | Metformin / rosiglitazone Oral Tablet [Avandamet 4/500] |
| RxNorm | 806295 | Metformin / rosiglitazone Oral Tablet [Avandamet 2/500] |
| RxNorm | 806283 | Metformin / rosiglitazone Oral Tablet [Avandamet 4/1000] |
| RxNorm | 748765 | Metformin 500 MG / sitagliptin 64.3 MG Oral Tablet [Janumet] |
| RxNorm | 805674 | Metformin / repaglinide Oral Tablet [PrandiMet 2/500] |
| RxNorm | 645112 | Metformin 500 MG Oral Tablet [Glumetza] |
| RxNorm | 705137 | Metformin 500 MG / sitagliptin 50 MG [Janumet] |
| RxNorm | 759752 | Metformin 500 MG / rosiglitazone 2 MG [Adandamet 2/500] |
| RxNorm | 602415 | Metformin 850 MG / pioglitazone 15 MG [Actoplus Met] |
| RxNorm | 849593 | Glipizide / Metformin Oral Tablet [Metaglip 2.5 MG/500 MG] |
| RxNorm | 748764 | Metformin 500 MG / sitagliptin 64.3 MG [Janumet] |
| RxNorm | 806275 | Metformin / rosiglitazone Oral Tablet [Avandamet 1/500] |
| RxNorm | 860996 | 24 HR Metformin hydrochloride 1000 MG Extended Release Oral Tablet |

Continued on next page

**Table24 – continued from previous page**

| CodeSystem | ConceptCode | ConceptName |
|---|---|---|
| RxNorm | 849589 | Glipizide / Metformin Oral Tablet [Metaglip 5 MG/500 MG] |
| RxNorm | 861746 | Glyburide / Metformin Oral Tablet [Glucovance 1.25 MG/250 MG] |
| RxNorm | 861751 | Glyburide / Metformin Oral Tablet [Glucovance 2.5 MG/500 MG] |
| RxNorm | 899999 | Metformin / pioglitazone Extended Release Tablet [Actoplus Met 30/1000] |
| RxNorm | 1043581 | Metformin / saxagliptin Extended Release Tablet [Kombiglyze 5/500] |
| RxNorm | 1043573 | Metformin / saxagliptin Extended Release Tablet [Kombiglyze 5/1000] |
| RxNorm | 1185627 | Metaglip 5 MG/500 MG Oral Product |
| RxNorm | 1243031 | Jentadueto 2.5/500 Oral Product |
| RxNorm | 1169924 | Actoplus Met 15/850 Oral Product |
| RxNorm | 1243025 | Jentadueto 2.5/1000 Pill |
| RxNorm | 1184630 | PrandiMet 2/500 Pill |
| RxNorm | 1185628 | Metaglip 5 MG/500 MG Pill |
| RxNorm | 1169921 | Actoplus Met 15/1000 Pill |
| RxNorm | 1175023 | Avandamet 4/1000 Pill |
| RxNorm | 1175018 | Avandamet 2/1000 Oral Product |
| RxNorm | 1169927 | Actoplus Met 30/1000 Pill |
| RxNorm | 1175019 | Avandamet 2/1000 Pill |
| RxNorm | 1175024 | Avandamet 4/500 Oral Product |
| RxNorm | 1169922 | Actoplus Met 15/500 Oral Product |
| RxNorm | 1173547 | Kombiglyze 5/500 Oral Product |
| RxNorm | 1185625 | Metaglip 2.5 MG/500 MG Oral Product |
| RxNorm | 1171251 | Glucovance 2.5 MG/500 MG Pill |
| RxNorm | 1172859 | Kombiglyze 2.5/1000 Oral Product |
| RxNorm | 1169926 | Actoplus Met 30/1000 Oral Product |
| RxNorm | 1173546 | Kombiglyze 5/1000 Pill |
| RxNorm | 1175025 | Avandamet 4/500 Pill |
| RxNorm | 1243832 | Janumet 100/1000 Pill |
| RxNorm | 1175022 | Avandamet 4/1000 Oral Product |
| RxNorm | 1184629 | PrandiMet 2/500 Oral Product |
| RxNorm | 1175020 | Avandamet 2/500 Oral Product |
| RxNorm | 1185626 | Metaglip 2.5 MG/500 MG Pill |
| RxNorm | 1175017 | Avandamet 1/500 Pill |
| RxNorm | 1171253 | Glucovance 5 MG/500 MG Pill |
| RxNorm | 1171250 | Glucovance 2.5 MG/500 MG Oral Product |
| RxNorm | 1167812 | Janumet 50/500 Oral Product |
| RxNorm | 1243032 | Jentadueto 2.5/500 Pill |
| RxNorm | 1169925 | Actoplus Met 15/850 Pill |

Continued on next page

**Table24 – continued from previous page**

| CodeSystem | ConceptCode | ConceptName |
|---|---|---|
| RxNorm | 1173548 | Kombiglyze 5/500 Pill |
| RxNorm | 1167813 | Janumet 50/500 Pill |
| RxNorm | 1171252 | Glucovance 5 MG/500 MG Oral Product |
| RxNorm | 1243024 | Jentadueto 2.5/1000 Oral Product |
| RxNorm | 1243831 | Janumet 100/1000 Oral Product |
| RxNorm | 1172860 | Kombiglyze Pill |
| RxNorm | 1172861 | Kombiglyze Oral Product |
| RxNorm | 1167810 | Janumet Oral Product |
| RxNorm | 1167811 | Janumet Pill |
| RxNorm | 1184627 | PrandiMet Oral Product |
| RxNorm | 1184628 | PrandiMet Pill |
| RxNorm | 1182890 | Orabet Metformin Oral Product |
| RxNorm | 1182891 | Orabet Metformin Pill |
| RxNorm | 1185049 | Metaglip Oral Product |
| RxNorm | 1185325 | Riomet Oral Liquid Product |
| RxNorm | 1185326 | Riomet Oral Product |
| RxNorm | 1185624 | Metaglip Pill |
| RxNorm | 1185653 | Metforming Oral Product |
| RxNorm | 1185654 | Metforming Pill |
| RxNorm | 1243016 | linagliptin / metformin Oral Product |
| RxNorm | 1243017 | linagliptin / metformin Pill |
| RxNorm | 1243018 | linagliptin / metformin Oral Tablet |
| RxNorm | 1243019 | linagliptin / metformin |
| RxNorm | 1243020 | linagliptin 2.5 MG / metformin hydrochloride 1000 MG Oral Tablet |
| RxNorm | 1243022 | linagliptin 2.5 MG / metformin hydrochloride 1000 MG [Jentadueto] |
| RxNorm | 1243026 | linagliptin 2.5 MG / metformin hydrochloride 1000 MG Oral Tablet [Jentadueto] |
| RxNorm | 1243027 | linagliptin 2.5 MG / metformin hydrochloride 500 MG Oral Tablet |
| RxNorm | 1243029 | linagliptin 2.5 MG / metformin hydrochloride 500 MG [Jentadueto] |
| RxNorm | 1243033 | linagliptin 2.5 MG / metformin hydrochloride 500 MG Oral Tablet [Jentadueto] |
| RxNorm | 1243034 | linagliptin 2.5 MG / metformin hydrochloride 850 MG Oral Tablet |
| RxNorm | 1243036 | linagliptin 2.5 MG / metformin hydrochloride 850 MG [Jentadueto] |
| RxNorm | 1243037 | linagliptin / metformin Oral Tablet [Jentadueto] |
| RxNorm | 1243038 | Jentadueto Oral Product |
| RxNorm | 1243039 | Jentadueto Pill |

Continued on next page

**Table24 – continued from previous page**

| CodeSystem | ConceptCode | ConceptName |
|---|---|---|
| RxNorm | 1243040 | linagliptin 2.5 MG / metformin hydrochloride 850 MG Oral Tablet [Jentadueto] |
| RxNorm | 1243826 | metformin / sitagliptin Extended Release Oral Tablet |
| RxNorm | 1243827 | 24 HR metformin hydrochloride 1000 MG / sitagliptin 100 MG Extended Release Oral Tablet |
| RxNorm | 1243829 | metformin hydrochloride 1000 MG / sitagliptin 100 MG [Janumet] |
| RxNorm | 1243833 | 24 HR metformin hydrochloride 1000 MG / sitagliptin 100 MG Extended Release Oral Tablet [Janumet] |
| RxNorm | 1243834 | metformin hydrochloride 1000 MG / sitagliptin 100 MG Extended Release Oral Tablet |
| RxNorm | 1243835 | metformin hydrochloride 1000 MG / sitagliptin 100 MG Extended Release Oral Tablet [Janumet] |
| RxNorm | 1243839 | metformin / sitagliptin Extended Release Oral Tablet [Janumet] |
| RxNorm | 1243842 | 24 HR metformin hydrochloride 1000 MG / sitagliptin 50 MG Extended Release Oral Tablet |
| RxNorm | 1243843 | 24 HR metformin hydrochloride 1000 MG / sitagliptin 50 MG Extended Release Oral Tablet [Janumet] |
| RxNorm | 1243844 | metformin hydrochloride 1000 MG / sitagliptin 50 MG Extended Release Oral Tablet |
| RxNorm | 1243845 | metformin hydrochloride 1000 MG / sitagliptin 50 MG Extended Release Oral Tablet [Janumet] |
| RxNorm | 1243846 | 24 HR metformin hydrochloride 500 MG / sitagliptin 50 MG Extended Release Oral Tablet |
| RxNorm | 1243848 | 24 HR metformin hydrochloride 500 MG / sitagliptin 50 MG Extended Release Oral Tablet [Janumet] |
| RxNorm | 1243849 | metformin hydrochloride 500 MG / sitagliptin 50 MG Extended Release Oral Tablet |
| RxNorm | 1243850 | metformin hydrochloride 500 MG / sitagliptin 50 MG Extended Release Oral Tablet [Janumet] |
| RxNorm | 1368381 | alogliptin / metformin Oral Product |
| RxNorm | 1368382 | alogliptin / metformin Pill |
| RxNorm | 1368383 | alogliptin / metformin Oral Tablet |
| RxNorm | 1368384 | alogliptin / metformin |
| RxNorm | 1368385 | alogliptin 12.5 MG / metformin hydrochloride 1000 MG Oral Tablet |
| RxNorm | 1368387 | alogliptin 12.5 MG / metformin hydrochloride 1000 MG [Kazano] |
| RxNorm | 1368391 | alogliptin 12.5 MG / metformin hydrochloride 1000 MG Oral Tablet [Kazano] |
| RxNorm | 1368392 | alogliptin 12.5 MG / metformin hydrochloride 500 MG Oral Tablet |

Continued on next page

**Table24 – continued from previous page**

| CodeSystem | ConceptCode | ConceptName |
| --- | --- | --- |
| RxNorm | 1368394 | alogliptin 12.5 MG / metformin hydrochloride 500 MG [Kazano] |
| RxNorm | 1368395 | alogliptin / metformin Oral Tablet [Kazano] |
| RxNorm | 1368396 | Kazano Oral Product |
| RxNorm | 1368397 | Kazano Pill |
| RxNorm | 1368398 | alogliptin 12.5 MG / metformin hydrochloride 500 MG Oral Tablet [Kazano] |
| RxNorm | 1372692 | Kazano |
| RxNorm | 1372706 | Jentadueto |
| RxNorm | 1372716 | PrandiMet |
| RxNorm | 1372730 | Kombiglyze |
| RxNorm | 1372738 | Janumet |
| RxNorm | 1486436 | dapagliflozin / metformin |
| RxNorm | 151825 | Glucamet |
| RxNorm | 151827 | Glucophage |
| RxNorm | 1545146 | canagliflozin / metformin Oral Product |
| RxNorm | 1545147 | canagliflozin / metformin Pill |
| RxNorm | 1545148 | canagliflozin / metformin Oral Tablet |
| RxNorm | 1545149 | canagliflozin / metformin |
| RxNorm | 1545150 | canagliflozin 150 MG / metformin hydrochloride 1000 MG Oral Tablet |
| RxNorm | 1545151 | Invokamet |
| RxNorm | 1545152 | canagliflozin 150 MG / metformin hydrochloride 1000 MG [Invokamet] |
| RxNorm | 1545153 | canagliflozin / metformin Oral Tablet [Invokamet] |
| RxNorm | 1545154 | Invokamet Oral Product |
| RxNorm | 1545155 | Invokamet Pill |
| RxNorm | 1545156 | canagliflozin 150 MG / metformin hydrochloride 1000 MG Oral Tablet [Invokamet] |
| RxNorm | 1545157 | canagliflozin 150 MG / metformin hydrochloride 500 MG Oral Tablet |
| RxNorm | 1545158 | canagliflozin 150 MG / metformin hydrochloride 500 MG [Invokamet] |
| RxNorm | 1545159 | canagliflozin 150 MG / metformin hydrochloride 500 MG Oral Tablet [Invokamet] |
| RxNorm | 1545161 | canagliflozin 50 MG / metformin hydrochloride 1000 MG Oral Tablet |
| RxNorm | 1545162 | canagliflozin 50 MG / metformin hydrochloride 1000 MG [Invokamet] |
| RxNorm | 1545163 | canagliflozin 50 MG / metformin hydrochloride 1000 MG Oral Tablet [Invokamet] |
| RxNorm | 1545164 | canagliflozin 50 MG / metformin hydrochloride 500 MG Oral Tablet |

Continued on next page

**Table24 – continued from previous page**

| CodeSystem | ConceptCode | ConceptName |
|---|---|---|
| RxNorm | 1545165 | canagliflozin 50 MG / metformin hydrochloride 500 MG [Invokamet] |
| RxNorm | 1545166 | canagliflozin 50 MG / metformin hydrochloride 500 MG Oral Tablet [Invokamet] |
| RxNorm | 152161 | Orabet Metformin |
| RxNorm | 1593057 | dapagliflozin / metformin Extended Release Oral Tablet |
| RxNorm | 1593058 | 24 HR dapagliflozin 10 MG / metformin hydrochloride 1000 MG Extended Release Oral Tablet |
| RxNorm | 1593059 | dapagliflozin 10 MG / metformin hydrochloride 1000 MG Extended Release Oral Tablet |
| RxNorm | 1593068 | 24 HR dapagliflozin 10 MG / metformin hydrochloride 500 MG Extended Release Oral Tablet |
| RxNorm | 1593069 | dapagliflozin 10 MG / metformin hydrochloride 500 MG Extended Release Oral Tablet |
| RxNorm | 1593070 | 24 HR dapagliflozin 5 MG / metformin hydrochloride 1000 MG Extended Release Oral Tablet |
| RxNorm | 1593071 | dapagliflozin 5 MG / metformin hydrochloride 1000 MG Extended Release Oral Tablet |
| RxNorm | 1593072 | 24 HR dapagliflozin 5 MG / metformin hydrochloride 500 MG Extended Release Oral Tablet |
| RxNorm | 1593073 | dapagliflozin 5 MG / metformin hydrochloride 500 MG Extended Release Oral Tablet |
| RxNorm | 1593774 | dapagliflozin / metformin Extended Release Oral Tablet [Xigduo] |
| RxNorm | 1593775 | 24 HR dapagliflozin 10 MG / metformin hydrochloride 1000 MG Extended Release Oral Tablet [Xigduo] |
| RxNorm | 1593776 | dapagliflozin 10 MG / metformin hydrochloride 1000 MG Extended Release Oral Tablet [Xigduo] |
| RxNorm | 1593826 | dapagliflozin 10 MG / metformin hydrochloride 500 MG [Xigduo] |
| RxNorm | 1593827 | dapagliflozin 10 MG / metformin hydrochloride 500 MG Extended Release Oral Tablet [Xigduo] |
| RxNorm | 1593828 | dapagliflozin 5 MG / metformin hydrochloride 1000 MG [Xigduo] |
| RxNorm | 1593829 | dapagliflozin 5 MG / metformin hydrochloride 1000 MG Extended Release Oral Tablet [Xigduo] |
| RxNorm | 1593830 | dapagliflozin 5 MG / metformin hydrochloride 500 MG [Xigduo] |
| RxNorm | 1593831 | 24 HR dapagliflozin 5 MG / metformin hydrochloride 500 MG Extended Release Oral Tablet [Xigduo] |
| RxNorm | 1593832 | dapagliflozin 5 MG / metformin hydrochloride 500 MG Extended Release Oral Tablet [Xigduo] |

Continued on next page

**Table24 – continued from previous page**

| CodeSystem | ConceptCode | ConceptName |
|---|---|---|
| RxNorm | 1593833 | 24 HR dapagliflozin 5 MG / metformin hydrochloride 1000 MG Extended Release Oral Tablet [Xigduo] |
| RxNorm | 1593835 | 24 HR dapagliflozin 10 MG / metformin hydrochloride 500 MG Extended Release Oral Tablet [Xigduo] |
| RxNorm | 1592709 | dapagliflozin / metformin Oral Product |
| RxNorm | 1592710 | dapagliflozin / metformin Pill |
| RxNorm | 1592713 | Xigduo |
| RxNorm | 1592716 | Xigduo Oral Product |
| RxNorm | 1592717 | Xigduo Pill |
| RxNorm | 1592722 | dapagliflozin 10 MG / metformin hydrochloride 1000 MG [Xigduo] |
| RxNorm | 1664311 | empagliflozin / metformin Oral Product |
| RxNorm | 1664312 | empagliflozin / metformin Pill |
| RxNorm | 1664313 | empagliflozin / metformin Oral Tablet |
| RxNorm | 1664314 | empagliflozin / metformin |
| RxNorm | 1664315 | empagliflozin 5 MG / metformin hydrochloride 500 MG Oral Tablet |
| RxNorm | 1664316 | Synjardy |
| RxNorm | 1664317 | empagliflozin 5 MG / metformin hydrochloride 500 MG [Synjardy] |
| RxNorm | 1664318 | empagliflozin / metformin Oral Tablet [Synjardy] |
| RxNorm | 1664319 | Synjardy Oral Product |
| RxNorm | 1664320 | Synjardy Pill |
| RxNorm | 1664321 | empagliflozin 5 MG / metformin hydrochloride 500 MG Oral Tablet [Synjardy] |
| RxNorm | 1664323 | empagliflozin 12.5 MG / metformin hydrochloride 500 MG Oral Tablet |
| RxNorm | 1664324 | empagliflozin 12.5 MG / metformin hydrochloride 500 MG [Synjardy] |
| RxNorm | 1664325 | empagliflozin 12.5 MG / metformin hydrochloride 500 MG Oral Tablet [Synjardy] |
| RxNorm | 1664326 | empagliflozin 5 MG / metformin hydrochloride 1000 MG Oral Tablet |
| RxNorm | 1664327 | empagliflozin 5 MG / metformin hydrochloride 1000 MG [Synjardy] |
| RxNorm | 1664328 | empagliflozin 5 MG / metformin hydrochloride 1000 MG Oral Tablet [Synjardy] |
| RxNorm | 1665367 | empagliflozin 12.5 MG / metformin hydrochloride 1000 MG Oral Tablet |
| RxNorm | 1665368 | empagliflozin 12.5 MG / metformin hydrochloride 1000 MG [Synjardy] |
| RxNorm | 1665369 | empagliflozin 12.5 MG / metformin hydrochloride 1000 MG Oral Tablet [Synjardy] |

Continued on next page

**Table24 – continued from previous page**

| CodeSystem | ConceptCode | ConceptName |
|---|---|---|
| RxNorm | 1796088 | linagliptin / metformin Extended Release Oral Tablet |
| RxNorm | 1796089 | 24 HR linagliptin 2.5 MG / metformin hydrochloride 1000 MG Extended Release Oral Tablet |
| RxNorm | 1796090 | linagliptin / metformin Extended Release Oral Tablet [Jentadueto] |
| RxNorm | 1796091 | 24 HR linagliptin 2.5 MG / metformin hydrochloride 1000 MG Extended Release Oral Tablet [Jentadueto] |
| RxNorm | 1796092 | linagliptin 2.5 MG / metformin hydrochloride 1000 MG Extended Release Oral Tablet |
| RxNorm | 1796093 | linagliptin 2.5 MG / metformin hydrochloride 1000 MG Extended Release Oral Tablet [Jentadueto] |
| RxNorm | 1796094 | 24 HR linagliptin 5 MG / metformin hydrochloride 1000 MG Extended Release Oral Tablet |
| RxNorm | 1796095 | linagliptin 5 MG / metformin hydrochloride 1000 MG [Jentadueto] |
| RxNorm | 1796096 | 24 HR linagliptin 5 MG / metformin hydrochloride 1000 MG Extended Release Oral Tablet [Jentadueto] |
| RxNorm | 1796097 | linagliptin 5 MG / metformin hydrochloride 1000 MG Extended Release Oral Tablet |
| RxNorm | 1796098 | linagliptin 5 MG / metformin hydrochloride 1000 MG Extended Release Oral Tablet [Jentadueto] |
| RxNorm | 1807888 | Modified 24 HR metformin hydrochloride 1000 MG Extended Release Oral Tablet |
| RxNorm | 1807894 | Osmotic 24 HR metformin hydrochloride 1000 MG Extended Release Oral Tablet |
| RxNorm | 1807915 | Modified 24 HR metformin hydrochloride 500 MG Extended Release Oral Tablet |
| RxNorm | 1807917 | Osmotic 24 HR metformin hydrochloride 500 MG Extended Release Oral Tablet |
| RxNorm | 1810996 | canagliflozin / metformin Extended Release Oral Tablet |
| RxNorm | 1810997 | 24 HR canagliflozin 150 MG / metformin hydrochloride 1000 MG Extended Release Oral Tablet |
| RxNorm | 1810998 | canagliflozin / metformin Extended Release Oral Tablet [Invokamet] |
| RxNorm | 1810999 | 24 HR canagliflozin 150 MG / metformin hydrochloride 1000 MG Extended Release Oral Tablet [Invokamet] |
| RxNorm | 1811000 | canagliflozin 150 MG / metformin hydrochloride 1000 MG Extended Release Oral Tablet |
| RxNorm | 1811001 | canagliflozin 150 MG / metformin hydrochloride 1000 MG Extended Release Oral Tablet [Invokamet] |
| RxNorm | 1811002 | 24 HR canagliflozin 150 MG / metformin hydrochloride 500 MG Extended Release Oral Tablet |

Continued on next page

**Table24 – continued from previous page**

| CodeSystem | ConceptCode | ConceptName |
|---|---|---|
| RxNorm | 1811003 | 24 HR canagliflozin 150 MG / metformin hydrochloride 500 MG Extended Release Oral Tablet [Invokamet] |
| RxNorm | 1811004 | canagliflozin 150 MG / metformin hydrochloride 500 MG Extended Release Oral Tablet |
| RxNorm | 1811005 | canagliflozin 150 MG / metformin hydrochloride 500 MG Extended Release Oral Tablet [Invokamet] |
| RxNorm | 1811006 | 24 HR canagliflozin 50 MG / metformin hydrochloride 1000 MG Extended Release Oral Tablet |
| RxNorm | 1811007 | 24 HR canagliflozin 50 MG / metformin hydrochloride 1000 MG Extended Release Oral Tablet [Invokamet] |
| RxNorm | 1862684 | empagliflozin / metformin Extended Release Oral Tablet |
| RxNorm | 1862685 | 24 HR empagliflozin 10 MG / metformin hydrochloride 1000 MG Extended Release Oral Tablet |
| RxNorm | 1862686 | empagliflozin 10 MG / metformin hydrochloride 1000 MG [Synjardy] |
| RxNorm | 1862687 | empagliflozin / metformin Extended Release Oral Tablet [Synjardy] |
| RxNorm | 1862688 | 24 HR empagliflozin 10 MG / metformin hydrochloride 1000 MG Extended Release Oral Tablet [Synjardy] |
| RxNorm | 1862689 | empagliflozin 10 MG / metformin hydrochloride 1000 MG Extended Release Oral Tablet |
| RxNorm | 1862690 | empagliflozin 10 MG / metformin hydrochloride 1000 MG Extended Release Oral Tablet [Synjardy] |
| RxNorm | 1862691 | 24 HR empagliflozin 12.5 MG / metformin hydrochloride 1000 MG Extended Release Oral Tablet |
| RxNorm | 1862692 | 24 HR empagliflozin 12.5 MG / metformin hydrochloride 1000 MG Extended Release Oral Tablet [Synjardy] |
| RxNorm | 1862693 | empagliflozin 12.5 MG / metformin hydrochloride 1000 MG Extended Release Oral Tablet |
| RxNorm | 1811008 | canagliflozin 50 MG / metformin hydrochloride 1000 MG Extended Release Oral Tablet |
| RxNorm | 1811009 | canagliflozin 50 MG / metformin hydrochloride 1000 MG Extended Release Oral Tablet [Invokamet] |
| RxNorm | 1811010 | 24 HR canagliflozin 50 MG / metformin hydrochloride 500 MG Extended Release Oral Tablet |
| RxNorm | 1811011 | 24 HR canagliflozin 50 MG / metformin hydrochloride 500 MG Extended Release Oral Tablet [Invokamet] |
| RxNorm | 1811012 | canagliflozin 50 MG / metformin hydrochloride 500 MG Extended Release Oral Tablet |
| RxNorm | 1811013 | canagliflozin 50 MG / metformin hydrochloride 500 MG Extended Release Oral Tablet [Invokamet] |
| RxNorm | 1862694 | empagliflozin 12.5 MG / metformin hydrochloride 1000 MG Extended Release Oral Tablet [Synjardy] |

Continued on next page

**Table24 – continued from previous page**

| CodeSystem | ConceptCode | ConceptName |
|---|---|---|
| RxNorm | 1862695 | 24 HR empagliflozin 25 MG / metformin hydrochloride 1000 MG Extended Release Oral Tablet |
| RxNorm | 1862696 | empagliflozin 25 MG / metformin hydrochloride 1000 MG [Synjardy] |
| RxNorm | 1862697 | 24 HR empagliflozin 25 MG / metformin hydrochloride 1000 MG Extended Release Oral Tablet [Synjardy] |
| RxNorm | 1862698 | empagliflozin 25 MG / metformin hydrochloride 1000 MG Extended Release Oral Tablet |
| RxNorm | 1862699 | empagliflozin 25 MG / metformin hydrochloride 1000 MG Extended Release Oral Tablet [Synjardy] |
| RxNorm | 1862700 | 24 HR empagliflozin 5 MG / metformin hydrochloride 1000 MG Extended Release Oral Tablet |
| RxNorm | 1862701 | 24 HR empagliflozin 5 MG / metformin hydrochloride 1000 MG Extended Release Oral Tablet [Synjardy] |
| RxNorm | 1862702 | empagliflozin 5 MG / metformin hydrochloride 1000 MG Extended Release Oral Tablet |
| RxNorm | 1862703 | empagliflozin 5 MG / metformin hydrochloride 1000 MG Extended Release Oral Tablet [Synjardy] |
| RxNorm | 1940496 | 24 HR dapagliflozin 2.5 MG / metformin hydrochloride 1000 MG Extended Release Oral Tablet |
| RxNorm | 1940497 | dapagliflozin 2.5 MG / metformin hydrochloride 1000 MG [Xigduo] |
| RxNorm | 1940498 | 24 HR dapagliflozin 2.5 MG / metformin hydrochloride 1000 MG Extended Release Oral Tablet [Xigduo] |
| RxNorm | 1940499 | dapagliflozin 2.5 MG / metformin hydrochloride 1000 MG Extended Release Oral Tablet |
| RxNorm | 1940500 | dapagliflozin 2.5 MG / metformin hydrochloride 1000 MG Extended Release Oral Tablet [Xigduo] |
| RxNorm | 1992681 | ertugliflozin / metformin Oral Product |
| RxNorm | 1992682 | ertugliflozin / metformin Pill |
| RxNorm | 1992683 | ertugliflozin / metformin Oral Tablet |
| RxNorm | 1992684 | ertugliflozin / metformin |
| RxNorm | 1992685 | ertugliflozin 2.5 MG / metformin hydrochloride 1000 MG Oral Tablet |
| RxNorm | 1992686 | Segluromet |
| RxNorm | 1992687 | ertugliflozin 2.5 MG / metformin hydrochloride 1000 MG [Segluromet] |
| RxNorm | 1992688 | ertugliflozin / metformin Oral Tablet [Segluromet] |
| RxNorm | 1992689 | Segluromet Oral Product |
| RxNorm | 1992690 | Segluromet Pill |
| RxNorm | 1992691 | ertugliflozin 2.5 MG / metformin hydrochloride 1000 MG Oral Tablet [Segluromet] |

Continued on next page

**Table24 – continued from previous page**

| CodeSystem | ConceptCode | ConceptName |
|---|---|---|
| RxNorm | 1992693 | ertugliflozin 2.5 MG / metformin hydrochloride 500 MG Oral Tablet |
| RxNorm | 1992694 | ertugliflozin 2.5 MG / metformin hydrochloride 500 MG [Segluromet] |
| RxNorm | 1992695 | ertugliflozin 2.5 MG / metformin hydrochloride 500 MG Oral Tablet [Segluromet] |
| RxNorm | 1992698 | ertugliflozin 7.5 MG / metformin hydrochloride 1000 MG Oral Tablet |
| RxNorm | 1992699 | ertugliflozin 7.5 MG / metformin hydrochloride 1000 MG [Segluromet] |
| RxNorm | 1992700 | ertugliflozin 7.5 MG / metformin hydrochloride 1000 MG Oral Tablet [Segluromet] |
| RxNorm | 1992701 | ertugliflozin 7.5 MG / metformin hydrochloride 500 MG Oral Tablet |
| RxNorm | 1992702 | ertugliflozin 7.5 MG / metformin hydrochloride 500 MG [Segluromet] |
| RxNorm | 1992703 | ertugliflozin 7.5 MG / metformin hydrochloride 500 MG Oral Tablet [Segluromet] |
| RxNorm | 204045 | metformin hydrochloride 500 MG Oral Tablet [Orabet Metformin] |
| RxNorm | 204047 | metformin hydrochloride 850 MG Oral Tablet [Orabet Metformin] |
| RxNorm | 2117292 | dapagliflozin / metformin / saxagliptin |
| RxNorm | 2200517 | metformin Extended Release Suspension |
| RxNorm | 2200518 | metformin hydrochloride 100 MG/ML Extended Release Suspension |
| RxNorm | 2200519 | metformin Extended Release Suspension [Riomet] |
| RxNorm | 2200520 | metformin hydrochloride 100 MG/ML Extended Release Suspension [Riomet] |
| RxNorm | 2281864 | empagliflozin / linagliptin / metformin |
| RxNorm | 2371722 | dapagliflozin / metformin / saxagliptin Oral Product |
| RxNorm | 2371723 | dapagliflozin / metformin / saxagliptin Pill |
| RxNorm | 2371724 | dapagliflozin / metformin / saxagliptin Extended Release Oral Tablet |
| RxNorm | 2371725 | 24 HR dapagliflozin 10 MG / metformin hydrochloride 1000 MG / saxagliptin 5 MG Extended Release Oral Tablet |
| RxNorm | 2371726 | Qternmet |
| RxNorm | 2371727 | dapagliflozin 10 MG / metformin hydrochloride 1000 MG / saxagliptin 5 MG [Qternmet] |
| RxNorm | 2371728 | dapagliflozin / metformin / saxagliptin Extended Release Oral Tablet [Qternmet] |
| RxNorm | 2371729 | Qternmet Oral Product |
| RxNorm | 2371730 | Qternmet Pill |

Continued on next page

**Table24 – continued from previous page**

| CodeSystem | ConceptCode | ConceptName |
|---|---|---|
| RxNorm | 2371731 | 24 HR dapagliflozin 10 MG / metformin hydrochloride 1000 MG / saxagliptin 5 MG Extended Release Oral Tablet [Qternmet] |
| RxNorm | 2371732 | dapagliflozin 10 MG / metformin hydrochloride 1000 MG / saxagliptin 5 MG Extended Release Oral Tablet |
| RxNorm | 2371733 | dapagliflozin 10 MG / metformin hydrochloride 1000 MG / saxagliptin 5 MG Extended Release Oral Tablet [Qternmet] |
| RxNorm | 2371734 | 24 HR dapagliflozin 2.5 MG / metformin hydrochloride 1000 MG / saxagliptin 2.5 MG Extended Release Oral Tablet |
| RxNorm | 2371735 | dapagliflozin 2.5 MG / metformin hydrochloride 1000 MG / saxagliptin 2.5 MG [Qternmet] |
| RxNorm | 2371736 | 24 HR dapagliflozin 2.5 MG / metformin hydrochloride 1000 MG / saxagliptin 2.5 MG Extended Release Oral Tablet [Qternmet] |
| RxNorm | 2371737 | dapagliflozin 2.5 MG / metformin hydrochloride 1000 MG / saxagliptin 2.5 MG Extended Release Oral Tablet |
| RxNorm | 2371738 | dapagliflozin 2.5 MG / metformin hydrochloride 1000 MG / saxagliptin 2.5 MG Extended Release Oral Tablet [Qternmet] |
| RxNorm | 2371740 | 24 HR dapagliflozin 5 MG / metformin hydrochloride 1000 MG / saxagliptin 2.5 MG Extended Release Oral Tablet |
| RxNorm | 2371741 | dapagliflozin 5 MG / metformin hydrochloride 1000 MG / saxagliptin 2.5 MG [Qternmet] |
| RxNorm | 2371742 | 24 HR dapagliflozin 5 MG / metformin hydrochloride 1000 MG / saxagliptin 2.5 MG Extended Release Oral Tablet [Qternmet] |
| RxNorm | 2371743 | dapagliflozin 5 MG / metformin hydrochloride 1000 MG / saxagliptin 2.5 MG Extended Release Oral Tablet |
| RxNorm | 2371744 | dapagliflozin 5 MG / metformin hydrochloride 1000 MG / saxagliptin 2.5 MG Extended Release Oral Tablet [Qternmet] |
| RxNorm | 2371745 | 24 HR dapagliflozin 5 MG / metformin hydrochloride 1000 MG / saxagliptin 5 MG Extended Release Oral Tablet |
| RxNorm | 2371746 | dapagliflozin 5 MG / metformin hydrochloride 1000 MG / saxagliptin 5 MG [Qternmet] |
| RxNorm | 2371747 | 24 HR dapagliflozin 5 MG / metformin hydrochloride 1000 MG / saxagliptin 5 MG Extended Release Oral Tablet [Qternmet] |
| RxNorm | 2371748 | dapagliflozin 5 MG / metformin hydrochloride 1000 MG / saxagliptin 5 MG Extended Release Oral Tablet |
| RxNorm | 2371749 | dapagliflozin 5 MG / metformin hydrochloride 1000 MG / saxagliptin 5 MG Extended Release Oral Tablet [Qternmet] |
| RxNorm | 2359276 | empagliflozin / linagliptin / metformin Oral Product |

Continued on next page

**Table24 – continued from previous page**

| CodeSystem | ConceptCode | ConceptName |
|---|---|---|
| RxNorm | 2359277 | empagliflozin / linagliptin / metformin Pill |
| RxNorm | 2359278 | empagliflozin / linagliptin / metformin Extended Release Oral Tablet |
| RxNorm | 2359279 | 24 HR empagliflozin 10 MG / linagliptin 5 MG / metformin hydrochloride 1000 MG Extended Release Oral Tablet |
| RxNorm | 2359280 | Trijardy |
| RxNorm | 2359281 | empagliflozin 10 MG / linagliptin 5 MG / metformin hydrochloride 1000 MG [Trijardy] |
| RxNorm | 2359282 | empagliflozin / linagliptin / metformin Extended Release Oral Tablet [Trijardy] |
| RxNorm | 2359283 | Trijardy Oral Product |
| RxNorm | 2359284 | Trijardy Pill |
| RxNorm | 2359285 | 24 HR empagliflozin 10 MG / linagliptin 5 MG / metformin hydrochloride 1000 MG Extended Release Oral Tablet [Trijardy] |
| RxNorm | 2359286 | empagliflozin 10 MG / linagliptin 5 MG / metformin hydrochloride 1000 MG Extended Release Oral Tablet |
| RxNorm | 2359287 | empagliflozin 10 MG / linagliptin 5 MG / metformin hydrochloride 1000 MG Extended Release Oral Tablet [Trijardy] |
| RxNorm | 2359288 | 24 HR empagliflozin 12.5 MG / linagliptin 2.5 MG / metformin hydrochloride 1000 MG Extended Release Oral Tablet |
| RxNorm | 2359289 | empagliflozin 12.5 MG / linagliptin 2.5 MG / metformin hydrochloride 1000 MG [Trijardy] |
| RxNorm | 2359290 | 24 HR empagliflozin 12.5 MG / linagliptin 2.5 MG / metformin hydrochloride 1000 MG Extended Release Oral Tablet [Trijardy] |
| RxNorm | 2359291 | empagliflozin 12.5 MG / linagliptin 2.5 MG / metformin hydrochloride 1000 MG Extended Release Oral Tablet |
| RxNorm | 2359292 | empagliflozin 12.5 MG / linagliptin 2.5 MG / metformin hydrochloride 1000 MG Extended Release Oral Tablet [Trijardy] |
| RxNorm | 2359351 | 24 HR empagliflozin 25 MG / linagliptin 5 MG / metformin hydrochloride 1000 MG Extended Release Oral Tablet |
| RxNorm | 2359352 | empagliflozin 25 MG / linagliptin 5 MG / metformin hydrochloride 1000 MG [Trijardy] |
| RxNorm | 2359353 | 24 HR empagliflozin 25 MG / linagliptin 5 MG / metformin hydrochloride 1000 MG Extended Release Oral Tablet [Trijardy] |
| RxNorm | 2359354 | empagliflozin 25 MG / linagliptin 5 MG / metformin hydrochloride 1000 MG Extended Release Oral Tablet |

Continued on next page

**Table24 – continued from previous page**

| CodeSystem | ConceptCode | ConceptName |
|---|---|---|
| RxNorm | 2359355 | empagliflozin 25 MG / linagliptin 5 MG / metformin hydrochloride 1000 MG Extended Release Oral Tablet [Trijardy] |
| RxNorm | 2359356 | 24 HR empagliflozin 5 MG / linagliptin 2.5 MG / metformin hydrochloride 1000 MG Extended Release Oral Tablet |
| RxNorm | 2359357 | empagliflozin 5 MG / linagliptin 2.5 MG / metformin hydrochloride 1000 MG [Trijardy] |
| RxNorm | 2359358 | 24 HR empagliflozin 5 MG / linagliptin 2.5 MG / metformin hydrochloride 1000 MG Extended Release Oral Tablet [Trijardy] |
| RxNorm | 2359359 | empagliflozin 5 MG / linagliptin 2.5 MG / metformin hydrochloride 1000 MG Extended Release Oral Tablet |
| RxNorm | 2359360 | empagliflozin 5 MG / linagliptin 2.5 MG / metformin hydrochloride 1000 MG Extended Release Oral Tablet [Trijardy] |
| RxNorm | 246522 | chlorpropamide 125 MG / metformin 400 MG Oral Tablet |
| RxNorm | 250919 | glyburide 2.5 MG / metformin 400 MG Oral Tablet |
| RxNorm | 284743 | Glucovance |
| RxNorm | 285129 | glyburide / metformin |
| RxNorm | 330861 | metformin 250 MG |
| RxNorm | 316255 | metformin 1000 MG |
| RxNorm | 316256 | metformin 500 MG |
| RxNorm | 332809 | metformin 400 MG |
| RxNorm | 352381 | glipizide / metformin |
| RxNorm | 352450 | Avandamet |
| RxNorm | 352764 | Metaglip |
| RxNorm | 368254 | metformin Oral Tablet [Glucophage] |
| RxNorm | 361841 | metformin Oral Tablet [Orabet Metformin] |
| RxNorm | 368526 | metformin Oral Tablet [Glucamet] |
| RxNorm | 371466 | chlorpropamide / metformin Oral Tablet |
| RxNorm | 374635 | glyburide / metformin Oral Tablet |
| RxNorm | 372803 | metformin Oral Tablet |
| RxNorm | 372804 | metformin Extended Release Oral Tablet |
| RxNorm | 378729 | metformin / rosiglitazone Oral Tablet |
| RxNorm | 378730 | glipizide / metformin Oral Tablet |
| RxNorm | 405304 | Riomet |
| RxNorm | 406082 | metformin Oral Solution |
| RxNorm | 406257 | metformin Oral Solution [Riomet] |
| RxNorm | 431724 | 12 HR metformin hydrochloride 850 MG Extended Release Oral Tablet |
| RxNorm | 432366 | chlorpropamide 125 MG / metformin 513 MG Oral Tablet |
| RxNorm | 428759 | metformin 250 MG Oral Tablet |
| RxNorm | 432780 | chlorpropamide 125 MG / metformin 500 MG Oral Tablet |

Continued on next page

**Table24 – continued from previous page**

| CodeSystem | ConceptCode | ConceptName |
|---|---|---|
| RxNorm | 429841 | glyburide 5 MG / metformin 1000 MG Oral Tablet |
| RxNorm | 438507 | metformin 513 MG |
| RxNorm | 541766 | Fortamet |
| RxNorm | 541768 | metformin Extended Release Oral Tablet [Fortamet] |
| RxNorm | 541774 | metformin / ropinirole Oral Tablet |
| RxNorm | 541775 | metformin 500 MG / ropinirole 2 MG Oral Tablet |
| RxNorm | 583192 | Metforming |
| RxNorm | 583194 | metformin Oral Tablet [Metforming] |
| RxNorm | 583195 | metformin hydrochloride 850 MG Oral Tablet [Metforming] |
| RxNorm | 602411 | Actoplus Met |
| RxNorm | 607999 | metformin / pioglitazone |
| RxNorm | 614348 | metformin / rosiglitazone |
| RxNorm | 577093 | metformin / pioglitazone Oral Tablet |
| RxNorm | 645109 | Glumetza |
| RxNorm | 647241 | metformin Extended Release Oral Tablet [Glumetza] |
| RxNorm | 700516 | metformin / sitagliptin Oral Tablet |
| RxNorm | 729717 | metformin / sitagliptin |
| RxNorm | 731442 | metformin / pioglitazone Oral Tablet [Actoplus Met] |
| RxNorm | 757603 | metformin / sitagliptin Oral Tablet [Janumet] |
| RxNorm | 802051 | metformin Extended Release Oral Tablet [Glucophage] |
| RxNorm | 802646 | metformin / repaglinide |
| RxNorm | 802742 | metformin / repaglinide Oral Tablet |
| RxNorm | 849585 | glipizide / metformin Oral Tablet [Metaglip] |
| RxNorm | 805670 | metformin / repaglinide Oral Tablet [PrandiMet] |
| RxNorm | 806287 | metformin / rosiglitazone Oral Tablet [Avandamet] |
| RxNorm | 861730 | metformin hydrochloride 250 MG |
| RxNorm | 861731 | glipizide 2.5 MG / metformin hydrochloride 250 MG Oral Tablet |
| RxNorm | 861732 | glipizide 2.5 MG / metformin hydrochloride 250 MG [Metaglip] |
| RxNorm | 861733 | glipizide 2.5 MG / metformin hydrochloride 250 MG Oral Tablet [Metaglip] |
| RxNorm | 861736 | glipizide 2.5 MG / metformin hydrochloride 500 MG Oral Tablet |
| RxNorm | 861737 | glipizide 2.5 MG / metformin hydrochloride 500 MG [Metaglip] |
| RxNorm | 861738 | glipizide 2.5 MG / metformin hydrochloride 500 MG Oral Tablet [Metaglip] |
| RxNorm | 861740 | glipizide 5 MG / metformin hydrochloride 500 MG Oral Tablet |
| RxNorm | 861741 | glipizide 5 MG / metformin hydrochloride 500 MG [Metaglip] |

Continued on next page

**Table24 – continued from previous page**

| CodeSystem | ConceptCode | ConceptName |
|---|---|---|
| RxNorm | 861742 | glipizide 5 MG / metformin hydrochloride 500 MG Oral Tablet [Metaglip] |
| RxNorm | 861743 | glyburide 1.25 MG / metformin hydrochloride 250 MG Oral Tablet |
| RxNorm | 861745 | glyburide 1.25 MG / metformin hydrochloride 250 MG [Glucovance] |
| RxNorm | 861747 | glyburide 1.25 MG / metformin hydrochloride 250 MG Oral Tablet [Glucovance] |
| RxNorm | 861748 | glyburide 2.5 MG / metformin hydrochloride 500 MG Oral Tablet |
| RxNorm | 861750 | glyburide 2.5 MG / metformin hydrochloride 500 MG [Glucovance] |
| RxNorm | 861752 | glyburide 2.5 MG / metformin hydrochloride 500 MG Oral Tablet [Glucovance] |
| RxNorm | 861753 | glyburide 5 MG / metformin hydrochloride 500 MG Oral Tablet |
| RxNorm | 861755 | glyburide 5 MG / metformin hydrochloride 500 MG [Glucovance] |
| RxNorm | 861756 | glyburide / metformin Oral Tablet [Glucovance] |
| RxNorm | 861757 | glyburide 5 MG / metformin hydrochloride 500 MG Oral Tablet [Glucovance] |
| RxNorm | 861760 | metformin hydrochloride 1000 MG / rosiglitazone 2 MG Oral Tablet |
| RxNorm | 861761 | metformin hydrochloride 1000 MG / rosiglitazone 2 MG [Avandamet] |
| RxNorm | 861762 | metformin hydrochloride 1000 MG / rosiglitazone 2 MG Oral Tablet [Avandamet] |
| RxNorm | 861763 | metformin hydrochloride 1000 MG / rosiglitazone 4 MG Oral Tablet |
| RxNorm | 861764 | metformin hydrochloride 1000 MG / rosiglitazone 4 MG [Avandamet] |
| RxNorm | 875864 | metformin hydrochloride 500 MG [Glucamet] |
| RxNorm | 875865 | metformin hydrochloride 850 MG [Glucamet] |
| RxNorm | 861765 | metformin hydrochloride 1000 MG / rosiglitazone 4 MG Oral Tablet [Avandamet] |
| RxNorm | 861769 | metformin hydrochloride 1000 MG / sitagliptin 50 MG Oral Tablet |
| RxNorm | 861770 | metformin hydrochloride 1000 MG / sitagliptin 50 MG [Janumet] |
| RxNorm | 861771 | metformin hydrochloride 1000 MG / sitagliptin 50 MG Oral Tablet [Janumet] |
| RxNorm | 861783 | metformin hydrochloride 500 MG / pioglitazone 15 MG Oral Tablet |

Continued on next page

**Table24 – continued from previous page**

| CodeSystem | ConceptCode | ConceptName |
|---|---|---|
| RxNorm | 861784 | metformin hydrochloride 500 MG / pioglitazone 15 MG [Actoplus Met] |
| RxNorm | 861785 | metformin hydrochloride 500 MG / pioglitazone 15 MG Oral Tablet [Actoplus Met] |
| RxNorm | 861787 | metformin hydrochloride 500 MG / repaglinide 1 MG Oral Tablet |
| RxNorm | 861788 | metformin hydrochloride 500 MG / repaglinide 1 MG [PrandiMet] |
| RxNorm | 861789 | metformin hydrochloride 500 MG / repaglinide 1 MG Oral Tablet [PrandiMet] |
| RxNorm | 861790 | metformin hydrochloride 500 MG / repaglinide 2 MG Oral Tablet |
| RxNorm | 861791 | metformin hydrochloride 500 MG / repaglinide 2 MG [PrandiMet] |
| RxNorm | 861792 | metformin hydrochloride 500 MG / repaglinide 2 MG Oral Tablet [PrandiMet] |
| RxNorm | 861795 | metformin hydrochloride 500 MG / rosiglitazone 1 MG Oral Tablet |
| RxNorm | 861796 | metformin hydrochloride 500 MG / rosiglitazone 1 MG [Avandamet] |
| RxNorm | 861797 | metformin hydrochloride 500 MG / rosiglitazone 1 MG Oral Tablet [Avandamet] |
| RxNorm | 861806 | metformin hydrochloride 500 MG / rosiglitazone 2 MG Oral Tablet |
| RxNorm | 861807 | metformin hydrochloride 500 MG / rosiglitazone 2 MG [Avandamet] |
| RxNorm | 861808 | metformin hydrochloride 500 MG / rosiglitazone 2 MG Oral Tablet [Avandamet] |
| RxNorm | 861816 | metformin hydrochloride 500 MG / rosiglitazone 4 MG Oral Tablet |
| RxNorm | 861817 | metformin hydrochloride 500 MG / rosiglitazone 4 MG [Avandamet] |
| RxNorm | 861818 | metformin hydrochloride 500 MG / rosiglitazone 4 MG Oral Tablet [Avandamet] |
| RxNorm | 861819 | metformin hydrochloride 500 MG / sitagliptin 50 MG Oral Tablet |
| RxNorm | 861820 | metformin hydrochloride 500 MG / sitagliptin 50 MG [Janumet] |
| RxNorm | 861821 | metformin hydrochloride 500 MG / sitagliptin 50 MG Oral Tablet [Janumet] |
| RxNorm | 861822 | metformin hydrochloride 850 MG / pioglitazone 15 MG Oral Tablet |

Continued on next page

**Table24 – continued from previous page**

| CodeSystem | ConceptCode | ConceptName |
|---|---|---|
| RxNorm | 861823 | metformin hydrochloride 850 MG / pioglitazone 15 MG [Actoplus Met] |
| RxNorm | 861824 | metformin hydrochloride 850 MG / pioglitazone 15 MG Oral Tablet [Actoplus Met] |
| RxNorm | 876009 | metformin hydrochloride 500 MG [Orabet Metformin] |
| RxNorm | 876010 | metformin hydrochloride 850 MG [Orabet Metformin] |
| RxNorm | 876033 | metformin hydrochloride 850 MG [Metforming] |
| RxNorm | 860974 | metformin hydrochloride 500 MG |
| RxNorm | 860975 | 24 HR metformin hydrochloride 500 MG Extended Release Oral Tablet |
| RxNorm | 860976 | metformin hydrochloride 500 MG [Glucophage] |
| RxNorm | 860977 | 24 HR metformin hydrochloride 500 MG Extended Release Oral Tablet [Glucophage] |
| RxNorm | 860978 | metformin hydrochloride 500 MG Extended Release Oral Tablet |
| RxNorm | 860979 | metformin hydrochloride 500 MG Extended Release Oral Tablet [Glucophage] |
| RxNorm | 860980 | metformin hydrochloride 750 MG |
| RxNorm | 860981 | 24 HR metformin hydrochloride 750 MG Extended Release Oral Tablet |
| RxNorm | 860982 | metformin hydrochloride 750 MG [Glucophage] |
| RxNorm | 860983 | 24 HR metformin hydrochloride 750 MG Extended Release Oral Tablet [Glucophage] |
| RxNorm | 860984 | metformin hydrochloride 750 MG Extended Release Oral Tablet |
| RxNorm | 860985 | metformin hydrochloride 750 MG Extended Release Oral Tablet [Glucophage] |
| RxNorm | 860995 | metformin hydrochloride 1000 MG |
| RxNorm | 860997 | metformin hydrochloride 1000 MG [Fortamet] |
| RxNorm | 860998 | Osmotic 24 HR metformin hydrochloride 1000 MG Extended Release Oral Tablet [Fortamet] |
| RxNorm | 860999 | metformin hydrochloride 1000 MG Extended Release Oral Tablet |
| RxNorm | 861000 | metformin hydrochloride 1000 MG Extended Release Oral Tablet [Fortamet] |
| RxNorm | 861001 | metformin hydrochloride 500 MG [Fortamet] |
| RxNorm | 861002 | Osmotic 24 HR metformin hydrochloride 500 MG Extended Release Oral Tablet [Fortamet] |
| RxNorm | 861003 | metformin hydrochloride 500 MG Extended Release Oral Tablet [Fortamet] |
| RxNorm | 861004 | metformin hydrochloride 1000 MG Oral Tablet |
| RxNorm | 861005 | metformin hydrochloride 1000 MG [Glucophage] |

Continued on next page

**Table24 – continued from previous page**

| CodeSystem | ConceptCode | ConceptName |
|---|---|---|
| RxNorm | 861006 | metformin hydrochloride 1000 MG Oral Tablet [Glucophage] |
| RxNorm | 861007 | metformin hydrochloride 500 MG Oral Tablet |
| RxNorm | 861008 | metformin hydrochloride 500 MG Oral Tablet [Glucophage] |
| RxNorm | 861009 | metformin hydrochloride 850 MG |
| RxNorm | 861010 | metformin hydrochloride 850 MG Oral Tablet |
| RxNorm | 861011 | metformin hydrochloride 850 MG [Glucophage] |
| RxNorm | 861012 | metformin hydrochloride 850 MG Oral Tablet [Glucophage] |
| RxNorm | 861014 | metformin hydrochloride 1000 MG [Glumetza] |
| RxNorm | 861015 | Modified 24 HR metformin hydrochloride 1000 MG Extended Release Oral Tablet [Glumetza] |
| RxNorm | 861016 | metformin hydrochloride 1000 MG Extended Release Oral Tablet [Glumetza] |
| RxNorm | 861017 | metformin hydrochloride 500 MG [Glumetza] |
| RxNorm | 861018 | Modified 24 HR metformin hydrochloride 500 MG Extended Release Oral Tablet [Glumetza] |
| RxNorm | 861019 | metformin hydrochloride 500 MG Extended Release Oral Tablet [Glumetza] |
| RxNorm | 861020 | metformin hydrochloride 625 MG |
| RxNorm | 861021 | metformin hydrochloride 625 MG Oral Tablet |
| RxNorm | 861022 | metformin hydrochloride 625 MG [Glucophage] |
| RxNorm | 861023 | metformin hydrochloride 625 MG Oral Tablet [Glucophage] |
| RxNorm | 861024 | metformin hydrochloride 100 MG/ML |
| RxNorm | 861025 | metformin hydrochloride 100 MG/ML Oral Solution |
| RxNorm | 861026 | metformin hydrochloride 100 MG/ML [Riomet] |
| RxNorm | 861027 | metformin hydrochloride 100 MG/ML Oral Solution [Riomet] |
| RxNorm | 899988 | metformin / pioglitazone Extended Release Oral Tablet |
| RxNorm | 899989 | 24 HR metformin hydrochloride 1000 MG / pioglitazone 15 MG Extended Release Oral Tablet |
| RxNorm | 899991 | metformin hydrochloride 1000 MG / pioglitazone 15 MG [Actoplus Met] |
| RxNorm | 899992 | metformin / pioglitazone Extended Release Oral Tablet [Actoplus Met] |
| RxNorm | 899993 | 24 HR metformin hydrochloride 1000 MG / pioglitazone 15 MG Extended Release Oral Tablet [Actoplus Met] |
| RxNorm | 899994 | metformin hydrochloride 1000 MG / pioglitazone 15 MG Extended Release Oral Tablet |
| RxNorm | 899995 | metformin hydrochloride 1000 MG / pioglitazone 15 MG Extended Release Oral Tablet [Actoplus Met] |
| RxNorm | 899996 | 24 HR metformin hydrochloride 1000 MG / pioglitazone 30 MG Extended Release Oral Tablet |

Continued on next page

**Table24 – continued from previous page**

| CodeSystem | ConceptCode | ConceptName |
|---|---|---|
| RxNorm | 899998 | metformin hydrochloride 1000 MG / pioglitazone 30 MG [Actoplus Met] |
| RxNorm | 900000 | 24 HR metformin hydrochloride 1000 MG / pioglitazone 30 MG Extended Release Oral Tablet [Actoplus Met] |
| RxNorm | 900001 | metformin hydrochloride 1000 MG / pioglitazone 30 MG Extended Release Oral Tablet |
| RxNorm | 900002 | metformin hydrochloride 1000 MG / pioglitazone 30 MG Extended Release Oral Tablet [Actoplus Met] |
| RxNorm | 977566 | metformin hydrochloride 850 MG Extended Release Oral Tablet |
| RxNorm | 1007411 | chlorpropamide / metformin |

##### 8.5.2 Insulin

Table 25: Concept codes used to identify if a patient was prescribed insulin-medications.

| CodeSystem | ConceptCode | ConceptName |
|---|---|---|
| RxNorm | 253182 | insulin, regular, human |
| RxNorm | 575133 | Insulin, Isophane, Human 50 UNT/ML / Insulin, Regular, Human 50 UNT/ML [Humulin 50/50] |
| RxNorm | 106893 | Insulin, Isophane, Human 70 UNT/ML / Insulin, Regular, Human 30 UNT/ML Injectable Suspension [Human Mixtard 30] |
| RxNorm | 108812 | Insulin, Isophane, Human 70 UNT/ML / Insulin, Regular, Human 30 UNT/ML Injectable Suspension [Human Mixtard 30 ge] |
| RxNorm | 847195 | 3 ML insulin human, isophane 70 UNT/ML / Regular Insulin, Human 30 UNT/ML Prefilled Syringe [Novolin] |
| RxNorm | 213441 | Insulin, Isophane, Human 70 UNT/ML / Insulin, Regular, Human 30 UNT/ML Injectable Suspension [Humulin 70/30] |
| RxNorm | 153122 | Insulin, Isophane, Human 50 UNT/ML / Insulin, Regular, Human 50 UNT/ML Injectable Suspension [Human Mixtard 50] |
| RxNorm | 311016 | Insulin, Isophane, Human 50 UNT/ML / Insulin, Regular, Human 50 UNT/ML Injectable Suspension [Humulin 50/50] |
| RxNorm | 564604 | Insulin, Isophane, Human 50 UNT/ML / Insulin, Regular, Human 50 UNT/ML [Human Mixtard Penfill] |
| RxNorm | 564394 | Insulin, Isophane, Human 70 UNT/ML / Insulin, Regular, Human 30 UNT/ML [Human Mixtard 30] |
| RxNorm | 564600 | Insulin, Isophane, Human 70 UNT/ML / Insulin, Regular, Human 30 UNT/ML [Human Mixtard 30 ge] |

Continued on next page

**Table25 – continued from previous page**

| CodeSystem | ConceptCode | ConceptName |
|---|---|---|
| RxNorm | 564393 | Insulin, Isophane, Human 70 UNT/ML / Insulin, Regular, Human 30 UNT/ML [Humulin M3] |
| RxNorm | 1544487 | Regular Insulin, Human 0.7 MG/ACTUAT |
| RxNorm | 1544489 | Regular Insulin, Human 0.7 MG/ACTUAT [Afrezza] |
| RxNorm | 1543204 | Regular Insulin, Human 0.35 MG/ACTUAT [Afrezza] |
| RxNorm | 1543201 | Regular Insulin, Human Inhalant Powder |
| RxNorm | 1543205 | Regular Insulin, Human Inhalant Powder [Afrezza] |
| RxNorm | 1543199 | Regular Insulin, Human 0.35 MG/ACTUAT |
| RxNorm | 380934 | Insulin, Isophane, Human / Insulin, Regular, Human Injectable Suspension [Human Mixtard Penfill] |
| RxNorm | 379754 | Insulin, Isophane, Human / Insulin, Regular, Human Injectable Suspension [Novolin 70/30 PenFill] |
| RxNorm | 365578 | Insulin, Isophane, Human / Insulin, Regular, Human Injectable Suspension [Humulin M1] |
| RxNorm | 365580 | Insulin, Isophane, Human / Insulin, Regular, Human Injectable Suspension [Pur-In Mix 50/50] |
| RxNorm | 365577 | Insulin, Isophane, Human / Insulin, Regular, Human Injectable Suspension [Humulin M2] |
| RxNorm | 365575 | Insulin, Isophane, Human / Insulin, Regular, Human Injectable Suspension [Humulin M5] |
| RxNorm | 379741 | Insulin, Isophane, Human / Insulin, Regular, Human Injectable Suspension [Human Mixtard 50] |
| RxNorm | 365673 | Insulin, Isophane, Human / Insulin, Regular, Human Injectable Suspension [Humulin 70/30] |
| RxNorm | 311023 | Insulin, Isophane, Human 70 UNT/ML / Insulin, Regular, Human 30 UNT/ML Injectable Suspension [Novolin 70/30 PenFill] |
| RxNorm | 1008501 | insulin isophane / insulin, regular, human |
| RxNorm | 108813 | insulin isophane, human 90 UNT/ML / insulin, regular, human 10 UNT/ML Injectable Suspension [Human Mixtard] |
| RxNorm | 108814 | insulin isophane, human 80 UNT/ML / insulin, regular, human 20 UNT/ML Injectable Suspension [Human Mixtard] |
| RxNorm | 108815 | insulin isophane, human 70 UNT/ML / insulin, regular, human 30 UNT/ML Injectable Suspension [Human Mixtard] |
| RxNorm | 108816 | insulin isophane, human 50 UNT/ML / insulin, regular, human 50 UNT/ML Injectable Suspension [Human Mixtard] |
| RxNorm | 108822 | insulin isophane, human 60 UNT/ML / insulin, regular, human 40 UNT/ML Injectable Suspension [Human Mixtard] |
| RxNorm | 11160 | Velosulin |
| RxNorm | 106892 | insulin isophane, human 70 UNT/ML / insulin, regular, human 30 UNT/ML Injectable Suspension [Humulin] |
| RxNorm | 106894 | insulin isophane, human 85 UNT/ML / insulin, regular, human 15 UNT/ML Injectable Suspension [Pur-In Mix] |

Continued on next page

**Table25 – continued from previous page**

| CodeSystem | ConceptCode | ConceptName |
|---|---|---|
| RxNorm | 106895 | insulin isophane, human 75 UNT/ML / insulin, regular, human 25 UNT/ML Injectable Suspension [Pur-In Mix] |
| RxNorm | 106896 | insulin isophane, human 50 UNT/ML / insulin, regular, human 50 UNT/ML Injectable Suspension [Pur-In Mix] |
| RxNorm | 106899 | insulin isophane, human 90 UNT/ML / insulin, regular, human 10 UNT/ML Injectable Suspension [Humulin] |
| RxNorm | 106900 | insulin isophane, human 80 UNT/ML / insulin, regular, human 20 UNT/ML Injectable Suspension [Humulin] |
| RxNorm | 106901 | insulin isophane, human 60 UNT/ML / insulin, regular, human 40 UNT/ML Injectable Suspension [Humulin] |
| RxNorm | 108407 | insulin isophane, human 50 UNT/ML / insulin, regular, human 50 UNT/ML Injectable Suspension [Humulin] |
| RxNorm | 1164824 | insulin, regular, human Injectable Product |
| RxNorm | 1171292 | Human Actrapid Injectable Product |
| RxNorm | 1171293 | Human Actrapid Penfill Injectable Product |
| RxNorm | 1171296 | Human Mixtard Injectable Product |
| RxNorm | 1171997 | Humulin Injectable Product |
| RxNorm | 1172692 | Humulin R Injectable Product |
| RxNorm | 1172693 | Humulin S Injectable Product |
| RxNorm | 379742 | Insulin, Isophane, Human / Insulin, Regular, Human Injectable Suspension [Human Mixtard 30 ge] |
| RxNorm | 365582 | Insulin, Isophane, Human / Insulin, Regular, Human Injectable Suspension [Humulin M3] |
| RxNorm | 365576 | Insulin, Isophane, Human / Insulin, Regular, Human Injectable Suspension [Humulin M4] |
| RxNorm | 365684 | Insulin, Isophane, Human / Insulin, Regular, Human Injectable Suspension [Pur-In Mix 25/75] |
| RxNorm | 575139 | Insulin, Isophane, Human 70 UNT/ML / Insulin, Regular, Human 30 UNT/ML [Novolin 70/30 PenFill] |
| RxNorm | 724341 | Insulin, Isophane, Human / Insulin, Regular, Human Injectable Suspension [ReliOn/Novolin 70/30] |
| RxNorm | 727909 | 1.5 ML Insulin, Isophane, Human 70 UNT/ML / Insulin, Regular, Human 30 UNT/ML Prefilled Syringe |
| RxNorm | 727917 | 3 ML Insulin, Regular, Human 100 UNT/ML Prefilled Syringe |
| RxNorm | 727770 | 3 ML Insulin, Isophane, Human 50 UNT/ML / Insulin, Regular, Human 50 UNT/ML Prefilled Syringe |
| RxNorm | 727766 | 3 ML Insulin, Isophane, Human 85 UNT/ML / Insulin, Regular, Human 15 UNT/ML Prefilled Syringe |
| RxNorm | 727626 | Insulin, Isophane, Human / Insulin, Regular, Human Prefilled Syringe |
| RxNorm | 723552 | Insulin, Regular, Human Injectable Solution [ReliOn R/Novolin] |

Continued on next page

**Table25 – continued from previous page**

| CodeSystem | ConceptCode | ConceptName |
|---|---|---|
| RxNorm | 727895 | Insulin, Isophane, Human / Insulin, Regular, Human Prefilled Syringe [Humulin 70/30] |
| RxNorm | 727916 | Insulin, Regular, Human Prefilled Syringe |
| RxNorm | 727769 | 3 ML Insulin, Isophane, Human 60 UNT/ML / Insulin, Regular, Human 40 UNT/ML Prefilled Syringe |
| RxNorm | 727918 | 1.5 ML Insulin, Regular, Human 100 UNT/ML Prefilled Syringe |
| RxNorm | 727627 | 3 ML Insulin, Isophane, Human 70 UNT/ML / Insulin, Regular, Human 30 UNT/ML Prefilled Syringe |
| RxNorm | 727755 | 3 ML Insulin, Isophane, Human 90 UNT/ML / Insulin, Regular, Human 10 UNT/ML Prefilled Syringe |
| RxNorm | 727628 | Insulin, Isophane, Human / Insulin, Regular, Human Prefilled Syringe [Novolin 70/30] |
| RxNorm | 727896 | 3 ML Insulin, Isophane, Human 70 UNT/ML / Insulin, Regular, Human 30 UNT/ML Prefilled Syringe [Humulin 70/30] |
| RxNorm | 724342 | Insulin, Isophane, Human 70 UNT/ML / Insulin, Regular, Human 30 UNT/ML Injectable Suspension [ReliOn/Novolin 70/30] |
| RxNorm | 727629 | 3 ML Insulin, Isophane, Human 70 UNT/ML / Insulin, Regular, Human 30 UNT/ML Prefilled Syringe [Novolin 70/30] |
| RxNorm | 727768 | 3 ML Insulin, Isophane, Human 75 UNT/ML / Insulin, Regular, Human 25 UNT/ML Prefilled Syringe |
| RxNorm | 723553 | Insulin, Regular, Human 100 UNT/ML Injectable Solution [ReliOn R/Novolin] |
| RxNorm | 727910 | 1.5 ML Insulin, Isophane, Human 70 UNT/ML / Insulin, Regular, Human 30 UNT/ML Prefilled Syringe [Novolin 70/30] |
| RxNorm | 723551 | Insulin, Regular, Human 100 UNT/ML [ReliOn R/Novolin] |
| RxNorm | 727767 | 3 ML Insulin, Isophane, Human 80 UNT/ML / Insulin, Regular, Human 20 UNT/ML Prefilled Syringe |
| RxNorm | 724340 | Insulin, Isophane, Human 70 UNT/ML / Insulin, Regular, Human 30 UNT/ML [ReliOn/Novolin 70/30] |
| RxNorm | 1172687 | Humulin M2 Injectable Product |
| RxNorm | 1172688 | Humulin M3 Injectable Product |
| RxNorm | 1156396 | NPH Insulin, Human / Regular Insulin, Human Injectable Product |
| RxNorm | 1172689 | Humulin M4 Injectable Product |
| RxNorm | 1171298 | Human Mixtard 50 Injectable Product |
| RxNorm | 1172690 | Humulin M5 Injectable Product |
| RxNorm | 1183567 | Pur-In Mix 50/50 Injectable Product |
| RxNorm | 1183565 | Pur-In Mix 15/85 Injectable Product |
| RxNorm | 1172683 | Humulin 70/30 Injectable Product |
| RxNorm | 1172686 | Humulin M1 Injectable Product |

Continued on next page

**Table25 – continued from previous page**

| CodeSystem | ConceptCode | ConceptName |
|---|---|---|
| RxNorm | 1171299 | Human Mixtard Penfill Injectable Product |
| RxNorm | 1171297 | Human Mixtard 30 ge Injectable Product |
| RxNorm | 1183566 | Pur-In Mix Injectable Product |
| RxNorm | 1183568 | Pur-In Neutral Injectable Product |
| RxNorm | 1178125 | Novolin Injectable Product |
| RxNorm | 1178128 | Novolin R Injectable Product |
| RxNorm | 1187762 | Velosulin Injectable Product |
| RxNorm | 1359720 | insulin isophane, human 70 UNT/ML / insulin, regular, human 30 UNT/ML Prefilled Syringe [Humulin] |
| RxNorm | 1360172 | insulin isophane, human 70 UNT/ML / insulin, regular, human 30 UNT/ML Prefilled Syringe |
| RxNorm | 1360226 | insulin isophane, human 70 UNT/ML / insulin, regular, human 30 UNT/ML Prefilled Syringe [Novolin] |
| RxNorm | 1360482 | insulin, regular, human 100 UNT/ML Prefilled Syringe |
| RxNorm | 1360435 | insulin, regular, human 100 UNT/ML Prefilled Syringe [Novolin R] |
| RxNorm | 1372685 | Human Mixtard |
| RxNorm | 1372723 | Novolin |
| RxNorm | 1372744 | Humulin |
| RxNorm | 1372761 | Pur-In Mix |
| RxNorm | 150659 | insulin, regular, human 100 UNT/ML Injectable Solution [Pur-In Neutral] |
| RxNorm | 150831 | insulin, regular, human 100 UNT/ML Injectable Solution [Human Actrapid Penfill] |
| RxNorm | 150973 | insulin, regular, human 100 UNT/ML Injectable Solution [Human Actrapid] |
| RxNorm | 150974 | insulin, regular, human 100 UNT/ML Injectable Solution [Humulin S] |
| RxNorm | 1544488 | insulin, regular, human 8 UNT Inhalation Powder |
| RxNorm | 1544490 | insulin, regular, human 8 UNT Inhalation Powder [Afrezza] |
| RxNorm | 1544568 | {60 (insulin, regular, human 4 UNT Inhalation Powder) / 30 (insulin, regular, human 8 UNT Inhalation Powder) } Pack |
| RxNorm | 1544569 | {60 (insulin, regular, human 4 UNT Inhalation Powder [Afrezza]) / 30 (insulin, regular, human 8 UNT Inhalation Powder [Afrezza]) } Pack [Afrezza 90 - 60 (4 UNT), 30 (8 UNT)] |
| RxNorm | 1544570 | {30 (insulin, regular, human 4 UNT Inhalation Powder) / 60 (insulin, regular, human 8 UNT Inhalation Powder) } Pack |

Continued on next page

**Table25 – continued from previous page**

| CodeSystem | ConceptCode | ConceptName |
|---|---|---|
| RxNorm | 1544571 | {30 (insulin, regular, human 4 UNT Inhalation Powder [Afrezza]) / 60 (insulin, regular, human 8 UNT Inhalation Powder [Afrezza]) } Pack [Afrezza 90 - 30 (4 UNT), 60 (8 UNT)] |
| RxNorm | 1543200 | insulin, regular, human Inhalant Product |
| RxNorm | 1543202 | insulin, regular, human 4 UNT Inhalation Powder |
| RxNorm | 1543203 | Afrezza |
| RxNorm | 1543206 | Afrezza Inhalant Product |
| RxNorm | 1543207 | insulin, regular, human 4 UNT Inhalation Powder [Afrezza] |
| RxNorm | 152644 | Human Actrapid |
| RxNorm | 1650256 | insulin, regular, human 4 UNT |
| RxNorm | 1650260 | insulin, regular, human 4 UNT [Afrezza] |
| RxNorm | 1650262 | insulin, regular, human 8 UNT |
| RxNorm | 1650264 | insulin, regular, human 8 UNT [Afrezza] |
| RxNorm | 1653899 | insulin isophane / insulin, regular, human Injectable Product |
| RxNorm | 1654651 | insulin isophane / insulin, regular, human Pen Injector |
| RxNorm | 1654855 | insulin isophane / insulin, regular, human Pen Injector [Humulin] |
| RxNorm | 1654857 | insulin isophane, human 70 UNT/ML / insulin, regular, human 30 UNT/ML Pen Injector |
| RxNorm | 1654858 | insulin isophane, human 70 UNT/ML / insulin, regular, human 30 UNT/ML Pen Injector [Humulin] |
| RxNorm | 1654909 | insulin, regular, human 12 UNT |
| RxNorm | 1654910 | insulin, regular, human 12 UNT Inhalation Powder |
| RxNorm | 1654911 | insulin, regular, human 12 UNT [Afrezza] |
| RxNorm | 1654912 | insulin, regular, human 12 UNT Inhalation Powder [Afrezza] |
| RxNorm | 1656705 | {30 (insulin, regular, human 12 UNT Inhalation Powder) / 60 (insulin, regular, human 8 UNT Inhalation Powder) } Pack |
| RxNorm | 1656706 | {30 (insulin, regular, human 12 UNT Inhalation Powder [Afrezza]) / 60 (insulin, regular, human 8 UNT Inhalation Powder [Afrezza]) } Pack [Afrezza 90 - 60 (8 UNT), 30 (12 UNT)] |
| RxNorm | 1731314 | insulin, regular, human Pen Injector |
| RxNorm | 1731315 | 3 ML insulin, regular, human 500 UNT/ML Pen Injector |
| RxNorm | 1731316 | insulin, regular, human Pen Injector [Humulin R] |
| RxNorm | 1731317 | 3 ML insulin, regular, human 500 UNT/ML Pen Injector [Humulin R] |
| RxNorm | 1731318 | insulin, regular, human 500 UNT/ML Pen Injector |
| RxNorm | 1731319 | insulin, regular, human 500 UNT/ML Pen Injector [Humulin R] |

Continued on next page

**Table25 – continued from previous page**

| CodeSystem | ConceptCode | ConceptName |
|---|---|---|
| RxNorm | 1798387 | {90 (insulin, regular, human 4 UNT Inhalation Powder) / 90 (insulin, regular, human 8 UNT Inhalation Powder) } Pack |
| RxNorm | 1798388 | {90 (insulin, regular, human 4 UNT Inhalation Powder [Afrezza]) / 90 (insulin, regular, human 8 UNT Inhalation Powder [Afrezza]) } Pack [Afrezza Titration Pack] |
| RxNorm | 1862101 | {60 (insulin, regular, human 12 UNT Inhalation Powder) / 60 (insulin, regular, human 4 UNT Inhalation Powder) / 60 (insulin, regular, human 8 UNT Inhalation Powder) } Pack |
| RxNorm | 1862102 | {60 (insulin, regular, human 12 UNT Inhalation Powder [Afrezza]) / 60 (insulin, regular, human 4 UNT Inhalation Powder [Afrezza]) / 60 (insulin, regular, human 8 UNT Inhalation Powder [Afrezza]) } Pack [Afrezza 180 Titration Pack- 60 (4 UNT), 60 (8 UNT), 60 (12 UNT)] |
| RxNorm | 203209 | Human Actrapid Penfill |
| RxNorm | 2049379 | insulin isophane / insulin, regular, human Pen Injector [Novolin] |
| RxNorm | 2049380 | 3 ML insulin isophane, human 70 UNT/ML / insulin, regular, human 30 UNT/ML Pen Injector [Novolin] |
| RxNorm | 2049381 | insulin isophane, human 70 UNT/ML / insulin, regular, human 30 UNT/ML Pen Injector [Novolin] |
| RxNorm | 2100028 | {90 (insulin, regular, human 12 UNT Inhalation Powder) / 90 (insulin, regular, human 8 UNT Inhalation Powder) } Pack |
| RxNorm | 2100029 | {90 (insulin, regular, human 12 UNT Inhalation Powder [Afrezza]) / 90 (insulin, regular, human 8 UNT Inhalation Powder [Afrezza]) } Pack [Afrezza 180 - 90 (8 UNT), 90 (12 UNT)] |
| RxNorm | 2108525 | insulin, regular, human Inhalation Powder |
| RxNorm | 2108527 | insulin, regular, human Inhalation Powder [Afrezza] |
| RxNorm | 213442 | insulin isophane, human 70 UNT/ML / insulin, regular, human 30 UNT/ML Injectable Suspension [Novolin] |
| RxNorm | 2179742 | insulin, regular, human 1 UNT/ML |
| RxNorm | 2179743 | insulin, regular, human Injection |
| RxNorm | 2179744 | 100 ML insulin, regular, human 1 UNT/ML Injection |
| RxNorm | 2179745 | Myxredlin |
| RxNorm | 2179746 | insulin, regular, human 1 UNT/ML [Myxredlin] |
| RxNorm | 2179747 | insulin, regular, human Injection [Myxredlin] |
| RxNorm | 2179748 | Myxredlin Injectable Product |
| RxNorm | 2179749 | 100 ML insulin, regular, human 1 UNT/ML Injection [Myxredlin] |
| RxNorm | 2179750 | insulin, regular, human 1 UNT/ML Injection |
| RxNorm | 2179751 | insulin, regular, human 1 UNT/ML Injection [Myxredlin] |

Continued on next page

**Table25 – continued from previous page**

| CodeSystem | ConceptCode | ConceptName |
|---|---|---|
| RxNorm | 2206090 | 3 ML insulin, regular, human 100 UNT/ML Pen Injector |
| RxNorm | 2206091 | insulin, regular, human Pen Injector [Novolin R] |
| RxNorm | 2206092 | 3 ML insulin, regular, human 100 UNT/ML Pen Injector [Novolin R] |
| RxNorm | 2206093 | insulin, regular, human 100 UNT/ML Pen Injector |
| RxNorm | 2206094 | insulin, regular, human 100 UNT/ML Pen Injector [Novolin R] |
| RxNorm | 247511 | insulin isophane, human 90 UNT/ML / insulin, regular, human 10 UNT/ML Injectable Suspension |
| RxNorm | 247512 | insulin isophane, human 80 UNT/ML / insulin, regular, human 20 UNT/ML Injectable Suspension |
| RxNorm | 247513 | insulin isophane, human 60 UNT/ML / insulin, regular, human 40 UNT/ML Injectable Suspension |
| RxNorm | 249026 | insulin, regular, human 40 UNT/ML Injectable Solution |
| RxNorm | 245264 | insulin isophane, human 85 UNT/ML / insulin, regular, human 15 UNT/ML Injectable Suspension |
| RxNorm | 245265 | insulin isophane, human 50 UNT/ML / insulin, regular, human 50 UNT/ML Injectable Suspension |
| RxNorm | 249220 | insulin, regular, human 500 UNT/ML Injectable Solution |
| RxNorm | 311033 | insulin, regular, human 100 UNT/ML Injectable Solution [Novolin R] |
| RxNorm | 311034 | insulin, regular, human 100 UNT/ML Injectable Solution |
| RxNorm | 311035 | insulin, regular, human 100 UNT/ML Injectable Solution [Velosulin] |
| RxNorm | 311036 | insulin, regular, human 100 UNT/ML Injectable Solution [Humulin R] |
| RxNorm | 311048 | insulin isophane, human 70 UNT/ML / insulin, regular, human 30 UNT/ML Injectable Suspension |
| RxNorm | 340325 | insulin, regular, human 100 UNT/ML |
| RxNorm | 340326 | insulin, regular, human 40 UNT/ML |
| RxNorm | 340327 | insulin, regular, human 500 UNT/ML |
| RxNorm | 351859 | insulin, regular, human 500 UNT/ML Injectable Solution [Humulin R] |
| RxNorm | 362622 | insulin, regular, human Injectable Solution [Humulin R] |
| RxNorm | 363120 | insulin, regular, human Injectable Solution [Velosulin] |
| RxNorm | 363221 | insulin, regular, human Injectable Solution [Novolin R] |
| RxNorm | 343083 | insulin, regular, human 30 UNT/ML |
| RxNorm | 343258 | insulin, regular, human 50 UNT/ML |
| RxNorm | 360894 | insulin, regular, human 25 UNT/ML |
| RxNorm | 359125 | insulin, regular, human 15 UNT/ML |
| RxNorm | 359126 | insulin, regular, human 10 UNT/ML |
| RxNorm | 359127 | insulin, regular, human 20 UNT/ML |

Continued on next page

**Table25 – continued from previous page**

| CodeSystem | ConceptCode | ConceptName |
|---|---|---|
| RxNorm | 365668 | insulin isophane / insulin, regular, human Injectable Suspension [Pur-In Mix] |
| RxNorm | 365672 | insulin isophane / insulin, regular, human Injectable Suspension [Novolin] |
| RxNorm | 365679 | insulin isophane / insulin, regular, human Injectable Suspension [Humulin] |
| RxNorm | 392660 | insulin isophane, human 75 UNT/ML / insulin, regular, human 25 UNT/ML Injectable Suspension |
| RxNorm | 378857 | insulin isophane / insulin, regular, human Injectable Suspension |
| RxNorm | 376915 | insulin, regular, human Injectable Solution |
| RxNorm | 412978 | insulin isophane, human 16 UNT/ML / insulin, regular, human 24 UNT/ML Injectable Suspension |
| RxNorm | 379734 | insulin, regular, human Injectable Solution [Humulin S] |
| RxNorm | 379740 | insulin isophane / insulin, regular, human Injectable Suspension [Human Mixtard] |
| RxNorm | 379744 | insulin, regular, human Injectable Solution [Human Actrapid] |
| RxNorm | 379747 | insulin, regular, human Injectable Solution [Human Actrapid Penfill] |
| RxNorm | 385895 | Pur-In Neutral |
| RxNorm | 385896 | insulin, regular, human Injectable Solution [Pur-In Neutral] |
| RxNorm | 412453 | insulin, regular, human 5 UNT/ML Injectable Solution |
| RxNorm | 415088 | insulin isophane, human 28 UNT/ML / insulin, regular, human 12 UNT/ML Injectable Suspension |
| RxNorm | 415089 | insulin isophane, human 30 UNT/ML / insulin, regular, human 10 UNT/ML Injectable Suspension |
| RxNorm | 415090 | insulin isophane, human 34 UNT/ML / insulin, regular, human 6 UNT/ML Injectable Suspension |
| RxNorm | 415184 | insulin isophane, human 32 UNT/ML / insulin, regular, human 8 UNT/ML Injectable Suspension |
| RxNorm | 415185 | insulin isophane, human 36 UNT/ML / insulin, regular, human 4 UNT/ML Injectable Suspension |
| RxNorm | 440399 | insulin, regular, human 24 UNT/ML |
| RxNorm | 440650 | insulin, regular, human 12 UNT/ML |
| RxNorm | 440653 | insulin, regular, human 6 UNT/ML |
| RxNorm | 440654 | insulin, regular, human 5 UNT/ML |
| RxNorm | 451437 | insulin, regular, human 8 UNT/ML |
| RxNorm | 451439 | insulin, regular, human 4 UNT/ML |
| RxNorm | 564395 | insulin isophane, human 85 UNT/ML / insulin, regular, human 15 UNT/ML [Pur-In Mix] |
| RxNorm | 564396 | insulin isophane, human 75 UNT/ML / insulin, regular, human 25 UNT/ML [Pur-In Mix] |

Continued on next page

**Table25 – continued from previous page**

| CodeSystem | ConceptCode | ConceptName |
|---|---|---|
| RxNorm | 564397 | insulin isophane, human 50 UNT/ML / insulin, regular, human 50 UNT/ML [Pur-In Mix] |
| RxNorm | 564399 | insulin isophane, human 90 UNT/ML / insulin, regular, human 10 UNT/ML [Humulin] |
| RxNorm | 564400 | insulin isophane, human 80 UNT/ML / insulin, regular, human 20 UNT/ML [Humulin] |
| RxNorm | 564401 | insulin isophane, human 60 UNT/ML / insulin, regular, human 40 UNT/ML [Humulin] |
| RxNorm | 5459 | Humulin S |
| RxNorm | 564531 | insulin isophane, human 50 UNT/ML / insulin, regular, human 50 UNT/ML [Humulin] |
| RxNorm | 564601 | insulin isophane, human 90 UNT/ML / insulin, regular, human 10 UNT/ML [Human Mixtard] |
| RxNorm | 564602 | insulin isophane, human 80 UNT/ML / insulin, regular, human 20 UNT/ML [Human Mixtard] |
| RxNorm | 564603 | insulin isophane, human 70 UNT/ML / insulin, regular, human 30 UNT/ML [Human Mixtard] |
| RxNorm | 564605 | insulin isophane, human 60 UNT/ML / insulin, regular, human 40 UNT/ML [Human Mixtard] |
| RxNorm | 564766 | insulin, regular, human 100 UNT/ML [Pur-In Neutral] |
| RxNorm | 564820 | insulin, regular, human 100 UNT/ML [Human Actrapid Penfill] |
| RxNorm | 564881 | insulin, regular, human 100 UNT/ML [Human Actrapid] |
| RxNorm | 564882 | insulin, regular, human 100 UNT/ML [Humulin S] |
| RxNorm | 565176 | insulin isophane, human 50 UNT/ML / insulin, regular, human 50 UNT/ML [Human Mixtard] |
| RxNorm | 575146 | insulin, regular, human 100 UNT/ML [Novolin R] |
| RxNorm | 575147 | insulin, regular, human 100 UNT/ML [Velosulin] |
| RxNorm | 575148 | insulin, regular, human 100 UNT/ML [Humulin R] |
| RxNorm | 575628 | insulin, regular, human 500 UNT/ML [Humulin R] |
| RxNorm | 573330 | insulin isophane, human 70 UNT/ML / insulin, regular, human 30 UNT/ML [Humulin] |
| RxNorm | 573331 | insulin isophane, human 70 UNT/ML / insulin, regular, human 30 UNT/ML [Novolin] |
| RxNorm | 847186 | insulin isophane / insulin, regular, human Prefilled Syringe |
| RxNorm | 847187 | 3 ML insulin isophane, human 70 UNT/ML / insulin, regular, human 30 UNT/ML Pen Injector |
| RxNorm | 847188 | insulin isophane / insulin, regular, human Prefilled Syringe [Humulin] |
| RxNorm | 847189 | 3 ML insulin isophane, human 70 UNT/ML / insulin, regular, human 30 UNT/ML Pen Injector [Humulin] |
| RxNorm | 847194 | insulin isophane / insulin, regular, human Prefilled Syringe [Novolin] |

Continued on next page

**Table25 – continued from previous page**

| CodeSystem | ConceptCode | ConceptName |
|---|---|---|
| RxNorm | 847202 | insulin, regular, human Prefilled Syringe |
| RxNorm | 847203 | 3 ML insulin, regular, human 100 UNT/ML Prefilled Syringe |
| RxNorm | 847204 | insulin, regular, human Prefilled Syringe [Novolin R] |
| RxNorm | 847205 | 3 ML insulin, regular, human 100 UNT/ML Prefilled Syringe [Novolin R] |
| RxNorm | 847256 | 1.5 ML insulin isophane, human 70 UNT/ML / insulin, regular, human 30 UNT/ML Prefilled Syringe |
| RxNorm | 847257 | 1.5 ML insulin isophane, human 70 UNT/ML / insulin, regular, human 30 UNT/ML Prefilled Syringe [Novolin] |
| RxNorm | 847343 | 1.5 ML insulin isophane, human 70 UNT/ML / insulin, regular, human 30 UNT/ML Prefilled Syringe [Humulin] |
| RxNorm | 847417 | 1.5 ML insulin, regular, human 100 UNT/ML Prefilled Syringe |
| RxNorm | 92881 | Humulin R |
| RxNorm | 93560 | Novolin R |

##### 8.5.3 Sulfonylurea

Table 26: Concept codes used to identify if a patient was prescribed sulfonylurea medications.

| CodeSystem | ConceptCode | ConceptName |
|---|---|---|
| RxNorm | 10633 | tolazamide |
| RxNorm | 25789 | glimepiride |
| RxNorm | 4815 | glyburide |
| RxNorm | 4821 | glipizide |
| RxNorm | 284433 | Glyburide 5 MG / Metformin 500 MG Oral Tablet [Glucovance] |
| RxNorm | 284432 | Glyburide 2.5 MG / Metformin 500 MG Oral Tablet [Glucovance] |
| RxNorm | 351273 | Glipizide 2.5 MG / Metformin 500 MG Oral Tablet |
| RxNorm | 351275 | Glipizide 2.5 MG / Metformin 250 MG Oral Tablet |
| RxNorm | 261311 | Glipizide 2.5 MG 24 Hour Extended Release Tablet [Glucotrol XL] |
| RxNorm | 205829 | Glipizide 5 MG 24 Hour Extended Release Tablet [Glucotrol XL] |
| RxNorm | 284431 | Glyburide 1.25 MG / Metformin 250 MG Oral Tablet [Glucovance] |
| RxNorm | 205881 | Glyburide 6 MG Oral Tablet [Glynase Pres-Tab] |
| RxNorm | 205877 | Glyburide 3 MG Oral Tablet [Glynase Pres-Tab] |

Continued on next page

**Table26 – continued from previous page**

| CodeSystem | ConceptCode | ConceptName |
|---|---|---|
| RxNorm | 205831 | Glipizide 10 MG 24 Hour Extended Release Tablet [Glucotrol XL] |
| RxNorm | 205874 | Glyburide 1.5 MG Oral Tablet [Glynase Pres-Tab] |
| RxNorm | 351274 | Glipizide 5 MG / Metformin 500 MG Oral Tablet |
| RxNorm | 566719 | Glipizide 5 MG [Glucotrol XL] |
| RxNorm | 566721 | Glipizide 10 MG [Glucotrol XL] |
| RxNorm | 574533 | Glipizide 2.5 MG [Glucotrol XL] |
| RxNorm | 566763 | Glyburide 1.5 MG [Glynase Pres-Tab] |
| RxNorm | 574869 | Glyburide 1.25 MG / Metformin 250 MG [Glucovance] |
| RxNorm | 566770 | Glyburide 6 MG [Glynase Pres-Tab] |
| RxNorm | 574871 | Glyburide 5 MG / Metformin 500 MG [Glucovance] |
| RxNorm | 566766 | Glyburide 3 MG [Glynase Pres-Tab] |
| RxNorm | 574870 | Glyburide 2.5 MG / Metformin 500 MG [Glucovance] |
| RxNorm | 310538 | Glyburide 5 MG / Metformin 500 MG Oral Tablet |
| RxNorm | 310535 | Glyburide 2.5 MG / Metformin 500 MG Oral Tablet |
| RxNorm | 310533 | Glyburide 1.25 MG / Metformin 250 MG Oral Tablet |
| RxNorm | 368567 | Glipizide / Metformin Oral Tablet [metaglip] |
| RxNorm | 352301 | Glipizide 5 MG / Metformin 500 MG Oral Tablet [metaglip] |
| RxNorm | 352302 | Glipizide 2.5 MG / Metformin 250 MG Oral Tablet [metaglip] |
| RxNorm | 379801 | Glipizide 24 Hour Extended Release Tablet [Glucotrol XL] |
| RxNorm | 369372 | Glyburide Oral Tablet [Glynase Pres-Tab] |
| RxNorm | 379805 | Glipizide 24 Hour Extended Release Tablet |
| RxNorm | 368169 | Glyburide / Metformin Oral Tablet [Glucovance] |
| RxNorm | 352300 | Glipizide 2.5 MG / Metformin 500 MG Oral Tablet [metaglip] |
| RxNorm | 1008873 | glyburide / phenformin |
| RxNorm | 102845 | glyburide 5 MG Oral Tablet [Daonil] |
| RxNorm | 105371 | glyburide 2.5 MG Oral Tablet [Calabren] |
| RxNorm | 105372 | glyburide 5 MG Oral Tablet [Calabren] |
| RxNorm | 105373 | glipizide 5 MG Oral Tablet [Minidiab] |
| RxNorm | 1157121 | tolazamide Oral Product |
| RxNorm | 1157122 | tolazamide Pill |
| RxNorm | 1156197 | glyburide / metformin Pill |
| RxNorm | 1156198 | glyburide / phenformin Oral Product |
| RxNorm | 1156199 | glyburide / phenformin Pill |
| RxNorm | 1156200 | glyburide Oral Product |
| RxNorm | 1156201 | glyburide Pill |
| RxNorm | 1157240 | glimepiride / pioglitazone Oral Product |
| RxNorm | 1157241 | glimepiride / pioglitazone Pill |
| RxNorm | 1157242 | glimepiride / rosiglitazone Oral Product |
| RxNorm | 1157243 | glimepiride / rosiglitazone Pill |
| RxNorm | 1157244 | glimepiride Oral Product |

Continued on next page

**Table26 – continued from previous page**

| CodeSystem | ConceptCode | ConceptName |
|---|---|---|
| RxNorm | 1157245 | glimepiride Pill |
| RxNorm | 1166403 | Duetact Oral Product |
| RxNorm | 1166404 | Duetact Pill |
| RxNorm | 1165205 | glipizide / metformin Oral Product |
| RxNorm | 1165206 | glipizide / metformin Pill |
| RxNorm | 1165207 | glipizide Oral Product |
| RxNorm | 1165208 | glipizide Pill |
| RxNorm | 1165845 | glyburide / metformin Oral Product |
| RxNorm | 1170663 | Amaryl Oral Product |
| RxNorm | 1170664 | Amaryl Pill |
| RxNorm | 1170866 | Calabren Oral Product |
| RxNorm | 1170867 | Calabren Pill |
| RxNorm | 1171148 | Euglucon Oral Product |
| RxNorm | 1171149 | Euglucon Pill |
| RxNorm | 1171233 | Glibenese Oral Product |
| RxNorm | 1171234 | Glibenese Pill |
| RxNorm | 1171246 | Glucotrol Oral Product |
| RxNorm | 1171247 | Glucotrol Pill |
| RxNorm | 1171248 | Glucovance Oral Product |
| RxNorm | 1171249 | Glucovance Pill |
| RxNorm | 1171929 | Glycron Oral Product |
| RxNorm | 1171930 | Glycron Pill |
| RxNorm | 1171933 | Glynase Oral Product |
| RxNorm | 1171934 | Glynase Pill |
| RxNorm | 1171919 | Glyburase Oral Product |
| RxNorm | 1171920 | Glyburase Pill |
| RxNorm | 1175658 | Avandaryl Oral Product |
| RxNorm | 1175659 | Avandaryl Pill |
| RxNorm | 1175878 | Diabeta Oral Product |
| RxNorm | 1175879 | Diabeta Pill |
| RxNorm | 1175880 | Diabetamide Oral Product |
| RxNorm | 1175881 | Diabetamide Pill |
| RxNorm | 1168162 | Daonil Oral Product |
| RxNorm | 1168163 | Daonil Pill |
| RxNorm | 1179291 | Minidiab Oral Product |
| RxNorm | 1179292 | Minidiab Pill |
| RxNorm | 1185049 | Metaglip Oral Product |
| RxNorm | 1179740 | Malix Oral Product |
| RxNorm | 1179741 | Malix Pill |
| RxNorm | 1177973 | Tolinase Oral Product |
| RxNorm | 1177974 | Tolinase Pill |
| RxNorm | 1186169 | Libanil Oral Product |
| RxNorm | 1186170 | Libanil Pill |

Continued on next page

**Table26 – continued from previous page**

| CodeSystem | ConceptCode | ConceptName |
|---|---|---|
| RxNorm | 1178082 | Micronase Oral Product |
| RxNorm | 1178083 | Micronase Pill |
| RxNorm | 1178368 | Semi-Daonil Oral Product |
| RxNorm | 1178369 | Semi-Daonil Pill |
| RxNorm | 1185624 | Metaglip Pill |
| RxNorm | 1361492 | glimepiride 6 MG |
| RxNorm | 1361493 | glimepiride 6 MG Oral Tablet |
| RxNorm | 1361494 | glimepiride 8 MG |
| RxNorm | 1361495 | glimepiride 8 MG Oral Tablet |
| RxNorm | 151466 | Calabren |
| RxNorm | 151615 | Diabetamide |
| RxNorm | 151725 | Euglucon |
| RxNorm | 151822 | Glibenese |
| RxNorm | 153842 | glimepiride 3 MG Oral Tablet |
| RxNorm | 153843 | glimepiride 1 MG Oral Tablet [Amaryl] |
| RxNorm | 153844 | glimepiride 3 MG Oral Tablet [Amaryl] |
| RxNorm | 153845 | glimepiride 4 MG Oral Tablet [Amaryl] |
| RxNorm | 152651 | Malix |
| RxNorm | 153591 | glimepiride 2 MG Oral Tablet [Amaryl] |
| RxNorm | 153592 | Amaryl |
| RxNorm | 197737 | glyburide 1.25 MG Oral Tablet |
| RxNorm | 199245 | glimepiride 1 MG Oral Tablet |
| RxNorm | 199246 | glimepiride 2 MG Oral Tablet |
| RxNorm | 199247 | glimepiride 4 MG Oral Tablet |
| RxNorm | 198291 | tolazamide 100 MG Oral Tablet |
| RxNorm | 198292 | tolazamide 250 MG Oral Tablet |
| RxNorm | 198293 | tolazamide 500 MG Oral Tablet |
| RxNorm | 203289 | Micronase |
| RxNorm | 203295 | Diabeta |
| RxNorm | 203296 | Daonil |
| RxNorm | 201921 | glipizide 5 MG Oral Tablet [Glibenese] |
| RxNorm | 201922 | glipizide 2.5 MG Oral Tablet [Minidiab] |
| RxNorm | 203679 | Minidiab |
| RxNorm | 203680 | Glucotrol |
| RxNorm | 201056 | glyburide 2.5 MG Oral Tablet [Libanil] |
| RxNorm | 201057 | glyburide 2.5 MG Oral Tablet [Semi-Daonil] |
| RxNorm | 201058 | glyburide 2.5 MG Oral Tablet [Euglucon] |
| RxNorm | 201059 | glyburide 2.5 MG Oral Tablet [Malix] |
| RxNorm | 201060 | glyburide 2.5 MG Oral Tablet [Diabetamide] |
| RxNorm | 201061 | glyburide 5 MG Oral Tablet [Libanil] |
| RxNorm | 201062 | glyburide 5 MG Oral Tablet [Euglucon] |
| RxNorm | 201063 | glyburide 5 MG Oral Tablet [Malix] |
| RxNorm | 201064 | glyburide 5 MG Oral Tablet [Diabetamide] |

Continued on next page

**Table26 – continued from previous page**

| CodeSystem | ConceptCode | ConceptName |
|---|---|---|
| RxNorm | 205828 | glipizide 5 MG Oral Tablet [Glucotrol] |
| RxNorm | 205830 | glipizide 10 MG Oral Tablet [Glucotrol] |
| RxNorm | 205872 | glyburide 1.25 MG Oral Tablet [Diabeta] |
| RxNorm | 205873 | glyburide 1.25 MG Oral Tablet [Micronase] |
| RxNorm | 205875 | glyburide 2.5 MG Oral Tablet [Diabeta] |
| RxNorm | 205876 | glyburide 2.5 MG Oral Tablet [Micronase] |
| RxNorm | 205879 | glyburide 5 MG Oral Tablet [Diabeta] |
| RxNorm | 205880 | glyburide 5 MG Oral Tablet [Micronase] |
| RxNorm | 207953 | tolazamide 100 MG Oral Tablet [Tolinase] |
| RxNorm | 207954 | tolazamide 250 MG Oral Tablet [Tolinase] |
| RxNorm | 207955 | tolazamide 500 MG Oral Tablet [Tolinase] |
| RxNorm | 220338 | Tolinase |
| RxNorm | 225613 | Semi-Daonil |
| RxNorm | 246391 | glyburide 2.5 MG / phenformin 25 MG Oral Tablet |
| RxNorm | 246524 | glyburide 5 MG / phenformin 50 MG Oral Tablet |
| RxNorm | 250919 | glyburide 2.5 MG / metformin 400 MG Oral Tablet |
| RxNorm | 284743 | Glucovance |
| RxNorm | 261351 | glyburide 4.5 MG Oral Tablet [Glycron] |
| RxNorm | 252960 | glyburide 4.5 MG Oral Tablet |
| RxNorm | 285129 | glyburide / metformin |
| RxNorm | 261532 | Glycron |
| RxNorm | 261974 | glyburide 3 MG Oral Tablet [Glycron] |
| RxNorm | 260286 | glyburide 1.5 MG Oral Tablet [Glycron] |
| RxNorm | 260287 | glyburide 6 MG Oral Tablet [Glycron] |
| RxNorm | 310488 | glipizide 10 MG Oral Tablet |
| RxNorm | 310489 | 24 HR glipizide 2.5 MG Extended Release Oral Tablet |
| RxNorm | 310490 | glipizide 5 MG Oral Tablet |
| RxNorm | 310534 | glyburide 2.5 MG Oral Tablet |
| RxNorm | 310536 | glyburide 3 MG Oral Tablet |
| RxNorm | 310537 | glyburide 5 MG Oral Tablet |
| RxNorm | 310539 | glyburide 6 MG Oral Tablet |
| RxNorm | 313418 | tolazamide 250 MG Oral Capsule |
| RxNorm | 314000 | glyburide 1.5 MG Oral Tablet |
| RxNorm | 314006 | 24 HR glipizide 5 MG Extended Release Oral Tablet |
| RxNorm | 315107 | 24 HR glipizide 10 MG Extended Release Oral Tablet |
| RxNorm | 316832 | tolazamide 100 MG |
| RxNorm | 316833 | tolazamide 250 MG |
| RxNorm | 316834 | tolazamide 500 MG |
| RxNorm | 315978 | glimepiride 1 MG |
| RxNorm | 315979 | glimepiride 4 MG |
| RxNorm | 315980 | glipizide 5 MG |
| RxNorm | 315987 | glyburide 1.25 MG |
| RxNorm | 315988 | glyburide 1.5 MG |

Continued on next page

**Table26 – continued from previous page**

| CodeSystem | ConceptCode | ConceptName |
|---|---|---|
| RxNorm | 315989 | glyburide 2.5 MG |
| RxNorm | 315990 | glyburide 3 MG |
| RxNorm | 315991 | glyburide 5 MG |
| RxNorm | 315992 | glyburide 6 MG |
| RxNorm | 541898 | Glyburide 1.5 MG Oral Tablet [Glyguride] |
| RxNorm | 542065 | Glyburide 6 MG [Gyburide] |
| RxNorm | 575998 | Glipizide 2.5 MG / Metformin 250 MG [metaglip] |
| RxNorm | 575996 | Glipizide 2.5 MG / Metformin 500 MG [metaglip] |
| RxNorm | 541896 | Glyburide 1.5 MG [Glyguride] |
| RxNorm | 542067 | Glyburide 6 MG Oral Tablet [Gyburide] |
| RxNorm | 542066 | Glyburide Oral Tablet [Gyburide] |
| RxNorm | 541897 | Glyburide Oral Tablet [Glyguride] |
| RxNorm | 575997 | Glipizide 5 MG / Metformin 500 MG [metaglip] |
| RxNorm | 615211 | glimepiride / rosiglitazone Oral Tablet [Avandaryl] |
| RxNorm | 668563 | glimepiride 4 MG / pioglitazone 30 MG Oral Tablet [Duetact] |
| RxNorm | 706933 | glimepiride 2 MG / rosiglitazone 8 MG Oral Tablet [Avandaryl] |
| RxNorm | 668555 | glimepiride / pioglitazone Oral Tablet [Duetact] |
| RxNorm | 615918 | glimepiride 4 MG / rosiglitazone 4 MG [Avandaryl] |
| RxNorm | 615212 | glimepiride 1 MG / rosiglitazone 4 MG Oral Tablet [Avandaryl] |
| RxNorm | 668554 | glimepiride 2 MG / pioglitazone 30 MG [Duetact] |
| RxNorm | 668562 | glimepiride 4 MG / pioglitazone 30 MG [Duetact] |
| RxNorm | 700836 | Glipizide 5 MG Extended Release Tablet |
| RxNorm | 706932 | glimepiride 2 MG / rosiglitazone 8 MG [Avandaryl] |
| RxNorm | 731456 | glimepiride / pioglitazone Oral Tablet [Duetact 30/4] |
| RxNorm | 615214 | glimepiride 2 MG / rosiglitazone 4 MG Oral Tablet [Avandaryl] |
| RxNorm | 615919 | glimepiride 4 MG / rosiglitazone 4 MG Oral Tablet [Avandaryl] |
| RxNorm | 615210 | glimepiride 1 MG / rosiglitazone 4 MG [Avandaryl] |
| RxNorm | 668556 | glimepiride 2 MG / pioglitazone 30 MG Oral Tablet [Duetact] |
| RxNorm | 847723 | glimepiride / rosiglitazone Oral Tablet [Avandaryl 8/4] |
| RxNorm | 847719 | glimepiride / rosiglitazone Oral Tablet [Avandaryl 8/2] |
| RxNorm | 707181 | glimepiride 4 MG / rosiglitazone 8 MG [Avandaryl] |
| RxNorm | 615213 | glimepiride 2 MG / rosiglitazone 4 MG [Avandaryl] |
| RxNorm | 707182 | glimepiride 4 MG / rosiglitazone 8 MG Oral Tablet [Avandaryl] |
| RxNorm | 849593 | Glipizide / Metformin Oral Tablet [Metaglip 2.5 MG/500 MG] |
| RxNorm | 847715 | glimepiride / rosiglitazone Oral Tablet [Avandaryl 4/1] |

Continued on next page

**Table26 – continued from previous page**

| CodeSystem | ConceptCode | ConceptName |
|---|---|---|
| RxNorm | 847711 | glimepiride / rosiglitazone Oral Tablet [Avandaryl 4/2] |
| RxNorm | 849589 | Glipizide / Metformin Oral Tablet [Metaglip 5 MG/500 MG] |
| RxNorm | 861746 | Glyburide / Metformin Oral Tablet [Glucovance 1.25 MG/250 MG] |
| RxNorm | 861751 | Glyburide / Metformin Oral Tablet [Glucovance 2.5 MG/500 MG] |
| RxNorm | 1185627 | Metaglip 5 MG/500 MG Oral Product |
| RxNorm | 1175663 | Avandaryl 8/2 Pill |
| RxNorm | 1175657 | Avandaryl 4/1 Pill |
| RxNorm | 1175661 | Avandaryl 4/4 Pill |
| RxNorm | 1185628 | Metaglip 5 MG/500 MG Pill |
| RxNorm | 1175664 | Avandaryl 8/4 Oral Product |
| RxNorm | 1166405 | Duetact 30/4 Oral Product |
| RxNorm | 1185625 | Metaglip 2.5 MG/500 MG Oral Product |
| RxNorm | 1171251 | Glucovance 2.5 MG/500 MG Pill |
| RxNorm | 1166406 | Duetact 30/4 Pill |
| RxNorm | 1185626 | Metaglip 2.5 MG/500 MG Pill |
| RxNorm | 1175660 | Avandaryl 4/4 Oral Product |
| RxNorm | 1175662 | Avandaryl 8/2 Oral Product |
| RxNorm | 1171253 | Glucovance 5 MG/500 MG Pill |
| RxNorm | 1171250 | Glucovance 2.5 MG/500 MG Oral Product |
| RxNorm | 1175665 | Avandaryl 8/4 Pill |
| RxNorm | 1171252 | Glucovance 5 MG/500 MG Oral Product |
| RxNorm | 1175656 | Avandaryl 4/1 Oral Product |
| RxNorm | 317379 | glipizide 10 MG |
| RxNorm | 317637 | glimepiride 2 MG |
| RxNorm | 331496 | glyburide 4.5 MG |
| RxNorm | 330349 | glipizide 2.5 MG |
| RxNorm | 352381 | glipizide / metformin |
| RxNorm | 352764 | Metaglip |
| RxNorm | 367762 | glimepiride Oral Tablet [Amaryl] |
| RxNorm | 368204 | glyburide Oral Tablet [Glycron] |
| RxNorm | 369304 | tolazamide Oral Tablet [Tolinase] |
| RxNorm | 369373 | glyburide Oral Tablet [Diabeta] |
| RxNorm | 369500 | glipizide Oral Tablet [Glucotrol] |
| RxNorm | 369557 | glyburide Oral Tablet [Semi-Daonil] |
| RxNorm | 369562 | glyburide Oral Tablet [Daonil] |
| RxNorm | 374635 | glyburide / metformin Oral Tablet |
| RxNorm | 375952 | tolazamide Oral Capsule |
| RxNorm | 379803 | glipizide Oral Tablet [Glibenese] |
| RxNorm | 379804 | glipizide 2.5 MG Oral Tablet |
| RxNorm | 380849 | glimepiride 3 MG |

Continued on next page

**Table26 – continued from previous page**

| CodeSystem | ConceptCode | ConceptName |
|---|---|---|
| RxNorm | 372319 | glimepiride Oral Tablet |
| RxNorm | 372320 | glipizide Oral Tablet |
| RxNorm | 372333 | glyburide Oral Tablet |
| RxNorm | 372334 | glyburide / phenformin Oral Tablet |
| RxNorm | 378730 | glipizide / metformin Oral Tablet |
| RxNorm | 374149 | tolazamide Oral Tablet |
| RxNorm | 379559 | glyburide Oral Tablet [Euglucon] |
| RxNorm | 379565 | glyburide Oral Tablet [Diabetamide] |
| RxNorm | 379568 | glyburide Oral Tablet [Malix] |
| RxNorm | 379570 | glyburide Oral Tablet [Calabren] |
| RxNorm | 379571 | Libanil |
| RxNorm | 379572 | glyburide Oral Tablet [Libanil] |
| RxNorm | 379802 | glipizide Oral Tablet [Minidiab] |
| RxNorm | 429841 | glyburide 5 MG / metformin 1000 MG Oral Tablet |
| RxNorm | 430102 | glyburide 3.5 MG Oral Tablet |
| RxNorm | 430103 | glyburide 1.75 MG Oral Tablet |
| RxNorm | 430104 | glyburide 1 MG Oral Tablet |
| RxNorm | 440285 | glyburide 1.75 MG |
| RxNorm | 440286 | glyburide 1 MG |
| RxNorm | 440287 | glyburide 3.5 MG |
| RxNorm | 563154 | glyburide 5 MG [Daonil] |
| RxNorm | 566056 | glipizide 5 MG [Glibenese] |
| RxNorm | 566057 | glipizide 2.5 MG [Minidiab] |
| RxNorm | 565327 | glimepiride 2 MG [Amaryl] |
| RxNorm | 565408 | glimepiride 1 MG [Amaryl] |
| RxNorm | 565409 | glimepiride 3 MG [Amaryl] |
| RxNorm | 565410 | glimepiride 4 MG [Amaryl] |
| RxNorm | 568684 | tolazamide 100 MG [Tolinase] |
| RxNorm | 568685 | tolazamide 250 MG [Tolinase] |
| RxNorm | 568686 | tolazamide 500 MG [Tolinase] |
| RxNorm | 564036 | glyburide 2.5 MG [Calabren] |
| RxNorm | 564037 | glyburide 5 MG [Calabren] |
| RxNorm | 564038 | glipizide 5 MG [Minidiab] |
| RxNorm | 566718 | glipizide 5 MG [Glucotrol] |
| RxNorm | 566720 | glipizide 10 MG [Glucotrol] |
| RxNorm | 566761 | glyburide 1.25 MG [Diabeta] |
| RxNorm | 566762 | glyburide 1.25 MG [Micronase] |
| RxNorm | 566764 | glyburide 2.5 MG [Diabeta] |
| RxNorm | 566765 | glyburide 2.5 MG [Micronase] |
| RxNorm | 566768 | glyburide 5 MG [Diabeta] |
| RxNorm | 566769 | glyburide 5 MG [Micronase] |
| RxNorm | 565667 | glyburide 2.5 MG [Libanil] |
| RxNorm | 565668 | glyburide 2.5 MG [Semi-Daonil] |

Continued on next page

**Table26 – continued from previous page**

| CodeSystem | ConceptCode | ConceptName |
|---|---|---|
| RxNorm | 565669 | glyburide 2.5 MG [Euglucon] |
| RxNorm | 565670 | glyburide 2.5 MG [Malix] |
| RxNorm | 565671 | glyburide 2.5 MG [Diabetamide] |
| RxNorm | 565672 | glyburide 5 MG [Libanil] |
| RxNorm | 565673 | glyburide 5 MG [Euglucon] |
| RxNorm | 565674 | glyburide 5 MG [Malix] |
| RxNorm | 565675 | glyburide 5 MG [Diabetamide] |
| RxNorm | 574612 | glyburide 3 MG [Glycron] |
| RxNorm | 606253 | glimepiride / rosiglitazone |
| RxNorm | 602543 | glimepiride / rosiglitazone Oral Tablet |
| RxNorm | 602544 | glimepiride 1 MG / rosiglitazone 4 MG Oral Tablet |
| RxNorm | 602549 | glimepiride 2 MG / rosiglitazone 4 MG Oral Tablet |
| RxNorm | 602550 | glimepiride 4 MG / rosiglitazone 4 MG Oral Tablet |
| RxNorm | 607816 | Avandaryl |
| RxNorm | 574089 | glyburide 1.5 MG [Glycron] |
| RxNorm | 574090 | glyburide 6 MG [Glycron] |
| RxNorm | 574571 | glyburide 4.5 MG [Glycron] |
| RxNorm | 669980 | Glyburase |
| RxNorm | 669981 | glyburide 1.25 MG [Glyburase] |
| RxNorm | 669982 | glyburide Oral Tablet [Glyburase] |
| RxNorm | 669983 | glyburide 1.25 MG Oral Tablet [Glyburase] |
| RxNorm | 669984 | glyburide 2.5 MG [Glyburase] |
| RxNorm | 669985 | glyburide 2.5 MG Oral Tablet [Glyburase] |
| RxNorm | 669986 | glyburide 5 MG [Glyburase] |
| RxNorm | 669987 | glyburide 5 MG Oral Tablet [Glyburase] |
| RxNorm | 647208 | Duetact |
| RxNorm | 647235 | glimepiride / pioglitazone |
| RxNorm | 647236 | glimepiride / pioglitazone Oral Tablet |
| RxNorm | 647237 | glimepiride 2 MG / pioglitazone 30 MG Oral Tablet |
| RxNorm | 647239 | glimepiride 4 MG / pioglitazone 30 MG Oral Tablet |
| RxNorm | 700835 | glipizide Extended Release Oral Tablet |
| RxNorm | 706895 | glimepiride 2 MG / rosiglitazone 8 MG Oral Tablet |
| RxNorm | 706896 | glimepiride 4 MG / rosiglitazone 8 MG Oral Tablet |
| RxNorm | 731455 | glimepiride 4 MG / pioglitazone 30 MG [Duetact] |
| RxNorm | 731457 | glimepiride 4 MG / pioglitazone 30 MG Oral Tablet [Duetact] |
| RxNorm | 731461 | glimepiride 2 MG / pioglitazone 30 MG [Duetact] |
| RxNorm | 731462 | glimepiride / pioglitazone Oral Tablet [Duetact] |
| RxNorm | 731463 | glimepiride 2 MG / pioglitazone 30 MG Oral Tablet [Duetact] |
| RxNorm | 844809 | glipizide 2.5 MG Extended Release Oral Tablet |
| RxNorm | 844824 | glipizide 5 MG Extended Release Oral Tablet |
| RxNorm | 844827 | glipizide 10 MG Extended Release Oral Tablet |

Continued on next page

**Table26 – continued from previous page**

| CodeSystem | ConceptCode | ConceptName |
|---|---|---|
| RxNorm | 849585 | glipizide / metformin Oral Tablet [Metaglip] |
| RxNorm | 847708 | glimepiride 4 MG / rosiglitazone 4 MG Oral Tablet [Avandaryl] |
| RxNorm | 847706 | glimepiride 4 MG / rosiglitazone 4 MG [Avandaryl] |
| RxNorm | 847707 | glimepiride / rosiglitazone Oral Tablet [Avandaryl] |
| RxNorm | 847710 | glimepiride 2 MG / rosiglitazone 4 MG [Avandaryl] |
| RxNorm | 847712 | glimepiride 2 MG / rosiglitazone 4 MG Oral Tablet [Avandaryl] |
| RxNorm | 847714 | glimepiride 1 MG / rosiglitazone 4 MG [Avandaryl] |
| RxNorm | 847716 | glimepiride 1 MG / rosiglitazone 4 MG Oral Tablet [Avandaryl] |
| RxNorm | 847718 | glimepiride 2 MG / rosiglitazone 8 MG [Avandaryl] |
| RxNorm | 847720 | glimepiride 2 MG / rosiglitazone 8 MG Oral Tablet [Avandaryl] |
| RxNorm | 847722 | glimepiride 4 MG / rosiglitazone 8 MG [Avandaryl] |
| RxNorm | 847724 | glimepiride 4 MG / rosiglitazone 8 MG Oral Tablet [Avandaryl] |
| RxNorm | 861731 | glipizide 2.5 MG / metformin hydrochloride 250 MG Oral Tablet |
| RxNorm | 861732 | glipizide 2.5 MG / metformin hydrochloride 250 MG [Metaglip] |
| RxNorm | 861733 | glipizide 2.5 MG / metformin hydrochloride 250 MG Oral Tablet [Metaglip] |
| RxNorm | 861736 | glipizide 2.5 MG / metformin hydrochloride 500 MG Oral Tablet |
| RxNorm | 861737 | glipizide 2.5 MG / metformin hydrochloride 500 MG [Metaglip] |
| RxNorm | 861738 | glipizide 2.5 MG / metformin hydrochloride 500 MG Oral Tablet [Metaglip] |
| RxNorm | 861740 | glipizide 5 MG / metformin hydrochloride 500 MG Oral Tablet |
| RxNorm | 861741 | glipizide 5 MG / metformin hydrochloride 500 MG [Metaglip] |
| RxNorm | 861742 | glipizide 5 MG / metformin hydrochloride 500 MG Oral Tablet [Metaglip] |
| RxNorm | 861743 | glyburide 1.25 MG / metformin hydrochloride 250 MG Oral Tablet |
| RxNorm | 861745 | glyburide 1.25 MG / metformin hydrochloride 250 MG [Glucovance] |
| RxNorm | 861747 | glyburide 1.25 MG / metformin hydrochloride 250 MG Oral Tablet [Glucovance] |
| RxNorm | 861748 | glyburide 2.5 MG / metformin hydrochloride 500 MG Oral Tablet |

Continued on next page

**Table26 – continued from previous page**

| CodeSystem | ConceptCode | ConceptName |
|---|---|---|
| RxNorm | 861750 | glyburide 2.5 MG / metformin hydrochloride 500 MG [Glucovance] |
| RxNorm | 861752 | glyburide 2.5 MG / metformin hydrochloride 500 MG Oral Tablet [Glucovance] |
| RxNorm | 861753 | glyburide 5 MG / metformin hydrochloride 500 MG Oral Tablet |
| RxNorm | 861755 | glyburide 5 MG / metformin hydrochloride 500 MG [Glucovance] |
| RxNorm | 861756 | glyburide / metformin Oral Tablet [Glucovance] |
| RxNorm | 861757 | glyburide 5 MG / metformin hydrochloride 500 MG Oral Tablet [Glucovance] |
| RxNorm | 881404 | Glynase |
| RxNorm | 881405 | glyburide 1.5 MG [Glynase] |
| RxNorm | 881406 | glyburide Oral Tablet [Glynase] |
| RxNorm | 881407 | glyburide 1.5 MG Oral Tablet [Glynase] |
| RxNorm | 881408 | glyburide 3 MG [Glynase] |
| RxNorm | 881409 | glyburide 3 MG Oral Tablet [Glynase] |
| RxNorm | 881410 | glyburide 6 MG [Glynase] |
| RxNorm | 881411 | glyburide 6 MG Oral Tablet [Glynase] |
| RxNorm | 865567 | glipizide Extended Release Oral Tablet [Glucotrol] |
| RxNorm | 865568 | 24 HR glipizide 10 MG Extended Release Oral Tablet [Glucotrol] |
| RxNorm | 865569 | glipizide 10 MG Extended Release Oral Tablet [Glucotrol] |
| RxNorm | 865570 | glipizide 2.5 MG [Glucotrol] |
| RxNorm | 865571 | 24 HR glipizide 2.5 MG Extended Release Oral Tablet [Glucotrol] |
| RxNorm | 865572 | glipizide 2.5 MG Extended Release Oral Tablet [Glucotrol] |
| RxNorm | 865573 | 24 HR glipizide 5 MG Extended Release Oral Tablet [Glucotrol] |
| RxNorm | 865574 | glipizide 5 MG Extended Release Oral Tablet [Glucotrol] |
| RxNorm | 93312 | glyburide Oral Tablet [Micronase] |

##### 8.5.4 SGLT2 Inhibitors

Table 27: Concept codes used to identify if a patient was prescribed sodium-glucose-cotransporter-2-inhibitor medications.

| CodeSystem | ConceptCode | ConceptName |
|---|---|---|
| RxNorm | 1488564 | dapagliflozin |
| RxNorm | 1545653 | empagliflozin |
| RxNorm | 1546031 | canagliflozin anhydrous |

Continued on next page

**Table27 – continued from previous page**

| CodeSystem | ConceptCode | ConceptName |
|---|---|---|
| RxNorm | 1373458 | canagliflozin |
| RxNorm | 1373459 | canagliflozin 100 MG |
| RxNorm | 1373460 | canagliflozin Oral Product |
| RxNorm | 1373461 | canagliflozin Pill |
| RxNorm | 1373462 | canagliflozin Oral Tablet |
| RxNorm | 1373463 | canagliflozin 100 MG Oral Tablet |
| RxNorm | 1373464 | Invokana |
| RxNorm | 1373465 | canagliflozin 100 MG [Invokana] |
| RxNorm | 1373466 | canagliflozin Oral Tablet [Invokana] |
| RxNorm | 1373467 | Invokana Oral Product |
| RxNorm | 1373468 | Invokana Pill |
| RxNorm | 1373469 | canagliflozin 100 MG Oral Tablet [Invokana] |
| RxNorm | 1373470 | canagliflozin 300 MG |
| RxNorm | 1373471 | canagliflozin 300 MG Oral Tablet |
| RxNorm | 1373472 | canagliflozin 300 MG [Invokana] |
| RxNorm | 1373473 | canagliflozin 300 MG Oral Tablet [Invokana] |
| RxNorm | 1486436 | dapagliflozin / metformin |
| RxNorm | 1486972 | Farxiga |
| RxNorm | 1486975 | Farxiga Oral Product |
| RxNorm | 1486976 | Farxiga Pill |
| RxNorm | 1486977 | dapagliflozin 10 MG Oral Tablet [Farxiga] |
| RxNorm | 1486981 | dapagliflozin 5 MG Oral Tablet [Farxiga] |
| RxNorm | 1488565 | dapagliflozin 10 MG |
| RxNorm | 1488566 | dapagliflozin Oral Product |
| RxNorm | 1488567 | dapagliflozin Pill |
| RxNorm | 1488568 | dapagliflozin Oral Tablet |
| RxNorm | 1488569 | dapagliflozin 10 MG Oral Tablet |
| RxNorm | 1488573 | dapagliflozin 5 MG |
| RxNorm | 1488574 | dapagliflozin 5 MG Oral Tablet |
| RxNorm | 1534343 | dapagliflozin Oral Tablet [Farxiga] |
| RxNorm | 1534344 | dapagliflozin 5 MG [Farxiga] |
| RxNorm | 1534397 | dapagliflozin 10 MG [Farxiga] |
| RxNorm | 1545145 | canagliflozin 150 MG |
| RxNorm | 1545146 | canagliflozin / metformin Oral Product |
| RxNorm | 1545147 | canagliflozin / metformin Pill |
| RxNorm | 1545148 | canagliflozin / metformin Oral Tablet |
| RxNorm | 1545149 | canagliflozin / metformin |
| RxNorm | 1545150 | canagliflozin 150 MG / metformin hydrochloride 1000 MG Oral Tablet |
| RxNorm | 1545151 | Invokamet |
| RxNorm | 1545152 | canagliflozin 150 MG / metformin hydrochloride 1000 MG [Invokamet] |
| RxNorm | 1545153 | canagliflozin / metformin Oral Tablet [Invokamet] |

Continued on next page

**Table27 – continued from previous page**

| CodeSystem | ConceptCode | ConceptName |
|---|---|---|
| RxNorm | 1545154 | Invokamet Oral Product |
| RxNorm | 1545155 | Invokamet Pill |
| RxNorm | 1545156 | canagliflozin 150 MG / metformin hydrochloride 1000 MG Oral Tablet [Invokamet] |
| RxNorm | 1545157 | canagliflozin 150 MG / metformin hydrochloride 500 MG Oral Tablet |
| RxNorm | 1545158 | canagliflozin 150 MG / metformin hydrochloride 500 MG [Invokamet] |
| RxNorm | 1545159 | canagliflozin 150 MG / metformin hydrochloride 500 MG Oral Tablet [Invokamet] |
| RxNorm | 1545160 | canagliflozin 50 MG |
| RxNorm | 1545161 | canagliflozin 50 MG / metformin hydrochloride 1000 MG Oral Tablet |
| RxNorm | 1545162 | canagliflozin 50 MG / metformin hydrochloride 1000 MG [Invokamet] |
| RxNorm | 1545163 | canagliflozin 50 MG / metformin hydrochloride 1000 MG Oral Tablet [Invokamet] |
| RxNorm | 1545164 | canagliflozin 50 MG / metformin hydrochloride 500 MG Oral Tablet |
| RxNorm | 1545165 | canagliflozin 50 MG / metformin hydrochloride 500 MG [Invokamet] |
| RxNorm | 1545166 | canagliflozin 50 MG / metformin hydrochloride 500 MG Oral Tablet [Invokamet] |
| RxNorm | 1545654 | empagliflozin 10 MG |
| RxNorm | 1545655 | empagliflozin Oral Product |
| RxNorm | 1545656 | empagliflozin Pill |
| RxNorm | 1545657 | empagliflozin Oral Tablet |
| RxNorm | 1545658 | empagliflozin 10 MG Oral Tablet |
| RxNorm | 1545659 | Jardiance |
| RxNorm | 1545660 | empagliflozin 10 MG [Jardiance] |
| RxNorm | 1545661 | empagliflozin Oral Tablet [Jardiance] |
| RxNorm | 1545662 | Jardiance Oral Product |
| RxNorm | 1545663 | Jardiance Pill |
| RxNorm | 1545664 | empagliflozin 10 MG Oral Tablet [Jardiance] |
| RxNorm | 1545665 | empagliflozin 25 MG |
| RxNorm | 1545666 | empagliflozin 25 MG Oral Tablet |
| RxNorm | 1545667 | empagliflozin 25 MG [Jardiance] |
| RxNorm | 1545668 | empagliflozin 25 MG Oral Tablet [Jardiance] |
| RxNorm | 1593057 | dapagliflozin / metformin Extended Release Oral Tablet |
| RxNorm | 1593058 | 24 HR dapagliflozin 10 MG / metformin hydrochloride 1000 MG Extended Release Oral Tablet |
| RxNorm | 1593059 | dapagliflozin 10 MG / metformin hydrochloride 1000 MG Extended Release Oral Tablet |

Continued on next page

**Table27 – continued from previous page**

| CodeSystem | ConceptCode | ConceptName |
|---|---|---|
| RxNorm | 1593068 | 24 HR dapagliflozin 10 MG / metformin hydrochloride 500 MG Extended Release Oral Tablet |
| RxNorm | 1593069 | dapagliflozin 10 MG / metformin hydrochloride 500 MG Extended Release Oral Tablet |
| RxNorm | 1593070 | 24 HR dapagliflozin 5 MG / metformin hydrochloride 1000 MG Extended Release Oral Tablet |
| RxNorm | 1593071 | dapagliflozin 5 MG / metformin hydrochloride 1000 MG Extended Release Oral Tablet |
| RxNorm | 1593072 | 24 HR dapagliflozin 5 MG / metformin hydrochloride 500 MG Extended Release Oral Tablet |
| RxNorm | 1593073 | dapagliflozin 5 MG / metformin hydrochloride 500 MG Extended Release Oral Tablet |
| RxNorm | 1593774 | dapagliflozin / metformin Extended Release Oral Tablet [Xigduo] |
| RxNorm | 1593775 | 24 HR dapagliflozin 10 MG / metformin hydrochloride 1000 MG Extended Release Oral Tablet [Xigduo] |
| RxNorm | 1593776 | dapagliflozin 10 MG / metformin hydrochloride 1000 MG Extended Release Oral Tablet [Xigduo] |
| RxNorm | 1593826 | dapagliflozin 10 MG / metformin hydrochloride 500 MG [Xigduo] |
| RxNorm | 1593827 | dapagliflozin 10 MG / metformin hydrochloride 500 MG Extended Release Oral Tablet [Xigduo] |
| RxNorm | 1593828 | dapagliflozin 5 MG / metformin hydrochloride 1000 MG [Xigduo] |
| RxNorm | 1593829 | dapagliflozin 5 MG / metformin hydrochloride 1000 MG Extended Release Oral Tablet [Xigduo] |
| RxNorm | 1593830 | dapagliflozin 5 MG / metformin hydrochloride 500 MG [Xigduo] |
| RxNorm | 1593831 | 24 HR dapagliflozin 5 MG / metformin hydrochloride 500 MG Extended Release Oral Tablet [Xigduo] |
| RxNorm | 1593832 | dapagliflozin 5 MG / metformin hydrochloride 500 MG Extended Release Oral Tablet [Xigduo] |
| RxNorm | 1593833 | 24 HR dapagliflozin 5 MG / metformin hydrochloride 1000 MG Extended Release Oral Tablet [Xigduo] |
| RxNorm | 1593835 | 24 HR dapagliflozin 10 MG / metformin hydrochloride 500 MG Extended Release Oral Tablet [Xigduo] |
| RxNorm | 1598392 | empagliflozin / linagliptin |
| RxNorm | 1602106 | empagliflozin / linagliptin Oral Product |
| RxNorm | 1602107 | empagliflozin / linagliptin Pill |
| RxNorm | 1602108 | empagliflozin / linagliptin Oral Tablet |
| RxNorm | 1602109 | empagliflozin 10 MG / linagliptin 5 MG Oral Tablet |
| RxNorm | 1602110 | Glyxambi |
| RxNorm | 1602111 | empagliflozin 10 MG / linagliptin 5 MG [Glyxambi] |

Continued on next page

**Table27 – continued from previous page**

| CodeSystem | ConceptCode | ConceptName |
|---|---|---|
| RxNorm | 1602112 | empagliflozin / linagliptin Oral Tablet [Glyxambi] |
| RxNorm | 1602113 | Glyxambi Oral Product |
| RxNorm | 1602114 | Glyxambi Pill |
| RxNorm | 1602115 | empagliflozin 10 MG / linagliptin 5 MG Oral Tablet [Glyxambi] |
| RxNorm | 1602118 | empagliflozin 25 MG / linagliptin 5 MG Oral Tablet |
| RxNorm | 1602119 | empagliflozin 25 MG / linagliptin 5 MG [Glyxambi] |
| RxNorm | 1602120 | empagliflozin 25 MG / linagliptin 5 MG Oral Tablet [Glyxambi] |
| RxNorm | 1592709 | dapagliflozin / metformin Oral Product |
| RxNorm | 1592710 | dapagliflozin / metformin Pill |
| RxNorm | 1592713 | Xigduo |
| RxNorm | 1592716 | Xigduo Oral Product |
| RxNorm | 1592717 | Xigduo Pill |
| RxNorm | 1592722 | dapagliflozin 10 MG / metformin hydrochloride 1000 MG [Xigduo] |
| RxNorm | 1664310 | empagliflozin 5 MG |
| RxNorm | 1664311 | empagliflozin / metformin Oral Product |
| RxNorm | 1664312 | empagliflozin / metformin Pill |
| RxNorm | 1664313 | empagliflozin / metformin Oral Tablet |
| RxNorm | 1664314 | empagliflozin / metformin |
| RxNorm | 1664315 | empagliflozin 5 MG / metformin hydrochloride 500 MG Oral Tablet |
| RxNorm | 1664316 | Synjardy |
| RxNorm | 1664317 | empagliflozin 5 MG / metformin hydrochloride 500 MG [Synjardy] |
| RxNorm | 1664318 | empagliflozin / metformin Oral Tablet [Synjardy] |
| RxNorm | 1664319 | Synjardy Oral Product |
| RxNorm | 1664320 | Synjardy Pill |
| RxNorm | 1664321 | empagliflozin 5 MG / metformin hydrochloride 500 MG Oral Tablet [Synjardy] |
| RxNorm | 1664322 | empagliflozin 12.5 MG |
| RxNorm | 1664323 | empagliflozin 12.5 MG / metformin hydrochloride 500 MG Oral Tablet |
| RxNorm | 1664324 | empagliflozin 12.5 MG / metformin hydrochloride 500 MG [Synjardy] |
| RxNorm | 1664325 | empagliflozin 12.5 MG / metformin hydrochloride 500 MG Oral Tablet [Synjardy] |
| RxNorm | 1664326 | empagliflozin 5 MG / metformin hydrochloride 1000 MG Oral Tablet |
| RxNorm | 1664327 | empagliflozin 5 MG / metformin hydrochloride 1000 MG [Synjardy] |

Continued on next page

**Table27 – continued from previous page**

| CodeSystem | ConceptCode | ConceptName |
|---|---|---|
| RxNorm | 1664328 | empagliflozin 5 MG / metformin hydrochloride 1000 MG Oral Tablet [Synjardy] |
| RxNorm | 1665367 | empagliflozin 12.5 MG / metformin hydrochloride 1000 MG Oral Tablet |
| RxNorm | 1665368 | empagliflozin 12.5 MG / metformin hydrochloride 1000 MG [Synjardy] |
| RxNorm | 1665369 | empagliflozin 12.5 MG / metformin hydrochloride 1000 MG Oral Tablet [Synjardy] |
| RxNorm | 1727500 | dapagliflozin / saxagliptin |
| RxNorm | 1810996 | canagliflozin / metformin Extended Release Oral Tablet |
| RxNorm | 1810997 | 24 HR canagliflozin 150 MG / metformin hydrochloride 1000 MG Extended Release Oral Tablet |
| RxNorm | 1810998 | canagliflozin / metformin Extended Release Oral Tablet [Invokamet] |
| RxNorm | 1810999 | 24 HR canagliflozin 150 MG / metformin hydrochloride 1000 MG Extended Release Oral Tablet [Invokamet] |
| RxNorm | 1811000 | canagliflozin 150 MG / metformin hydrochloride 1000 MG Extended Release Oral Tablet |
| RxNorm | 1811001 | canagliflozin 150 MG / metformin hydrochloride 1000 MG Extended Release Oral Tablet [Invokamet] |
| RxNorm | 1811002 | 24 HR canagliflozin 150 MG / metformin hydrochloride 500 MG Extended Release Oral Tablet |
| RxNorm | 1811003 | 24 HR canagliflozin 150 MG / metformin hydrochloride 500 MG Extended Release Oral Tablet [Invokamet] |
| RxNorm | 1811004 | canagliflozin 150 MG / metformin hydrochloride 500 MG Extended Release Oral Tablet |
| RxNorm | 1811005 | canagliflozin 150 MG / metformin hydrochloride 500 MG Extended Release Oral Tablet [Invokamet] |
| RxNorm | 1811006 | 24 HR canagliflozin 50 MG / metformin hydrochloride 1000 MG Extended Release Oral Tablet |
| RxNorm | 1811007 | 24 HR canagliflozin 50 MG / metformin hydrochloride 1000 MG Extended Release Oral Tablet [Invokamet] |
| RxNorm | 1862684 | empagliflozin / metformin Extended Release Oral Tablet |
| RxNorm | 1862685 | 24 HR empagliflozin 10 MG / metformin hydrochloride 1000 MG Extended Release Oral Tablet |
| RxNorm | 1862686 | empagliflozin 10 MG / metformin hydrochloride 1000 MG [Synjardy] |
| RxNorm | 1862687 | empagliflozin / metformin Extended Release Oral Tablet [Synjardy] |
| RxNorm | 1862688 | 24 HR empagliflozin 10 MG / metformin hydrochloride 1000 MG Extended Release Oral Tablet [Synjardy] |
| RxNorm | 1862689 | empagliflozin 10 MG / metformin hydrochloride 1000 MG Extended Release Oral Tablet |

Continued on next page

**Table27 – continued from previous page**

| CodeSystem | ConceptCode | ConceptName |
| --- | --- | --- |
| RxNorm | 1862690 | empagliflozin 10 MG / metformin hydrochloride 1000 MG Extended Release Oral Tablet [Synjardy] |
| RxNorm | 1862691 | 24 HR empagliflozin 12.5 MG / metformin hydrochloride 1000 MG Extended Release Oral Tablet |
| RxNorm | 1862692 | 24 HR empagliflozin 12.5 MG / metformin hydrochloride 1000 MG Extended Release Oral Tablet [Synjardy] |
| RxNorm | 1862693 | empagliflozin 12.5 MG / metformin hydrochloride 1000 MG Extended Release Oral Tablet |
| RxNorm | 1811008 | canagliflozin 50 MG / metformin hydrochloride 1000 MG Extended Release Oral Tablet |
| RxNorm | 1811009 | canagliflozin 50 MG / metformin hydrochloride 1000 MG Extended Release Oral Tablet [Invokamet] |
| RxNorm | 1811010 | 24 HR canagliflozin 50 MG / metformin hydrochloride 500 MG Extended Release Oral Tablet |
| RxNorm | 1811011 | 24 HR canagliflozin 50 MG / metformin hydrochloride 500 MG Extended Release Oral Tablet [Invokamet] |
| RxNorm | 1811012 | canagliflozin 50 MG / metformin hydrochloride 500 MG Extended Release Oral Tablet |
| RxNorm | 1811013 | canagliflozin 50 MG / metformin hydrochloride 500 MG Extended Release Oral Tablet [Invokamet] |
| RxNorm | 1862694 | empagliflozin 12.5 MG / metformin hydrochloride 1000 MG Extended Release Oral Tablet [Synjardy] |
| RxNorm | 1862695 | 24 HR empagliflozin 25 MG / metformin hydrochloride 1000 MG Extended Release Oral Tablet |
| RxNorm | 1862696 | empagliflozin 25 MG / metformin hydrochloride 1000 MG [Synjardy] |
| RxNorm | 1862697 | 24 HR empagliflozin 25 MG / metformin hydrochloride 1000 MG Extended Release Oral Tablet [Synjardy] |
| RxNorm | 1862698 | empagliflozin 25 MG / metformin hydrochloride 1000 MG Extended Release Oral Tablet |
| RxNorm | 1862699 | empagliflozin 25 MG / metformin hydrochloride 1000 MG Extended Release Oral Tablet [Synjardy] |
| RxNorm | 1862700 | 24 HR empagliflozin 5 MG / metformin hydrochloride 1000 MG Extended Release Oral Tablet |
| RxNorm | 1862701 | 24 HR empagliflozin 5 MG / metformin hydrochloride 1000 MG Extended Release Oral Tablet [Synjardy] |
| RxNorm | 1862702 | empagliflozin 5 MG / metformin hydrochloride 1000 MG Extended Release Oral Tablet |
| RxNorm | 1862703 | empagliflozin 5 MG / metformin hydrochloride 1000 MG Extended Release Oral Tablet [Synjardy] |
| RxNorm | 1925495 | dapagliflozin / saxagliptin Oral Product |
| RxNorm | 1925496 | dapagliflozin / saxagliptin Pill |
| RxNorm | 1925497 | dapagliflozin / saxagliptin Oral Tablet |

Continued on next page

**Table27 – continued from previous page**

| CodeSystem | ConceptCode | ConceptName |
|---|---|---|
| RxNorm | 1925498 | dapagliflozin 10 MG / saxagliptin 5 MG Oral Tablet |
| RxNorm | 1925499 | Qtern |
| RxNorm | 1925500 | dapagliflozin 10 MG / saxagliptin 5 MG [Qtern] |
| RxNorm | 1925501 | dapagliflozin / saxagliptin Oral Tablet [Qtern] |
| RxNorm | 1925502 | Qtern Oral Product |
| RxNorm | 1925503 | Qtern Pill |
| RxNorm | 1925504 | dapagliflozin 10 MG / saxagliptin 5 MG Oral Tablet [Qtern] |
| RxNorm | 1940495 | dapagliflozin 2.5 MG |
| RxNorm | 1940496 | 24 HR dapagliflozin 2.5 MG / metformin hydrochloride 1000 MG Extended Release Oral Tablet |
| RxNorm | 1940497 | dapagliflozin 2.5 MG / metformin hydrochloride 1000 MG [Xigduo] |
| RxNorm | 1940498 | 24 HR dapagliflozin 2.5 MG / metformin hydrochloride 1000 MG Extended Release Oral Tablet [Xigduo] |
| RxNorm | 1940499 | dapagliflozin 2.5 MG / metformin hydrochloride 1000 MG Extended Release Oral Tablet |
| RxNorm | 1940500 | dapagliflozin 2.5 MG / metformin hydrochloride 1000 MG Extended Release Oral Tablet [Xigduo] |
| RxNorm | 2117292 | dapagliflozin / metformin / saxagliptin |
| RxNorm | 2169274 | dapagliflozin 5 MG / saxagliptin 5 MG Oral Tablet |
| RxNorm | 2169275 | dapagliflozin 5 MG / saxagliptin 5 MG [Qtern] |
| RxNorm | 2169276 | dapagliflozin 5 MG / saxagliptin 5 MG Oral Tablet [Qtern] |
| RxNorm | 2281864 | empagliflozin / linagliptin / metformin |
| RxNorm | 2371722 | dapagliflozin / metformin / saxagliptin Oral Product |
| RxNorm | 2371723 | dapagliflozin / metformin / saxagliptin Pill |
| RxNorm | 2371724 | dapagliflozin / metformin / saxagliptin Extended Release Oral Tablet |
| RxNorm | 2371725 | 24 HR dapagliflozin 10 MG / metformin hydrochloride 1000 MG / saxagliptin 5 MG Extended Release Oral Tablet |
| RxNorm | 2371726 | Qternmet |
| RxNorm | 2371727 | dapagliflozin 10 MG / metformin hydrochloride 1000 MG / saxagliptin 5 MG [Qternmet] |
| RxNorm | 2371728 | dapagliflozin / metformin / saxagliptin Extended Release Oral Tablet [Qternmet] |
| RxNorm | 2371729 | Qternmet Oral Product |
| RxNorm | 2371730 | Qternmet Pill |
| RxNorm | 2371731 | 24 HR dapagliflozin 10 MG / metformin hydrochloride 1000 MG / saxagliptin 5 MG Extended Release Oral Tablet [Qternmet] |
| RxNorm | 2371732 | dapagliflozin 10 MG / metformin hydrochloride 1000 MG / saxagliptin 5 MG Extended Release Oral Tablet |
| RxNorm | 2371733 | dapagliflozin 10 MG / metformin hydrochloride 1000 MG / saxagliptin 5 MG Extended Release Oral Tablet [Qternmet] |

Continued on next page

**Table27 – continued from previous page**

| CodeSystem | ConceptCode | ConceptName |
|---|---|---|
| RxNorm | 2371734 | 24 HR dapagliflozin 2.5 MG / metformin hydrochloride 1000 MG / saxagliptin 2.5 MG Extended Release Oral Tablet |
| RxNorm | 2371735 | dapagliflozin 2.5 MG / metformin hydrochloride 1000 MG / saxagliptin 2.5 MG [Qternmet] |
| RxNorm | 2371736 | 24 HR dapagliflozin 2.5 MG / metformin hydrochloride 1000 MG / saxagliptin 2.5 MG Extended Release Oral Tablet [Qternmet] |
| RxNorm | 2371737 | dapagliflozin 2.5 MG / metformin hydrochloride 1000 MG / saxagliptin 2.5 MG Extended Release Oral Tablet |
| RxNorm | 2371738 | dapagliflozin 2.5 MG / metformin hydrochloride 1000 MG / saxagliptin 2.5 MG Extended Release Oral Tablet [Qternmet] |
| RxNorm | 2371740 | 24 HR dapagliflozin 5 MG / metformin hydrochloride 1000 MG / saxagliptin 2.5 MG Extended Release Oral Tablet |
| RxNorm | 2371741 | dapagliflozin 5 MG / metformin hydrochloride 1000 MG / saxagliptin 2.5 MG [Qternmet] |
| RxNorm | 2371742 | 24 HR dapagliflozin 5 MG / metformin hydrochloride 1000 MG / saxagliptin 2.5 MG Extended Release Oral Tablet [Qternmet] |
| RxNorm | 2371743 | dapagliflozin 5 MG / metformin hydrochloride 1000 MG / saxagliptin 2.5 MG Extended Release Oral Tablet |
| RxNorm | 2371744 | dapagliflozin 5 MG / metformin hydrochloride 1000 MG / saxagliptin 2.5 MG Extended Release Oral Tablet [Qternmet] |
| RxNorm | 2371745 | 24 HR dapagliflozin 5 MG / metformin hydrochloride 1000 MG / saxagliptin 5 MG Extended Release Oral Tablet |
| RxNorm | 2371746 | dapagliflozin 5 MG / metformin hydrochloride 1000 MG / saxagliptin 5 MG [Qternmet] |
| RxNorm | 2371747 | 24 HR dapagliflozin 5 MG / metformin hydrochloride 1000 MG / saxagliptin 5 MG Extended Release Oral Tablet [Qternmet] |
| RxNorm | 2371748 | dapagliflozin 5 MG / metformin hydrochloride 1000 MG / saxagliptin 5 MG Extended Release Oral Tablet |
| RxNorm | 2371749 | dapagliflozin 5 MG / metformin hydrochloride 1000 MG / saxagliptin 5 MG Extended Release Oral Tablet [Qternmet] |
| RxNorm | 2359276 | empagliflozin / linagliptin / metformin Oral Product |
| RxNorm | 2359277 | empagliflozin / linagliptin / metformin Pill |
| RxNorm | 2359278 | empagliflozin / linagliptin / metformin Extended Release Oral Tablet |
| RxNorm | 2359279 | 24 HR empagliflozin 10 MG / linagliptin 5 MG / metformin hydrochloride 1000 MG Extended Release Oral Tablet |
| RxNorm | 2359280 | Trijardy |

Continued on next page

**Table27 – continued from previous page**

| CodeSystem | ConceptCode | ConceptName |
|---|---|---|
| RxNorm | 2359281 | empagliflozin 10 MG / linagliptin 5 MG / metformin hydrochloride 1000 MG [Trijardy] |
| RxNorm | 2359282 | empagliflozin / linagliptin / metformin Extended Release Oral Tablet [Trijardy] |
| RxNorm | 2359283 | Trijardy Oral Product |
| RxNorm | 2359284 | Trijardy Pill |
| RxNorm | 2359285 | 24 HR empagliflozin 10 MG / linagliptin 5 MG / metformin hydrochloride 1000 MG Extended Release Oral Tablet [Trijardy] |
| RxNorm | 2359286 | empagliflozin 10 MG / linagliptin 5 MG / metformin hydrochloride 1000 MG Extended Release Oral Tablet |
| RxNorm | 2359287 | empagliflozin 10 MG / linagliptin 5 MG / metformin hydrochloride 1000 MG Extended Release Oral Tablet [Trijardy] |
| RxNorm | 2359288 | 24 HR empagliflozin 12.5 MG / linagliptin 2.5 MG / metformin hydrochloride 1000 MG Extended Release Oral Tablet |
| RxNorm | 2359289 | empagliflozin 12.5 MG / linagliptin 2.5 MG / metformin hydrochloride 1000 MG [Trijardy] |
| RxNorm | 2359290 | 24 HR empagliflozin 12.5 MG / linagliptin 2.5 MG / metformin hydrochloride 1000 MG Extended Release Oral Tablet [Trijardy] |
| RxNorm | 2359291 | empagliflozin 12.5 MG / linagliptin 2.5 MG / metformin hydrochloride 1000 MG Extended Release Oral Tablet |
| RxNorm | 2359292 | empagliflozin 12.5 MG / linagliptin 2.5 MG / metformin hydrochloride 1000 MG Extended Release Oral Tablet [Trijardy] |
| RxNorm | 2359351 | 24 HR empagliflozin 25 MG / linagliptin 5 MG / metformin hydrochloride 1000 MG Extended Release Oral Tablet |
| RxNorm | 2359352 | empagliflozin 25 MG / linagliptin 5 MG / metformin hydrochloride 1000 MG [Trijardy] |
| RxNorm | 2359353 | 24 HR empagliflozin 25 MG / linagliptin 5 MG / metformin hydrochloride 1000 MG Extended Release Oral Tablet [Trijardy] |
| RxNorm | 2359354 | empagliflozin 25 MG / linagliptin 5 MG / metformin hydrochloride 1000 MG Extended Release Oral Tablet |
| RxNorm | 2359355 | empagliflozin 25 MG / linagliptin 5 MG / metformin hydrochloride 1000 MG Extended Release Oral Tablet [Trijardy] |
| RxNorm | 2359356 | 24 HR empagliflozin 5 MG / linagliptin 2.5 MG / metformin hydrochloride 1000 MG Extended Release Oral Tablet |
| RxNorm | 2359357 | empagliflozin 5 MG / linagliptin 2.5 MG / metformin hydrochloride 1000 MG [Trijardy] |

Continued on next page

**Table27 – continued from previous page**

| CodeSystem | ConceptCode | ConceptName |
| --- | --- | --- |
| RxNorm | 2359358 | 24 HR empagliflozin 5 MG / linagliptin 2.5 MG / metformin hydrochloride 1000 MG Extended Release Oral Tablet [Trijardy] |
| RxNorm | 2359359 | empagliflozin 5 MG / linagliptin 2.5 MG / metformin hydrochloride 1000 MG Extended Release Oral Tablet |
| RxNorm | 2359360 | empagliflozin 5 MG / linagliptin 2.5 MG / metformin hydrochloride 1000 MG Extended Release Oral Tablet [Trijardy] |

##### 8.5.5 DPP-4

Table 28: Concept codes used to identify if a patient was prescribed dipeptidyl-peptidase-4-inhibitor medications.

| CodeSystem | ConceptCode | ConceptName |
| --- | --- | --- |
| RxNorm | 1100699 | linagliptin |
| RxNorm | 1368001 | alogliptin |
| RxNorm | 1546030 | saxagliptin anhydrous |
| RxNorm | 593411 | sitagliptin |
| RxNorm | 700518 | Metformin 1000 MG / sitagliptin 50 MG Oral Tablet |
| RxNorm | 700517 | Metformin 500 MG / sitagliptin 50 MG Oral Tablet |
| RxNorm | 757606 | Metformin 500 MG / sitagliptin 50 MG [Janumet 50 mg/500 mg] |
| RxNorm | 757602 | Metformin 1000 MG / sitagliptin 50 MG [Janumet 50 mg/1000 mg] |
| RxNorm | 757608 | Metformin 500 MG / sitagliptin 50 MG Oral Tablet [Janumet 50 mg/500 mg] |
| RxNorm | 757604 | Metformin 1000 MG / sitagliptin 50 MG Oral Tablet [Janumet 50 mg/1000 mg] |
| RxNorm | 1243830 | Metformin / sitagliptin Extended Release Tablet [Janumet 100/1000] |
| RxNorm | 1243847 | Metformin / sitagliptin Extended Release Tablet [Janumet 50/500] |
| RxNorm | 1243023 | Linagliptin / Metformin Oral Tablet [Jentadueto 2.5/1000] |
| RxNorm | 1243030 | Linagliptin / Metformin Oral Tablet [Jentadueto 2.5/500] |
| RxNorm | 1043561 | metformin / saxagliptin Extended Release Oral Tablet |
| RxNorm | 1043562 | metformin / saxagliptin |
| RxNorm | 1043563 | 24 HR metformin hydrochloride 1000 MG / saxagliptin 2.5 MG Extended Release Oral Tablet |
| RxNorm | 1043565 | metformin hydrochloride 1000 MG / saxagliptin 2.5 MG [Kombiglyze] |

Continued on next page

**Table28 – continued from previous page**

| CodeSystem | ConceptCode | ConceptName |
|---|---|---|
| RxNorm | 1043566 | metformin / saxagliptin Extended Release Oral Tablet [Kombiglyze] |
| RxNorm | 1043567 | 24 HR metformin hydrochloride 1000 MG / saxagliptin 2.5 MG Extended Release Oral Tablet [Kombiglyze] |
| RxNorm | 1043568 | metformin hydrochloride 1000 MG / saxagliptin 2.5 MG Extended Release Oral Tablet |
| RxNorm | 1043569 | metformin hydrochloride 1000 MG / saxagliptin 2.5 MG Extended Release Oral Tablet [Kombiglyze] |
| RxNorm | 1043570 | 24 HR metformin hydrochloride 1000 MG / saxagliptin 5 MG Extended Release Oral Tablet |
| RxNorm | 1043572 | metformin hydrochloride 1000 MG / saxagliptin 5 MG [Kombiglyze] |
| RxNorm | 1043574 | 24 HR metformin hydrochloride 1000 MG / saxagliptin 5 MG Extended Release Oral Tablet [Kombiglyze] |
| RxNorm | 1043575 | metformin hydrochloride 1000 MG / saxagliptin 5 MG Extended Release Oral Tablet |
| RxNorm | 1043576 | metformin hydrochloride 1000 MG / saxagliptin 5 MG Extended Release Oral Tablet [Kombiglyze] |
| RxNorm | 1043578 | 24 HR metformin hydrochloride 500 MG / saxagliptin 5 MG Extended Release Oral Tablet |
| RxNorm | 1043580 | metformin hydrochloride 500 MG / saxagliptin 5 MG [Kombiglyze] |
| RxNorm | 1043582 | 24 HR metformin hydrochloride 500 MG / saxagliptin 5 MG Extended Release Oral Tablet [Kombiglyze] |
| RxNorm | 1043583 | metformin hydrochloride 500 MG / saxagliptin 5 MG Extended Release Oral Tablet |
| RxNorm | 1043584 | metformin hydrochloride 500 MG / saxagliptin 5 MG Extended Release Oral Tablet [Kombiglyze] |
| RxNorm | 1100700 | linagliptin 5 MG |
| RxNorm | 1100701 | linagliptin Oral Tablet |
| RxNorm | 1100702 | linagliptin 5 MG Oral Tablet |
| RxNorm | 1100703 | Tradjenta |
| RxNorm | 1100704 | linagliptin 5 MG [Tradjenta] |
| RxNorm | 1100705 | linagliptin Oral Tablet [Tradjenta] |
| RxNorm | 1100706 | linagliptin 5 MG Oral Tablet [Tradjenta] |
| RxNorm | 1158518 | saxagliptin Oral Product |
| RxNorm | 1158519 | saxagliptin Pill |
| RxNorm | 1159662 | sitagliptin Oral Product |
| RxNorm | 1159663 | sitagliptin Pill |
| RxNorm | 1161605 | metformin / saxagliptin Oral Product |
| RxNorm | 1161606 | metformin / saxagliptin Pill |
| RxNorm | 1161607 | metformin / sitagliptin Oral Product |
| RxNorm | 1161608 | metformin / sitagliptin Pill |

Continued on next page

**Table28 – continued from previous page**

| CodeSystem | ConceptCode | ConceptName |
|---|---|---|
| RxNorm | 1164670 | linagliptin Oral Product |
| RxNorm | 1164671 | linagliptin Pill |
| RxNorm | 1181729 | Onglyza Oral Product |
| RxNorm | 1181730 | Onglyza Pill |
| RxNorm | 1172860 | Kombiglyze Pill |
| RxNorm | 1172861 | Kombiglyze Oral Product |
| RxNorm | 1167810 | Janumet Oral Product |
| RxNorm | 1167811 | Janumet Pill |
| RxNorm | 1167814 | Januvia Oral Product |
| RxNorm | 1167815 | Januvia Pill |
| RxNorm | 1179163 | Tradjenta Oral Product |
| RxNorm | 1179164 | Tradjenta Pill |
| RxNorm | 1189800 | simvastatin / sitagliptin Oral Product |
| RxNorm | 1189801 | simvastatin / sitagliptin Pill |
| RxNorm | 1189802 | simvastatin / sitagliptin Oral Tablet |
| RxNorm | 1189803 | simvastatin / sitagliptin |
| RxNorm | 1189804 | simvastatin 10 MG / sitagliptin 100 MG Oral Tablet |
| RxNorm | 1189806 | simvastatin 10 MG / sitagliptin 100 MG [Juvisync] |
| RxNorm | 1189808 | simvastatin 20 MG / sitagliptin 100 MG Oral Tablet |
| RxNorm | 1189810 | simvastatin 20 MG / sitagliptin 100 MG [Juvisync] |
| RxNorm | 1189811 | simvastatin / sitagliptin Oral Tablet [Juvisync] |
| RxNorm | 1189812 | Juvisync Oral Product |
| RxNorm | 1189813 | Juvisync Pill |
| RxNorm | 1189814 | simvastatin 20 MG / sitagliptin 100 MG Oral Tablet [Juvisync] |
| RxNorm | 1189818 | simvastatin 10 MG / sitagliptin 100 MG Oral Tablet [Juvisync] |
| RxNorm | 1189821 | simvastatin 40 MG / sitagliptin 100 MG Oral Tablet |
| RxNorm | 1189823 | simvastatin 40 MG / sitagliptin 100 MG [Juvisync] |
| RxNorm | 1189827 | simvastatin 40 MG / sitagliptin 100 MG Oral Tablet [Juvisync] |
| RxNorm | 1243015 | linagliptin 2.5 MG |
| RxNorm | 1243016 | linagliptin / metformin Oral Product |
| RxNorm | 1243017 | linagliptin / metformin Pill |
| RxNorm | 1243018 | linagliptin / metformin Oral Tablet |
| RxNorm | 1243019 | linagliptin / metformin |
| RxNorm | 1243020 | linagliptin 2.5 MG / metformin hydrochloride 1000 MG Oral Tablet |
| RxNorm | 1243022 | linagliptin 2.5 MG / metformin hydrochloride 1000 MG [Jentadueto] |
| RxNorm | 1243026 | linagliptin 2.5 MG / metformin hydrochloride 1000 MG Oral Tablet [Jentadueto] |

Continued on next page

**Table28 – continued from previous page**

| CodeSystem | ConceptCode | ConceptName |
|---|---|---|
| RxNorm | 1243027 | linagliptin 2.5 MG / metformin hydrochloride 500 MG Oral Tablet |
| RxNorm | 1243029 | linagliptin 2.5 MG / metformin hydrochloride 500 MG [Jentadueto] |
| RxNorm | 1243033 | linagliptin 2.5 MG / metformin hydrochloride 500 MG Oral Tablet [Jentadueto] |
| RxNorm | 1243034 | linagliptin 2.5 MG / metformin hydrochloride 850 MG Oral Tablet |
| RxNorm | 1243036 | linagliptin 2.5 MG / metformin hydrochloride 850 MG [Jentadueto] |
| RxNorm | 1243037 | linagliptin / metformin Oral Tablet [Jentadueto] |
| RxNorm | 1243038 | Jentadueto Oral Product |
| RxNorm | 1243039 | Jentadueto Pill |
| RxNorm | 1243040 | linagliptin 2.5 MG / metformin hydrochloride 850 MG Oral Tablet [Jentadueto] |
| RxNorm | 1243826 | metformin / sitagliptin Extended Release Oral Tablet |
| RxNorm | 1243827 | 24 HR metformin hydrochloride 1000 MG / sitagliptin 100 MG Extended Release Oral Tablet |
| RxNorm | 1243829 | metformin hydrochloride 1000 MG / sitagliptin 100 MG [Janumet] |
| RxNorm | 1243833 | 24 HR metformin hydrochloride 1000 MG / sitagliptin 100 MG Extended Release Oral Tablet [Janumet] |
| RxNorm | 1243834 | metformin hydrochloride 1000 MG / sitagliptin 100 MG Extended Release Oral Tablet |
| RxNorm | 1243835 | metformin hydrochloride 1000 MG / sitagliptin 100 MG Extended Release Oral Tablet [Janumet] |
| RxNorm | 1243839 | metformin / sitagliptin Extended Release Oral Tablet [Janumet] |
| RxNorm | 1243842 | 24 HR metformin hydrochloride 1000 MG / sitagliptin 50 MG Extended Release Oral Tablet |
| RxNorm | 1243843 | 24 HR metformin hydrochloride 1000 MG / sitagliptin 50 MG Extended Release Oral Tablet [Janumet] |
| RxNorm | 1243844 | metformin hydrochloride 1000 MG / sitagliptin 50 MG Extended Release Oral Tablet |
| RxNorm | 1243845 | metformin hydrochloride 1000 MG / sitagliptin 50 MG Extended Release Oral Tablet [Janumet] |
| RxNorm | 1243846 | 24 HR metformin hydrochloride 500 MG / sitagliptin 50 MG Extended Release Oral Tablet |
| RxNorm | 1243848 | 24 HR metformin hydrochloride 500 MG / sitagliptin 50 MG Extended Release Oral Tablet [Janumet] |
| RxNorm | 1243849 | metformin hydrochloride 500 MG / sitagliptin 50 MG Extended Release Oral Tablet |

Continued on next page

**Table28 – continued from previous page**

| CodeSystem | ConceptCode | ConceptName |
|---|---|---|
| RxNorm | 1243850 | metformin hydrochloride 500 MG / sitagliptin 50 MG Extended Release Oral Tablet [Janumet] |
| RxNorm | 1312409 | simvastatin 10 MG / sitagliptin 50 MG Oral Tablet |
| RxNorm | 1312411 | simvastatin 10 MG / sitagliptin 50 MG [Juvisync] |
| RxNorm | 1312415 | simvastatin 10 MG / sitagliptin 50 MG Oral Tablet [Juvisync] |
| RxNorm | 1312416 | simvastatin 20 MG / sitagliptin 50 MG Oral Tablet |
| RxNorm | 1312418 | simvastatin 20 MG / sitagliptin 50 MG [Juvisync] |
| RxNorm | 1312422 | simvastatin 20 MG / sitagliptin 50 MG Oral Tablet [Juvisync] |
| RxNorm | 1312423 | simvastatin 40 MG / sitagliptin 50 MG Oral Tablet |
| RxNorm | 1312425 | simvastatin 40 MG / sitagliptin 50 MG [Juvisync] |
| RxNorm | 1312429 | simvastatin 40 MG / sitagliptin 50 MG Oral Tablet [Juvisync] |
| RxNorm | 1368002 | alogliptin 25 MG |
| RxNorm | 1368003 | alogliptin Oral Product |
| RxNorm | 1368004 | alogliptin Pill |
| RxNorm | 1368005 | alogliptin Oral Tablet |
| RxNorm | 1368006 | alogliptin 25 MG Oral Tablet |
| RxNorm | 1368007 | Nesina |
| RxNorm | 1368008 | alogliptin 25 MG [Nesina] |
| RxNorm | 1368009 | alogliptin Oral Tablet [Nesina] |
| RxNorm | 1368010 | Nesina Oral Product |
| RxNorm | 1368011 | Nesina Pill |
| RxNorm | 1368012 | alogliptin 25 MG Oral Tablet [Nesina] |
| RxNorm | 1368017 | alogliptin 6.25 MG |
| RxNorm | 1368018 | alogliptin 6.25 MG Oral Tablet |
| RxNorm | 1368019 | alogliptin 6.25 MG [Nesina] |
| RxNorm | 1368020 | alogliptin 6.25 MG Oral Tablet [Nesina] |
| RxNorm | 1368033 | alogliptin 12.5 MG |
| RxNorm | 1368034 | alogliptin 12.5 MG Oral Tablet |
| RxNorm | 1368035 | alogliptin 12.5 MG [Nesina] |
| RxNorm | 1368036 | alogliptin 12.5 MG Oral Tablet [Nesina] |
| RxNorm | 1368381 | alogliptin / metformin Oral Product |
| RxNorm | 1368382 | alogliptin / metformin Pill |
| RxNorm | 1368383 | alogliptin / metformin Oral Tablet |
| RxNorm | 1368384 | alogliptin / metformin |
| RxNorm | 1368385 | alogliptin 12.5 MG / metformin hydrochloride 1000 MG Oral Tablet |
| RxNorm | 1368387 | alogliptin 12.5 MG / metformin hydrochloride 1000 MG [Kazano] |
| RxNorm | 1368391 | alogliptin 12.5 MG / metformin hydrochloride 1000 MG Oral Tablet [Kazano] |

Continued on next page

**Table28 – continued from previous page**

| CodeSystem | ConceptCode | ConceptName |
|---|---|---|
| RxNorm | 1368392 | alogliptin 12.5 MG / metformin hydrochloride 500 MG Oral Tablet |
| RxNorm | 1368394 | alogliptin 12.5 MG / metformin hydrochloride 500 MG [Kazano] |
| RxNorm | 1368395 | alogliptin / metformin Oral Tablet [Kazano] |
| RxNorm | 1368396 | Kazano Oral Product |
| RxNorm | 1368397 | Kazano Pill |
| RxNorm | 1368398 | alogliptin 12.5 MG / metformin hydrochloride 500 MG Oral Tablet [Kazano] |
| RxNorm | 1368399 | alogliptin / pioglitazone Oral Product |
| RxNorm | 1368400 | alogliptin / pioglitazone Pill |
| RxNorm | 1368401 | alogliptin / pioglitazone Oral Tablet |
| RxNorm | 1368402 | alogliptin / pioglitazone |
| RxNorm | 1368403 | alogliptin 12.5 MG / pioglitazone 15 MG Oral Tablet |
| RxNorm | 1368405 | alogliptin 12.5 MG / pioglitazone 15 MG [Oseni] |
| RxNorm | 1368409 | alogliptin 12.5 MG / pioglitazone 15 MG Oral Tablet [Oseni] |
| RxNorm | 1368410 | alogliptin 12.5 MG / pioglitazone 30 MG Oral Tablet |
| RxNorm | 1368412 | alogliptin 12.5 MG / pioglitazone 30 MG [Oseni] |
| RxNorm | 1368416 | alogliptin 12.5 MG / pioglitazone 30 MG Oral Tablet [Oseni] |
| RxNorm | 1368417 | alogliptin 12.5 MG / pioglitazone 45 MG Oral Tablet |
| RxNorm | 1368419 | alogliptin 12.5 MG / pioglitazone 45 MG [Oseni] |
| RxNorm | 1368423 | alogliptin 12.5 MG / pioglitazone 45 MG Oral Tablet [Oseni] |
| RxNorm | 1368424 | alogliptin 25 MG / pioglitazone 15 MG Oral Tablet |
| RxNorm | 1368426 | alogliptin 25 MG / pioglitazone 15 MG [Oseni] |
| RxNorm | 1368430 | alogliptin 25 MG / pioglitazone 15 MG Oral Tablet [Oseni] |
| RxNorm | 1368431 | alogliptin 25 MG / pioglitazone 30 MG Oral Tablet |
| RxNorm | 1368433 | alogliptin 25 MG / pioglitazone 30 MG [Oseni] |
| RxNorm | 1368434 | alogliptin / pioglitazone Oral Tablet [Oseni] |
| RxNorm | 1368435 | Oseni Oral Product |
| RxNorm | 1368436 | Oseni Pill |
| RxNorm | 1368437 | alogliptin 25 MG / pioglitazone 30 MG Oral Tablet [Oseni] |
| RxNorm | 1368438 | alogliptin 25 MG / pioglitazone 45 MG Oral Tablet |
| RxNorm | 1368440 | alogliptin 25 MG / pioglitazone 45 MG [Oseni] |
| RxNorm | 1368444 | alogliptin 25 MG / pioglitazone 45 MG Oral Tablet [Oseni] |
| RxNorm | 1372692 | Kazano |
| RxNorm | 1372706 | Jentadueto |
| RxNorm | 1372717 | Oseni |
| RxNorm | 1372730 | Kombiglyze |
| RxNorm | 1372738 | Janumet |
| RxNorm | 1372754 | Juvisync |
| RxNorm | 1598392 | empagliflozin / linagliptin |
| RxNorm | 1602106 | empagliflozin / linagliptin Oral Product |
| RxNorm | 1602107 | empagliflozin / linagliptin Pill |

Continued on next page

**Table28 – continued from previous page**

| CodeSystem | ConceptCode | ConceptName |
|---|---|---|
| RxNorm | 1602108 | empagliflozin / linagliptin Oral Tablet |
| RxNorm | 1602109 | empagliflozin 10 MG / linagliptin 5 MG Oral Tablet |
| RxNorm | 1602110 | Glyxambi |
| RxNorm | 1602111 | empagliflozin 10 MG / linagliptin 5 MG [Glyxambi] |
| RxNorm | 1602112 | empagliflozin / linagliptin Oral Tablet [Glyxambi] |
| RxNorm | 1602113 | Glyxambi Oral Product |
| RxNorm | 1602114 | Glyxambi Pill |
| RxNorm | 1602115 | empagliflozin 10 MG / linagliptin 5 MG Oral Tablet [Glyxambi] |
| RxNorm | 1602118 | empagliflozin 25 MG / linagliptin 5 MG Oral Tablet |
| RxNorm | 1602119 | empagliflozin 25 MG / linagliptin 5 MG [Glyxambi] |
| RxNorm | 1602120 | empagliflozin 25 MG / linagliptin 5 MG Oral Tablet [Glyxambi] |
| RxNorm | 1727500 | dapagliflozin / saxagliptin |
| RxNorm | 1796088 | linagliptin / metformin Extended Release Oral Tablet |
| RxNorm | 1796089 | 24 HR linagliptin 2.5 MG / metformin hydrochloride 1000 MG Extended Release Oral Tablet |
| RxNorm | 1796090 | linagliptin / metformin Extended Release Oral Tablet [Jentadueto] |
| RxNorm | 1796091 | 24 HR linagliptin 2.5 MG / metformin hydrochloride 1000 MG Extended Release Oral Tablet [Jentadueto] |
| RxNorm | 1796092 | linagliptin 2.5 MG / metformin hydrochloride 1000 MG Extended Release Oral Tablet |
| RxNorm | 1796093 | linagliptin 2.5 MG / metformin hydrochloride 1000 MG Extended Release Oral Tablet [Jentadueto] |
| RxNorm | 1796094 | 24 HR linagliptin 5 MG / metformin hydrochloride 1000 MG Extended Release Oral Tablet |
| RxNorm | 1796095 | linagliptin 5 MG / metformin hydrochloride 1000 MG [Jentadueto] |
| RxNorm | 1796096 | 24 HR linagliptin 5 MG / metformin hydrochloride 1000 MG Extended Release Oral Tablet [Jentadueto] |
| RxNorm | 1796097 | linagliptin 5 MG / metformin hydrochloride 1000 MG Extended Release Oral Tablet |
| RxNorm | 1796098 | linagliptin 5 MG / metformin hydrochloride 1000 MG Extended Release Oral Tablet [Jentadueto] |
| RxNorm | 1925495 | dapagliflozin / saxagliptin Oral Product |
| RxNorm | 1925496 | dapagliflozin / saxagliptin Pill |
| RxNorm | 1925497 | dapagliflozin / saxagliptin Oral Tablet |
| RxNorm | 1925498 | dapagliflozin 10 MG / saxagliptin 5 MG Oral Tablet |
| RxNorm | 1925499 | Qtern |
| RxNorm | 1925500 | dapagliflozin 10 MG / saxagliptin 5 MG [Qtern] |
| RxNorm | 1925501 | dapagliflozin / saxagliptin Oral Tablet [Qtern] |
| RxNorm | 1925502 | Qtern Oral Product |

Continued on next page

**Table28 – continued from previous page**

| CodeSystem | ConceptCode | ConceptName |
|---|---|---|
| RxNorm | 1925503 | Qtern Pill |
| RxNorm | 1925504 | dapagliflozin 10 MG / saxagliptin 5 MG Oral Tablet [Qtern] |
| RxNorm | 1992822 | ertugliflozin / sitagliptin Oral Product |
| RxNorm | 1992823 | ertugliflozin / sitagliptin Pill |
| RxNorm | 1992824 | ertugliflozin / sitagliptin Oral Tablet |
| RxNorm | 1992825 | ertugliflozin / sitagliptin |
| RxNorm | 1992826 | ertugliflozin 15 MG / sitagliptin 100 MG Oral Tablet |
| RxNorm | 1992827 | Steglujan |
| RxNorm | 1992828 | ertugliflozin 15 MG / sitagliptin 100 MG [Steglujan] |
| RxNorm | 1992829 | ertugliflozin / sitagliptin Oral Tablet [Steglujan] |
| RxNorm | 1992830 | Steglujan Oral Product |
| RxNorm | 1992831 | Steglujan Pill |
| RxNorm | 1992832 | ertugliflozin 15 MG / sitagliptin 100 MG Oral Tablet [Steglujan] |
| RxNorm | 1992835 | ertugliflozin 5 MG / sitagliptin 100 MG Oral Tablet |
| RxNorm | 1992836 | ertugliflozin 5 MG / sitagliptin 100 MG [Steglujan] |
| RxNorm | 1992837 | ertugliflozin 5 MG / sitagliptin 100 MG Oral Tablet [Steglujan] |
| RxNorm | 2117292 | dapagliflozin / metformin / saxagliptin |
| RxNorm | 2169274 | dapagliflozin 5 MG / saxagliptin 5 MG Oral Tablet |
| RxNorm | 2169275 | dapagliflozin 5 MG / saxagliptin 5 MG [Qtern] |
| RxNorm | 2169276 | dapagliflozin 5 MG / saxagliptin 5 MG Oral Tablet [Qtern] |
| RxNorm | 2281864 | empagliflozin / linagliptin / metformin |
| RxNorm | 2371722 | dapagliflozin / metformin / saxagliptin Oral Product |
| RxNorm | 2371723 | dapagliflozin / metformin / saxagliptin Pill |
| RxNorm | 2371724 | dapagliflozin / metformin / saxagliptin Extended Release Oral Tablet |
| RxNorm | 2371725 | 24 HR dapagliflozin 10 MG / metformin hydrochloride 1000 MG / saxagliptin 5 MG Extended Release Oral Tablet |
| RxNorm | 2371726 | Qternmet |
| RxNorm | 2371727 | dapagliflozin 10 MG / metformin hydrochloride 1000 MG / saxagliptin 5 MG [Qternmet] |
| RxNorm | 2371728 | dapagliflozin / metformin / saxagliptin Extended Release Oral Tablet [Qternmet] |
| RxNorm | 2371729 | Qternmet Oral Product |
| RxNorm | 2371730 | Qternmet Pill |
| RxNorm | 2371731 | 24 HR dapagliflozin 10 MG / metformin hydrochloride 1000 MG / saxagliptin 5 MG Extended Release Oral Tablet [Qternmet] |
| RxNorm | 2371732 | dapagliflozin 10 MG / metformin hydrochloride 1000 MG / saxagliptin 5 MG Extended Release Oral Tablet |
| RxNorm | 2371733 | dapagliflozin 10 MG / metformin hydrochloride 1000 MG / saxagliptin 5 MG Extended Release Oral Tablet [Qternmet] |

Continued on next page

**Table28 – continued from previous page**

| CodeSystem | ConceptCode | ConceptName |
|---|---|---|
| RxNorm | 2371734 | 24 HR dapagliflozin 2.5 MG / metformin hydrochloride 1000 MG / saxagliptin 2.5 MG Extended Release Oral Tablet |
| RxNorm | 2371735 | dapagliflozin 2.5 MG / metformin hydrochloride 1000 MG / saxagliptin 2.5 MG [Qternmet] |
| RxNorm | 2371736 | 24 HR dapagliflozin 2.5 MG / metformin hydrochloride 1000 MG / saxagliptin 2.5 MG Extended Release Oral Tablet [Qternmet] |
| RxNorm | 2371737 | dapagliflozin 2.5 MG / metformin hydrochloride 1000 MG / saxagliptin 2.5 MG Extended Release Oral Tablet |
| RxNorm | 2371738 | dapagliflozin 2.5 MG / metformin hydrochloride 1000 MG / saxagliptin 2.5 MG Extended Release Oral Tablet [Qternmet] |
| RxNorm | 2371740 | 24 HR dapagliflozin 5 MG / metformin hydrochloride 1000 MG / saxagliptin 2.5 MG Extended Release Oral Tablet |
| RxNorm | 2371741 | dapagliflozin 5 MG / metformin hydrochloride 1000 MG / saxagliptin 2.5 MG [Qternmet] |
| RxNorm | 2371742 | 24 HR dapagliflozin 5 MG / metformin hydrochloride 1000 MG / saxagliptin 2.5 MG Extended Release Oral Tablet [Qternmet] |
| RxNorm | 2371743 | dapagliflozin 5 MG / metformin hydrochloride 1000 MG / saxagliptin 2.5 MG Extended Release Oral Tablet |
| RxNorm | 2371744 | dapagliflozin 5 MG / metformin hydrochloride 1000 MG / saxagliptin 2.5 MG Extended Release Oral Tablet [Qternmet] |
| RxNorm | 2371745 | 24 HR dapagliflozin 5 MG / metformin hydrochloride 1000 MG / saxagliptin 5 MG Extended Release Oral Tablet |
| RxNorm | 2371746 | dapagliflozin 5 MG / metformin hydrochloride 1000 MG / saxagliptin 5 MG [Qternmet] |
| RxNorm | 2371747 | 24 HR dapagliflozin 5 MG / metformin hydrochloride 1000 MG / saxagliptin 5 MG Extended Release Oral Tablet [Qternmet] |
| RxNorm | 2371748 | dapagliflozin 5 MG / metformin hydrochloride 1000 MG / saxagliptin 5 MG Extended Release Oral Tablet |
| RxNorm | 2371749 | dapagliflozin 5 MG / metformin hydrochloride 1000 MG / saxagliptin 5 MG Extended Release Oral Tablet [Qternmet] |
| RxNorm | 2359276 | empagliflozin / linagliptin / metformin Oral Product |
| RxNorm | 2359277 | empagliflozin / linagliptin / metformin Pill |
| RxNorm | 2359278 | empagliflozin / linagliptin / metformin Extended Release Oral Tablet |
| RxNorm | 2359279 | 24 HR empagliflozin 10 MG / linagliptin 5 MG / metformin hydrochloride 1000 MG Extended Release Oral Tablet |
| RxNorm | 2359280 | Trijardy |

Continued on next page

**Table28 – continued from previous page**

| CodeSystem | ConceptCode | ConceptName |
| --- | --- | --- |
| RxNorm | 2359281 | empagliflozin 10 MG / linagliptin 5 MG / metformin hydrochloride 1000 MG [Trijardy] |
| RxNorm | 2359282 | empagliflozin / linagliptin / metformin Extended Release Oral Tablet [Trijardy] |
| RxNorm | 2359283 | Trijardy Oral Product |
| RxNorm | 2359284 | Trijardy Pill |
| RxNorm | 2359285 | 24 HR empagliflozin 10 MG / linagliptin 5 MG / metformin hydrochloride 1000 MG Extended Release Oral Tablet [Trijardy] |
| RxNorm | 2359286 | empagliflozin 10 MG / linagliptin 5 MG / metformin hydrochloride 1000 MG Extended Release Oral Tablet |
| RxNorm | 2359287 | empagliflozin 10 MG / linagliptin 5 MG / metformin hydrochloride 1000 MG Extended Release Oral Tablet [Trijardy] |
| RxNorm | 2359288 | 24 HR empagliflozin 12.5 MG / linagliptin 2.5 MG / metformin hydrochloride 1000 MG Extended Release Oral Tablet |
| RxNorm | 2359289 | empagliflozin 12.5 MG / linagliptin 2.5 MG / metformin hydrochloride 1000 MG [Trijardy] |
| RxNorm | 2359290 | 24 HR empagliflozin 12.5 MG / linagliptin 2.5 MG / metformin hydrochloride 1000 MG Extended Release Oral Tablet [Trijardy] |
| RxNorm | 2359291 | empagliflozin 12.5 MG / linagliptin 2.5 MG / metformin hydrochloride 1000 MG Extended Release Oral Tablet |
| RxNorm | 2359292 | empagliflozin 12.5 MG / linagliptin 2.5 MG / metformin hydrochloride 1000 MG Extended Release Oral Tablet [Trijardy] |
| RxNorm | 2359351 | 24 HR empagliflozin 25 MG / linagliptin 5 MG / metformin hydrochloride 1000 MG Extended Release Oral Tablet |
| RxNorm | 2359352 | empagliflozin 25 MG / linagliptin 5 MG / metformin hydrochloride 1000 MG [Trijardy] |
| RxNorm | 2359353 | 24 HR empagliflozin 25 MG / linagliptin 5 MG / metformin hydrochloride 1000 MG Extended Release Oral Tablet [Trijardy] |
| RxNorm | 2359354 | empagliflozin 25 MG / linagliptin 5 MG / metformin hydrochloride 1000 MG Extended Release Oral Tablet |
| RxNorm | 2359355 | empagliflozin 25 MG / linagliptin 5 MG / metformin hydrochloride 1000 MG Extended Release Oral Tablet [Trijardy] |
| RxNorm | 2359356 | 24 HR empagliflozin 5 MG / linagliptin 2.5 MG / metformin hydrochloride 1000 MG Extended Release Oral Tablet |
| RxNorm | 2359357 | empagliflozin 5 MG / linagliptin 2.5 MG / metformin hydrochloride 1000 MG [Trijardy] |

Continued on next page

**Table28 – continued from previous page**

| CodeSystem | ConceptCode | ConceptName |
|---|---|---|
| RxNorm | 2359358 | 24 HR empagliflozin 5 MG / linagliptin 2.5 MG / metformin hydrochloride 1000 MG Extended Release Oral Tablet [Trijardy] |
| RxNorm | 2359359 | empagliflozin 5 MG / linagliptin 2.5 MG / metformin hydrochloride 1000 MG Extended Release Oral Tablet |
| RxNorm | 2359360 | empagliflozin 5 MG / linagliptin 2.5 MG / metformin hydrochloride 1000 MG Extended Release Oral Tablet [Trijardy] |
| RxNorm | 705136 | Metformin 1000 MG / sitagliptin 50 MG Oral Tablet [Janumet] |
| RxNorm | 705134 | Metformin 1000 MG / sitagliptin 50 MG [Janumet] |
| RxNorm | 705135 | Metformin / sitagliptin Oral Tablet [Janumet] |
| RxNorm | 705138 | Metformin 500 MG / sitagliptin 50 MG Oral Tablet [Janumet] |
| RxNorm | 748763 | Metformin 500 MG / sitagliptin 64.3 MG Oral Tablet |
| RxNorm | 757607 | Metformin / sitagliptin Oral Tablet [Janumet 50 mg/500 mg] |
| RxNorm | 748762 | sitagliptin 64.3 MG |
| RxNorm | 748765 | Metformin 500 MG / sitagliptin 64.3 MG Oral Tablet [Janumet] |
| RxNorm | 705137 | Metformin 500 MG / sitagliptin 50 MG [Janumet] |
| RxNorm | 748764 | Metformin 500 MG / sitagliptin 64.3 MG [Janumet] |
| RxNorm | 1043581 | Metformin / saxagliptin Extended Release Tablet [Kombiglyze 5/500] |
| RxNorm | 1043573 | Metformin / saxagliptin Extended Release Tablet [Kombiglyze 5/1000] |
| RxNorm | 1189807 | Simvastatin / sitagliptin Oral Tablet [Juvisync 100/10] |
| RxNorm | 1189824 | Simvastatin / sitagliptin Oral Tablet [Juvisync 100/40] |
| RxNorm | 1243031 | Jentadueto 2.5/500 Oral Product |
| RxNorm | 1243025 | Jentadueto 2.5/1000 Pill |
| RxNorm | 1173547 | Kombiglyze 5/500 Oral Product |
| RxNorm | 1189825 | Juvisync 100/40 Oral Product |
| RxNorm | 1172859 | Kombiglyze 2.5/1000 Oral Product |
| RxNorm | 1173546 | Kombiglyze 5/1000 Pill |
| RxNorm | 1243832 | Janumet 100/1000 Pill |
| RxNorm | 1189816 | Juvisync 100/10 Oral Product |
| RxNorm | 1189826 | Juvisync 100/40 Pill |
| RxNorm | 1189817 | Juvisync 100/10 Pill |
| RxNorm | 1167812 | Janumet 50/500 Oral Product |
| RxNorm | 1243032 | Jentadueto 2.5/500 Pill |
| RxNorm | 1173548 | Kombiglyze 5/500 Pill |
| RxNorm | 1167813 | Janumet 50/500 Pill |
| RxNorm | 1243024 | Jentadueto 2.5/1000 Oral Product |

Continued on next page

**Table28 – continued from previous page**

| CodeSystem | ConceptCode | ConceptName |
|---|---|---|
| RxNorm | 1243831 | Janumet 100/1000 Oral Product |
| RxNorm | 665031 | sitagliptin 100 MG |
| RxNorm | 665032 | sitagliptin Oral Tablet |
| RxNorm | 665033 | sitagliptin 100 MG Oral Tablet |
| RxNorm | 665034 | sitagliptin 100 MG [Januvia] |
| RxNorm | 665035 | sitagliptin Oral Tablet [Januvia] |
| RxNorm | 665036 | sitagliptin 100 MG Oral Tablet [Januvia] |
| RxNorm | 665037 | sitagliptin 25 MG |
| RxNorm | 665038 | sitagliptin 25 MG Oral Tablet |
| RxNorm | 665039 | sitagliptin 25 MG [Januvia] |
| RxNorm | 665040 | sitagliptin 25 MG Oral Tablet [Januvia] |
| RxNorm | 665041 | sitagliptin 50 MG |
| RxNorm | 665042 | sitagliptin 50 MG Oral Tablet |
| RxNorm | 665043 | sitagliptin 50 MG [Januvia] |
| RxNorm | 665044 | sitagliptin 50 MG Oral Tablet [Januvia] |
| RxNorm | 638596 | Januvia |
| RxNorm | 700516 | metformin / sitagliptin Oral Tablet |
| RxNorm | 729717 | metformin / sitagliptin |
| RxNorm | 757603 | metformin / sitagliptin Oral Tablet [Janumet] |
| RxNorm | 857974 | saxagliptin |
| RxNorm | 858034 | saxagliptin 5 MG |
| RxNorm | 858035 | saxagliptin Oral Tablet |
| RxNorm | 858036 | saxagliptin 5 MG Oral Tablet |
| RxNorm | 858037 | Onglyza |
| RxNorm | 858038 | saxagliptin 5 MG [Onglyza] |
| RxNorm | 858039 | saxagliptin Oral Tablet [Onglyza] |
| RxNorm | 858040 | saxagliptin 5 MG Oral Tablet [Onglyza] |
| RxNorm | 858041 | saxagliptin 2.5 MG |
| RxNorm | 858042 | saxagliptin 2.5 MG Oral Tablet |
| RxNorm | 858043 | saxagliptin 2.5 MG [Onglyza] |
| RxNorm | 858044 | saxagliptin 2.5 MG Oral Tablet [Onglyza] |
| RxNorm | 861769 | metformin hydrochloride 1000 MG / sitagliptin 50 MG Oral Tablet |
| RxNorm | 861770 | metformin hydrochloride 1000 MG / sitagliptin 50 MG [Janumet] |
| RxNorm | 861771 | metformin hydrochloride 1000 MG / sitagliptin 50 MG Oral Tablet [Janumet] |
| RxNorm | 861819 | metformin hydrochloride 500 MG / sitagliptin 50 MG Oral Tablet |
| RxNorm | 861820 | metformin hydrochloride 500 MG / sitagliptin 50 MG [Janumet] |
| RxNorm | 861821 | metformin hydrochloride 500 MG / sitagliptin 50 MG Oral Tablet [Janumet] |

Continued on next page

**Table28 – continued from previous page**

| CodeSystem | ConceptCode | ConceptName |
|---|---|---|

#### 8.6 Anti-Obesity Medications (AOMS)

##### 8.6.1 Orlistat

Table 29: Concept codes used to identify if a patient was prescribed orlistat.

| CodeSystem | ConceptCode | ConceptName |
|---|---|---|
| RxNorm | 1159582 | orlistat Oral Product |
| RxNorm | 1159583 | orlistat Pill |
| RxNorm | 1168454 | Alli Oral Product |
| RxNorm | 1168455 | Alli Pill |
| RxNorm | 1186415 | Xenical Oral Product |
| RxNorm | 1186416 | Xenical Pill |
| RxNorm | 226917 | orlistat 120 MG Oral Capsule [Xenical] |
| RxNorm | 226918 | Xenical |
| RxNorm | 314153 | orlistat 120 MG Oral Capsule |
| RxNorm | 330389 | orlistat 120 MG |
| RxNorm | 366436 | orlistat Oral Capsule [Xenical] |
| RxNorm | 373151 | orlistat Oral Capsule |
| RxNorm | 37925 | orlistat |
| RxNorm | 574045 | orlistat 120 MG [Xenical] |
| RxNorm | 692875 | orlistat 60 MG |
| RxNorm | 692876 | orlistat 60 MG Oral Capsule |
| RxNorm | 708361 | Alli |
| RxNorm | 723844 | orlistat 60 MG [Alli] |
| RxNorm | 723845 | orlistat Oral Capsule [Alli] |
| RxNorm | 723846 | orlistat 60 MG Oral Capsule [Alli] |
| SNOMED CT | 116093009 | Orlistat-containing product |
| SNOMED CT | 387007000 | Orlistat |
| SNOMED CT | 317893003 | Orlistat 120mg capsule |
| SNOMED CT | 426638000 | Orlistat 60mg capsule |
| SNOMED CT | 768105006 | Orlistat-containing product in oral dose form |
| SNOMED CT | 777002008 | Orlistat only product |
| SNOMED CT | 780075002 | Orlistat only product in oral dose form |

##### 8.6.2 Phentermine Topiramate

Table 30: Concept codes used to identify if a patient was prescribed phentermine-topiramate.

| CodeSystem | ConceptCode | ConceptName |
|---|---|---|
| RxNorm | 1302868 | Phentermine 11.25 MG / topiramate 69 MG Extended Release Capsule |
| RxNorm | 1302827 | 24 HR phentermine 7.5 MG / topiramate 46 MG Extended Release Oral Capsule |
| RxNorm | 1302839 | 24 HR phentermine 3.75 MG / topiramate 23 MG Extended Release Oral Capsule |
| RxNorm | 1302850 | 24 HR phentermine 15 MG / topiramate 92 MG Extended Release Oral Capsule |
| RxNorm | 1313059 | 24 HR phentermine 11.25 MG / topiramate 69 MG Extended Release Oral Capsule |
| RxNorm | 1302833 | 24 HR phentermine 7.5 MG / topiramate 46 MG Extended Release Oral Capsule [Qsymia] |
| RxNorm | 1302845 | 24 HR phentermine 3.75 MG / topiramate 23 MG Extended Release Oral Capsule [Qsymia] |
| RxNorm | 1302856 | 24 HR phentermine 15 MG / topiramate 92 MG Extended Release Oral Capsule [Qsymia] |
| RxNorm | 1313061 | 24 HR phentermine 11.25 MG / topiramate 69 MG Extended Release Oral Capsule [Qsymia] |
